# Forecasting global progress towards the United Nations hypertension control target for 2030

**DOI:** 10.64898/2026.09.22.26363661

**Authors:** Thiago André Carniel, Aline Mânica, Walter Strobel Neto, Elisângela Pinheiro, Junir Antonio Lutinski, Fátima Kremer Ferretti, Samuel Spiegelberg Zuge, Letícia de Lima Trindade, Clodoaldo Antônio De Sá

## Abstract

In 2025, the United Nations General Assembly adopted a target of 150 million additional people with hypertension under control by 2030, but did not specify the baseline year. We quantified how baseline choice affects estimated progress towards this target and forecast the number of adults with controlled hypertension globally through 2030. Age- and sex-specific hypertension prevalence and control estimates from the NCD Risk Factor Collaboration for 1990–2019 were combined with United Nations World Population Prospects 2024 for adults aged 30–79 years in 200 countries and territories. Bayesian state-space models generated separate prevalence and control forecasts, propagated through 100 000 posterior draws. We evaluated baselines of 2020, 2023, 2024 and 2025 and decomposed the 2024–2030 change using continuous-change decomposition. Holding the 2030 forecast constant, estimated target attainment ranged from 94.4% with a 2020 baseline to 51.4% with a 2025 baseline. Using the prespecified 2024 baseline, the number of adults with controlled hypertension was forecast to increase from 332.4 million in 2024 to 423.9 million in 2030, an increase of 91.5 million (95% uncertainty interval 75.2–110.0 million), corresponding to 61.0% (95% uncertainty interval 50.2–73.3%) of the target and a median shortfall of 58.5 million. The control-rate improvement component accounted for 66.3% of the increase and demographic change for 44.0%, while declining age–sex-specific hypertension prevalence offset 10.4%. Progress towards this absolute target cannot be interpreted unambiguously without an explicit baseline year. From a 2024 baseline, current forecast trajectories fall short of the 2030 target.

## Introduction

Hypertension is a leading modifiable risk factor for cardiovascular disease and premature death worldwide [1, 2]. In 2024, about 1.4 billion adults aged 30–79 years had hypertension, and approximately 23% had their blood pressure controlled [3]. Increasing the number of people with controlled hypertension is now a formal international commitment.

In December 2025, the United Nations (UN) General Assembly committed to 150 million additional people with hypertension under control by 2030 [4]. The commitment is expressed as an absolute count, but the resolution does not specify the baseline year. Its predecessor used a different metric and an explicit reference: the World Health Organization (WHO) global monitoring framework set a 25% relative reduction in raised blood pressure by 2025 against a 2010 baseline [5]. This distinction matters because absolute counts respond to demographic forces that prevalence rates do not.

Population growth and ageing can increase hypertension counts even when age-specific prevalence is stable or declining [6–8]. WHO estimates indicate that the number of adults with hypertension increased from 650 million in 1990 to 1.4 billion in 2024, largely because of growth in the number of older adults in low- and middle-income countries [3]. Among adults aged 20 years or older in high-income countries, age-standardized prevalence fell by 2.7 percentage points between 2000 and 2020 while the absolute number with hypertension increased by 76 million [9]. Progress towards an absolute-count target can therefore reflect, in part, changes in population size and structure even without improvement in age–sex-specific control rates.

To our knowledge, the combined implications of epidemiological trends, demographic change and analytical baseline choice for the new UN hypertension-control target have not been quantified globally. We therefore forecast the number of adults aged 30–79 years with controlled hypertension to 2030, assessed progress towards the 150-million target under alternative analytical baselines, and decomposed the forecast change into population size, age–sex composition, hypertension prevalence and control-rate improvement at global, regional and country levels.

## Methods

### Data sources and study population

We used age- and sex-specific estimates of hypertension prevalence and control from 1990 to 2019 produced by the NCD Risk Factor Collaboration (NCD-RisC), based on 1201 population-representative studies comprising 104 million participants [10]. Hypertension was defined as systolic blood pressure of at least 140 mmHg, diastolic blood pressure of at least 90 mmHg, or use of antihypertensive medication. Control was defined among people with hypertension as antihypertensive treatment with systolic and diastolic blood pressure below 140 and 90 mmHg, respectively [10].

Age- and sex-specific population estimates and projections were obtained from the United Nations (UN) World Population Prospects 2024 (WPP 2024) [11]. The analysis included 200 countries and territories and 20 age–sex strata per country, comprising men and women aged 30–79 years in ten 5-year age groups. These entities represented approximately 99.8% of the global population in this age range in 2024 and 2030. Country results were aggregated to the six WHO regions according to the WHO reporting convention detailed in Supplementary Methods S5. Following World Health Assembly resolution 78.25, Indonesia was assigned to the Western Pacific Region throughout the analytical series [12]. Prevalence and control were modelled separately and combined only when calculating absolute counts.

### Primary outcome and analytical baseline

For each country and posterior draw, the number of adults with controlled hypertension was calculated by summing across age–sex strata the product of population, hypertension prevalence, and the proportion of people with hypertension whose blood pressure was controlled. Country estimates were summed draw-by-draw to obtain regional and global totals. Within each draw, aggregate hypertension prevalence was calculated as the number of adults with hypertension divided by the corresponding population aged 30– 79 years, and aggregate control as the number with controlled hypertension divided by the number with hypertension.

The primary outcome was the change in the global number of adults with controlled hypertension between 2024 and 2030. The UN General Assembly established a global target of 150 million additional people with hypertension under control by 2030 but did not specify an analytical baseline [4]. All candidate baseline years after 2019 were model-based forecasts. We prespecified 2024 as the primary analytical baseline because it was the last completed calendar year entirely preceding adoption of the resolution on 15 December 2025. Using 2025 would place the baseline within the year in which the target itself was adopted. Baselines of 2020, 2023 and 2025 were examined in sensitivity analyses.

Progress towards the target was calculated within each posterior draw as the forecast increase in the number of adults with controlled hypertension divided by 150 million. Regional contributions were expressed as shares of this same global target and were not interpreted as region-specific targets. Countries highlighted as leading absolute contributors were ranked by posterior mean change. Full selection and geographical-display procedures are provided in Supplementary Methods S5.

### Bayesian forecasting model

Hypertension prevalence and control were forecast separately on the logit scale using Bayesian local-linear-trend state-space models. Each age–sex stratum had a latent level and time-varying slope, allowing both the direction and rate of epidemiological change to evolve over time rather than imposing a fixed linear trajectory. Bayesian approaches for temporally evolving population-level health measures, including blood-pressure outcomes, have previously been described [13].

Slope innovations were correlated across age–sex strata within each country and epidemiological series so that related subpopulations could evolve coherently while retaining stratum-specific variation. The primary forecasting and uncertainty specifications were fixed before global production and before calculation of global or regional study outcomes. Forecasts were anchored to the published NCD-RisC central estimates for 2019 and propagated sequentially through 2030. We generated 100 000 posterior draws for each reported analysis, allowing forecast uncertainty to accumulate with increasing horizon. Full model equations, priors, posterior computation and convergence criteria are provided in Supplementary Methods S5.

### Uncertainty and dependence assumptions

Published NCD-RisC uncertainty intervals were not treated as independent annual observational errors. The annual estimates and their 95% credible intervals are themselves outputs of a Bayesian hierarchical model with temporal smoothing [10]. Consequently, interval widths can reflect model structure and boundary behavior near the beginning and end of the estimated series rather than year-specific measurement uncertainty alone.

We therefore modelled uncertainty across the complete epidemiological trajectory. Uncertainty in the epidemiological level was represented through a separate asymmetric layer calibrated from an interior portion of the published NCD-RisC uncertainty intervals, reducing sensitivity to boundary-related interval geometry. Because prevalence–control and between-country dependence cannot be recovered from the published marginal uncertainty intervals, the primary analysis introduced no additional dependence between epidemiological series or countries. Alternative dependence assumptions were examined in sensitivity analyses. All reported 95% uncertainty intervals (95% UIs) were obtained from posterior draws. The full uncertainty architecture and dependence specifications are provided in Supplementary Methods S5.

### Decomposition analysis

We decomposed the change in the absolute number of adults with controlled hypertension using the continuous-change decomposition method [14]. Four components were quantified: total population size, age–sex population composition, age–sex-specific hypertension prevalence, and the control-rate improvement component among people with hypertension.

The two demographic components capture distinct aspects of demographic change. The population-size component captures the effect of growth or decline in the total number of adults aged 30–79 years, independently of changes in the population structure. The age–sex-composition component captures changes in the relative distribution of the population across age and sex strata, independently of changes in total population size. In this study, this component largely reflects population ageing, because a growing proportion of adults moves into older age groups, in which hypertension prevalence is higher, although the component also incorporates changes in sex composition. We therefore defined the demographic contribution as the sum of the population-size and age–sex-composition components.

The prevalence component captures changes in age–sex-specific hypertension prevalence, whereas the control-rate improvement component captures changes in the proportion of people with hypertension whose blood pressure is controlled. Thus, the decomposition distinguishes demographic changes in the size and structure of the population from epidemiological changes in hypertension prevalence and control.

Decomposition was performed within each posterior draw, and country-level components were summed draw-by-draw to obtain regional and global contributions. Component fractions were reported only when the 2.5th percentile of the corresponding total change was greater than zero. The prespecified temporal comparison used matched six-year periods, 2013–2019 and 2024–2030, with absolute changes expressed on an annualized basis and component-share differences calculated using aligned posterior draws. Computational implementation and numerical checks are described in Supplementary Methods S5.

### Model assessment, ethics and software

Posterior convergence, numerical consistency, uncertainty propagation and decomposition closure were assessed as described in Supplementary Methods S5. Analyses were performed in MATLAB R2024b. The study used only publicly available aggregate data: modelled estimates from NCD-RisC and population projections from WPP 2024. No individual-level data or personal identifiers were accessed, and ethics committee approval was not required for this secondary analysis. Reporting followed the Guidelines for Accurate and Transparent Health Estimates Reporting (GATHER) statement [15].

### Use of generative artificial intelligence

During manuscript preparation, the authors used ChatGPT (OpenAI) and Claude (Anthropic) to assist with drafting, language editing, grammar, clarity, and secondary numerical checks of reported values. Generative artificial intelligence was not used to generate the analytical results, which were produced by the authors’ MATLAB pipeline. All AI-assisted content was critically reviewed and verified by the authors, who take full responsibility for the final manuscript.

## Results

### Forecast global progress towards the target

The global number of adults aged 30–79 years with controlled hypertension was forecast to increase from 332.4 million in 2024 to 423.9 million in 2030 (Fig. 1A). This represented an increase of 91.5 million people (95% UI: 75.2 to 110.0 million), equivalent to 61.0% (95% UI: 50.2% to 73.3%) of the UN target of 150 million additional people with hypertension under control by 2030. Under the primary 2024 baseline, full attainment of the target would correspond to approximately 482.4 million adults with controlled hypertension in 2030, leaving the median forecast 58.5 million below this level.

**Figure 1.**
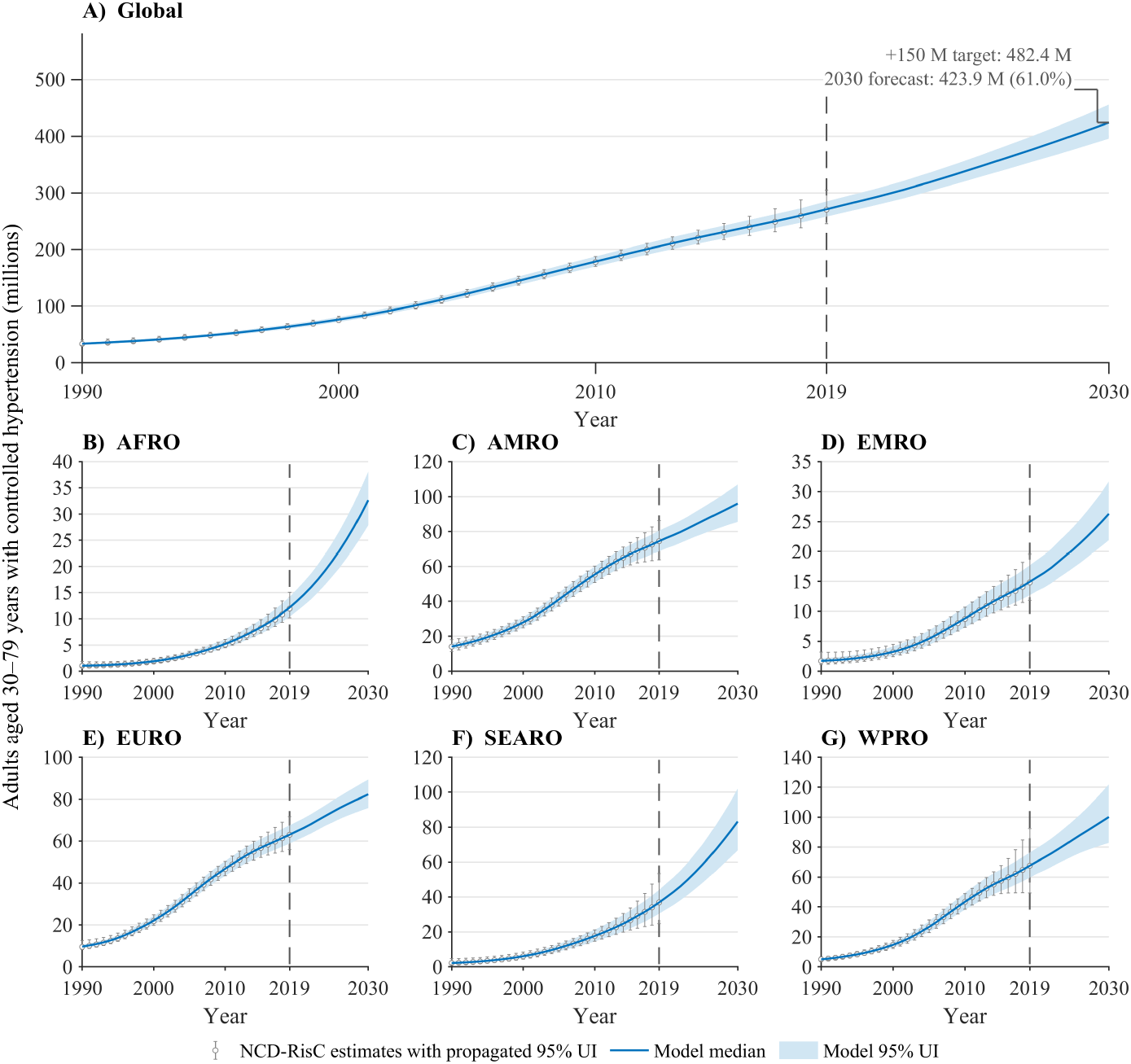
Estimated and forecast number of adults aged 30–79 years with controlled hypertension, globally and by World Health Organization (WHO) region, 1990–2030. (A) Global. (B) African Region (AFRO). (C) Region of the Americas (AMRO). (D) Eastern Mediterranean Region (EMRO). (E) European Region (EURO). (F) South-East Asia Region (SEARO). (G) Western Pacific Region (WPRO). Open circles and error bars show annual NCD-RisC estimates and 95% uncertainty intervals (UIs) for 1990–2019. The solid line and shaded band show the model median and 95% UI through 2030. The vertical dashed line marks 2019, the final year of NCD-RisC estimates. In panel A, the horizontal reference indicates the level corresponding to 150 million additional adults with controlled hypertension relative to the 2024 baseline.

### Dependence of target attainment on the baseline year

Estimated progress towards the target varied substantially with the analytical baseline year (Supplementary Fig. S1). Using baselines of 2020, 2023, 2024 and 2025 yielded forecast increases by 2030 of 141.6, 105.3, 91.5 and 77.1 million adults, respectively, corresponding to 94.4%, 70.2%, 61.0% and 51.4% of the target. Thus, estimated progress from a 2020 baseline was 1.84 times that obtained from 2025. This variation reflected differences in the starting values, while the 2030 forecast remained unchanged. The UN resolution did not specify which baseline year should be used. In contrast, the median demographic share spanned only 2.52 percentage points, and no individual component share varied by more than 3.09 percentage points across the four baselines.

### Decomposition of the forecast change

The forecast increase from 2024 to 2030 was driven primarily by improved control among people with hypertension and by demographic change (Fig. 2; Table 1). The control-rate improvement component accounted for 66.3% (95% UI: 59.6% to 72.8%) of the global increase. Demographic change accounted for 44.0% (95% UI: 38.2% to 51.5%), comprising 24.4% (95% UI: 20.9% to 28.7%) from population-size change and 19.6% (95% UI: 17.0% to 23.0%) from changes in age–sex composition. Age–sex compositional change therefore represented 44.5% of the demographic subtotal. Declining age–sex-specific hypertension prevalence partly offset these increases, contributing ™10.4% (95% UI: ™18.9% to ™3.0%).

**Figure 2.**
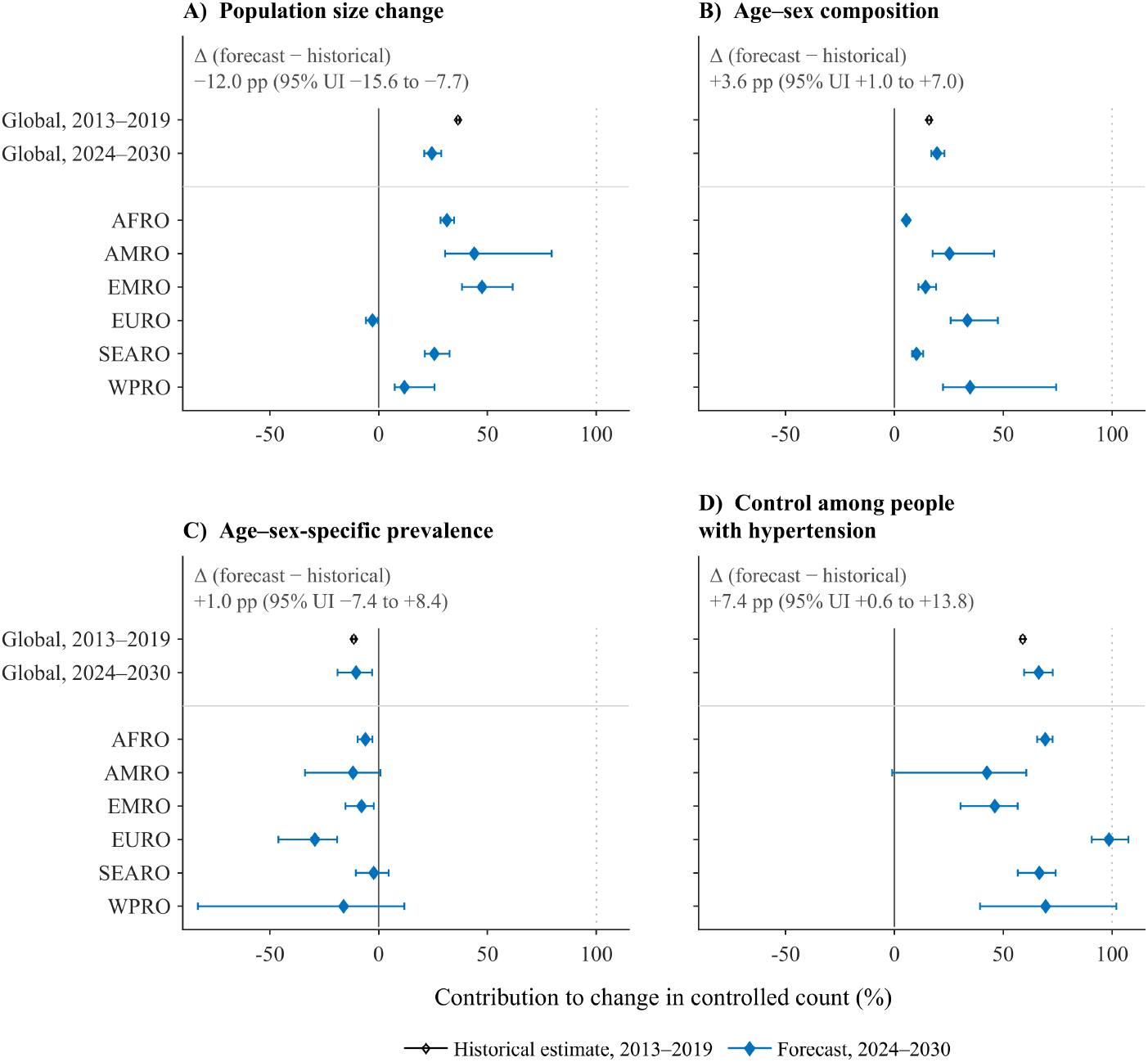
Horiuchi decomposition of the change in the number of adults aged 30–79 years with controlled hypertension, globally and by World Health Organization (WHO) region. (A) Population-size change. (B) Age–sex composition. (C) Age–sex-specific hypertension prevalence. (D) Control-rate improvement among people with hypertension. Open black diamonds show global component fractions for 2013–2019. Filled blue diamonds show component fractions for the 2024–2030 forecast globally and for the African Region (AFRO), Region of the Americas (AMRO), Eastern Mediterranean Region (EMRO), European Region (EURO), South-East Asia Region (SEARO), and Western Pacific Region (WPRO). Diamonds represent medians and error bars represent 95% uncertainty intervals (UIs). Text annotations show the paired forecast-minus-historical difference in the global component fraction, expressed in percentage points with 95% UI. Regional estimates refer to 2024–2030 only. Panels A and B together represent the demographic change component.

**Table 1.** Forecast increase in the number of adults aged 30–79 years with controlled hypertension and decomposition of the increase, globally and by WHO region, with Taiwan, China reported separately, 2024–2030.

| Geographical grouping | Forecast increase, millions (95% UI) | Contribution to the global 150-million target, % (95% UI) | Decomposition of the forecast increase, % (95% UI) |  |  |  |  |
| --- | --- | --- | --- | --- | --- | --- | --- |
|  |  |  | Demographic change |  |  | Age–sex-specific prevalence | Control among people with hypertension |
|  |  |  | Total | Population size change | Age–sex composition |  |  |
| Global | 91.5<br>(75.2 to 110.0) | 61.0<br>(50.2 to 73.3) | 44.0<br>(38.2 to 51.5) | 24.4<br>(20.9 to 28.7) | 19.6<br>(17.0 to 23.0) | –10.4<br>(–18.9 to –3.0) | 66.3<br>(59.6 to 72.8) |
| African Region | 13.4<br>(11.1 to 16.1) | 8.9<br>(7.4 to 10.7) | 36.8<br>(33.5 to 40.7) | 31.4<br>(28.4 to 34.7) | 5.4<br>(4.5 to 6.6) | –6.1<br>(–9.6 to –2.9) | 69.3<br>(65.6 to 72.7) |
| Region of the Americas | 12.2<br>(6.5 to 18.3) | 8.2<br>(4.3 to 12.2) | 69.2<br>(48.1 to 125.2) | 43.9<br>(30.5 to 79.5) | 25.3<br>(17.6 to 45.8) | –11.8<br>(–33.8 to 0.8) | 42.5<br>(–1.0 to 60.6) |
| Eastern Mediterranean Region | 7.0<br>(4.9 to 9.5) | 4.7<br>(3.3 to 6.3) | 61.8<br>(50.2 to 79.8) | 47.5<br>(38.3 to 61.6) | 14.3<br>(11.1 to 19.2) | –7.8<br>(–15.2 to –2.2) | 46.1<br>(30.4 to 56.6) |
| European Region | 10.3<br>(7.1 to 13.9) | 6.9<br>(4.7 to 9.3) | 30.7<br>(23.6 to 43.4) | –2.8<br>(–5.8 to –0.4) | 33.5<br>(25.9 to 47.5) | –29.3<br>(–46.1 to –19.1) | 98.6<br>(90.6 to 107.5) |
| South-East Asia Region | 29.3<br>(20.4 to 40.0) | 19.5<br>(13.6 to 26.6) | 35.7<br>(29.4 to 45.7) | 25.6<br>(21.2 to 32.6) | 10.1<br>(8.1 to 13.2) | –2.3<br>(–10.5 to 4.6) | 66.6<br>(56.7 to 74.1) |
| Western Pacific Region | 18.3<br>(7.8 to 32.0) | 12.2<br>(5.2 to 21.3) | 46.6<br>(30.0 to 99.5) | 11.9<br>(7.4 to 25.7) | 34.8<br>(22.3 to 74.3) | –16.1<br>(–83.1 to 11.8) | 69.5<br>(39.3 to 102.0) |
| Taiwan, China | 0.31<br>(–0.04 to 0.68) | 0.20<br>(–0.03 to 0.45) | — | — | — | — | — |

*Notes.* Values are medians with 95% uncertainty intervals (UIs). Forecast increase is the difference between the 2030 and 2024 counts. Regional contributions and the contribution of Taiwan, China are expressed as shares of the global target. The forecast increase was decomposed into population size ( $D_N$ ), age–sex composition ( $D_A$ ), age–sex-specific hypertension prevalence ( $D_P$ ), and control-rate improvement ( $D_C$ ). Demographic change is the sum of $D_N$ and $D_A$ . Component percentages are not reported for Taiwan, China because its 95% UI for the forecast increase includes zero. The global estimate includes the six WHO regions and Taiwan, China. Because medians summarize separate posterior distributions, regional medians and the median for Taiwan, China do not necessarily sum to the global median. Component percentages were calculated within posterior draws; therefore, the median of a combined component may differ slightly from the sum of the medians of its constituent components.
*Abbreviations.* UI, uncertainty interval; WHO, World Health Organization.

### Comparison with the matched historical benchmark

The annual increase in the number of adults with controlled hypertension was greater in the forecast period than in the matched historical benchmark, rising from 10.00 million per year (95% UI: 9.44 to 10.58 million) in 2013–2019 to 15.24 million per year (95% UI: 12.54 to 18.33 million) in 2024–2030 (Fig. 2; Table 2). Although the absolute annual demographic contribution increased from 5.24 to 6.71 million, its share of the total increase declined from 52.4% to 44.0%, a paired difference of ™8.4 percentage points (95% UI: ™14.4 to ™0.9). The population-size share decreased by 12.0 percentage points (95% UI: ™15.6 to ™7.7), whereas the age–sex-composition share increased by 3.6 percentage points (95% UI: 1.0 to 7.0). The control-rate improvement share increased by 7.4 percentage points (95% UI: 0.6 to 13.8). The corresponding change in the prevalence component was 1.0 percentage point (95% UI: ™7.4 to 8.4).

**Table 2.** Temporal comparison of the global annual increase in adults aged 30–79 years with controlled hypertension and its decomposition, 2013–2019 versus 2024–2030.

| Component of change | Annualized contribution, millions per year (95% UI) |  | Contribution to the total increase |  |  |  |
| --- | --- | --- | --- | --- | --- | --- |
|  | Historical benchmark,<br>2013–2019 | Forecast period,<br>2024–2030 | Historical benchmark,<br>2013–2019, % (95% UI) | Forecast period,<br>2024–2030, % (95% UI) | Paired difference,<br>percentage points (95%<br>UI) | Pr(forecast > historical<br>benchmark) |
| Total increase, $\Delta C$ | 10.00<br>(9.44 to 10.58) | 15.24<br>(12.54 to 18.33) | — | — | — | — |
| Population size change, $D_N$ | 3.65<br>(3.44 to 3.87) | 3.72<br>(3.41 to 4.07) | 36.5<br>(35.8 to 37.2) | 24.4<br>(20.9 to 28.7) | –12.0<br>(–15.6 to –7.7) | <0.0001 |
| Age–sex composition, $D_A$ | 1.59<br>(1.50 to 1.69) | 2.98<br>(2.77 to 3.22) | 16.0<br>(15.1 to 16.8) | 19.6<br>(17.0 to 23.0) | 3.6<br>(1.0 to 7.0) | 0.998 |
| Demographic change, $D_N + D_A$ | 5.24<br>(4.96 to 5.53) | 6.71<br>(6.25 to 7.21) | 52.4<br>(51.3 to 53.6) | 44.0<br>(38.2 to 51.5) | –8.4<br>(–14.4 to –0.9) | 0.015 |
| Age–sex-specific prevalence, $D_P$ | –1.14<br>(–1.23 to –1.05) | –1.57<br>(–2.63 to –0.50) | –11.4<br>(–12.2 to –10.6) | –10.4<br>(–18.9 to –3.0) | 1.0<br>(–7.4 to 8.4) | 0.605 |
| Control among people with<br>hypertension, $D_C$ | 5.90<br>(5.54 to 6.27) | 10.12<br>(7.86 to 12.62) | 59.0<br>(58.0 to 59.9) | 66.3<br>(59.6 to 72.8) | 7.4<br>(0.6 to 13.8) | 0.983 |

*Notes.* Values are medians with 95% uncertainty intervals (UIs). Both periods span six years. Annualized contributions are the period-specific components divided by six. Demographic change is the sum of the population-size and age–sex composition components. Component percentages were calculated within draws, so their medians may not sum exactly to 100%. Paired differences are the forecast-period fraction minus the historical-benchmark fraction. Pr(forecast > historical benchmark) is the proportion of paired draws with a positive difference and is not a frequentist p-value.
*Abbreviations.* UI, uncertainty interval; pp, percentage points; Pr, probability.

### Regional and country-level heterogeneity

The magnitude and component contributions to the forecast increase differed across WHO regions (Figs. 1, 3 and 4; Table 1). The South-East Asia Region had the largest absolute increase, at 29.3 million people, whereas the Eastern Mediterranean Region had the smallest, at 7.0 million. Demographic change accounted for 69.2% of the increase in the Region of the Americas. In contrast, population-size change contributed negatively in the European Region (™2.8%), where the control-rate improvement component accounted for 98.6% of the increase. Across regions, aggregate hypertension prevalence changed by no more than percentage points between 2019 and 2030, whereas the number of adults with controlled hypertension increased by factors ranging from 1.29 to 2.69.

**Figure 3.**
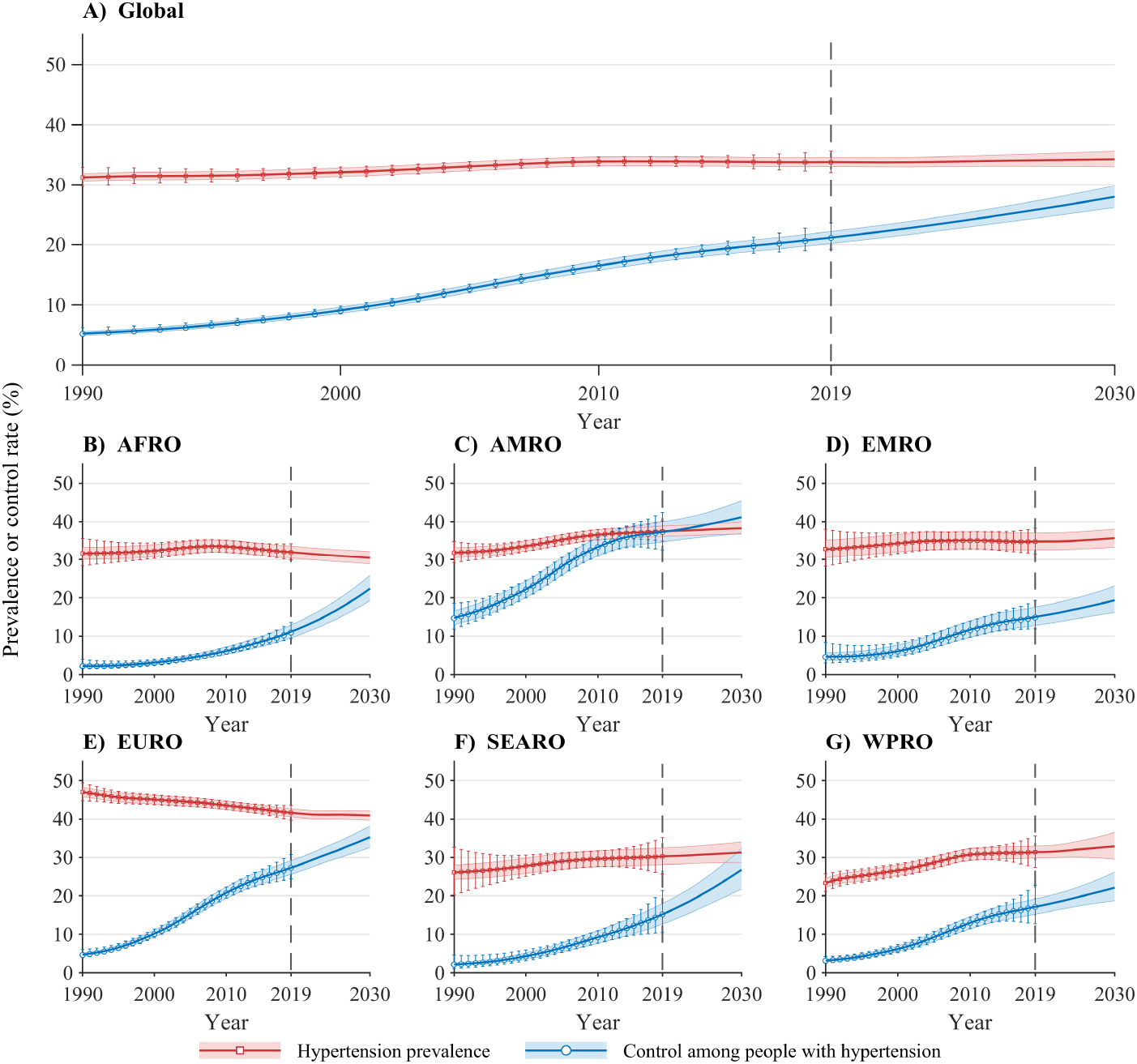
Estimated and forecast aggregate hypertension prevalence and control among adults aged 30–79 years, globally and by World Health Organization (WHO) region, 1990–2030. (A) Global. (B) African Region (AFRO). (C) Region of the Americas (AMRO). (D) Eastern Mediterranean Region (EMRO). (E) European Region (EURO). (F) South-East Asia Region (SEARO). (G) Western Pacific Region (WPRO). Open squares and error bars show annual NCD-RisC hypertension prevalence estimates and 95% uncertainty intervals (UIs) for 1990–2019. Open circles and error bars show the corresponding estimates of control among people with hypertension. Solid lines and shaded bands show model medians and 95% UIs through 2030. The vertical dashed line marks 2019, the final year of NCD-RisC estimates. Values for 2020–2030 are forecast. Prevalence is calculated among all adults aged 30–79 years, whereas control is calculated among adults with hypertension.

**Figure 4.**
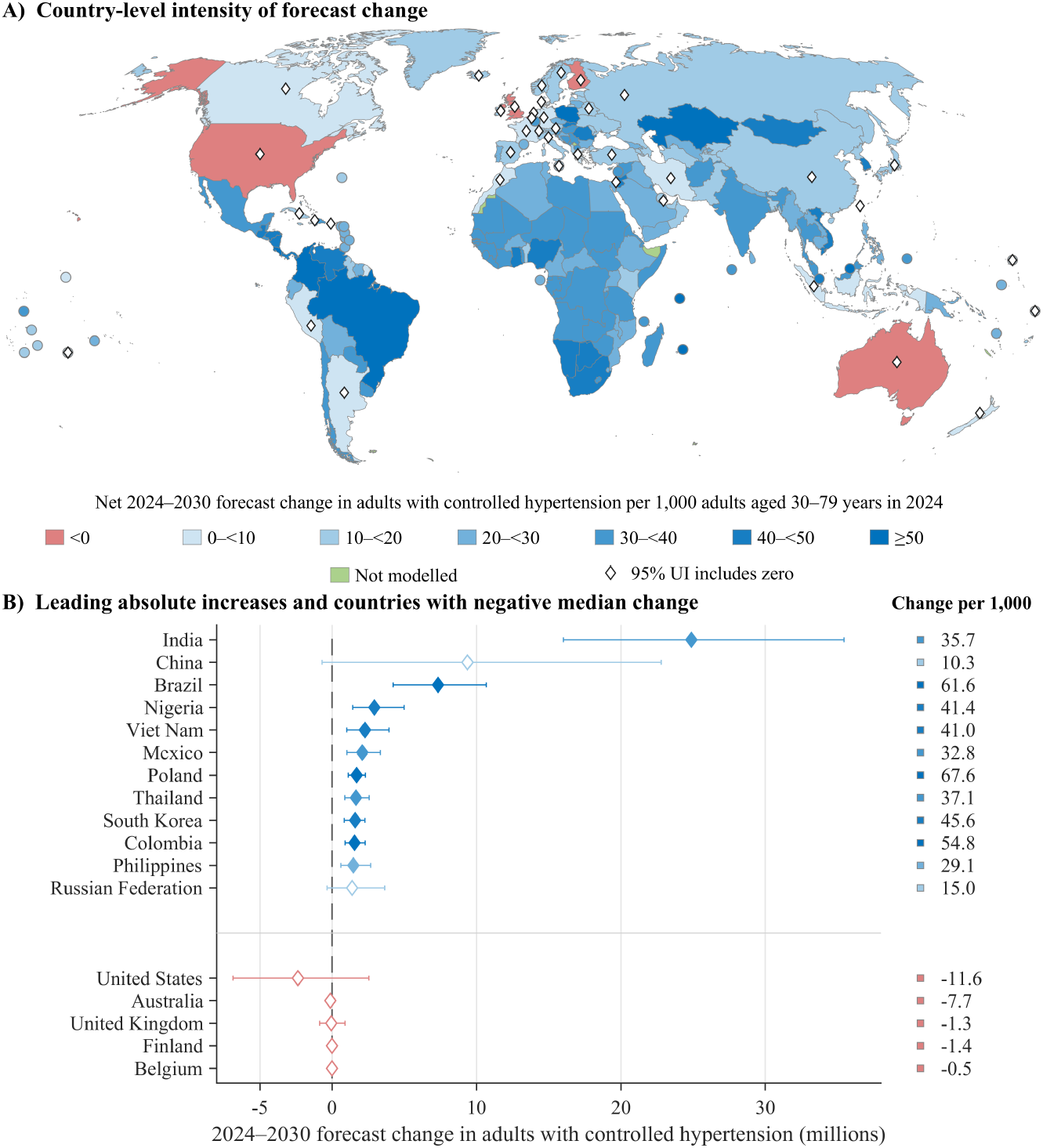
Country-level intensity and absolute contribution to the forecast increase in adults aged 30–79 years with controlled hypertension, 2024–2030. (A) Forecast change in the number of adults with controlled hypertension per 1000 adults in the 2024 population. Countries are grouped from negative change to ≥50 additional adults with controlled hypertension per 1000 adults. Open diamonds indicate countries whose 95% uncertainty interval (UI) for the absolute change includes zero. Green areas indicate countries or territories not modelled. (B) Absolute change, in millions, for the 12 largest contributors and the five countries with a negative median change. Diamonds show medians and horizontal bars show 95% UIs. Filled diamonds indicate that the 95% UI excludes zero, and open diamonds indicate that it includes zero. Colour squares correspond to the categories shown in panel A.

At country level, the 12 largest contributors accounted for 64.6% of the global posterior mean increase, with India, China and Brazil alone accounting for 46.1% of that same total (Fig. 4; Supplementary Table S4). Five countries had negative median changes, with the largest decline forecast for the United States of America, at ™2.36 million. Decomposition fractions were not reportable for 40 of the 200 countries because the 2.5th percentile of their total forecast change did not exceed zero. Country-specific observed and forecast trajectories are shown in Supplementary Figure S3.

### Sensitivity analyses

The central global forecast was stable across the six prespecified dependence scenarios, with median increases ranging from 90.49 to 92.35 million (Supplementary Fig. S2). Dependence assumptions had a larger effect on uncertainty: 95% UI widths ranged from 27.54 to 54.04 million, a 1.96-fold range. Positive between-country dependence increased interval width 1.32-fold globally and 2.04-fold in the African Region, whereas changes were small in most other regions. Alternative dependence structures therefore affected the precision of global and regional forecasts substantially more than their central estimates.

## Discussion

The principal finding of this study is that progress towards the new United Nations hypertension-control target cannot be interpreted unambiguously without an explicit baseline year. Holding the 2030 forecast constant, estimated attainment ranged from 51.4% with a 2025 baseline to 94.4% with a 2020 baseline, a 1.84-fold difference. Using the prespecified 2024 baseline, the forecast increase was 91.5 million adults with controlled hypertension, corresponding to 61.0% of the 150-million target and leaving a median shortfall of 58.5 million. These findings show that baseline specification is not a technical detail but a determinant of the headline measure used to monitor progress.

The decomposition provides a complementary and more stable interpretation of the forecast change. The control-rate improvement component accounted for 66.3% of the increase, demographic change for 44.0%, and declining age–sex-specific hypertension prevalence offset 10.4%. These component contributions varied much less across alternative baselines than the headline estimate of target attainment. The results therefore distinguish two separate questions: how much the number of adults with controlled hypertension is expected to increase, and which components account for that increase. This distinction is important because absolute gains in controlled hypertension can partly reflect population growth and changes in age–sex composition rather than improvements in hypertension control alone.

These findings extend evidence that favorable age-specific epidemiological trends can coexist with rising absolute health-care needs. Earlier global projections among adults aged 20 years or older showed that population growth and ageing could increase hypertension counts even when age- and sex-specific prevalence was held constant [7]. In six middle-income countries, demographic change alone was projected to add 319.7 million adults needing hypertension care by 2050 if age-specific prevalence remained unchanged. Importantly, treated individuals whose blood pressure was controlled were excluded from the population considered in need of care, so that outcome is not directly equivalent to ours [8]. Decomposition studies likewise show that population growth and ageing can offset improvements in age-specific rates [16–18]. A similar demographic offset has been reported across the Americas [19]. Our analysis extends this demographic logic to the number of people with controlled hypertension.

Compared with 2013–2019, the forecast period showed faster annual growth and a larger relative contribution from the control-rate improvement component. This component was substantial, but as an accounting component it does not identify why control changes. Large international differences in diagnosis, treatment and control remain despite improvements in several countries [10, 20, 21]. Modelling suggests that faster progress across the hypertension care cascade could avert substantial cardiovascular morbidity and mortality [22, 23]. In four middle-income countries, treatment discontinuation and subsequent loss of control were frequent [24]. Previous studies provided forecasts of global hypertension prevalence [25] and national care-cascade progress towards WHO 80–80–80 targets for 2030 [26]. We evaluated the UN global absolute-count target. Nevertheless, continuation of current forecast trajectories would still be insufficient to attain the new UN target.

Resolution A/RES/80/117 specifies 150 million additional people with hypertension under control by 2030 but does not define the year from which the increase should be measured [4]. We selected 2024 because it was the last completed calendar year entirely preceding adoption of the resolution. This is a defensible policy-temporal reference, but not the only possible choice. Quantitative monitoring therefore requires an explicitly defined and consistently applied baseline.

Regional heterogeneity further shows that a single global percentage conceals different pathways. Demographic forces dominated in some regions, whereas the control-rate improvement component predominated in others, consistent with large international disparities in hypertension control and health-system capacity [27, 28]. Almost two thirds of the global forecast increase was concentrated in 12 countries.

Several limitations should be considered. The NCD-RisC input series are themselves model-derived, temporally smoothed estimates rather than direct annual observations. Our forecasts therefore inherit uncertainty and structural assumptions from those estimates [10]. NCD-RisC estimates end in 2019, so all candidate baselines and the entire 2020–2030 period are model-based forecasts. Independent post-2019 NCD-RisC estimates were unavailable for validation, and a full-country rolling-origin validation was not performed for the global production model. Long-horizon calibration therefore remains uncertain. The coronavirus disease 2019 (COVID-19) pandemic may also have disrupted pre-pandemic trajectories, as reduced blood-pressure monitoring and worsening or flattening of control were documented during the pandemic [29, 30]. Population projections were treated as deterministic, and prevalence–control and between-country dependence could not be estimated from publicly available marginal uncertainty summaries. Finally, the model does not explicitly incorporate future policy changes, treatment innovations, health-system disruptions or accelerated control efforts undertaken to meet the UN target.

Overall, current trajectories imply substantial growth in the number of adults with controlled hypertension by 2030, driven principally by the control-rate improvement component but also shaped by population growth and ageing. Under the prespecified 2024 baseline, these trajectories would achieve about three fifths of the UN target. Explicit specification of the target baseline and further acceleration of hypertension control are therefore important for credible monitoring and for achieving sufficient absolute gains in a growing and ageing global population. Future research should update these forecasts as post-2019 population-representative estimates become available and assess how alternative policy and treatment trajectories could alter progress towards the 2030 target.

## Data Availability

All data used in this study are publicly available. Age- and sex-specific hypertension prevalence and control estimates were obtained from the NCD Risk Factor Collaboration (NCD-RisC), population estimates and projections from the United Nations World Population Prospects 2024, and cartographic boundaries from Natural Earth. The code used for model fitting, assembly, and decomposition is publicly available at Zenodo: https://doi.org/10.5281/zenodo.22278513.

https://ncdrisc.org/

https://population.un.org/wpp/

https://www.naturalearthdata.com/downloads/50m-cultural-vectors/

https://doi.org/10.5281/zenodo.22278513

## Acknowledgements

The authors acknowledge the Community University of Chapecó Region (Unochapecó), the Coordination for the Improvement of Higher Education Personnel (CAPES), the National Council for Scientific and Technological Development (CNPq), and the Foundation for Research and Innovation Support of the State of Santa Catarina (FAPESC) for their general support of postgraduate education and research in Brazil. No specific funding was received for this study.

## Data and code availability

All data used in this study are publicly available. Age- and sex-specific hypertension prevalence and control estimates were obtained from the NCD Risk Factor Collaboration [31], and population estimates and projections from the United Nations World Population Prospects 2024 [32]. Cartographic boundaries were obtained from Natural Earth 1:50m cultural vectors, version 5.1.1 [33]. The code for model fitting, assembly, and decomposition is available at Zenodo [34].

## Supplementary material

The supplementary material accompanying this manuscript includes Supplementary Methods S5, providing additional details on geographical classification and display procedures, Bayesian model specification and computation, uncertainty and dependence assumptions, decomposition implementation, and model assessment; Supplementary Figure S1, showing sensitivity of target attainment to the analytical baseline year; Supplementary Figure S2, showing sensitivity to alternative dependence assumptions; Supplementary Figure S3, showing country-specific observed and forecast trajectories; and Supplementary Table S4, providing country-level results.

