## Supplementary Figure S1 for "Forecasting global progress towards the United Nations hypertension control target for 2030"

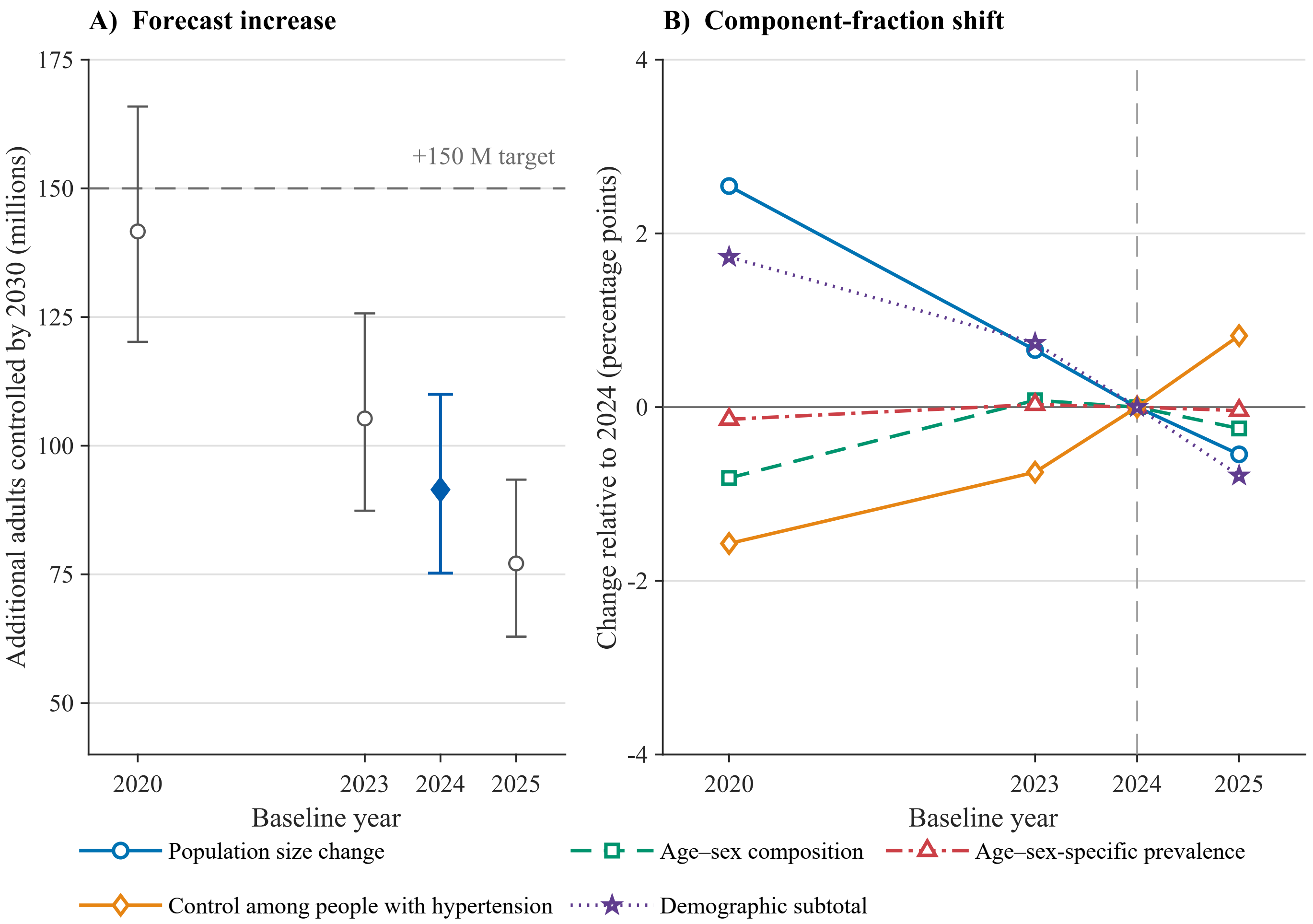


**Supplementary Figure S1. Sensitivity of the global forecast and decomposition to the baseline year.** (A) Forecast increase in the number of adults aged 30–79 years with controlled hypertension from each alternative baseline year (2020, 2023, 2024, and 2025) to 2030. Points and error bars show posterior medians and 95% uncertainty intervals; the filled marker identifies the primary 2024 baseline. The horizontal dashed line marks the target of 150 million additional adults with controlled hypertension. (B) Differences in the median fraction attributed to each Horiuchi component relative to the primary 2024 baseline, expressed in percentage points. These are descriptive differences between marginal posterior medians. Paired uncertainty intervals across baseline years were not produced in the frozen analytical output, although draw-by-draw differences are defined because all four baseline analyses derive from the same posterior. Across the four baselines, the ranges of the component medians were 3.09 percentage points for population size change ($D_N$), 2.52 for the demographic subtotal ($D_{\text{demo}}$), 2.39 for control among people with hypertension ($D_C$), 0.89 for age–sex composition ($D_A$), and 0.17 for age–sex-specific prevalence ($D_P$). Estimates use CORE-P25 and the prespecified main dependence assumptions.
