## Supplementary Figure S2 for "Forecasting global progress towards the United Nations hypertension control target for 2030"

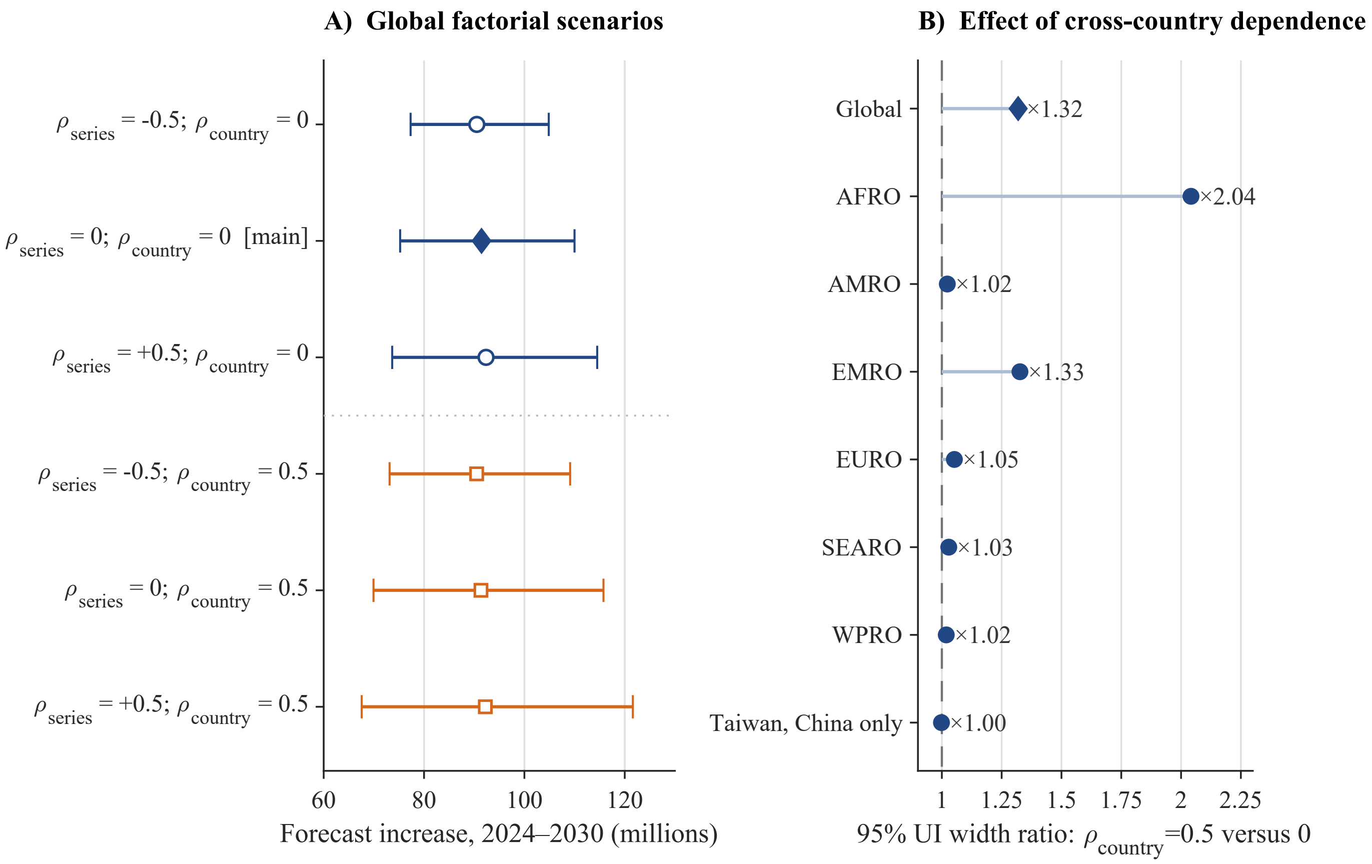


**Supplementary Figure S2. Sensitivity of the global forecast to the prespecified dependence assumptions.** (A) Forecast increase in the number of adults aged 30–79 years with controlled hypertension between 2024 and 2030 under six factorial scenarios crossing $\rho_{series} \in \{-0.5, 0, +0.5\}$ with $\rho_{country} \in \{0, 0.5\}$. Points and horizontal bars show posterior medians and 95% uncertainty intervals; the filled diamond identifies the main scenario ($\rho_{series} = 0$, $\rho_{country} = 0$). Global medians varied from 90.49 to 92.35 million, whereas uncertainty-interval widths varied from 27.54 to 54.04 million, a 1.96-fold range. (B) Ratio of the 95% uncertainty-interval width under $\rho_{country} = 0.5$ to that under the main assumption $\rho_{country} = 0$, holding $\rho_{series} = 0$. The vertical dashed line denotes a ratio of 1.00. Ratios were 1.32 globally, 2.04 in AFRO, 1.02 in AMRO, 1.33 in EMRO, 1.05 in EURO, 1.03 in SEARO, and 1.02 in WPRO. UNCLASSIFIED contains Taiwan, China only; its ratio of 1.00 (0.998 before rounding) is the expected negative control for cross-country dependence, and the departure from unity is below the Monte Carlo resolution of the estimate. Interval width is defined as the upper minus the lower limit. All estimates use CORE-P25 and the 2024–2030 window.
