## Supplementary Figure S3 for "Forecasting global progress towards the United Nations hypertension control target for 2030"

### Supplementary Figure S3. Country atlas of aggregate hypertension prevalence and control, 1990–2030

Countries are ordered alphabetically using the WHO display names applied in Supplementary Table S4. Each entry gives country [ISO3 · WHO region · atlas page].


Afghanistan [AFG · EMRO · 2]
Albania [ALB · EURO · 2]
Algeria [DZA · AFRO · 2]
American Samoa [ASM · WPRO · 2]
Andorra [AND · EURO · 3]
Angola [AGO · AFRO · 3]
Antigua and Barbuda [ATG · AMRO · 3]
Argentina [ARG · AMRO · 3]
Armenia [ARM · EURO · 4]
Australia [AUS · WPRO · 4]
Austria [AUT · EURO · 4]
Azerbaijan [AZE · EURO · 4]
Bahamas [BHS · AMRO · 5]
Bahrain [BHR · EMRO · 5]
Bangladesh [BGD · SEARO · 5]
Barbados [BRB · AMRO · 5]
Belarus [BLR · EURO · 6]
Belgium [BEL · EURO · 6]
Belize [BLZ · AMRO · 6]
Benin [BEN · AFRO · 6]
Bermuda [BMU · AMRO · 7]
Bhutan [BTN · SEARO · 7]
Bolivia [BOL · AMRO · 7]
Bosnia and Herzegovina [BIH · EURO · 7]
Botswana [BWA · AFRO · 8]
Brazil [BRA · AMRO · 8]
Brunei Darussalam [BRN · WPRO · 8]
Bulgaria [BGR · EURO · 8]
Burkina Faso [BFA · AFRO · 9]
Burundi [BDI · AFRO · 9]
Cabo Verde [CPV · AFRO · 9]
Cambodia [KHM · WPRO · 9]
Cameroon [CMR · AFRO · 10]
Canada [CAN · AMRO · 10]
Central African Republic [CAF · AFRO · 10]
Chad [TCD · AFRO · 10]
Chile [CHL · AMRO · 11]
China [CHN · WPRO · 11]
Colombia [COL · AMRO · 11]
Comoros [COM · AFRO · 11]
Congo [COG · AFRO · 12]
Cook Islands [COK · WPRO · 12]
Costa Rica [CRI · AMRO · 12]
Côte d'Ivoire [CIV · AFRO · 12]
Croatia [HRV · EURO · 13]
Cuba [CUB · AMRO · 13]
Cyprus [CYP · EURO · 13]
Czechia [CZE · EURO · 13]
Democratic People's Republic of Korea [PRK · SEARO · 14]
Democratic Republic of the Congo [COD · AFRO · 14]
Denmark [DNK · EURO · 14]
Djibouti [DJI · EMRO · 14]
Dominica [DMA · AMRO · 15]
Dominican Republic [DOM · AMRO · 15]
Ecuador [ECU · AMRO · 15]
Egypt [EGY · EMRO · 15]
El Salvador [SLV · AMRO · 16]
Equatorial Guinea [GNQ · AFRO · 16]
Eritrea [ERI · AFRO · 16]
Estonia [EST · EURO · 16]
Eswatini [SWZ · AFRO · 17]
Ethiopia [ETH · AFRO · 17]
Fiji [FJI · WPRO · 17]
Finland [FIN · EURO · 17]
France [FRA · EURO · 18]
French Polynesia [PYF · WPRO · 18]
Gabon [GAB · AFRO · 18]
Gambia [GMB · AFRO · 18]
Georgia [GEO · EURO · 19]
Germany [DEU · EURO · 19]
Ghana [GHA · AFRO · 19]
Greece [GRC · EURO · 19]
Greenland [GRL · EURO · 20]
Grenada [GRD · AMRO · 20]
Guatemala [GTM · AMRO · 20]
Guinea [GIN · AFRO · 20]
Guinea Bissau [GNB · AFRO · 21]
Guyana [GUY · AMRO · 21]
Haiti [HTI · AMRO · 21]
Honduras [HND · AMRO · 21]
Hungary [HUN · EURO · 22]
Iceland [ISL · EURO · 22]
India [IND · SEARO · 22]
Indonesia [IDN · WPRO · 22]
Iran (Islamic Republic of) [IRN · EMRO · 23]
Iraq [IRQ · EMRO · 23]
Ireland [IRL · EURO · 23]
Israel [ISR · EURO · 23]
Italy [ITA · EURO · 24]
Jamaica [JAM · AMRO · 24]
Japan [JPN · WPRO · 24]
Jordan [JOR · EMRO · 24]
Kazakhstan [KAZ · EURO · 25]
Kenya [KEN · AFRO · 25]
Kiribati [KIR · WPRO · 25]
Kuwait [KWT · EMRO · 25]
Kyrgyzstan [KGZ · EURO · 26]
Lao People's Democratic Republic [LAO · WPRO · 26]
Latvia [LVA · EURO · 26]
Lebanon [LBN · EMRO · 26]
Lesotho [LSO · AFRO · 27]
Liberia [LBR · AFRO · 27]
Libya [LBY · EMRO · 27]
Lithuania [LTU · EURO · 27]
Luxembourg [LUX · EURO · 28]
Madagascar [MDG · AFRO · 28]
Malawi [MWI · AFRO · 28]
Malaysia [MYS · WPRO · 28]
Maldives [MDV · SEARO · 29]
Mali [MLI · AFRO · 29]
Malta [MLT · EURO · 29]
Marshall Islands [MHL · WPRO · 29]
Mauritania [MRT · AFRO · 30]
Mauritius [MUS · AFRO · 30]
Mexico [MEX · AMRO · 30]
Micronesia (Federated States of) [FSM · WPRO · 30]
Moldova [MDA · EURO · 31]
Mongolia [MNG · WPRO · 31]
Montenegro [MNE · EURO · 31]
Morocco [MAR · EMRO · 31]
Mozambique [MOZ · AFRO · 32]
Myanmar [MMR · SEARO · 32]
Namibia [NAM · AFRO · 32]
Nauru [NRU · WPRO · 32]
Nepal [NPL · SEARO · 33]
Netherlands [NLD · EURO · 33]
New Zealand [NZL · WPRO · 33]
Nicaragua [NIC · AMRO · 33]
Niger [NER · AFRO · 34]
Nigeria [NGA · AFRO · 34]
Niue [NIU · WPRO · 34]
North Macedonia [MKD · EURO · 34]
Norway [NOR · EURO · 35]
occupied Palestinian territory [PSE · EMRO · 35]
Oman [OMN · EMRO · 35]
Pakistan [PAK · EMRO · 35]
Palau [PLW · WPRO · 36]
Panama [PAN · AMRO · 36]
Papua New Guinea [PNG · WPRO · 36]
Paraguay [PRY · AMRO · 36]
Peru [PER · AMRO · 37]
Philippines [PHL · WPRO · 37]
Poland [POL · EURO · 37]
Portugal [PRT · EURO · 37]
Puerto Rico [PRI · AMRO · 38]
Qatar [QAT · EMRO · 38]
Republic of Korea [KOR · WPRO · 38]
Romania [ROU · EURO · 38]
Russian Federation [RUS · EURO · 39]
Rwanda [RWA · AFRO · 39]
Saint Kitts and Nevis [KNA · AMRO · 39]
Saint Lucia [LCA · AMRO · 39]
Saint Vincent and the Grenadines [VCT · AMRO · 40]
Samoa [WSM · WPRO · 40]
Sao Tome and Principe [STP · AFRO · 40]
Saudi Arabia [SAU · EMRO · 40]
Senegal [SEN · AFRO · 41]
Serbia [SRB · EURO · 41]
Seychelles [SYC · AFRO · 41]
Sierra Leone [SLE · AFRO · 41]
Singapore [SGP · WPRO · 42]
Slovakia [SVK · EURO · 42]
Slovenia [SVN · EURO · 42]
Solomon Islands [SLB · WPRO · 42]
Somalia [SOM · EMRO · 43]
South Africa [ZAF · AFRO · 43]
South Sudan [SSD · AFRO · 43]
Spain [ESP · EURO · 43]
Sri Lanka [LKA · SEARO · 44]
Sudan [SDN · EMRO · 44]
Suriname [SUR · AMRO · 44]
Sweden [SWE · EURO · 44]
Switzerland [CHE · EURO · 45]
Syrian Arab Republic [SYR · EMRO · 45]
Taiwan, China [TWN · UNCLASSIFIED · 45]
Tajikistan [TJK · EURO · 45]
Tanzania [TZA · AFRO · 46]
Thailand [THA · SEARO · 46]
Timor-Leste [TLS · SEARO · 46]
Togo [TGO · AFRO · 46]
Tokelau [TKL · WPRO · 47]
Tonga [TON · WPRO · 47]
Trinidad and Tobago [TTO · AMRO · 47]
Tunisia [TUN · EMRO · 47]
Türkiye [TUR · EURO · 48]
Turkmenistan [TKM · EURO · 48]
Tuvalu [TUV · WPRO · 48]
Uganda [UGA · AFRO · 48]
Ukraine [UKR · EURO · 49]
United Arab Emirates [ARE · EMRO · 49]
United Kingdom [GBR · EURO · 49]
United States of America [USA · AMRO · 49]
Uruguay [URY · AMRO · 50]
Uzbekistan [UZB · EURO · 50]
Vanuatu [VUT · WPRO · 50]
Venezuela [VEN · AMRO · 50]
Viet Nam [VNM · WPRO · 51]
Yemen [YEM · EMRO · 51]
Zambia [ZMB · AFRO · 51]
Zimbabwe [ZWE · AFRO · 51]

**Supplementary Figure S3. Country-level trajectories of aggregate hypertension prevalence and control, 1990–2030**
Atlas panel page 1 of 50; countries 1–4 of 200.

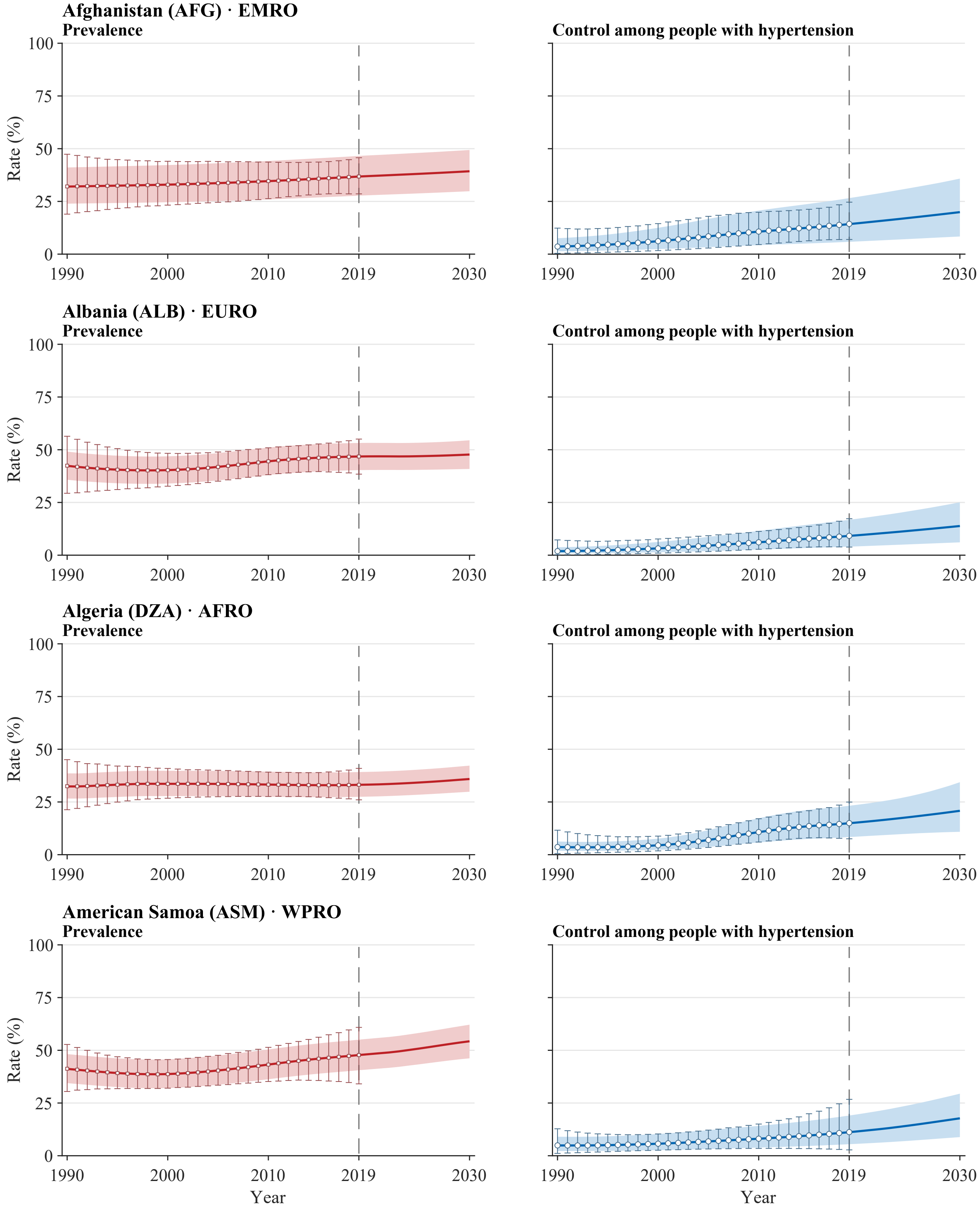


**Legend.** Open squares and error bars in prevalence panels and open circles and error bars in control panels show annual NCD-RisC central estimates with propagated 95% uncertainty intervals for 1990–2019. Solid red and blue lines and shaded bands show model medians and 95% uncertainty intervals for 1990–2030. The vertical dashed line marks 2019, the final year with reported estimates; values for 2020–2030 are forecast. Latent stratum-specific values in 2019 are anchored to the published central estimates. Propagated intervals for the published estimates are wider than the model bands during the historical period by construction of the external uncertainty layer. Model uncertainty represents a persistent level offset. All panels use a common 0–100% scale. Prevalence is defined among all adults aged 30–79 years, whereas control is defined among adults with hypertension. Estimates use CORE-P25 and the prespecified main dependence assumptions.

**Supplementary Figure S3. Country-level trajectories of aggregate hypertension prevalence and control, 1990–2030**
Atlas panel page 2 of 50; countries 5–8 of 200.

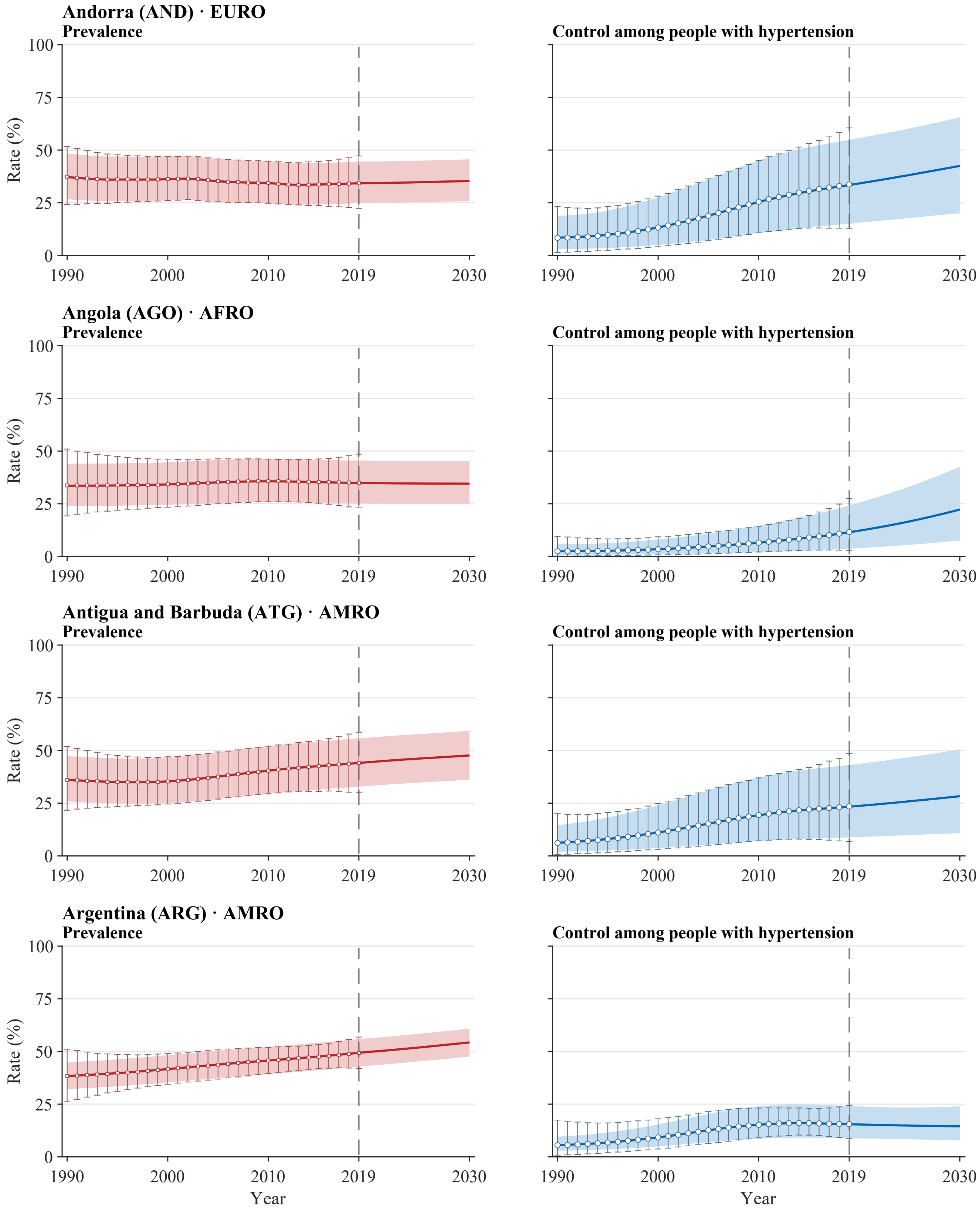


**Legend.** Open squares and error bars in prevalence panels and open circles and error bars in control panels show annual NCD-RisC central estimates with propagated 95% uncertainty intervals for 1990–2019. Solid red and blue lines and shaded bands show model medians and 95% uncertainty intervals for 1990–2030. The vertical dashed line marks 2019, the final year with reported estimates; values for 2020–2030 are forecast. Latent stratum-specific values in 2019 are anchored to the published central estimates. Propagated intervals for the published estimates are wider than the model bands during the historical period by construction of the external uncertainty layer. Model uncertainty represents a persistent level offset. All panels use a common 0–100% scale. Prevalence is defined among all adults aged 30–79 years, whereas control is defined among adults with hypertension. Estimates use CORE-P25 and the prespecified main dependence assumptions.

**Supplementary Figure S3. Country-level trajectories of aggregate hypertension prevalence and control, 1990–2030**
Atlas panel page 3 of 50; countries 9–12 of 200.

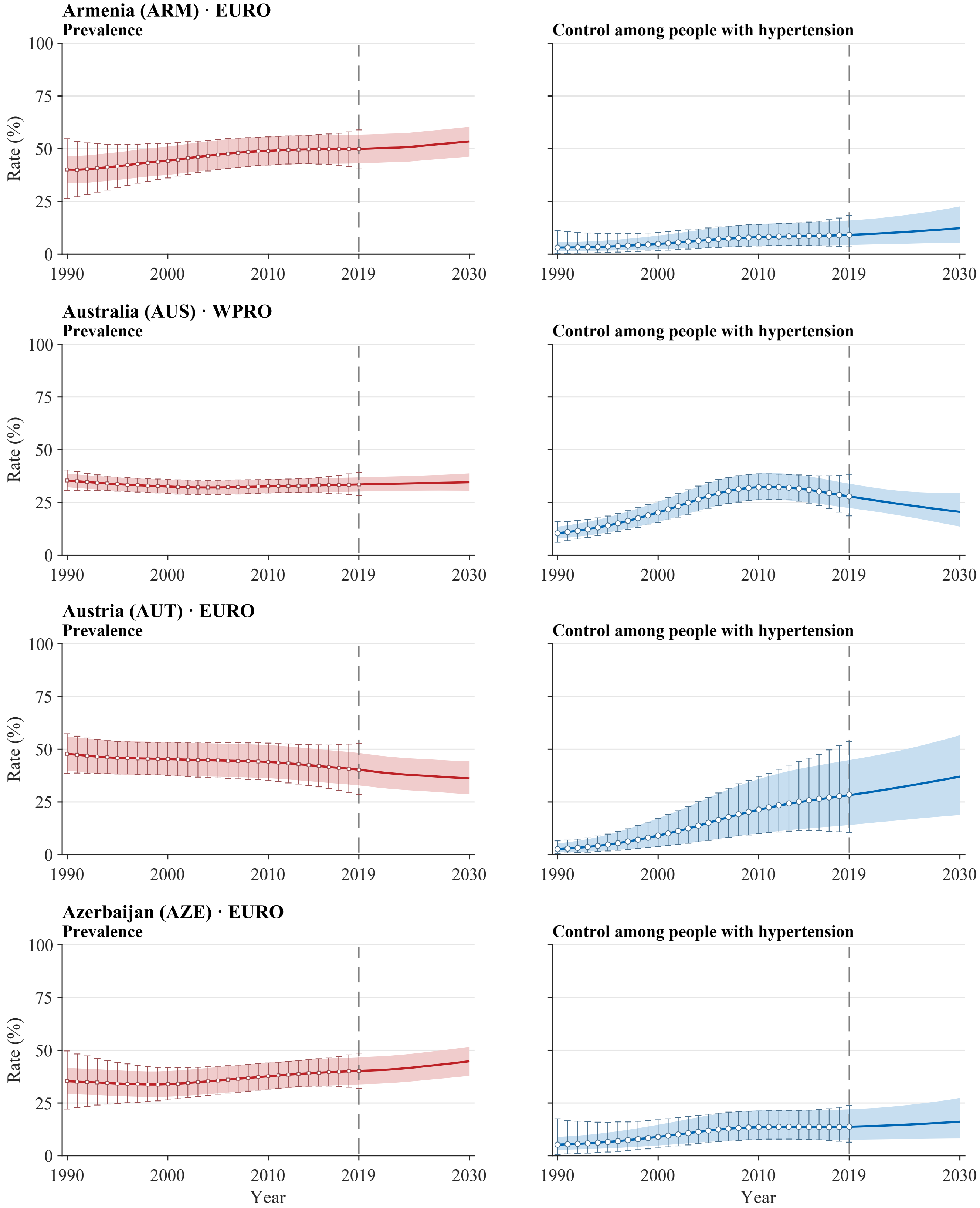


**Legend.** Open squares and error bars in prevalence panels and open circles and error bars in control panels show annual NCD-RisC central estimates with propagated 95% uncertainty intervals for 1990–2019. Solid red and blue lines and shaded bands show model medians and 95% uncertainty intervals for 1990–2030. The vertical dashed line marks 2019, the final year with reported estimates; values for 2020–2030 are forecast. Latent stratum-specific values in 2019 are anchored to the published central estimates. Propagated intervals for the published estimates are wider than the model bands during the historical period by construction of the external uncertainty layer. Model uncertainty represents a persistent level offset. All panels use a common 0–100% scale. Prevalence is defined among all adults aged 30–79 years, whereas control is defined among adults with hypertension. Estimates use CORE-P25 and the prespecified main dependence assumptions.

**Supplementary Figure S3. Country-level trajectories of aggregate hypertension prevalence and control, 1990–2030**

Atlas panel page 4 of 50; countries 13–16 of 200.

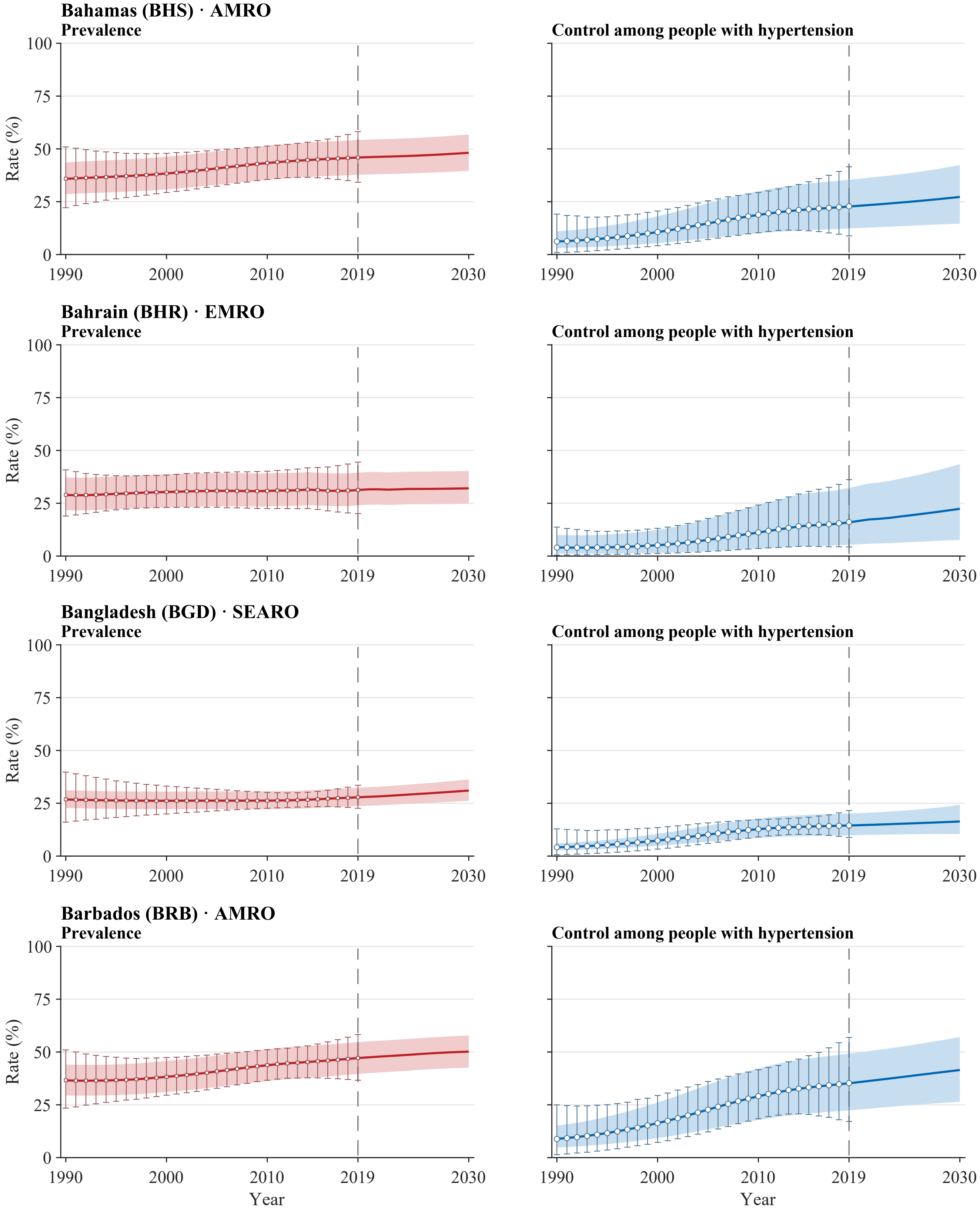


**Legend.** Open squares and error bars in prevalence panels and open circles and error bars in control panels show annual NCD-RisC central estimates with propagated 95% uncertainty intervals for 1990–2019. Solid red and blue lines and shaded bands show model medians and 95% uncertainty intervals for 1990–2030. The vertical dashed line marks 2019, the final year with reported estimates; values for 2020–2030 are forecast. Latent stratum-specific values in 2019 are anchored to the published central estimates. Propagated intervals for the published estimates are wider than the model bands during the historical period by construction of the external uncertainty layer. Model uncertainty represents a persistent level offset. All panels use a common 0–100% scale. Prevalence is defined among all adults aged 30–79 years, whereas control is defined among adults with hypertension. Estimates use CORE-P25 and the prespecified main dependence assumptions.

**Supplementary Figure S3. Country-level trajectories of aggregate hypertension prevalence and control, 1990–2030**
Atlas panel page 5 of 50; countries 17–20 of 200.

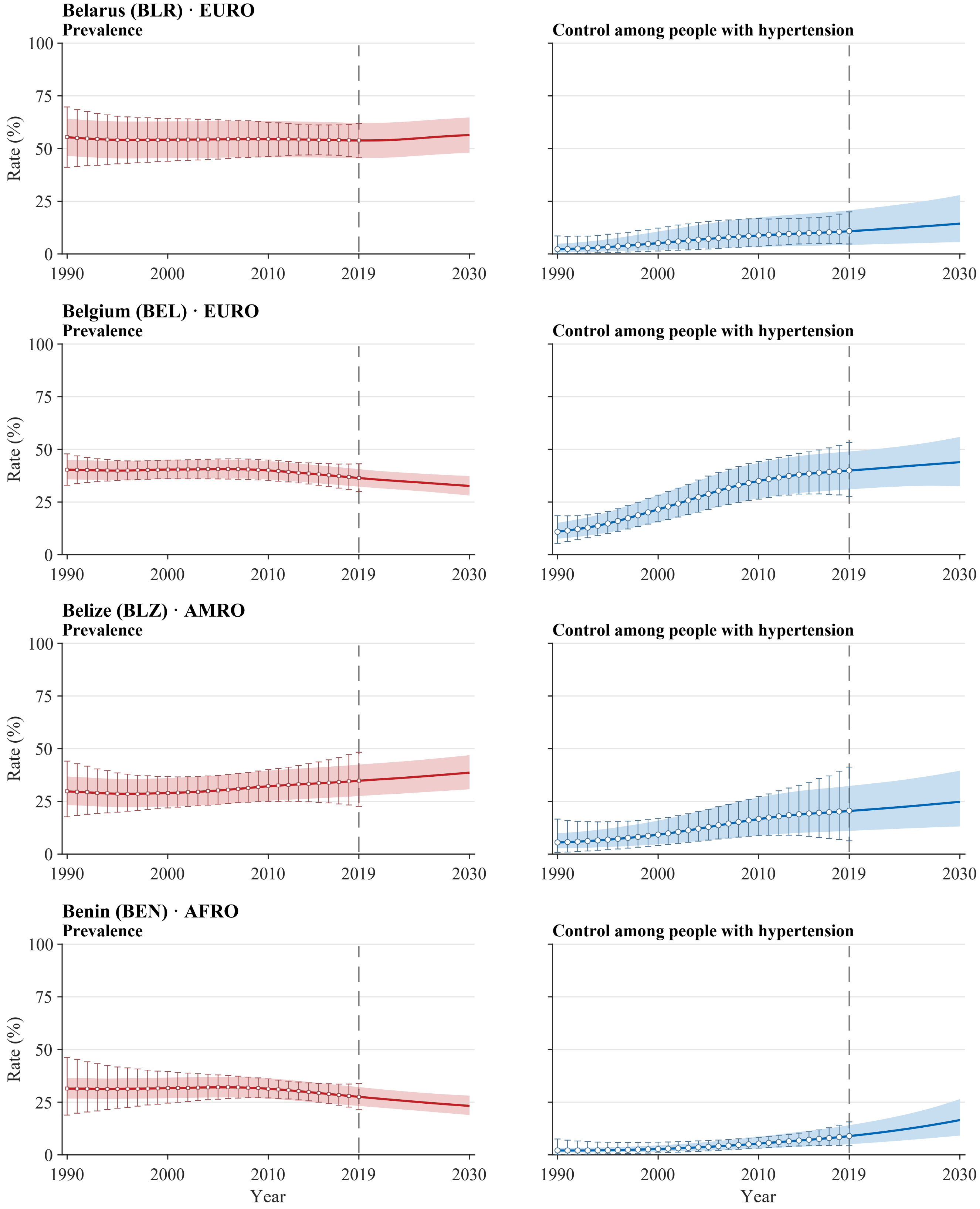


**Legend.** Open squares and error bars in prevalence panels and open circles and error bars in control panels show annual NCD-RisC central estimates with propagated 95% uncertainty intervals for 1990–2019. Solid red and blue lines and shaded bands show model medians and 95% uncertainty intervals for 1990–2030. The vertical dashed line marks 2019, the final year with reported estimates; values for 2020–2030 are forecast. Latent stratum-specific values in 2019 are anchored to the published central estimates. Propagated intervals for the published estimates are wider than the model bands during the historical period by construction of the external uncertainty layer. Model uncertainty represents a persistent level offset. All panels use a common 0–100% scale. Prevalence is defined among all adults aged 30–79 years, whereas control is defined among adults with hypertension. Estimates use CORE-P25 and the prespecified main dependence assumptions.

**Supplementary Figure S3. Country-level trajectories of aggregate hypertension prevalence and control, 1990–2030**
Atlas panel page 6 of 50; countries 21–24 of 200.

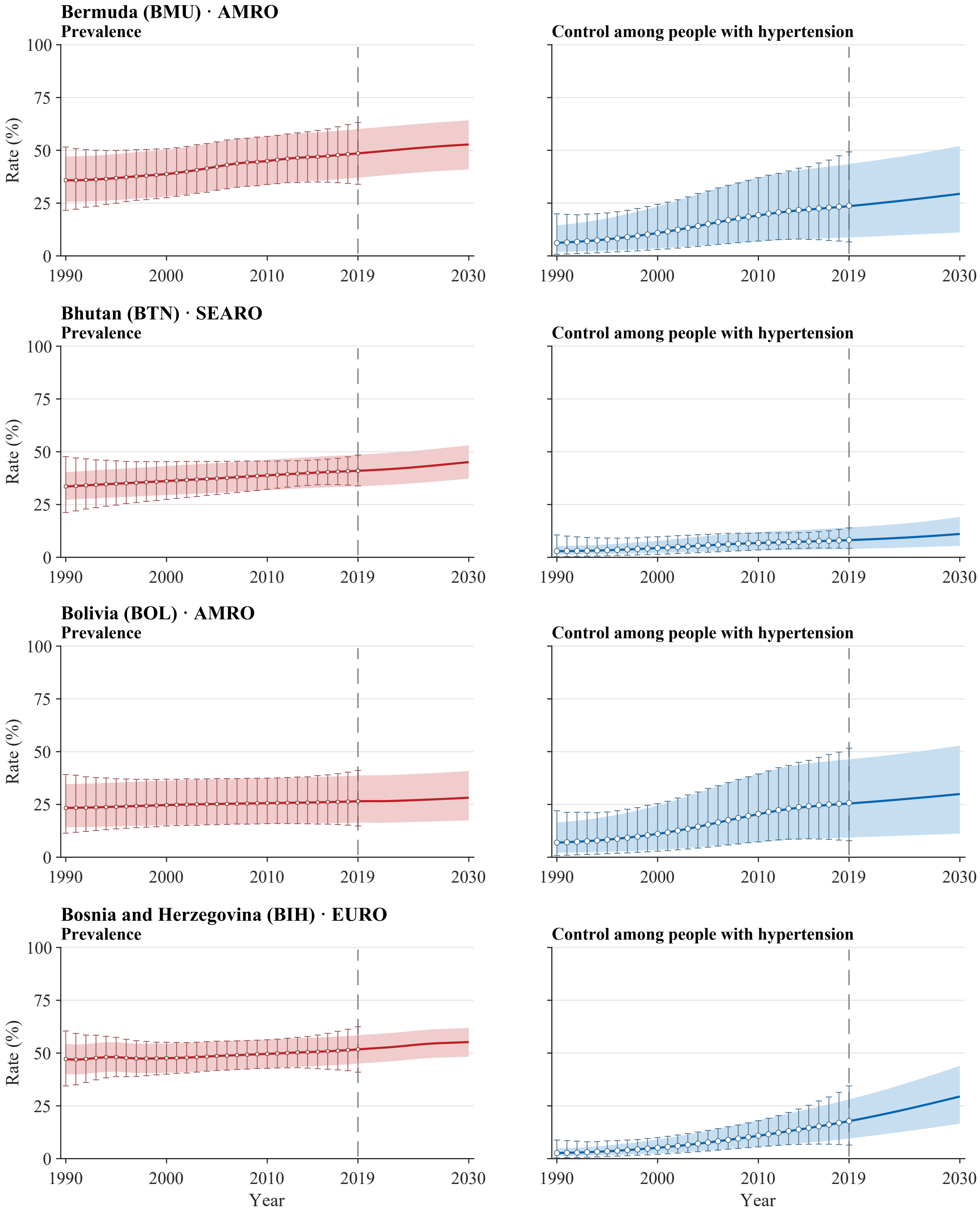


**Legend.** Open squares and error bars in prevalence panels and open circles and error bars in control panels show annual NCD-RisC central estimates with propagated 95% uncertainty intervals for 1990–2019. Solid red and blue lines and shaded bands show model medians and 95% uncertainty intervals for 1990–2030. The vertical dashed line marks 2019, the final year with reported estimates; values for 2020–2030 are forecast. Latent stratum-specific values in 2019 are anchored to the published central estimates. Propagated intervals for the published estimates are wider than the model bands during the historical period by construction of the external uncertainty layer. Model uncertainty represents a persistent level offset. All panels use a common 0–100% scale. Prevalence is defined among all adults aged 30–79 years, whereas control is defined among adults with hypertension. Estimates use CORE-P25 and the prespecified main dependence assumptions.

**Supplementary Figure S3. Country-level trajectories of aggregate hypertension prevalence and control, 1990–2030**
Atlas panel page 7 of 50; countries 25–28 of 200.

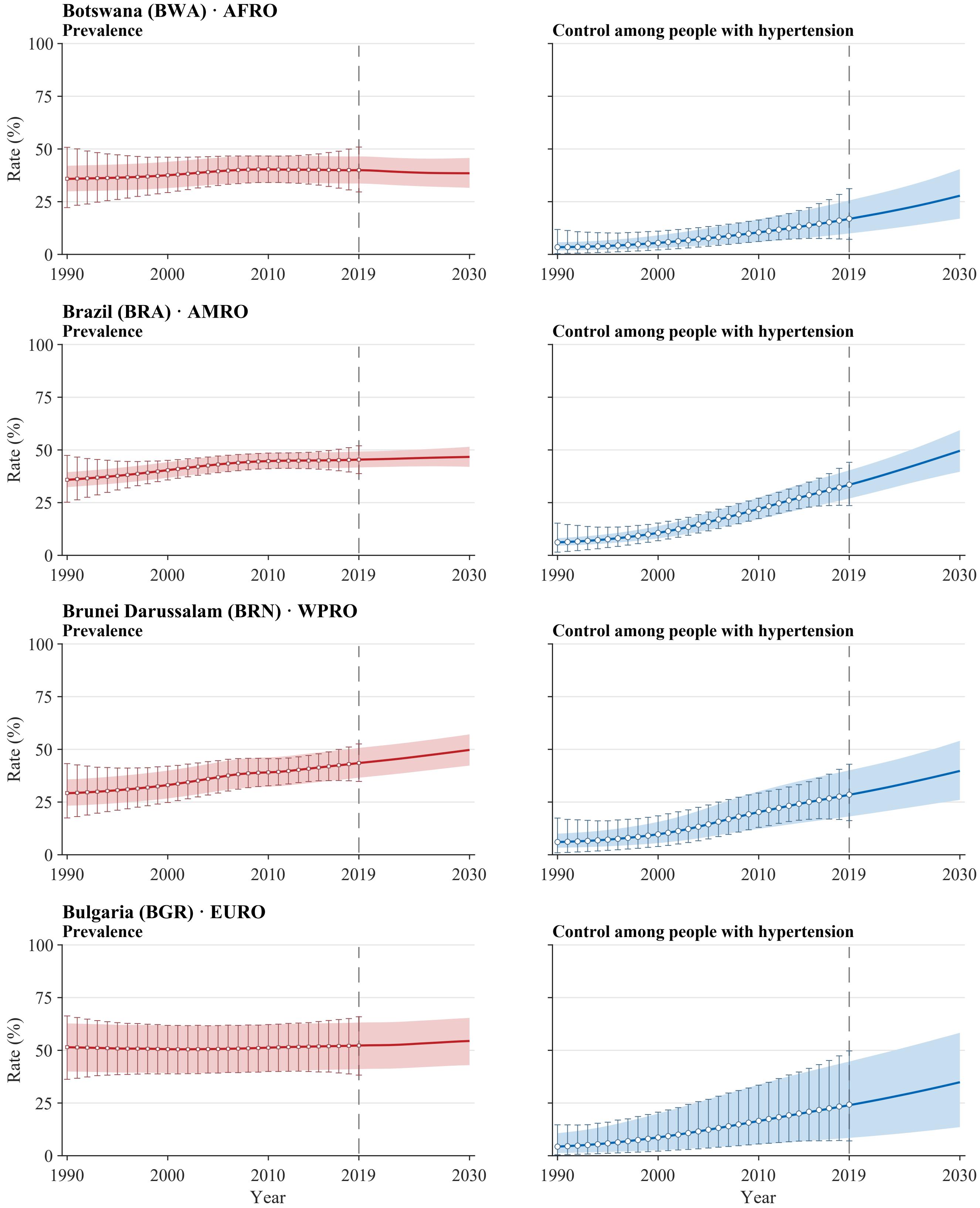


**Legend.** Open squares and error bars in prevalence panels and open circles and error bars in control panels show annual NCD-RisC central estimates with propagated 95% uncertainty intervals for 1990–2019. Solid red and blue lines and shaded bands show model medians and 95% uncertainty intervals for 1990–2030. The vertical dashed line marks 2019, the final year with reported estimates; values for 2020–2030 are forecast. Latent stratum-specific values in 2019 are anchored to the published central estimates. Propagated intervals for the published estimates are wider than the model bands during the historical period by construction of the external uncertainty layer. Model uncertainty represents a persistent level offset. All panels use a common 0–100% scale. Prevalence is defined among all adults aged 30–79 years, whereas control is defined among adults with hypertension. Estimates use CORE-P25 and the prespecified main dependence assumptions.

**Supplementary Figure S3. Country-level trajectories of aggregate hypertension prevalence and control, 1990–2030**

Atlas panel page 8 of 50; countries 29–32 of 200.

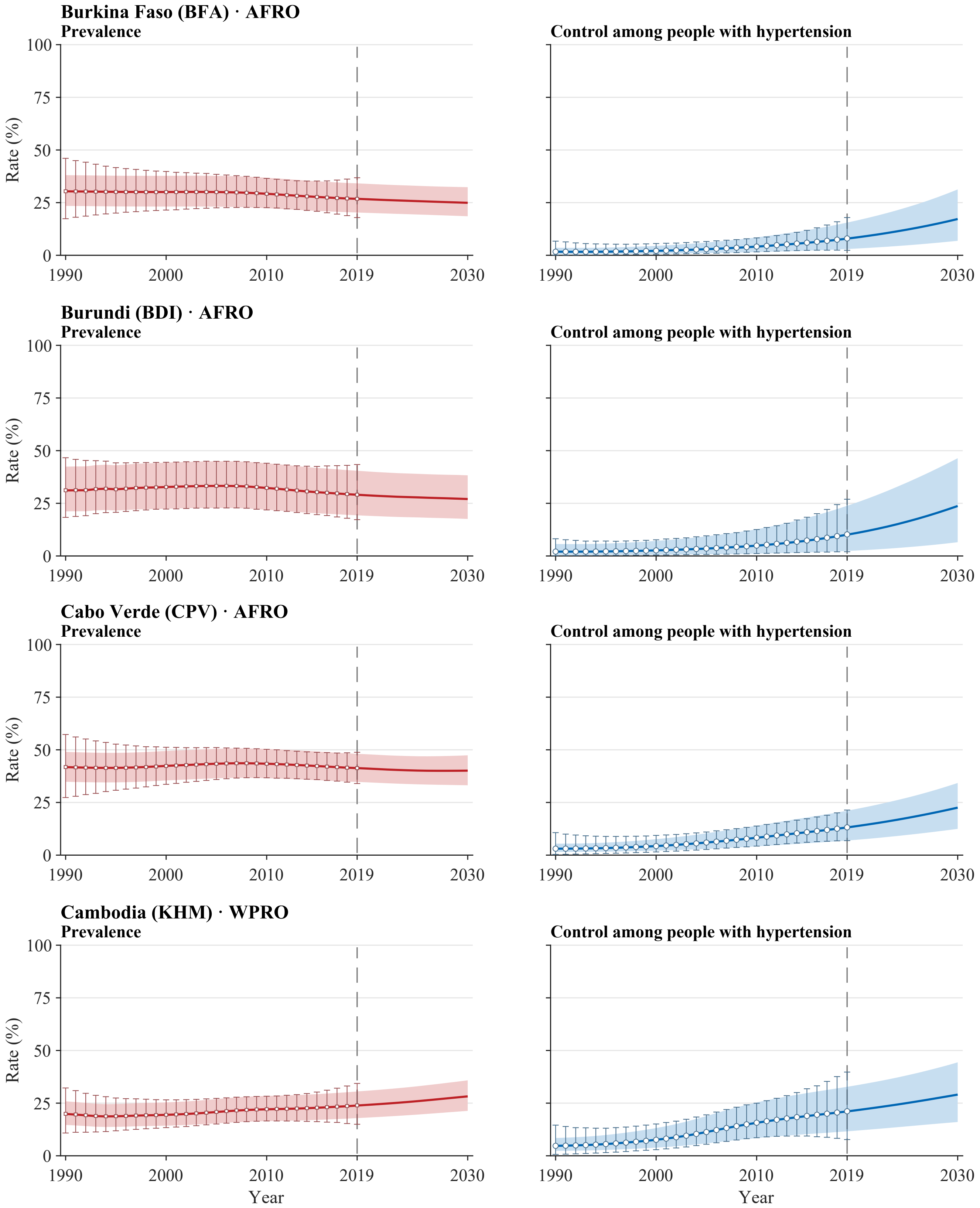


**Legend.** Open squares and error bars in prevalence panels and open circles and error bars in control panels show annual NCD-RisC central estimates with propagated 95% uncertainty intervals for 1990–2019. Solid red and blue lines and shaded bands show model medians and 95% uncertainty intervals for 1990–2030. The vertical dashed line marks 2019, the final year with reported estimates; values for 2020–2030 are forecast. Latent stratum-specific values in 2019 are anchored to the published central estimates. Propagated intervals for the published estimates are wider than the model bands during the historical period by construction of the external uncertainty layer. Model uncertainty represents a persistent level offset. All panels use a common 0–100% scale. Prevalence is defined among all adults aged 30–79 years, whereas control is defined among adults with hypertension. Estimates use CORE-P25 and the prespecified main dependence assumptions.

**Supplementary Figure S3. Country-level trajectories of aggregate hypertension prevalence and control, 1990–2030**
Atlas panel page 9 of 50; countries 33–36 of 200.

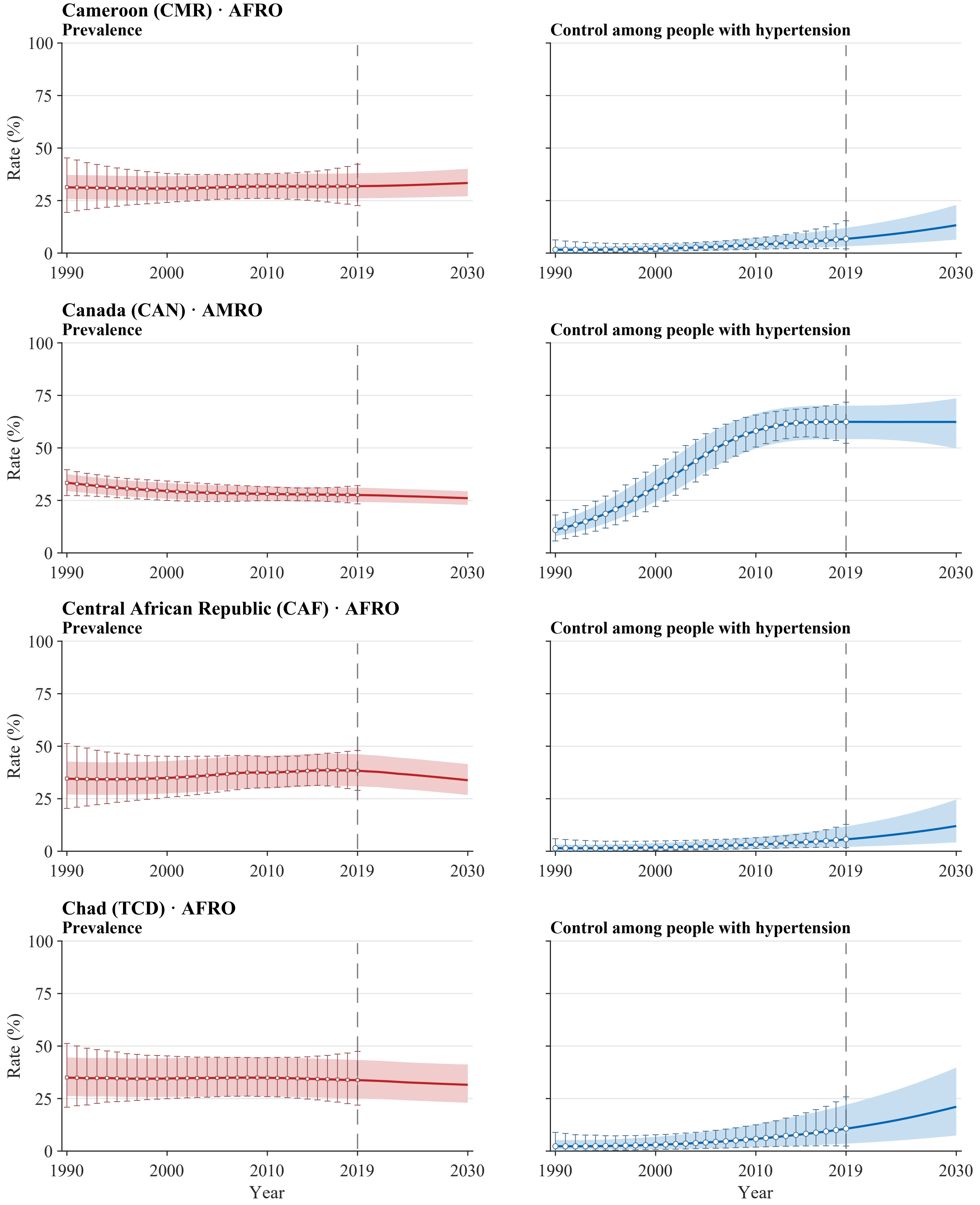


**Legend.** Open squares and error bars in prevalence panels and open circles and error bars in control panels show annual NCD-RisC central estimates with propagated 95% uncertainty intervals for 1990–2019. Solid red and blue lines and shaded bands show model medians and 95% uncertainty intervals for 1990–2030. The vertical dashed line marks 2019, the final year with reported estimates; values for 2020–2030 are forecast. Latent stratum-specific values in 2019 are anchored to the published central estimates. Propagated intervals for the published estimates are wider than the model bands during the historical period by construction of the external uncertainty layer. Model uncertainty represents a persistent level offset. All panels use a common 0–100% scale. Prevalence is defined among all adults aged 30–79 years, whereas control is defined among adults with hypertension. Estimates use CORE-P25 and the prespecified main dependence assumptions.

**Supplementary Figure S3. Country-level trajectories of aggregate hypertension prevalence and control, 1990–2030**
Atlas panel page 10 of 50; countries 37–40 of 200.

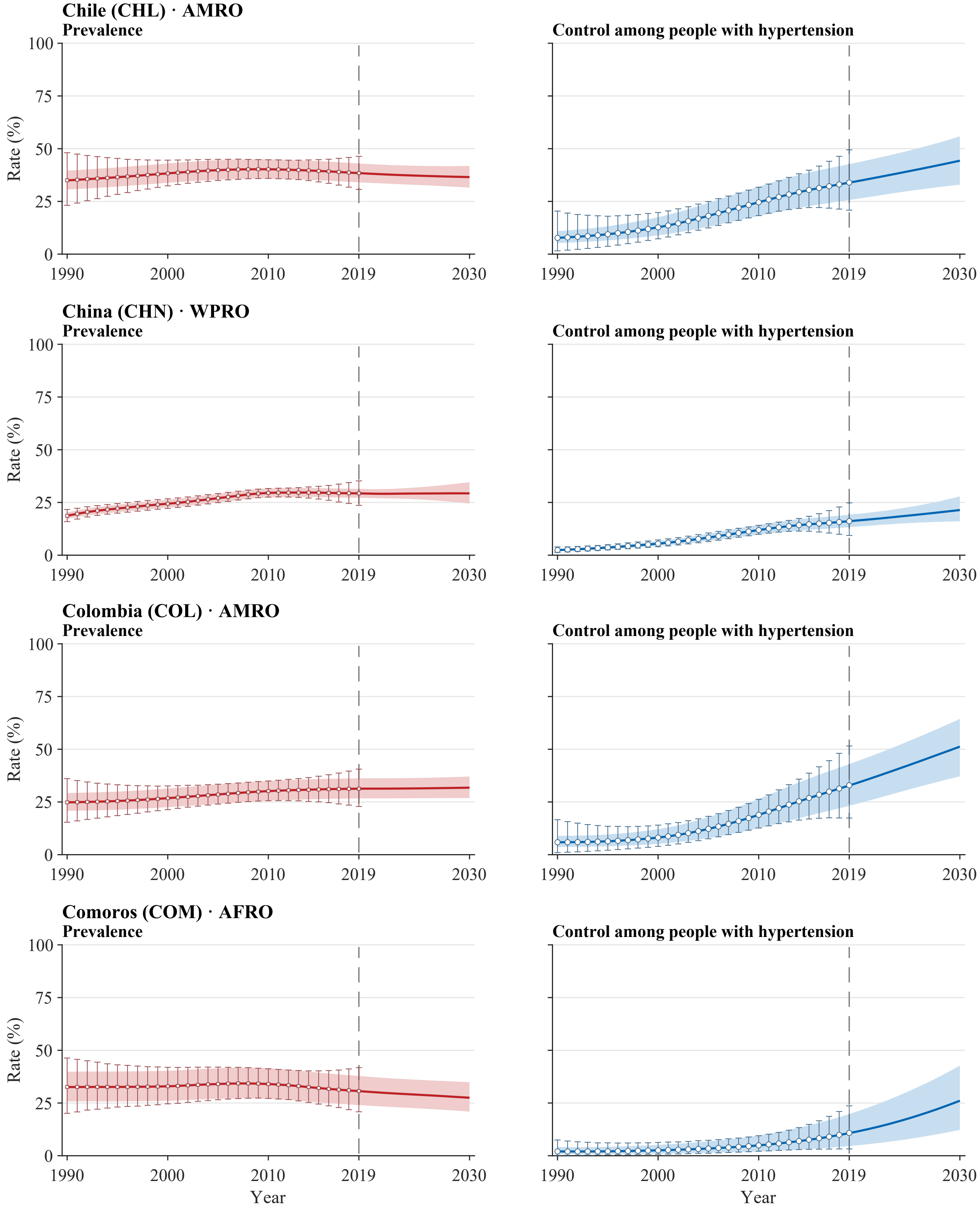


**Legend.** Open squares and error bars in prevalence panels and open circles and error bars in control panels show annual NCD-RisC central estimates with propagated 95% uncertainty intervals for 1990–2019. Solid red and blue lines and shaded bands show model medians and 95% uncertainty intervals for 1990–2030. The vertical dashed line marks 2019, the final year with reported estimates; values for 2020–2030 are forecast. Latent stratum-specific values in 2019 are anchored to the published central estimates. Propagated intervals for the published estimates are wider than the model bands during the historical period by construction of the external uncertainty layer. Model uncertainty represents a persistent level offset. All panels use a common 0–100% scale. Prevalence is defined among all adults aged 30–79 years, whereas control is defined among adults with hypertension. Estimates use CORE-P25 and the prespecified main dependence assumptions.

**Supplementary Figure S3. Country-level trajectories of aggregate hypertension prevalence and control, 1990–2030**
Atlas panel page 11 of 50; countries 41–44 of 200.

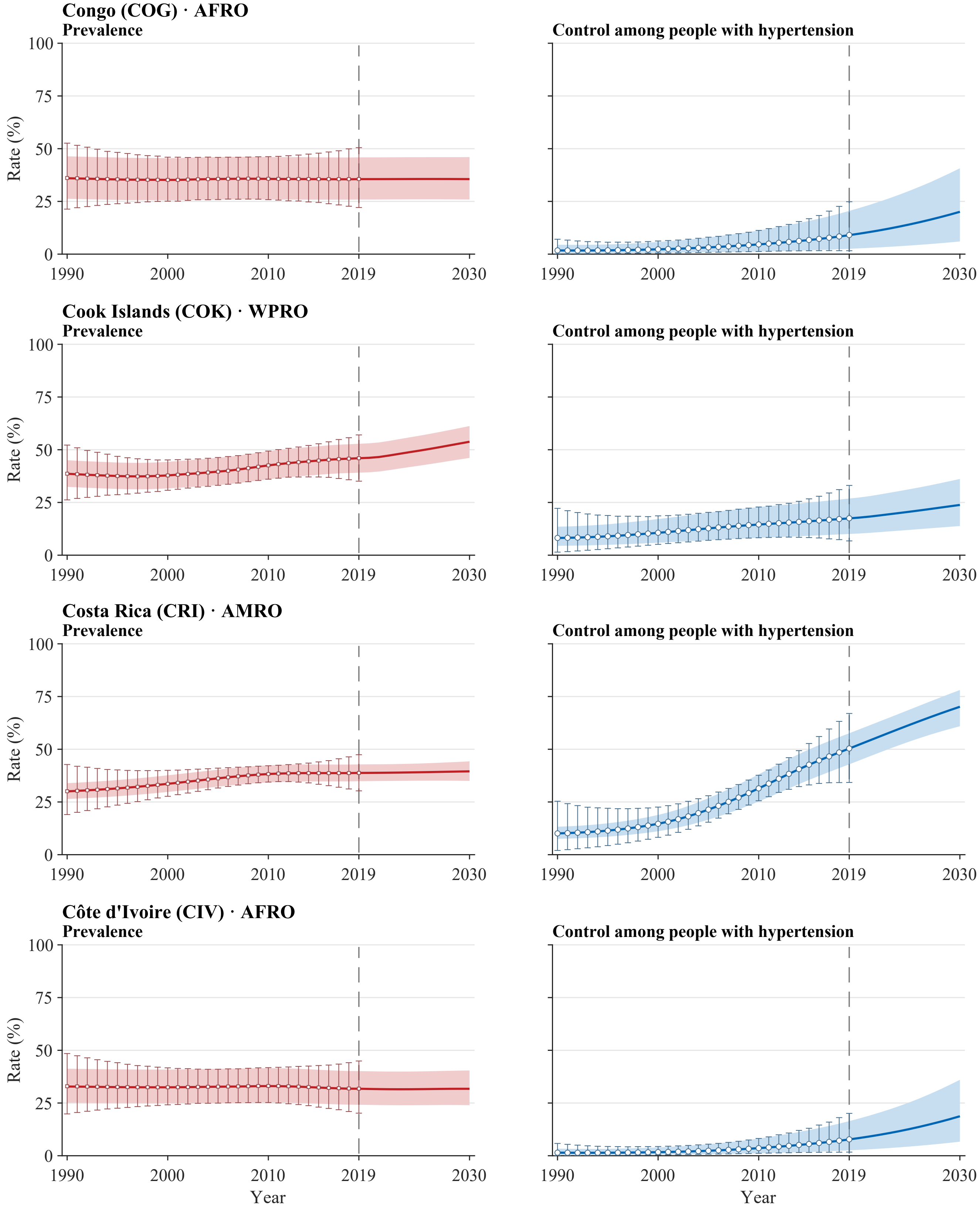


**Legend.** Open squares and error bars in prevalence panels and open circles and error bars in control panels show annual NCD-RisC central estimates with propagated 95% uncertainty intervals for 1990–2019. Solid red and blue lines and shaded bands show model medians and 95% uncertainty intervals for 1990–2030. The vertical dashed line marks 2019, the final year with reported estimates; values for 2020–2030 are forecast. Latent stratum-specific values in 2019 are anchored to the published central estimates. Propagated intervals for the published estimates are wider than the model bands during the historical period by construction of the external uncertainty layer. Model uncertainty represents a persistent level offset. All panels use a common 0–100% scale. Prevalence is defined among all adults aged 30–79 years, whereas control is defined among adults with hypertension. Estimates use CORE-P25 and the prespecified main dependence assumptions.

**Supplementary Figure S3. Country-level trajectories of aggregate hypertension prevalence and control, 1990–2030**

Atlas panel page 12 of 50; countries 45–48 of 200.

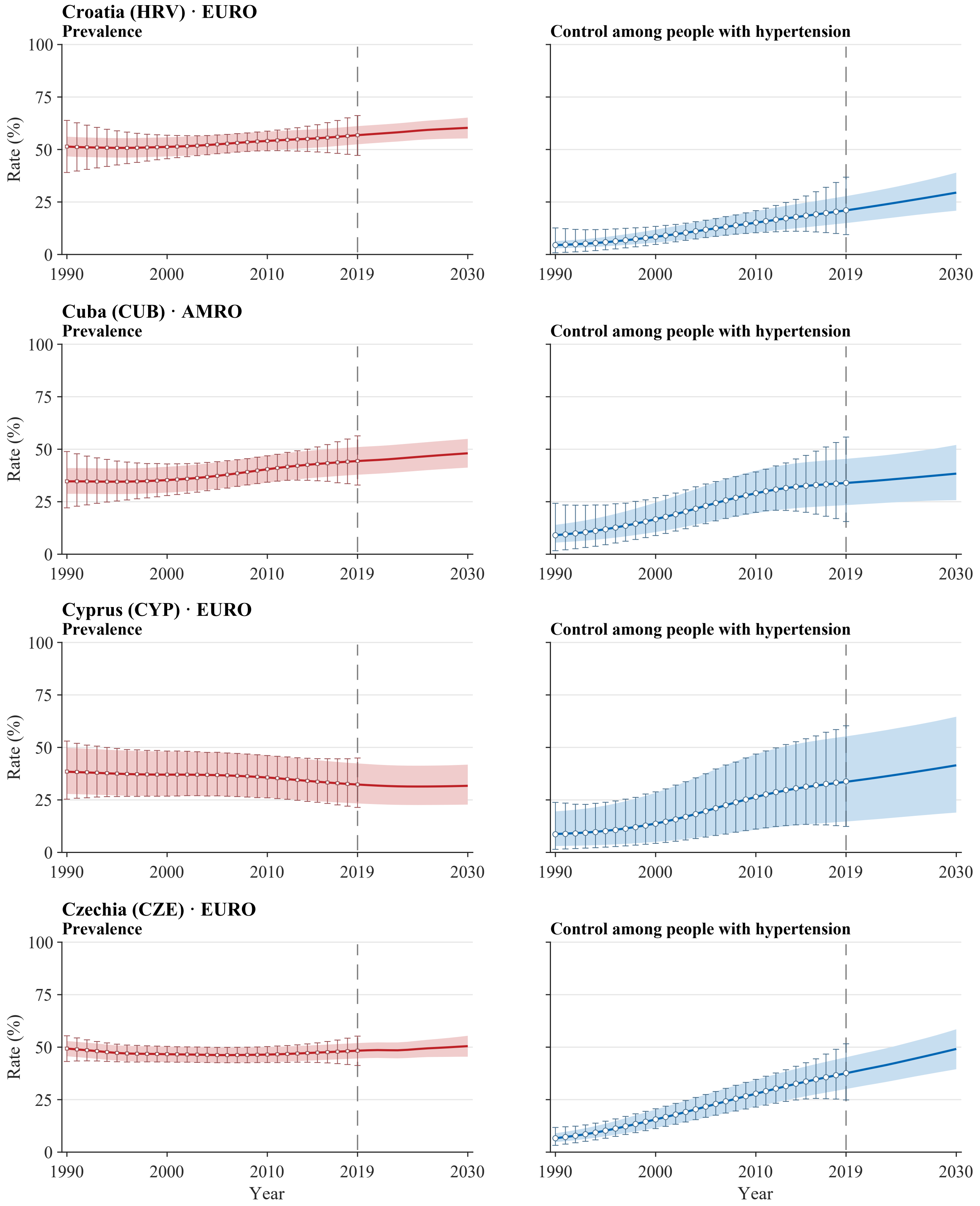


**Legend.** Open squares and error bars in prevalence panels and open circles and error bars in control panels show annual NCD-RisC central estimates with propagated 95% uncertainty intervals for 1990–2019. Solid red and blue lines and shaded bands show model medians and 95% uncertainty intervals for 1990–2030. The vertical dashed line marks 2019, the final year with reported estimates; values for 2020–2030 are forecast. Latent stratum-specific values in 2019 are anchored to the published central estimates. Propagated intervals for the published estimates are wider than the model bands during the historical period by construction of the external uncertainty layer. Model uncertainty represents a persistent level offset. All panels use a common 0–100% scale. Prevalence is defined among all adults aged 30–79 years, whereas control is defined among adults with hypertension. Estimates use CORE-P25 and the prespecified main dependence assumptions.

**Supplementary Figure S3. Country-level trajectories of aggregate hypertension prevalence and control, 1990–2030**

Atlas panel page 13 of 50; countries 49–52 of 200.

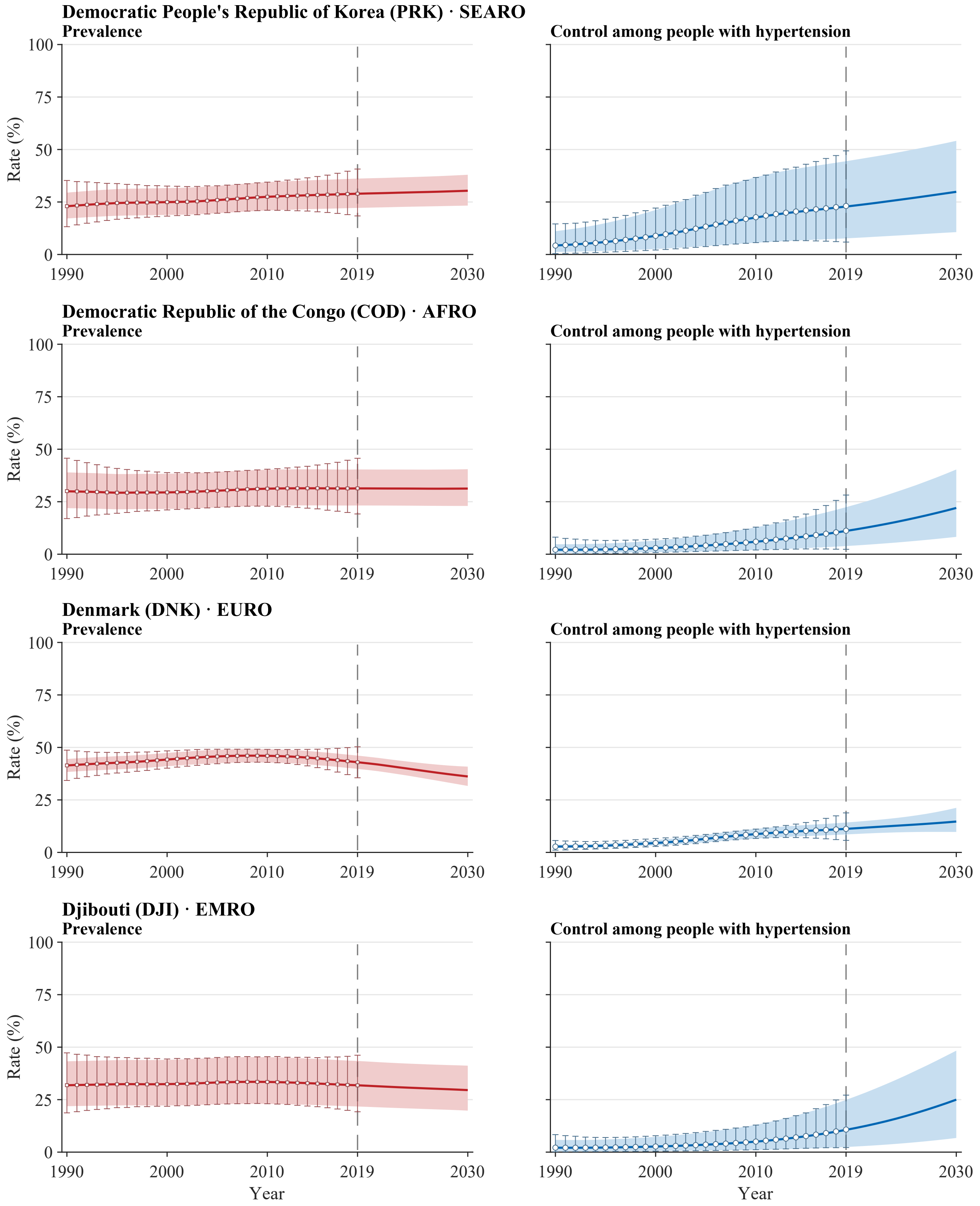


**Legend.** Open squares and error bars in prevalence panels and open circles and error bars in control panels show annual NCD-RisC central estimates with propagated 95% uncertainty intervals for 1990–2019. Solid red and blue lines and shaded bands show model medians and 95% uncertainty intervals for 1990–2030. The vertical dashed line marks 2019, the final year with reported estimates; values for 2020–2030 are forecast. Latent stratum-specific values in 2019 are anchored to the published central estimates. Propagated intervals for the published estimates are wider than the model bands during the historical period by construction of the external uncertainty layer. Model uncertainty represents a persistent level offset. All panels use a common 0–100% scale. Prevalence is defined among all adults aged 30–79 years, whereas control is defined among adults with hypertension. Estimates use CORE-P25 and the prespecified main dependence assumptions.

**Supplementary Figure S3. Country-level trajectories of aggregate hypertension prevalence and control, 1990–2030**

Atlas panel page 14 of 50; countries 53–56 of 200.

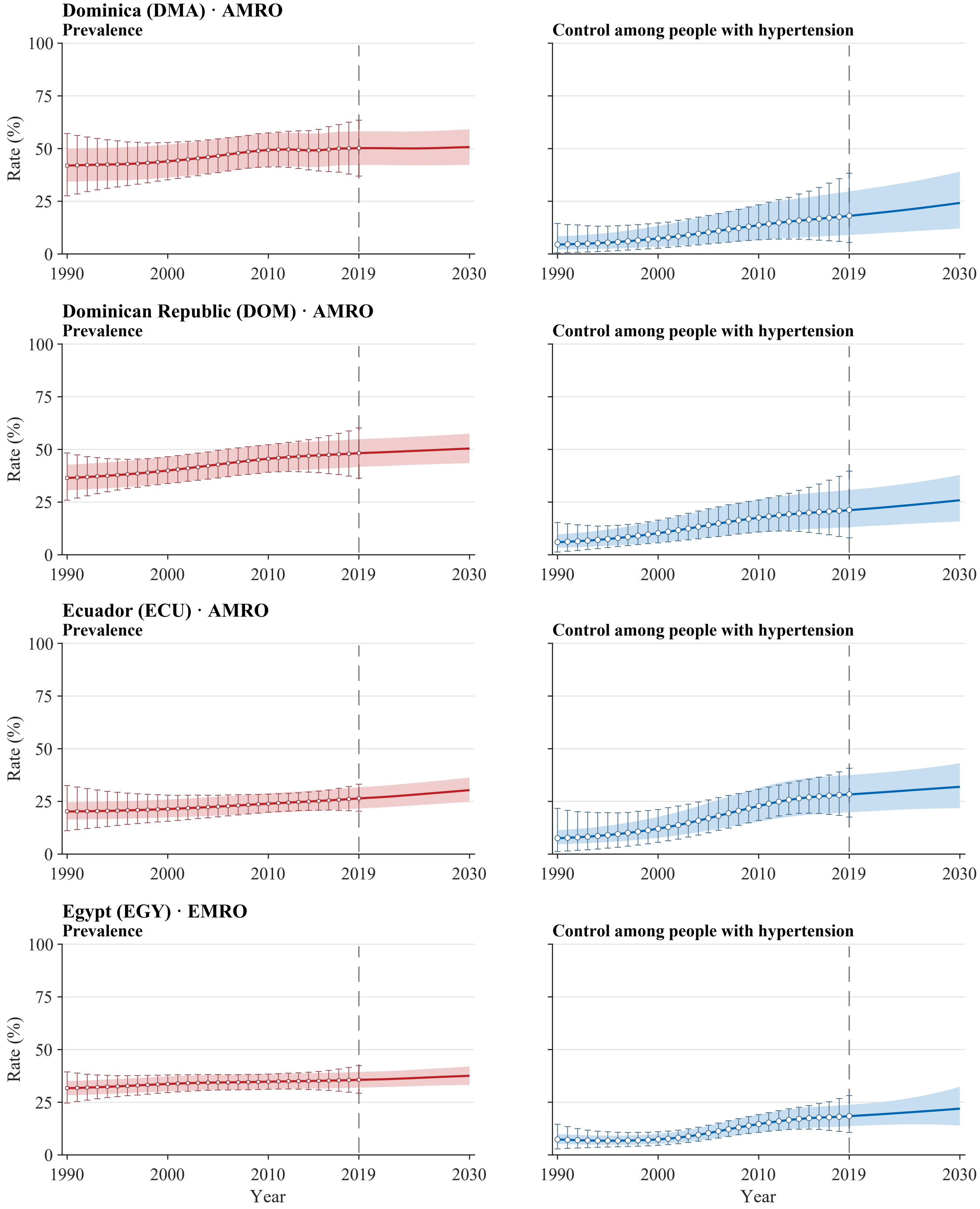


**Legend.** Open squares and error bars in prevalence panels and open circles and error bars in control panels show annual NCD-RisC central estimates with propagated 95% uncertainty intervals for 1990–2019. Solid red and blue lines and shaded bands show model medians and 95% uncertainty intervals for 1990–2030. The vertical dashed line marks 2019, the final year with reported estimates; values for 2020–2030 are forecast. Latent stratum-specific values in 2019 are anchored to the published central estimates. Propagated intervals for the published estimates are wider than the model bands during the historical period by construction of the external uncertainty layer. Model uncertainty represents a persistent level offset. All panels use a common 0–100% scale. Prevalence is defined among all adults aged 30–79 years, whereas control is defined among adults with hypertension. Estimates use CORE-P25 and the prespecified main dependence assumptions.

**Supplementary Figure S3. Country-level trajectories of aggregate hypertension prevalence and control, 1990–2030**

Atlas panel page 15 of 50; countries 57–60 of 200.

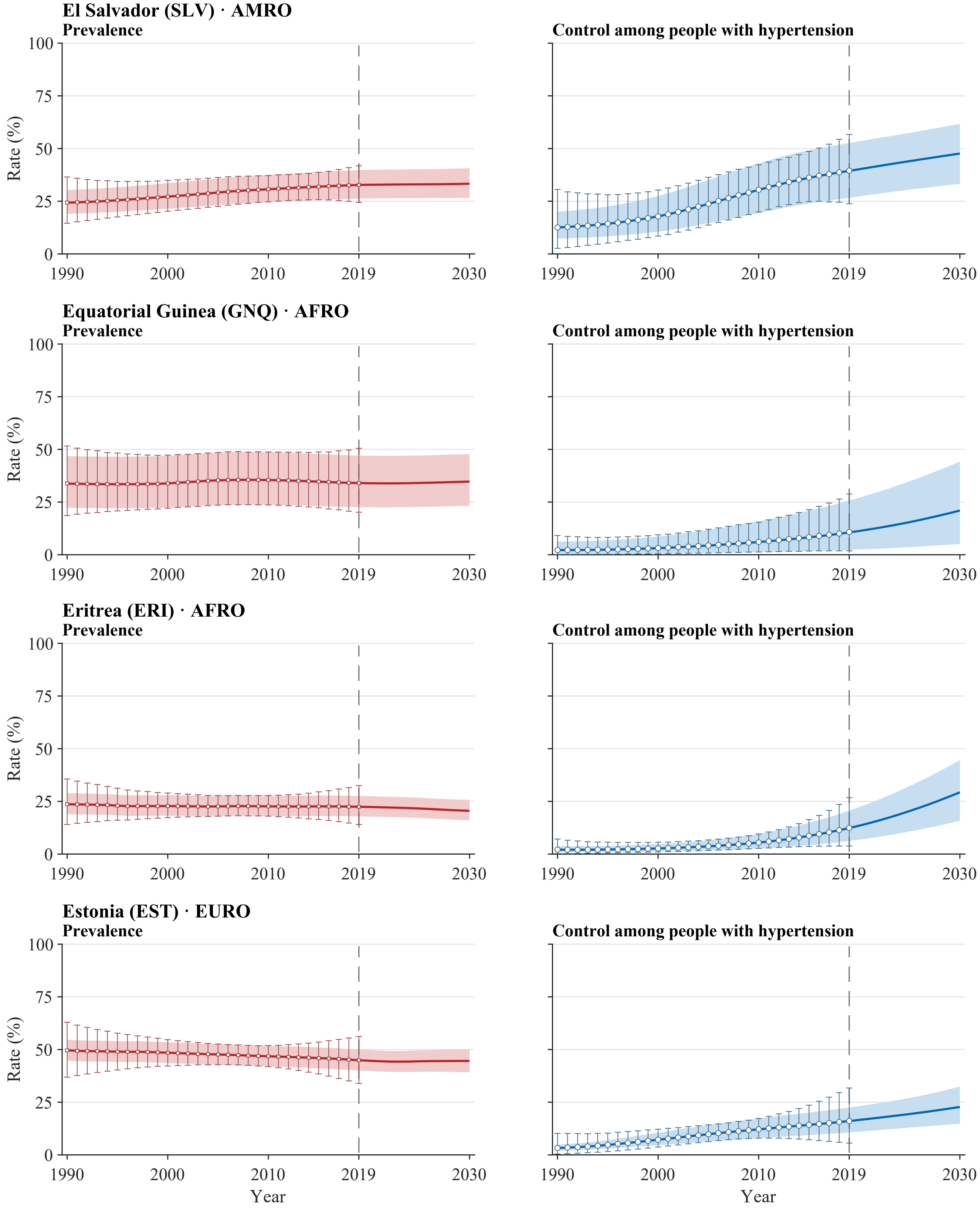


**Legend.** Open squares and error bars in prevalence panels and open circles and error bars in control panels show annual NCD-RisC central estimates with propagated 95% uncertainty intervals for 1990–2019. Solid red and blue lines and shaded bands show model medians and 95% uncertainty intervals for 1990–2030. The vertical dashed line marks 2019, the final year with reported estimates; values for 2020–2030 are forecast. Latent stratum-specific values in 2019 are anchored to the published central estimates. Propagated intervals for the published estimates are wider than the model bands during the historical period by construction of the external uncertainty layer. Model uncertainty represents a persistent level offset. All panels use a common 0–100% scale. Prevalence is defined among all adults aged 30–79 years, whereas control is defined among adults with hypertension. Estimates use CORE-P25 and the prespecified main dependence assumptions.

**Supplementary Figure S3. Country-level trajectories of aggregate hypertension prevalence and control, 1990–2030**

Atlas panel page 16 of 50; countries 61–64 of 200.

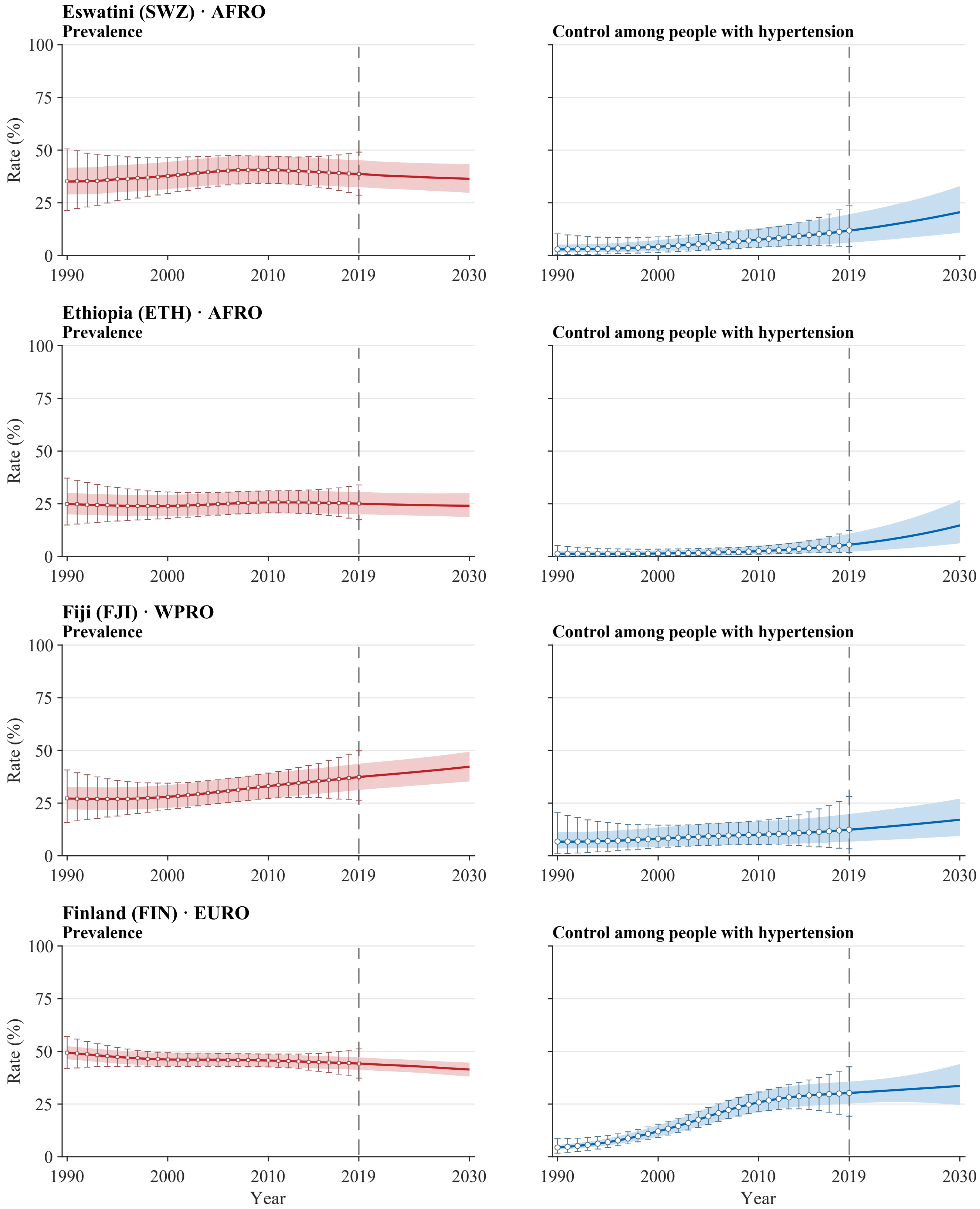


**Legend.** Open squares and error bars in prevalence panels and open circles and error bars in control panels show annual NCD-RisC central estimates with propagated 95% uncertainty intervals for 1990–2019. Solid red and blue lines and shaded bands show model medians and 95% uncertainty intervals for 1990–2030. The vertical dashed line marks 2019, the final year with reported estimates; values for 2020–2030 are forecast. Latent stratum-specific values in 2019 are anchored to the published central estimates. Propagated intervals for the published estimates are wider than the model bands during the historical period by construction of the external uncertainty layer. Model uncertainty represents a persistent level offset. All panels use a common 0–100% scale. Prevalence is defined among all adults aged 30–79 years, whereas control is defined among adults with hypertension. Estimates use CORE-P25 and the prespecified main dependence assumptions.

**Supplementary Figure S3. Country-level trajectories of aggregate hypertension prevalence and control, 1990–2030**
Atlas panel page 17 of 50; countries 65–68 of 200.

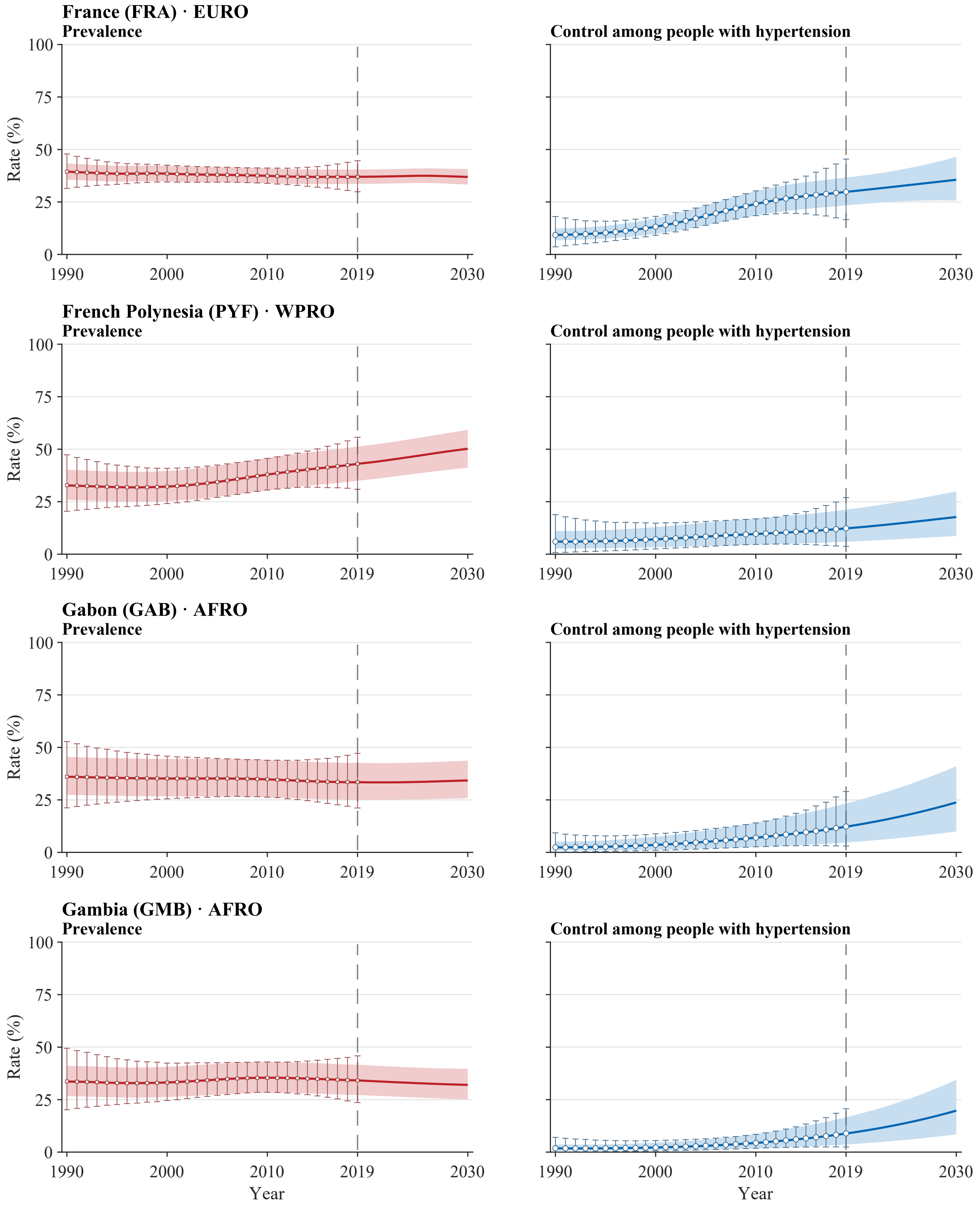


**Legend.** Open squares and error bars in prevalence panels and open circles and error bars in control panels show annual NCD-RisC central estimates with propagated 95% uncertainty intervals for 1990–2019. Solid red and blue lines and shaded bands show model medians and 95% uncertainty intervals for 1990–2030. The vertical dashed line marks 2019, the final year with reported estimates; values for 2020–2030 are forecast. Latent stratum-specific values in 2019 are anchored to the published central estimates. Propagated intervals for the published estimates are wider than the model bands during the historical period by construction of the external uncertainty layer. Model uncertainty represents a persistent level offset. All panels use a common 0–100% scale. Prevalence is defined among all adults aged 30–79 years, whereas control is defined among adults with hypertension. Estimates use CORE-P25 and the prespecified main dependence assumptions.

**Supplementary Figure S3. Country-level trajectories of aggregate hypertension prevalence and control, 1990–2030**

Atlas panel page 18 of 50; countries 69–72 of 200.

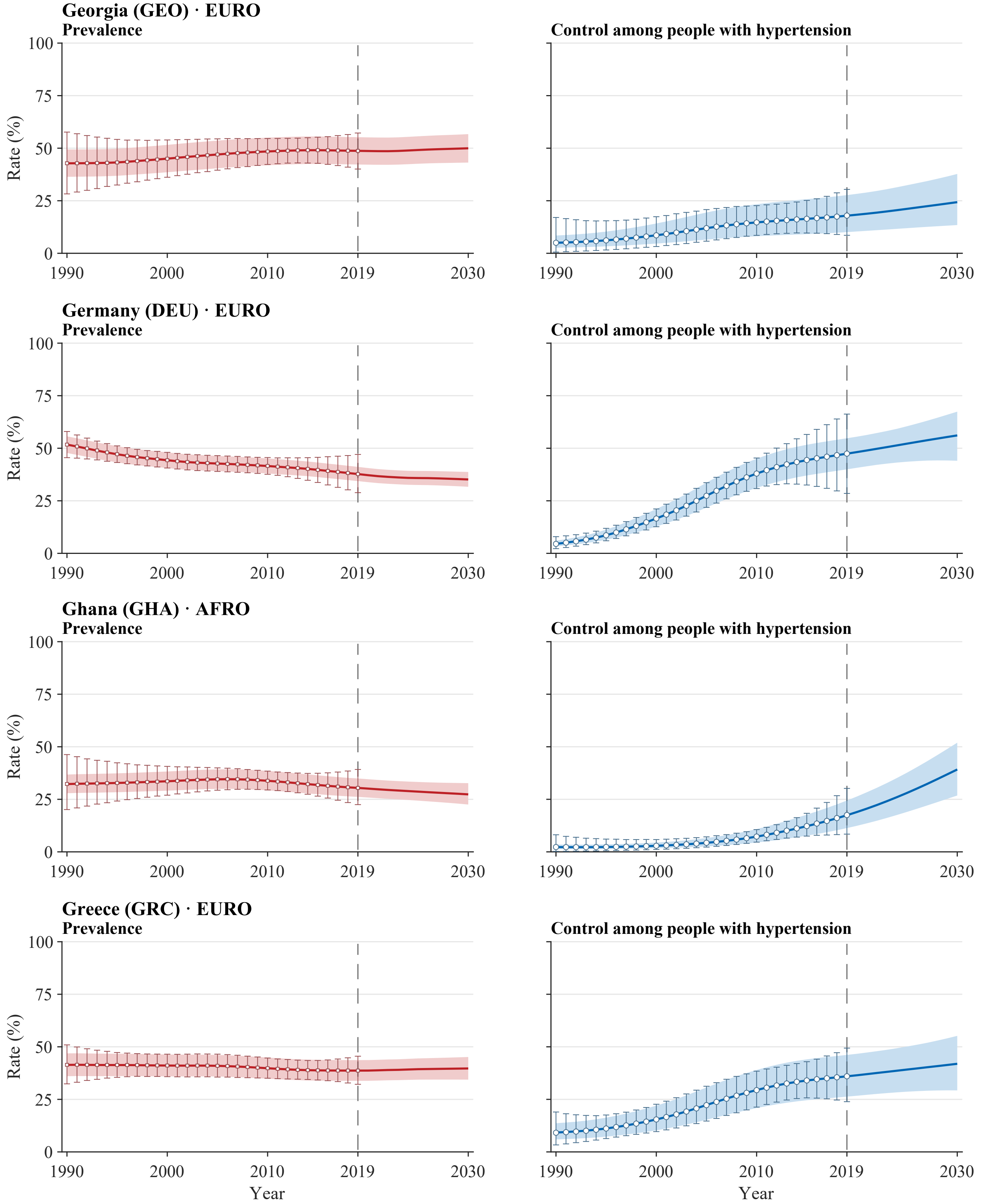


**Legend.** Open squares and error bars in prevalence panels and open circles and error bars in control panels show annual NCD-RisC central estimates with propagated 95% uncertainty intervals for 1990–2019. Solid red and blue lines and shaded bands show model medians and 95% uncertainty intervals for 1990–2030. The vertical dashed line marks 2019, the final year with reported estimates; values for 2020–2030 are forecast. Latent stratum-specific values in 2019 are anchored to the published central estimates. Propagated intervals for the published estimates are wider than the model bands during the historical period by construction of the external uncertainty layer. Model uncertainty represents a persistent level offset. All panels use a common 0–100% scale. Prevalence is defined among all adults aged 30–79 years, whereas control is defined among adults with hypertension. Estimates use CORE-P25 and the prespecified main dependence assumptions.

**Supplementary Figure S3. Country-level trajectories of aggregate hypertension prevalence and control, 1990–2030**

Atlas panel page 19 of 50; countries 73–76 of 200.

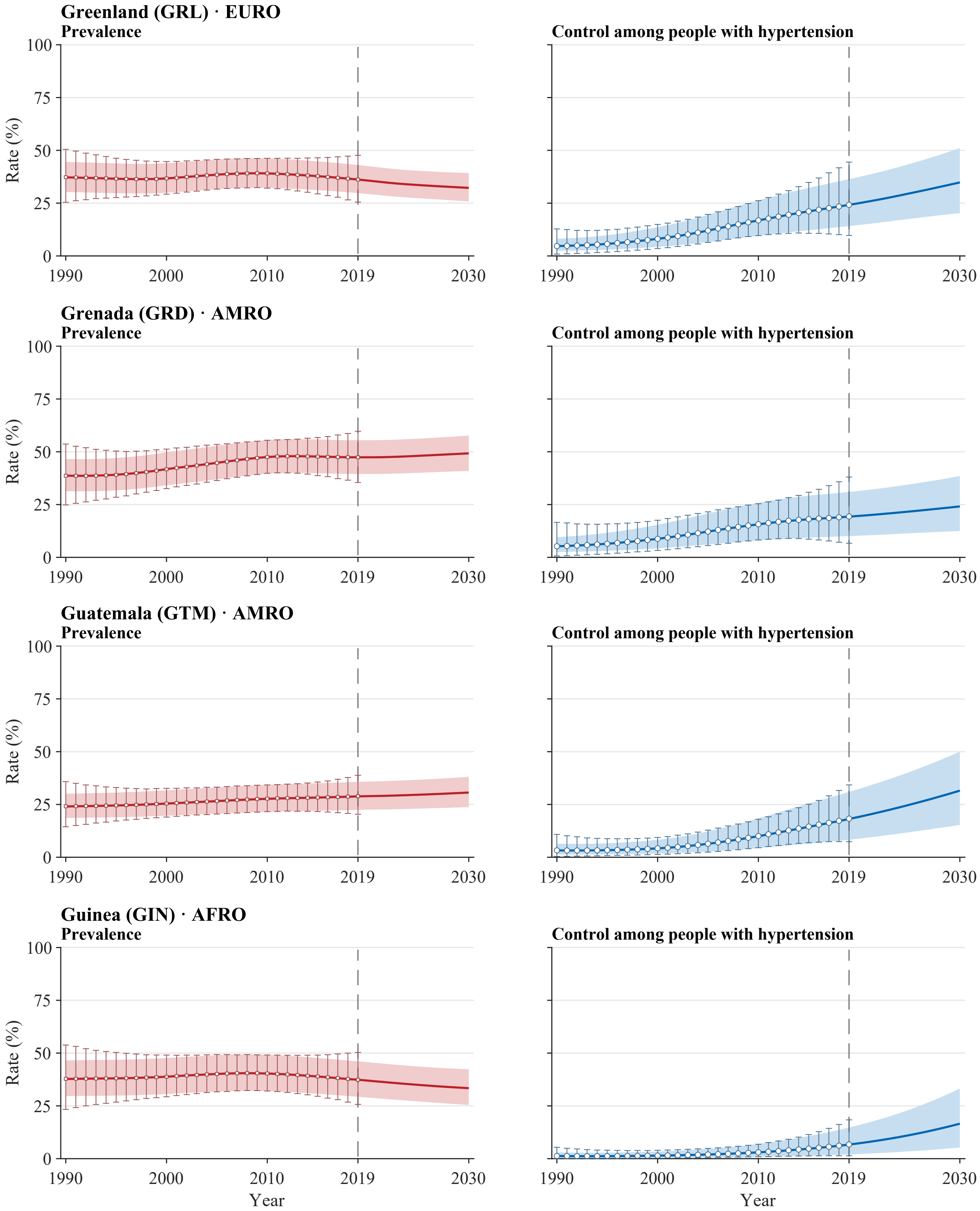


**Legend.** Open squares and error bars in prevalence panels and open circles and error bars in control panels show annual NCD-RisC central estimates with propagated 95% uncertainty intervals for 1990–2019. Solid red and blue lines and shaded bands show model medians and 95% uncertainty intervals for 1990–2030. The vertical dashed line marks 2019, the final year with reported estimates; values for 2020–2030 are forecast. Latent stratum-specific values in 2019 are anchored to the published central estimates. Propagated intervals for the published estimates are wider than the model bands during the historical period by construction of the external uncertainty layer. Model uncertainty represents a persistent level offset. All panels use a common 0–100% scale. Prevalence is defined among all adults aged 30–79 years, whereas control is defined among adults with hypertension. Estimates use CORE-P25 and the prespecified main dependence assumptions.

**Supplementary Figure S3. Country-level trajectories of aggregate hypertension prevalence and control, 1990–2030**
Atlas panel page 20 of 50; countries 77–80 of 200.

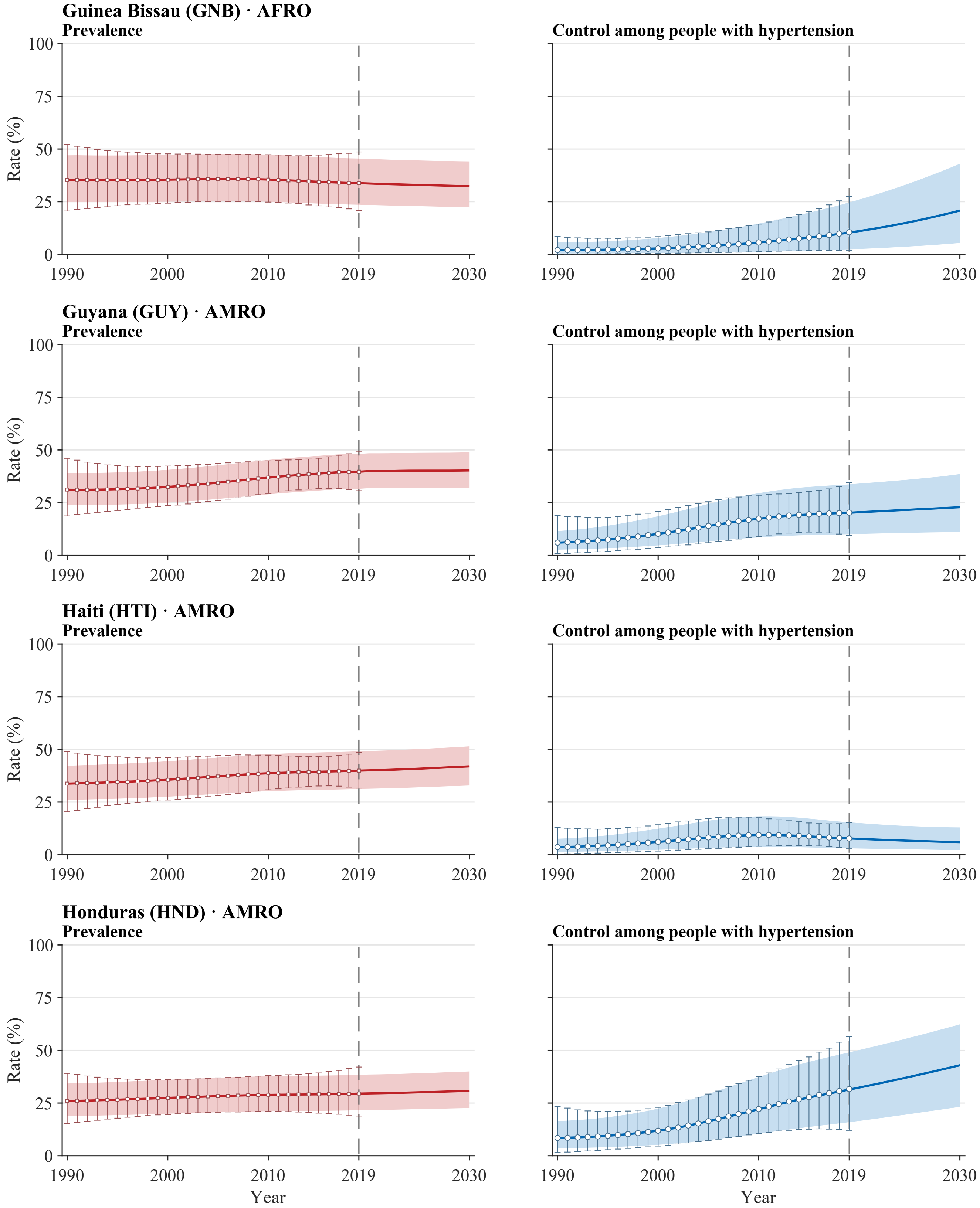


**Legend.** Open squares and error bars in prevalence panels and open circles and error bars in control panels show annual NCD-RisC central estimates with propagated 95% uncertainty intervals for 1990–2019. Solid red and blue lines and shaded bands show model medians and 95% uncertainty intervals for 1990–2030. The vertical dashed line marks 2019, the final year with reported estimates; values for 2020–2030 are forecast. Latent stratum-specific values in 2019 are anchored to the published central estimates. Propagated intervals for the published estimates are wider than the model bands during the historical period by construction of the external uncertainty layer. Model uncertainty represents a persistent level offset. All panels use a common 0–100% scale. Prevalence is defined among all adults aged 30–79 years, whereas control is defined among adults with hypertension. Estimates use CORE-P25 and the prespecified main dependence assumptions.

**Supplementary Figure S3. Country-level trajectories of aggregate hypertension prevalence and control, 1990–2030**

Atlas panel page 21 of 50; countries 81–84 of 200.

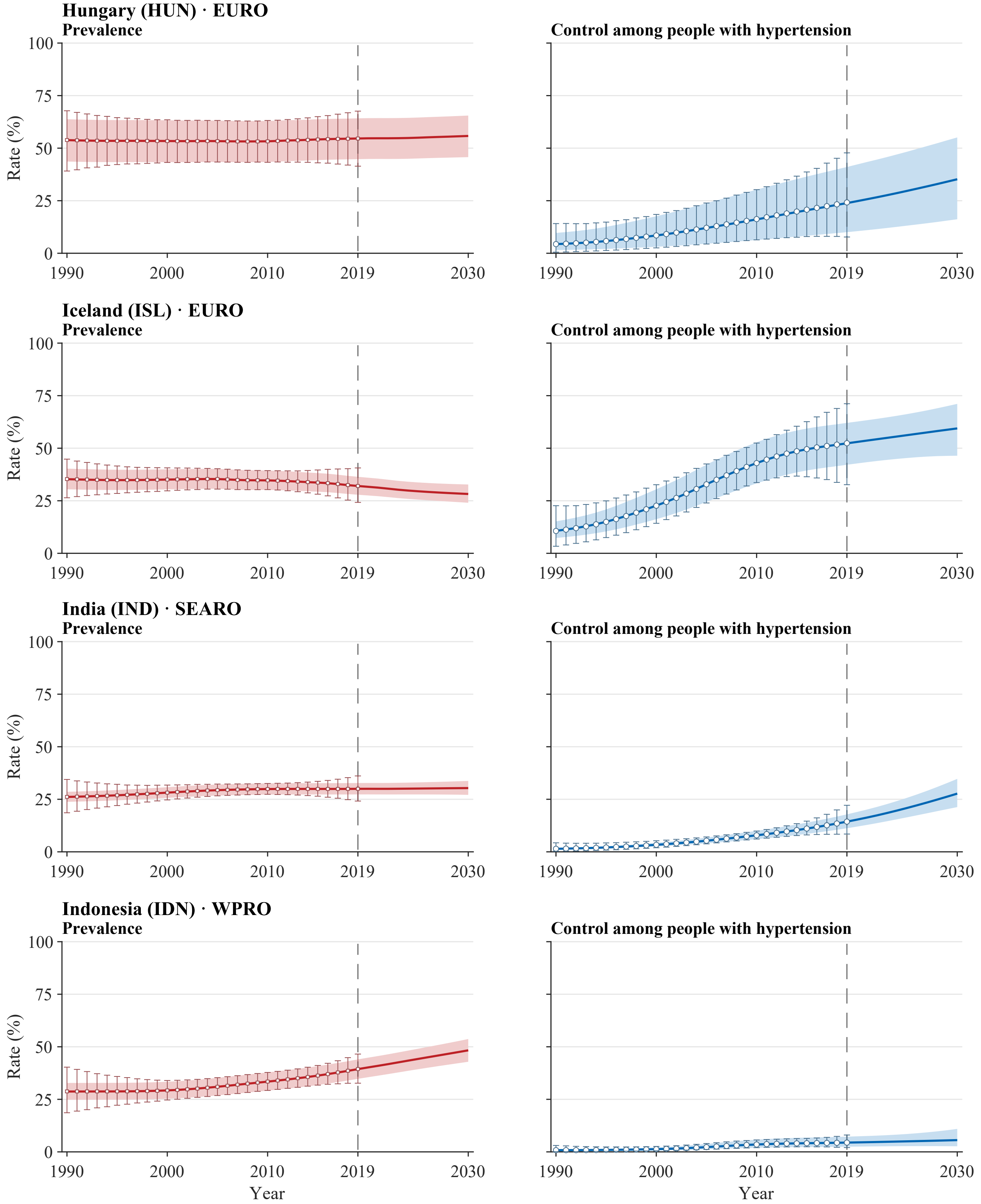


**Legend.** Open squares and error bars in prevalence panels and open circles and error bars in control panels show annual NCD-RisC central estimates with propagated 95% uncertainty intervals for 1990–2019. Solid red and blue lines and shaded bands show model medians and 95% uncertainty intervals for 1990–2030. The vertical dashed line marks 2019, the final year with reported estimates; values for 2020–2030 are forecast. Latent stratum-specific values in 2019 are anchored to the published central estimates. Propagated intervals for the published estimates are wider than the model bands during the historical period by construction of the external uncertainty layer. Model uncertainty represents a persistent level offset. All panels use a common 0–100% scale. Prevalence is defined among all adults aged 30–79 years, whereas control is defined among adults with hypertension. Estimates use CORE-P25 and the prespecified main dependence assumptions.

**Supplementary Figure S3. Country-level trajectories of aggregate hypertension prevalence and control, 1990–2030**

Atlas panel page 22 of 50; countries 85–88 of 200.

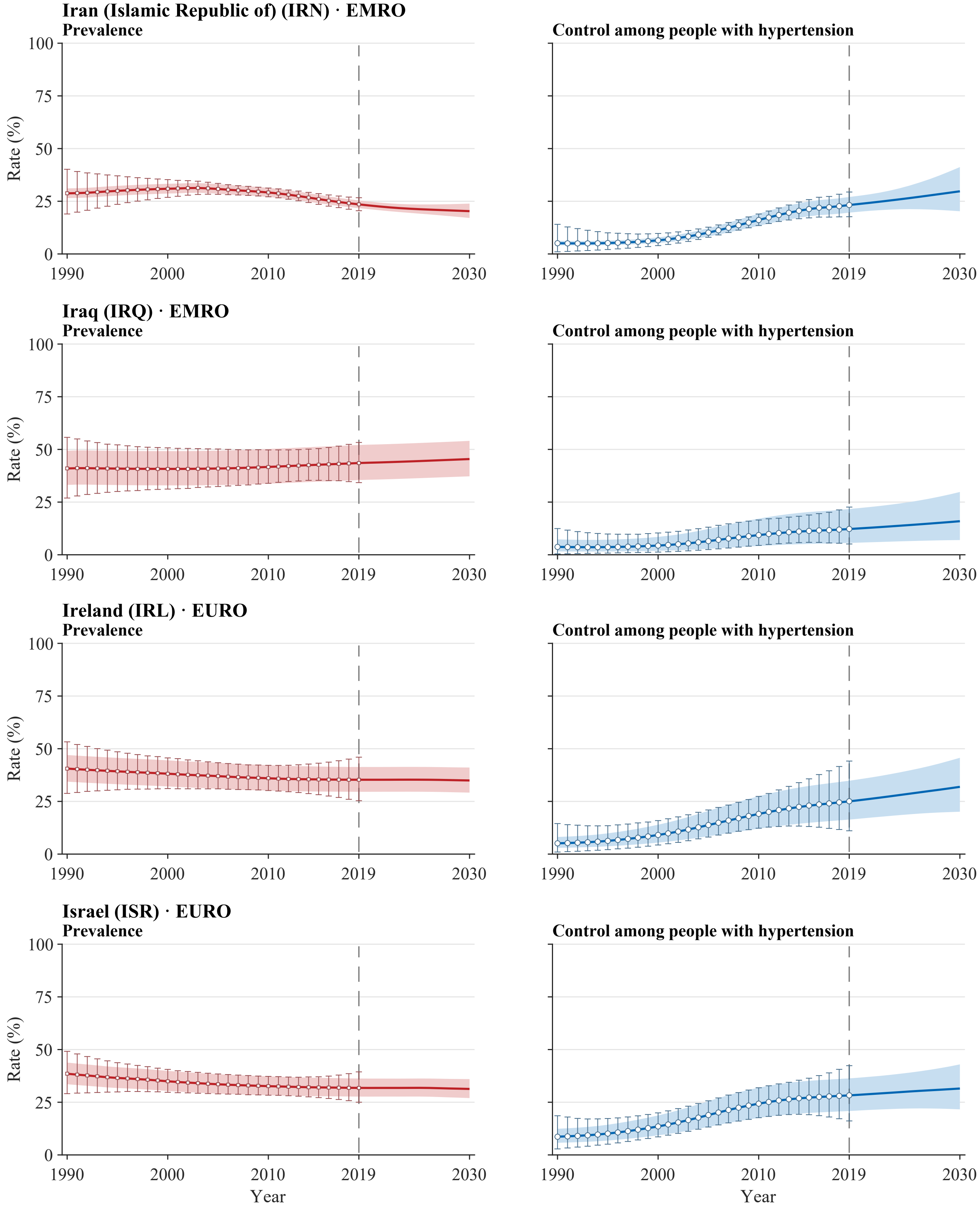


**Legend.** Open squares and error bars in prevalence panels and open circles and error bars in control panels show annual NCD-RisC central estimates with propagated 95% uncertainty intervals for 1990–2019. Solid red and blue lines and shaded bands show model medians and 95% uncertainty intervals for 1990–2030. The vertical dashed line marks 2019, the final year with reported estimates; values for 2020–2030 are forecast. Latent stratum-specific values in 2019 are anchored to the published central estimates. Propagated intervals for the published estimates are wider than the model bands during the historical period by construction of the external uncertainty layer. Model uncertainty represents a persistent level offset. All panels use a common 0–100% scale. Prevalence is defined among all adults aged 30–79 years, whereas control is defined among adults with hypertension. Estimates use CORE-P25 and the prespecified main dependence assumptions.

**Supplementary Figure S3. Country-level trajectories of aggregate hypertension prevalence and control, 1990–2030**
Atlas panel page 23 of 50; countries 89–92 of 200.

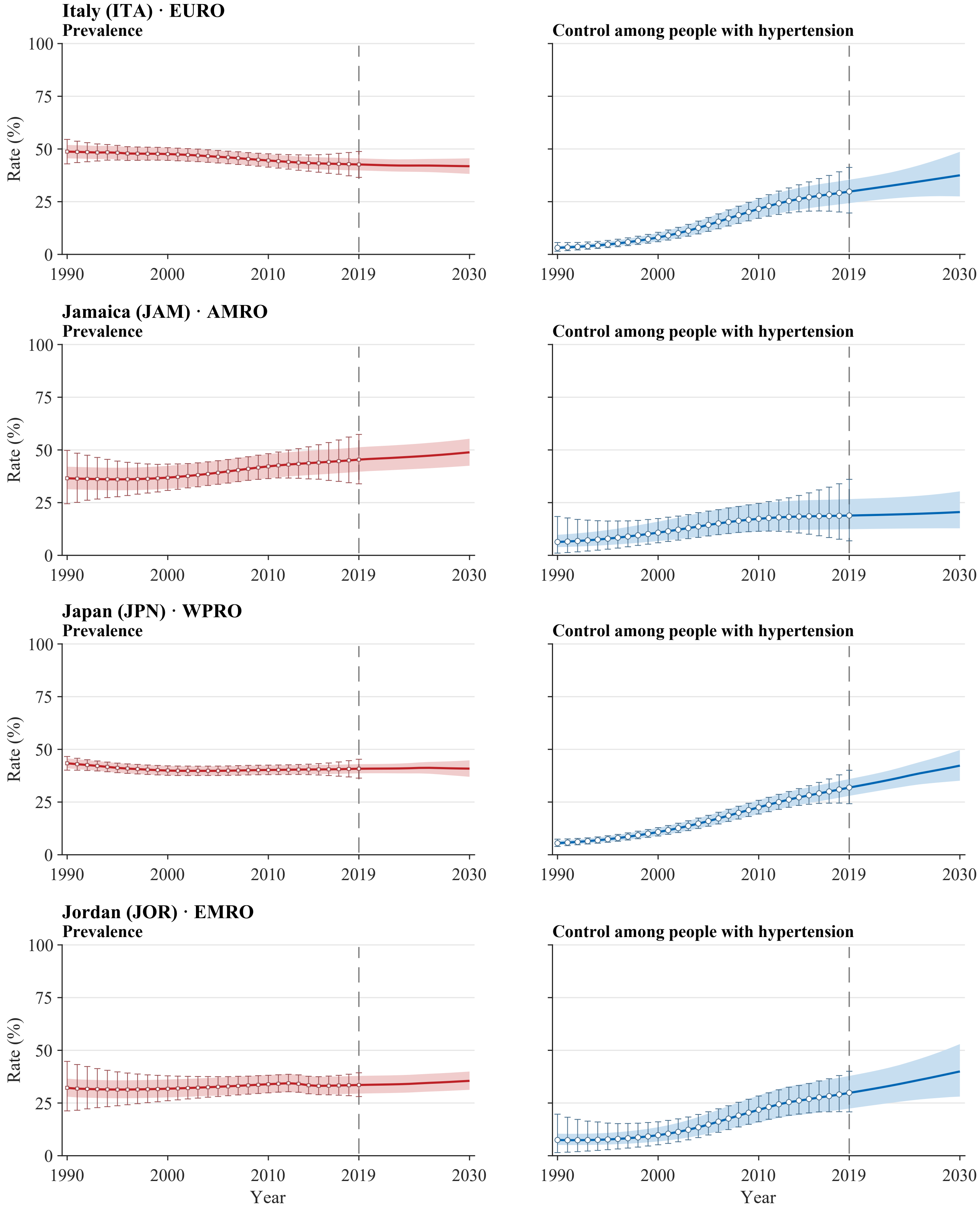


**Legend.** Open squares and error bars in prevalence panels and open circles and error bars in control panels show annual NCD-RisC central estimates with propagated 95% uncertainty intervals for 1990–2019. Solid red and blue lines and shaded bands show model medians and 95% uncertainty intervals for 1990–2030. The vertical dashed line marks 2019, the final year with reported estimates; values for 2020–2030 are forecast. Latent stratum-specific values in 2019 are anchored to the published central estimates. Propagated intervals for the published estimates are wider than the model bands during the historical period by construction of the external uncertainty layer. Model uncertainty represents a persistent level offset. All panels use a common 0–100% scale. Prevalence is defined among all adults aged 30–79 years, whereas control is defined among adults with hypertension. Estimates use CORE-P25 and the prespecified main dependence assumptions.

**Supplementary Figure S3. Country-level trajectories of aggregate hypertension prevalence and control, 1990–2030**
Atlas panel page 24 of 50; countries 93–96 of 200.

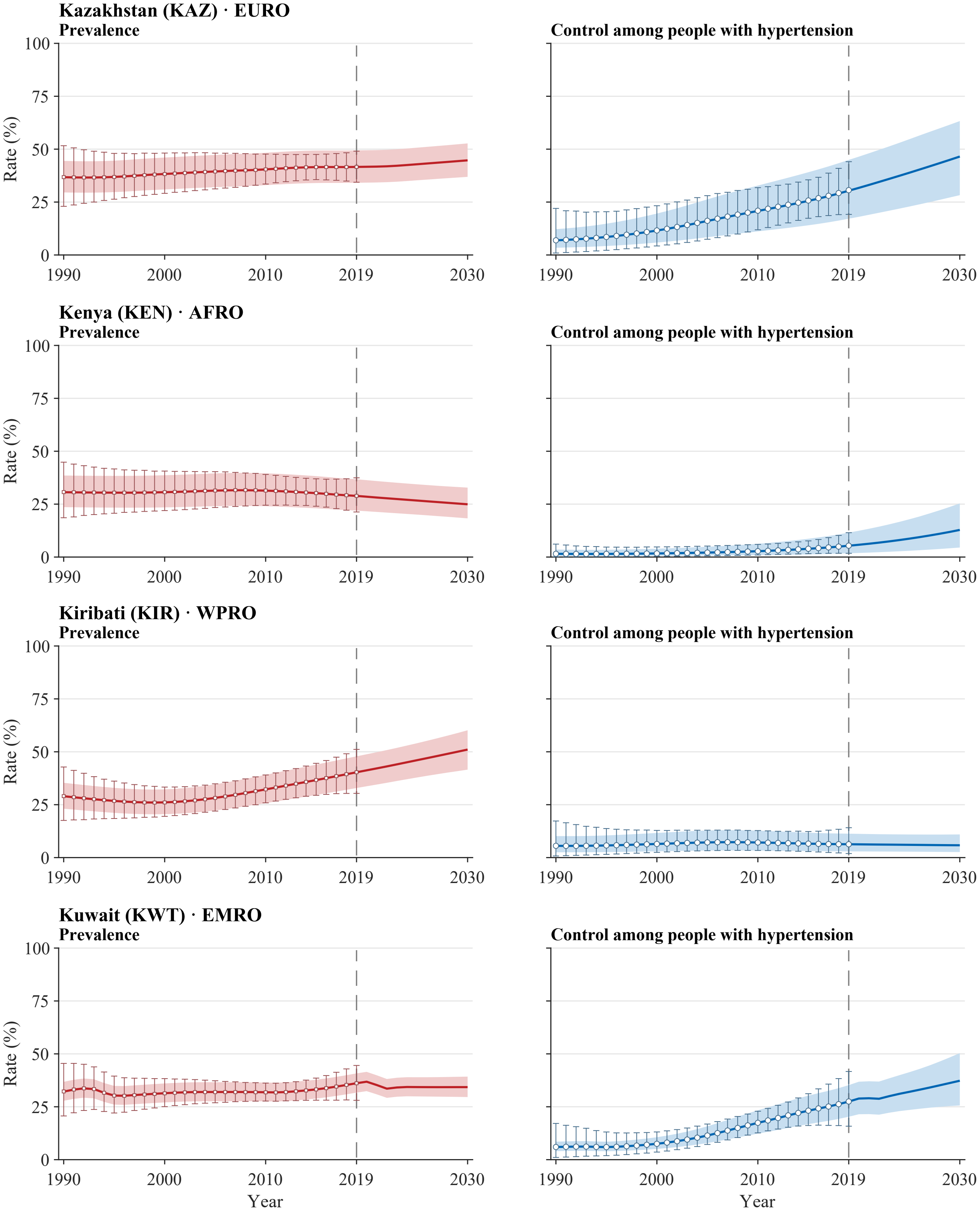


**Legend.** Open squares and error bars in prevalence panels and open circles and error bars in control panels show annual NCD-RisC central estimates with propagated 95% uncertainty intervals for 1990–2019. Solid red and blue lines and shaded bands show model medians and 95% uncertainty intervals for 1990–2030. The vertical dashed line marks 2019, the final year with reported estimates; values for 2020–2030 are forecast. Latent stratum-specific values in 2019 are anchored to the published central estimates. Propagated intervals for the published estimates are wider than the model bands during the historical period by construction of the external uncertainty layer. Model uncertainty represents a persistent level offset. All panels use a common 0–100% scale. Prevalence is defined among all adults aged 30–79 years, whereas control is defined among adults with hypertension. Estimates use CORE-P25 and the prespecified main dependence assumptions.

**Supplementary Figure S3. Country-level trajectories of aggregate hypertension prevalence and control, 1990–2030**
Atlas panel page 25 of 50; countries 97–100 of 200.

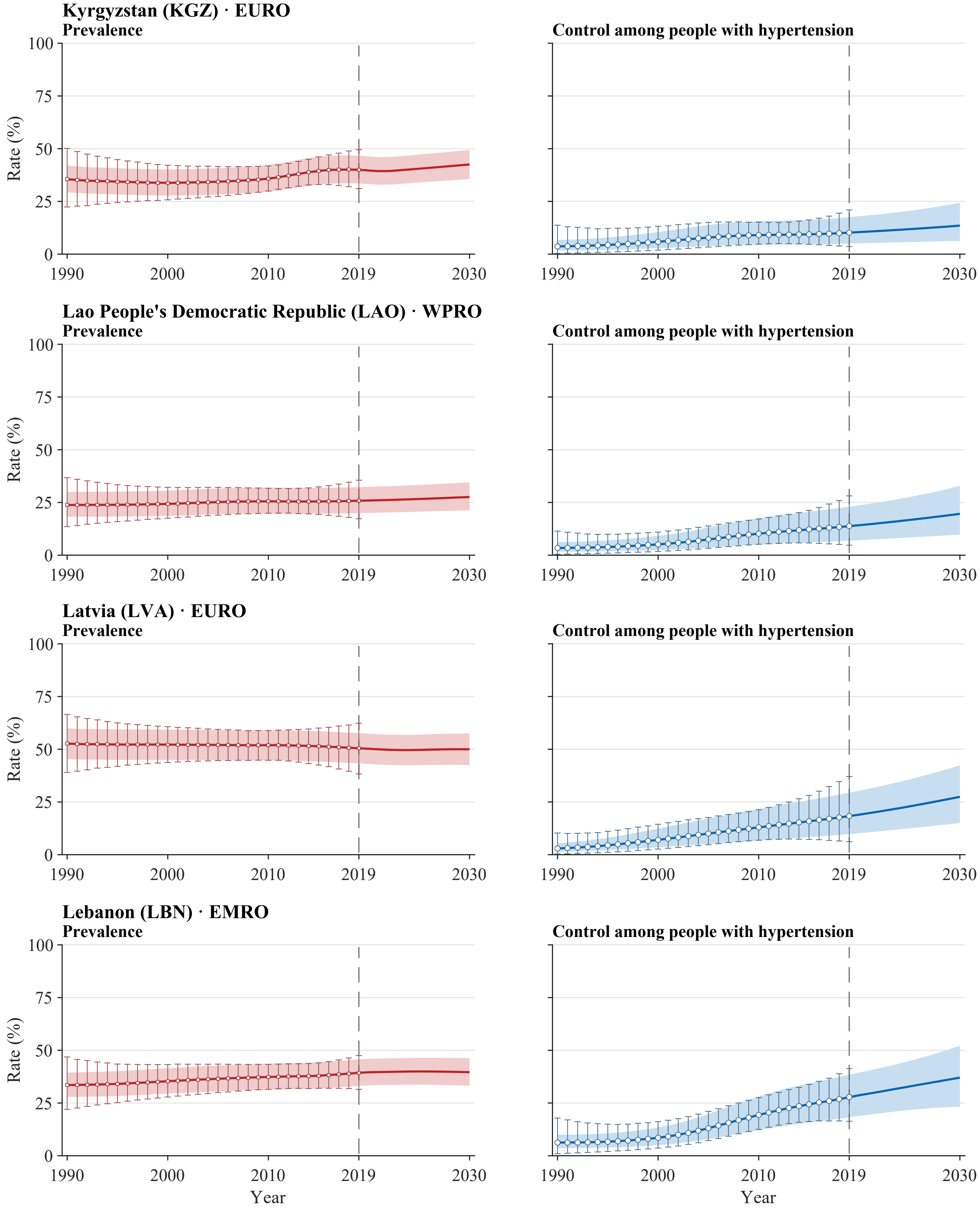


**Legend.** Open squares and error bars in prevalence panels and open circles and error bars in control panels show annual NCD-RisC central estimates with propagated 95% uncertainty intervals for 1990–2019. Solid red and blue lines and shaded bands show model medians and 95% uncertainty intervals for 1990–2030. The vertical dashed line marks 2019, the final year with reported estimates; values for 2020–2030 are forecast. Latent stratum-specific values in 2019 are anchored to the published central estimates. Propagated intervals for the published estimates are wider than the model bands during the historical period by construction of the external uncertainty layer. Model uncertainty represents a persistent level offset. All panels use a common 0–100% scale. Prevalence is defined among all adults aged 30–79 years, whereas control is defined among adults with hypertension. Estimates use CORE-P25 and the prespecified main dependence assumptions.

**Supplementary Figure S3. Country-level trajectories of aggregate hypertension prevalence and control, 1990–2030**
Atlas panel page 26 of 50; countries 101–104 of 200.

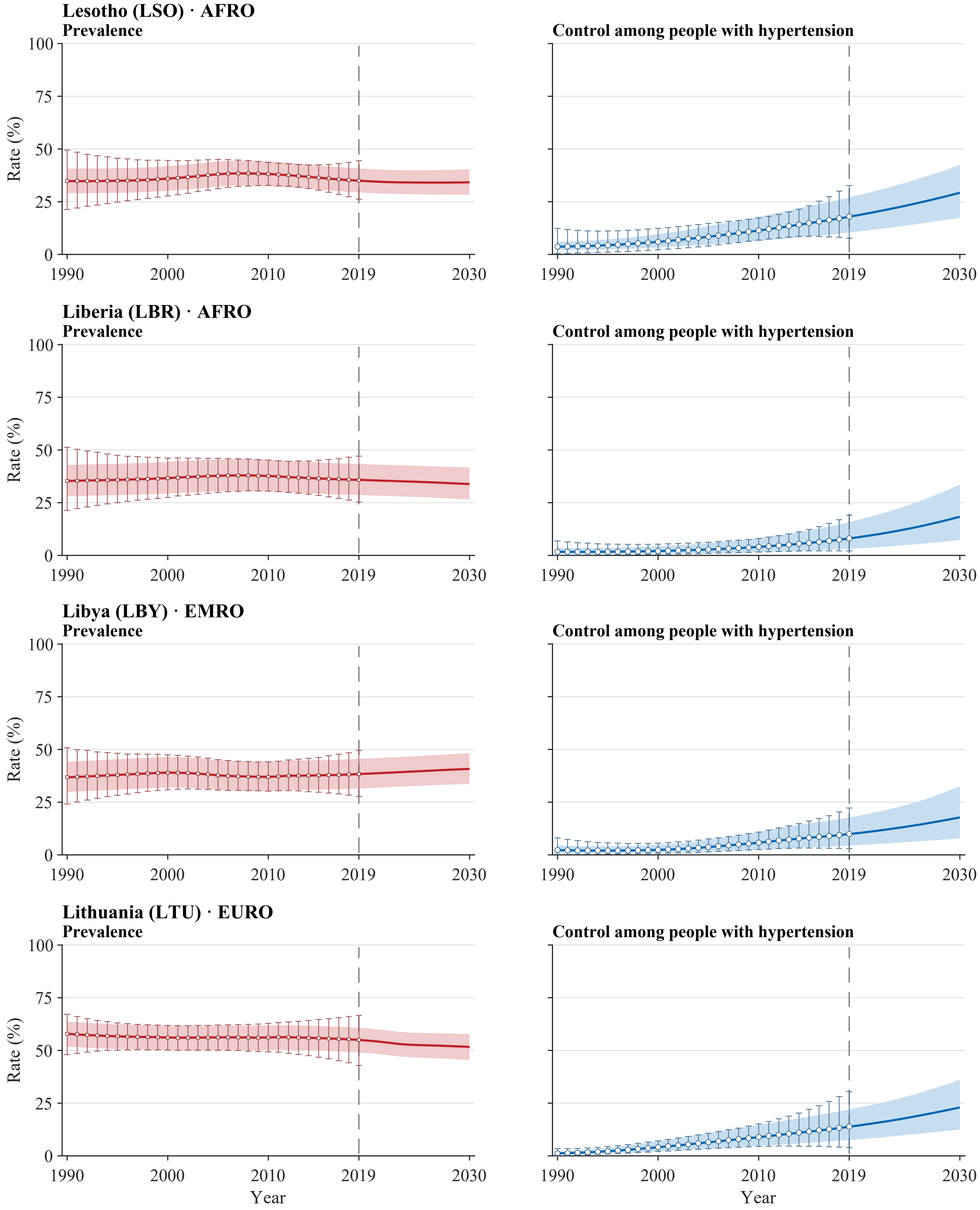


**Legend.** Open squares and error bars in prevalence panels and open circles and error bars in control panels show annual NCD-RisC central estimates with propagated 95% uncertainty intervals for 1990–2019. Solid red and blue lines and shaded bands show model medians and 95% uncertainty intervals for 1990–2030. The vertical dashed line marks 2019, the final year with reported estimates; values for 2020–2030 are forecast. Latent stratum-specific values in 2019 are anchored to the published central estimates. Propagated intervals for the published estimates are wider than the model bands during the historical period by construction of the external uncertainty layer. Model uncertainty represents a persistent level offset. All panels use a common 0–100% scale. Prevalence is defined among all adults aged 30–79 years, whereas control is defined among adults with hypertension. Estimates use CORE-P25 and the prespecified main dependence assumptions.

**Supplementary Figure S3. Country-level trajectories of aggregate hypertension prevalence and control, 1990–2030**
Atlas panel page 27 of 50; countries 105–108 of 200.

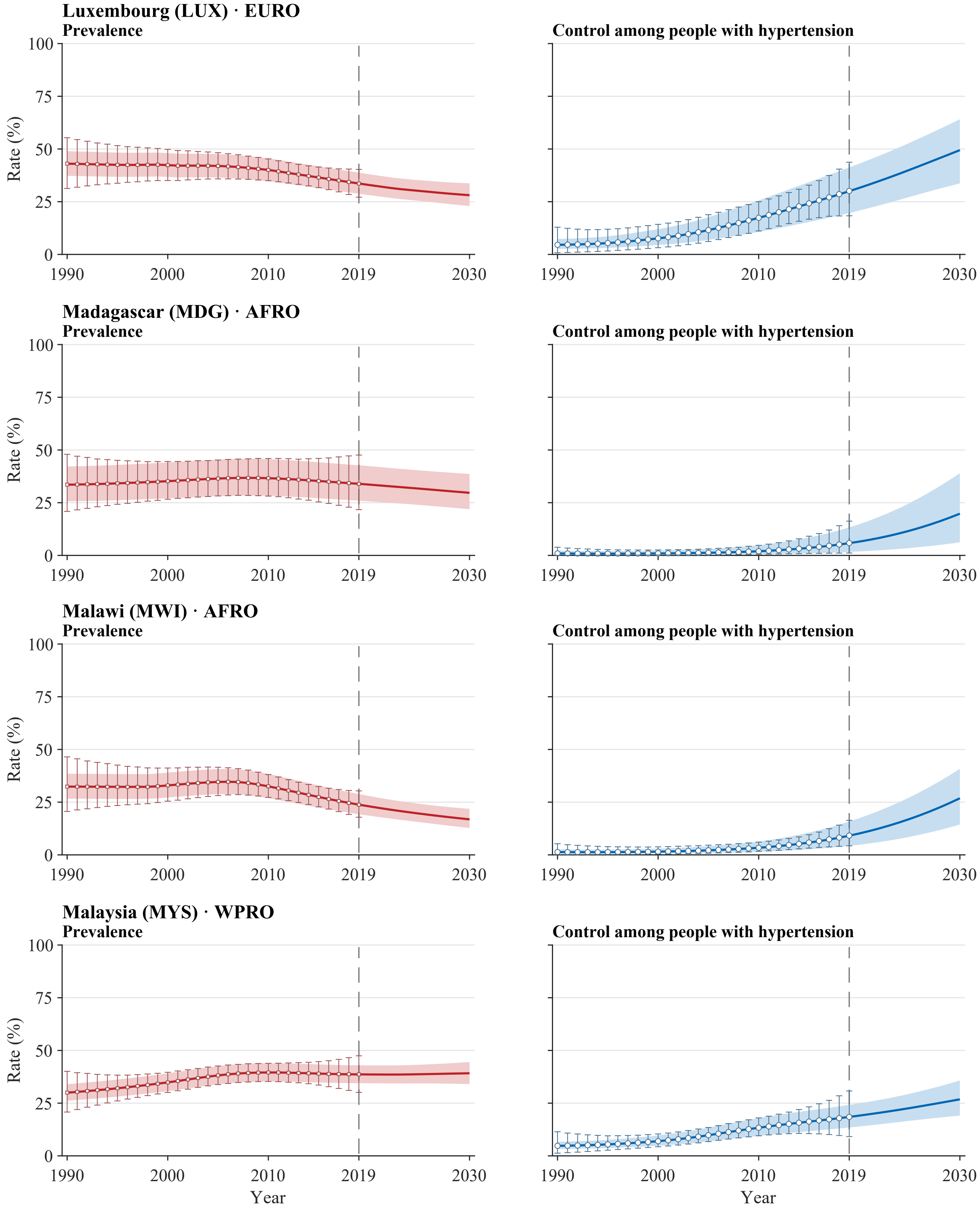


**Legend.** Open squares and error bars in prevalence panels and open circles and error bars in control panels show annual NCD-RisC central estimates with propagated 95% uncertainty intervals for 1990–2019. Solid red and blue lines and shaded bands show model medians and 95% uncertainty intervals for 1990–2030. The vertical dashed line marks 2019, the final year with reported estimates; values for 2020–2030 are forecast. Latent stratum-specific values in 2019 are anchored to the published central estimates. Propagated intervals for the published estimates are wider than the model bands during the historical period by construction of the external uncertainty layer. Model uncertainty represents a persistent level offset. All panels use a common 0–100% scale. Prevalence is defined among all adults aged 30–79 years, whereas control is defined among adults with hypertension. Estimates use CORE-P25 and the prespecified main dependence assumptions.

**Supplementary Figure S3. Country-level trajectories of aggregate hypertension prevalence and control, 1990–2030**
Atlas panel page 28 of 50; countries 109–112 of 200.

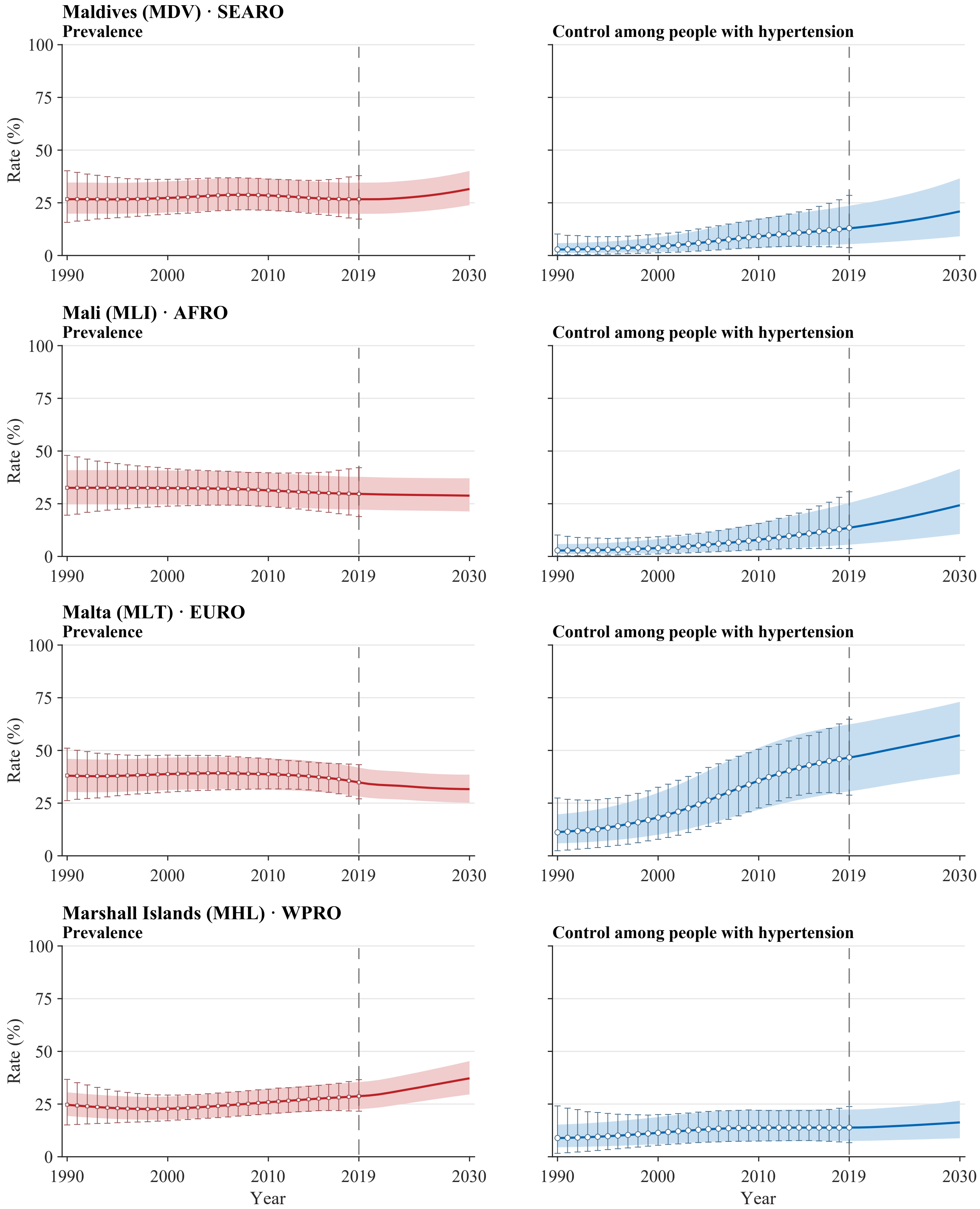


**Legend.** Open squares and error bars in prevalence panels and open circles and error bars in control panels show annual NCD-RisC central estimates with propagated 95% uncertainty intervals for 1990–2019. Solid red and blue lines and shaded bands show model medians and 95% uncertainty intervals for 1990–2030. The vertical dashed line marks 2019, the final year with reported estimates; values for 2020–2030 are forecast. Latent stratum-specific values in 2019 are anchored to the published central estimates. Propagated intervals for the published estimates are wider than the model bands during the historical period by construction of the external uncertainty layer. Model uncertainty represents a persistent level offset. All panels use a common 0–100% scale. Prevalence is defined among all adults aged 30–79 years, whereas control is defined among adults with hypertension. Estimates use CORE-P25 and the prespecified main dependence assumptions.

**Supplementary Figure S3. Country-level trajectories of aggregate hypertension prevalence and control, 1990–2030**
Atlas panel page 29 of 50; countries 113–116 of 200.

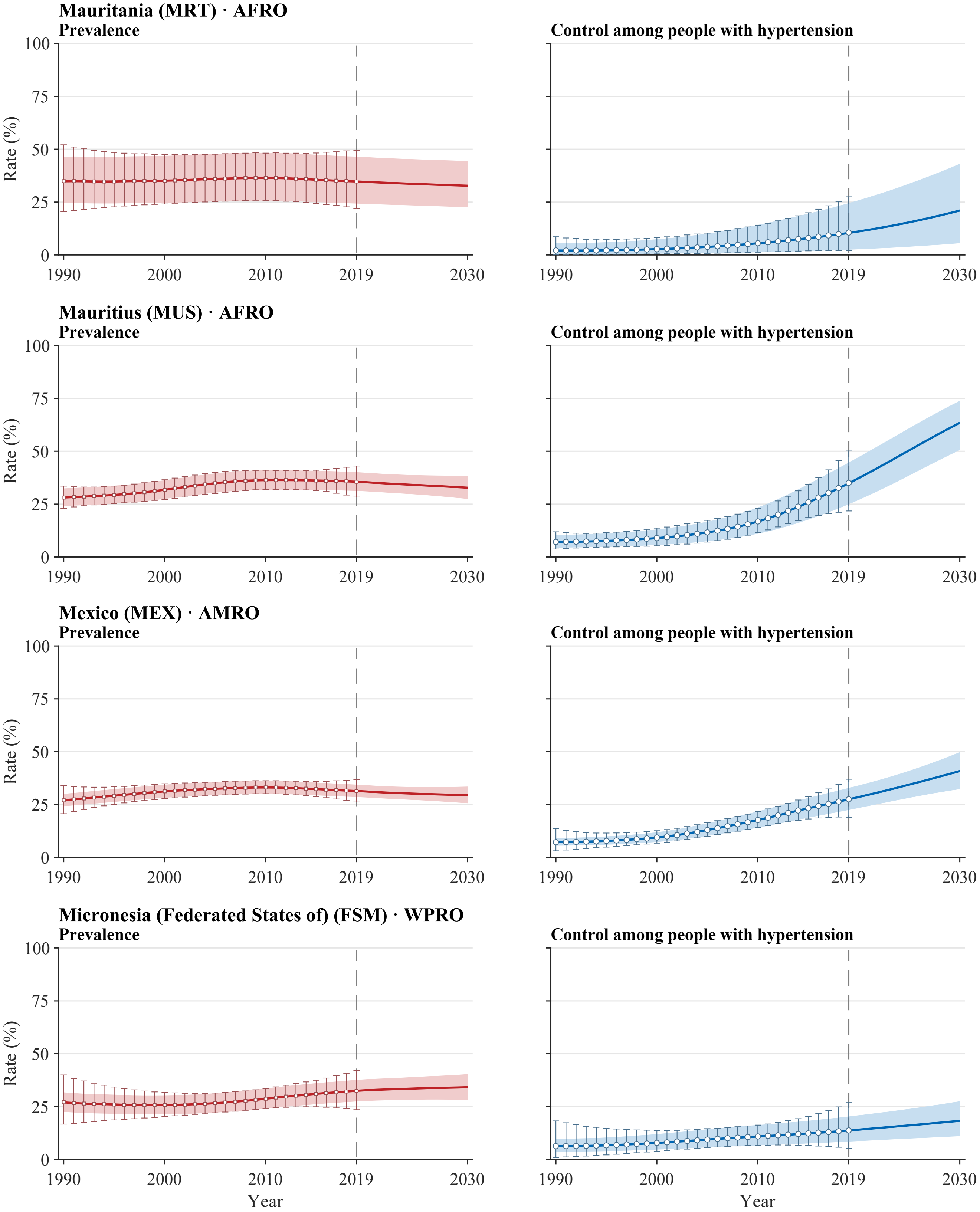


**Legend.** Open squares and error bars in prevalence panels and open circles and error bars in control panels show annual NCD-RisC central estimates with propagated 95% uncertainty intervals for 1990–2019. Solid red and blue lines and shaded bands show model medians and 95% uncertainty intervals for 1990–2030. The vertical dashed line marks 2019, the final year with reported estimates; values for 2020–2030 are forecast. Latent stratum-specific values in 2019 are anchored to the published central estimates. Propagated intervals for the published estimates are wider than the model bands during the historical period by construction of the external uncertainty layer. Model uncertainty represents a persistent level offset. All panels use a common 0–100% scale. Prevalence is defined among all adults aged 30–79 years, whereas control is defined among adults with hypertension. Estimates use CORE-P25 and the prespecified main dependence assumptions.

**Supplementary Figure S3. Country-level trajectories of aggregate hypertension prevalence and control, 1990–2030**

Atlas panel page 30 of 50; countries 117–120 of 200.

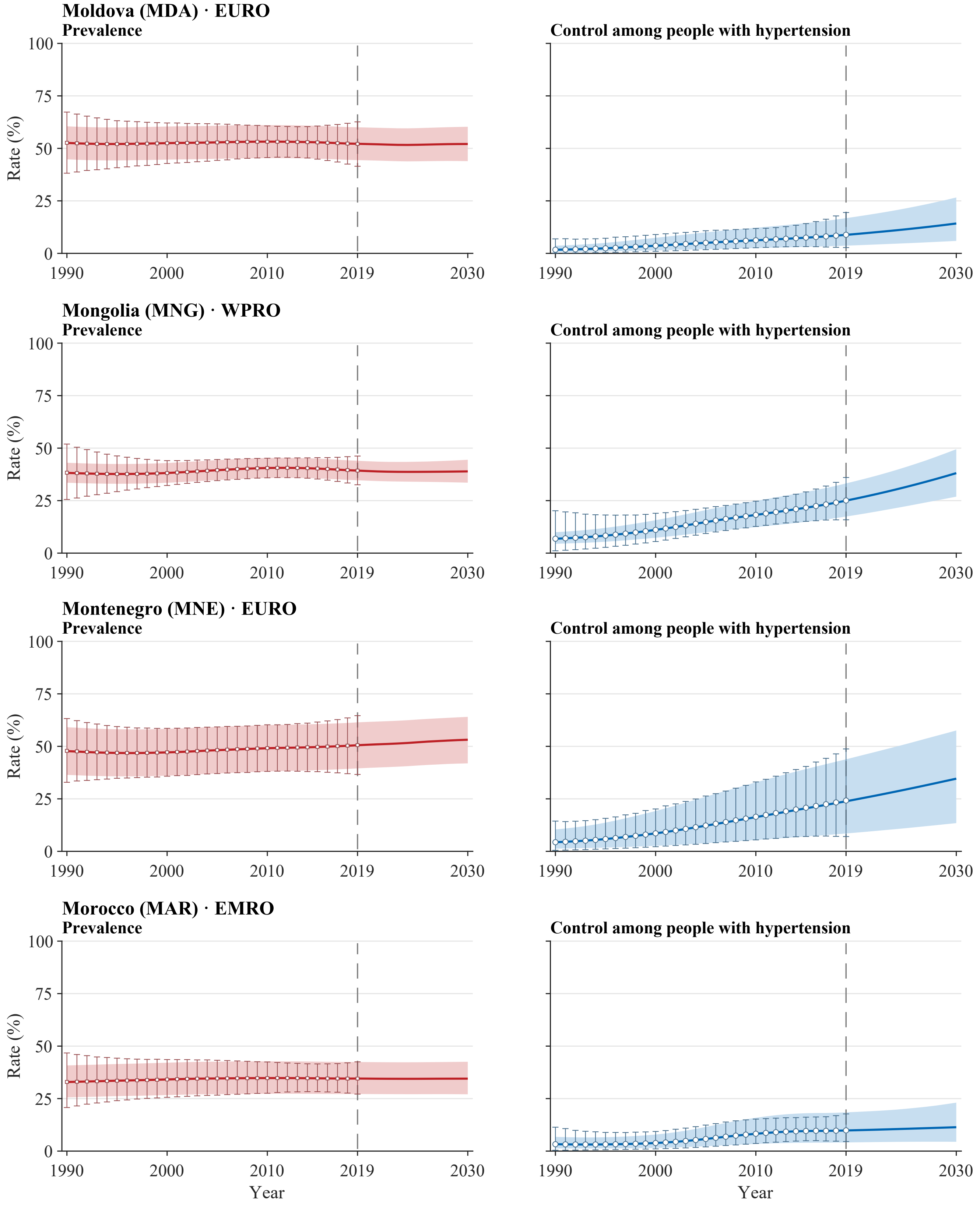


**Legend.** Open squares and error bars in prevalence panels and open circles and error bars in control panels show annual NCD-RisC central estimates with propagated 95% uncertainty intervals for 1990–2019. Solid red and blue lines and shaded bands show model medians and 95% uncertainty intervals for 1990–2030. The vertical dashed line marks 2019, the final year with reported estimates; values for 2020–2030 are forecast. Latent stratum-specific values in 2019 are anchored to the published central estimates. Propagated intervals for the published estimates are wider than the model bands during the historical period by construction of the external uncertainty layer. Model uncertainty represents a persistent level offset. All panels use a common 0–100% scale. Prevalence is defined among all adults aged 30–79 years, whereas control is defined among adults with hypertension. Estimates use CORE-P25 and the prespecified main dependence assumptions.

**Supplementary Figure S3. Country-level trajectories of aggregate hypertension prevalence and control, 1990–2030**

Atlas panel page 31 of 50; countries 121–124 of 200.

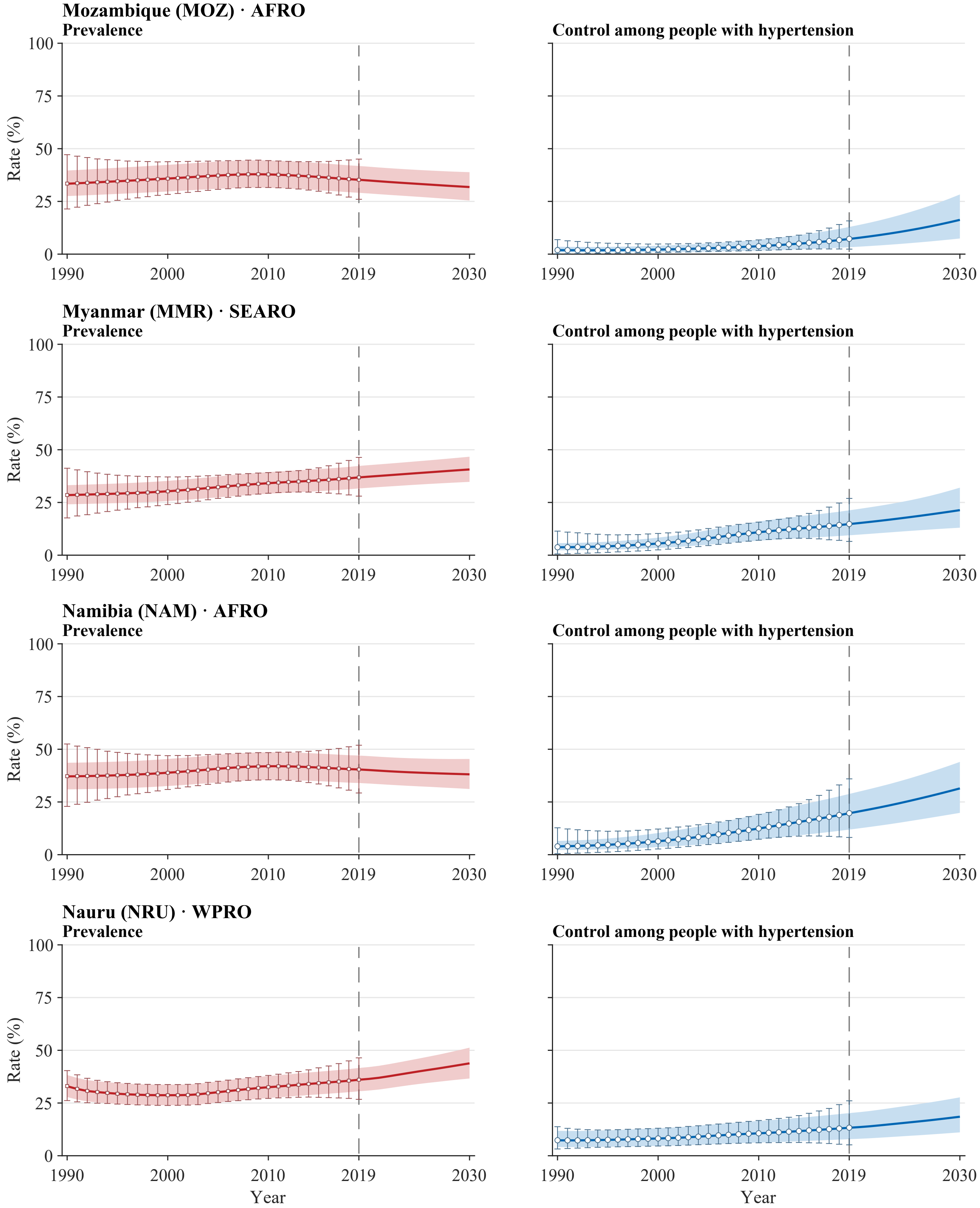


**Legend.** Open squares and error bars in prevalence panels and open circles and error bars in control panels show annual NCD-RisC central estimates with propagated 95% uncertainty intervals for 1990–2019. Solid red and blue lines and shaded bands show model medians and 95% uncertainty intervals for 1990–2030. The vertical dashed line marks 2019, the final year with reported estimates; values for 2020–2030 are forecast. Latent stratum-specific values in 2019 are anchored to the published central estimates. Propagated intervals for the published estimates are wider than the model bands during the historical period by construction of the external uncertainty layer. Model uncertainty represents a persistent level offset. All panels use a common 0–100% scale. Prevalence is defined among all adults aged 30–79 years, whereas control is defined among adults with hypertension. Estimates use CORE-P25 and the prespecified main dependence assumptions.

**Supplementary Figure S3. Country-level trajectories of aggregate hypertension prevalence and control, 1990–2030**

Atlas panel page 32 of 50; countries 125–128 of 200.

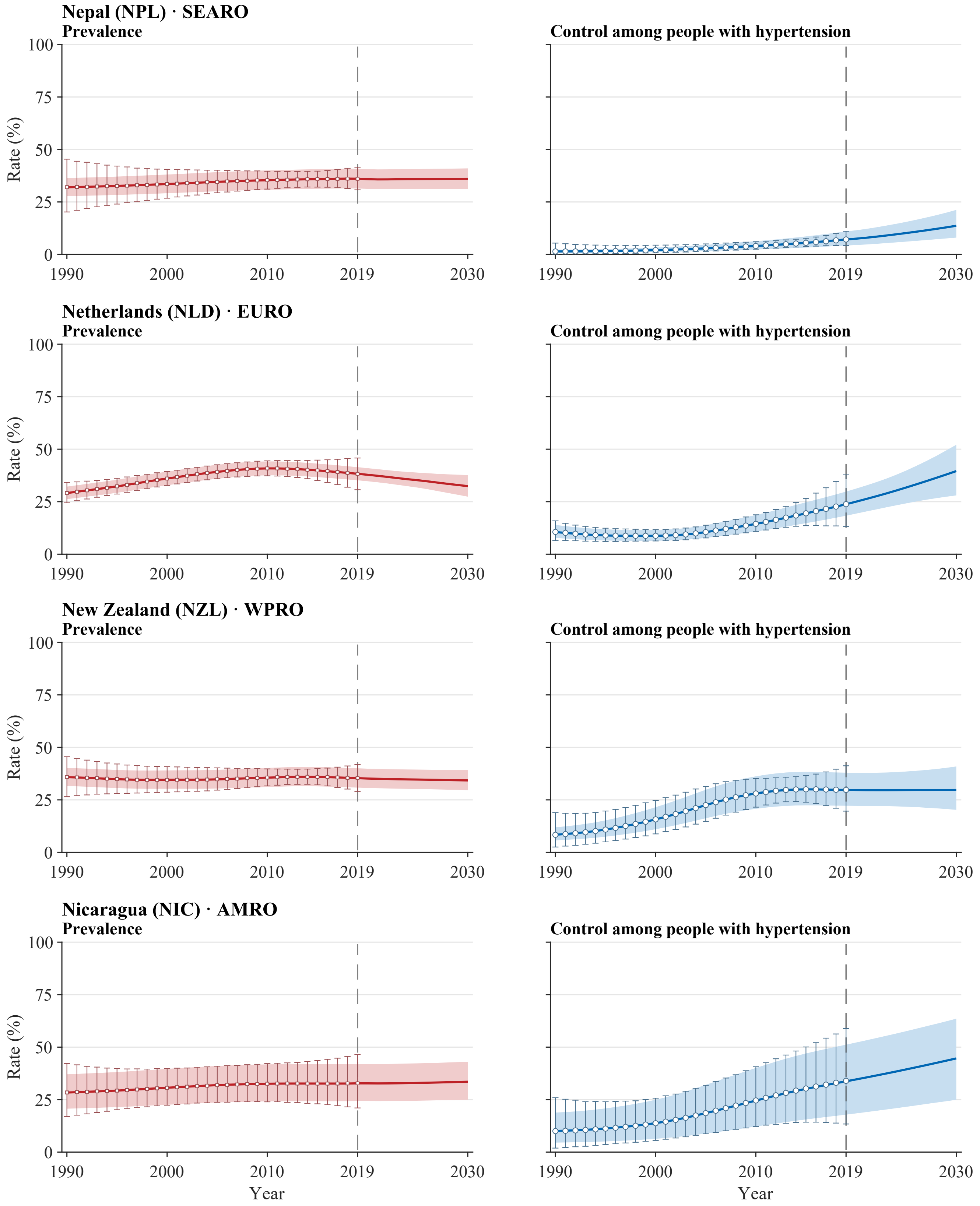


**Legend.** Open squares and error bars in prevalence panels and open circles and error bars in control panels show annual NCD-RisC central estimates with propagated 95% uncertainty intervals for 1990–2019. Solid red and blue lines and shaded bands show model medians and 95% uncertainty intervals for 1990–2030. The vertical dashed line marks 2019, the final year with reported estimates; values for 2020–2030 are forecast. Latent stratum-specific values in 2019 are anchored to the published central estimates. Propagated intervals for the published estimates are wider than the model bands during the historical period by construction of the external uncertainty layer. Model uncertainty represents a persistent level offset. All panels use a common 0–100% scale. Prevalence is defined among all adults aged 30–79 years, whereas control is defined among adults with hypertension. Estimates use CORE-P25 and the prespecified main dependence assumptions.

**Supplementary Figure S3. Country-level trajectories of aggregate hypertension prevalence and control, 1990–2030**

Atlas panel page 33 of 50; countries 129–132 of 200.

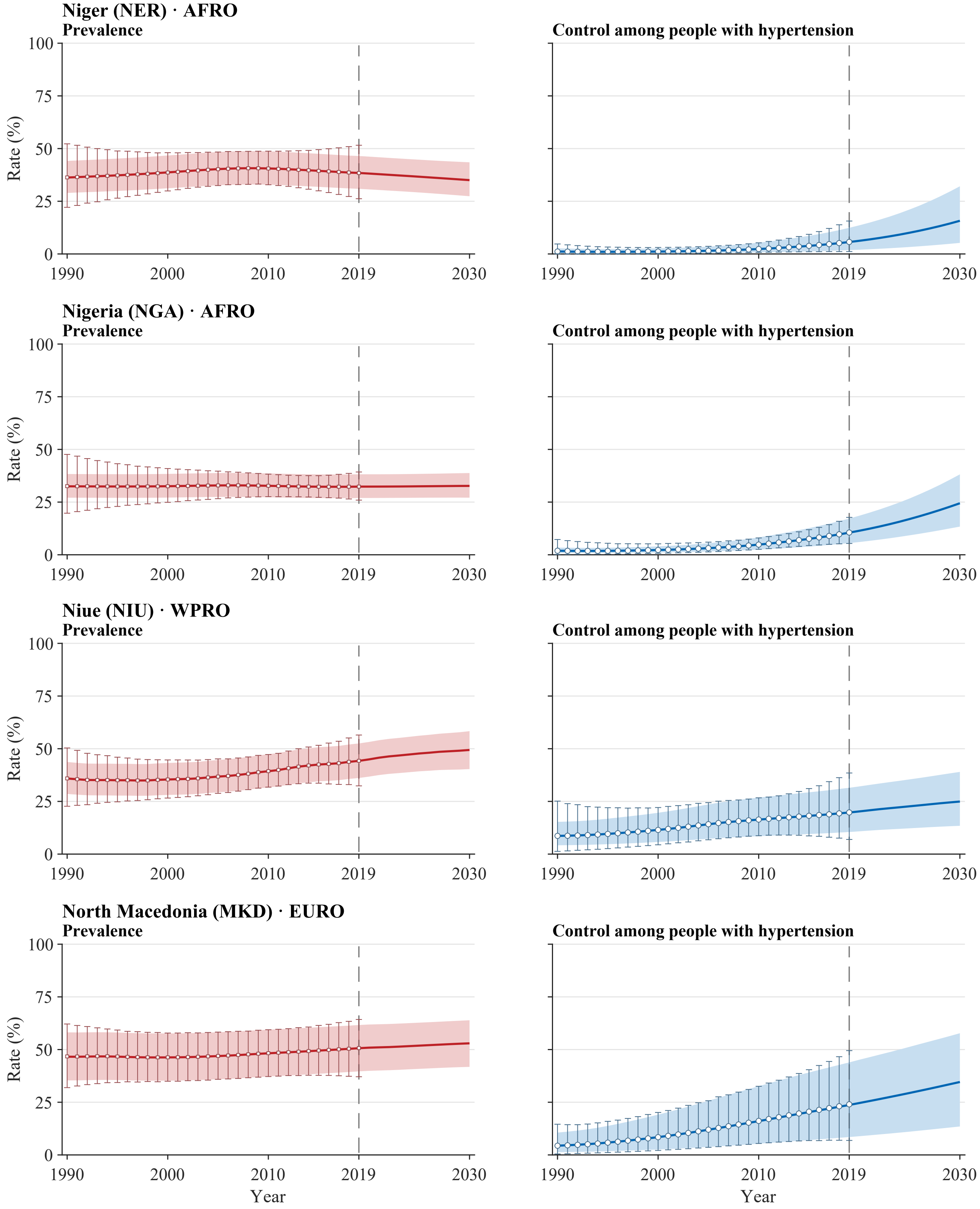


**Legend.** Open squares and error bars in prevalence panels and open circles and error bars in control panels show annual NCD-RisC central estimates with propagated 95% uncertainty intervals for 1990–2019. Solid red and blue lines and shaded bands show model medians and 95% uncertainty intervals for 1990–2030. The vertical dashed line marks 2019, the final year with reported estimates; values for 2020–2030 are forecast. Latent stratum-specific values in 2019 are anchored to the published central estimates. Propagated intervals for the published estimates are wider than the model bands during the historical period by construction of the external uncertainty layer. Model uncertainty represents a persistent level offset. All panels use a common 0–100% scale. Prevalence is defined among all adults aged 30–79 years, whereas control is defined among adults with hypertension. Estimates use CORE-P25 and the prespecified main dependence assumptions.

**Supplementary Figure S3. Country-level trajectories of aggregate hypertension prevalence and control, 1990–2030**
Atlas panel page 34 of 50; countries 133–136 of 200.

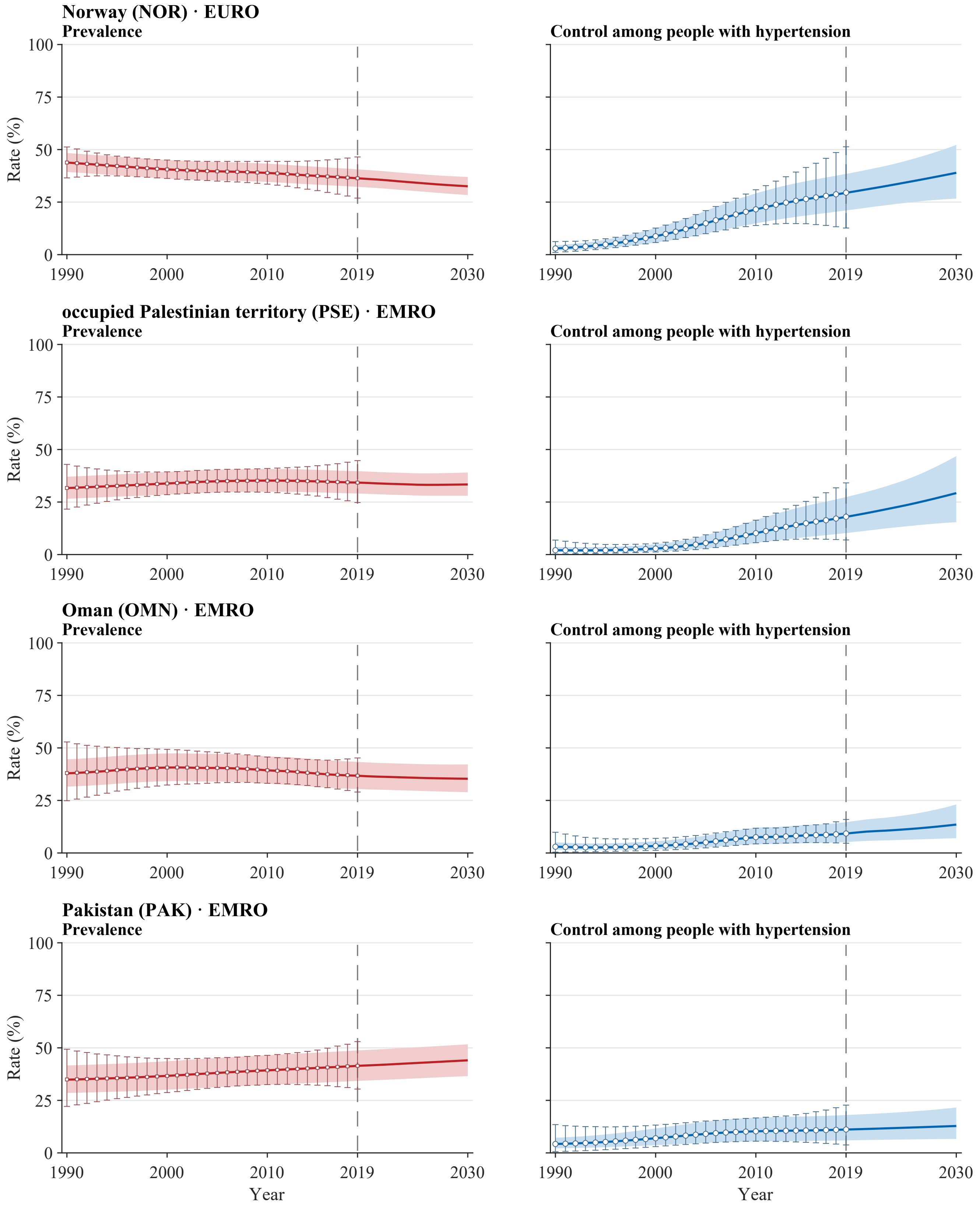


**Legend.** Open squares and error bars in prevalence panels and open circles and error bars in control panels show annual NCD-RisC central estimates with propagated 95% uncertainty intervals for 1990–2019. Solid red and blue lines and shaded bands show model medians and 95% uncertainty intervals for 1990–2030. The vertical dashed line marks 2019, the final year with reported estimates; values for 2020–2030 are forecast. Latent stratum-specific values in 2019 are anchored to the published central estimates. Propagated intervals for the published estimates are wider than the model bands during the historical period by construction of the external uncertainty layer. Model uncertainty represents a persistent level offset. All panels use a common 0–100% scale. Prevalence is defined among all adults aged 30–79 years, whereas control is defined among adults with hypertension. Estimates use CORE-P25 and the prespecified main dependence assumptions.

**Supplementary Figure S3. Country-level trajectories of aggregate hypertension prevalence and control, 1990–2030**

Atlas panel page 35 of 50; countries 137–140 of 200.

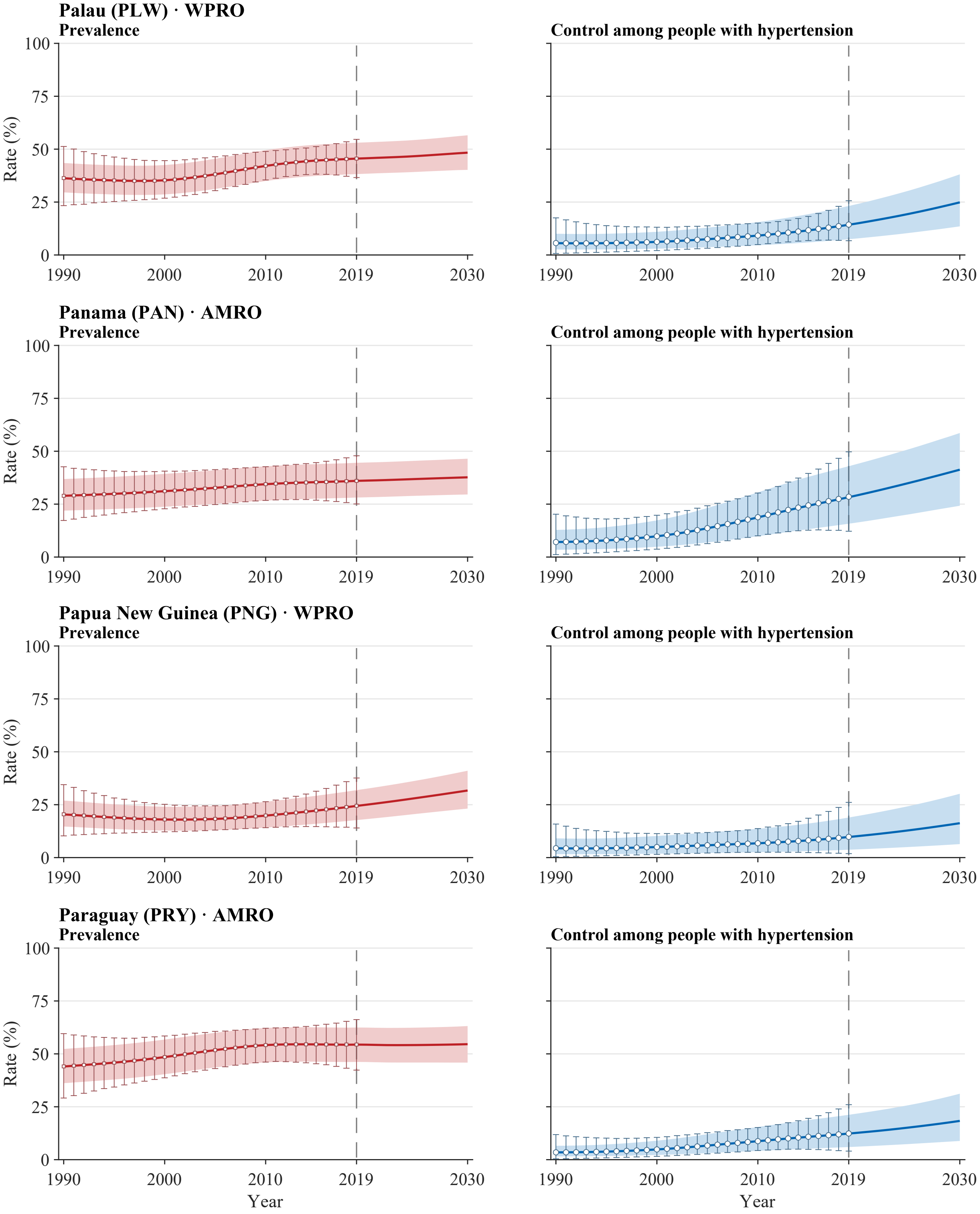


**Legend.** Open squares and error bars in prevalence panels and open circles and error bars in control panels show annual NCD-RisC central estimates with propagated 95% uncertainty intervals for 1990–2019. Solid red and blue lines and shaded bands show model medians and 95% uncertainty intervals for 1990–2030. The vertical dashed line marks 2019, the final year with reported estimates; values for 2020–2030 are forecast. Latent stratum-specific values in 2019 are anchored to the published central estimates. Propagated intervals for the published estimates are wider than the model bands during the historical period by construction of the external uncertainty layer. Model uncertainty represents a persistent level offset. All panels use a common 0–100% scale. Prevalence is defined among all adults aged 30–79 years, whereas control is defined among adults with hypertension. Estimates use CORE-P25 and the prespecified main dependence assumptions.

**Supplementary Figure S3. Country-level trajectories of aggregate hypertension prevalence and control, 1990–2030**
Atlas panel page 36 of 50; countries 141–144 of 200.

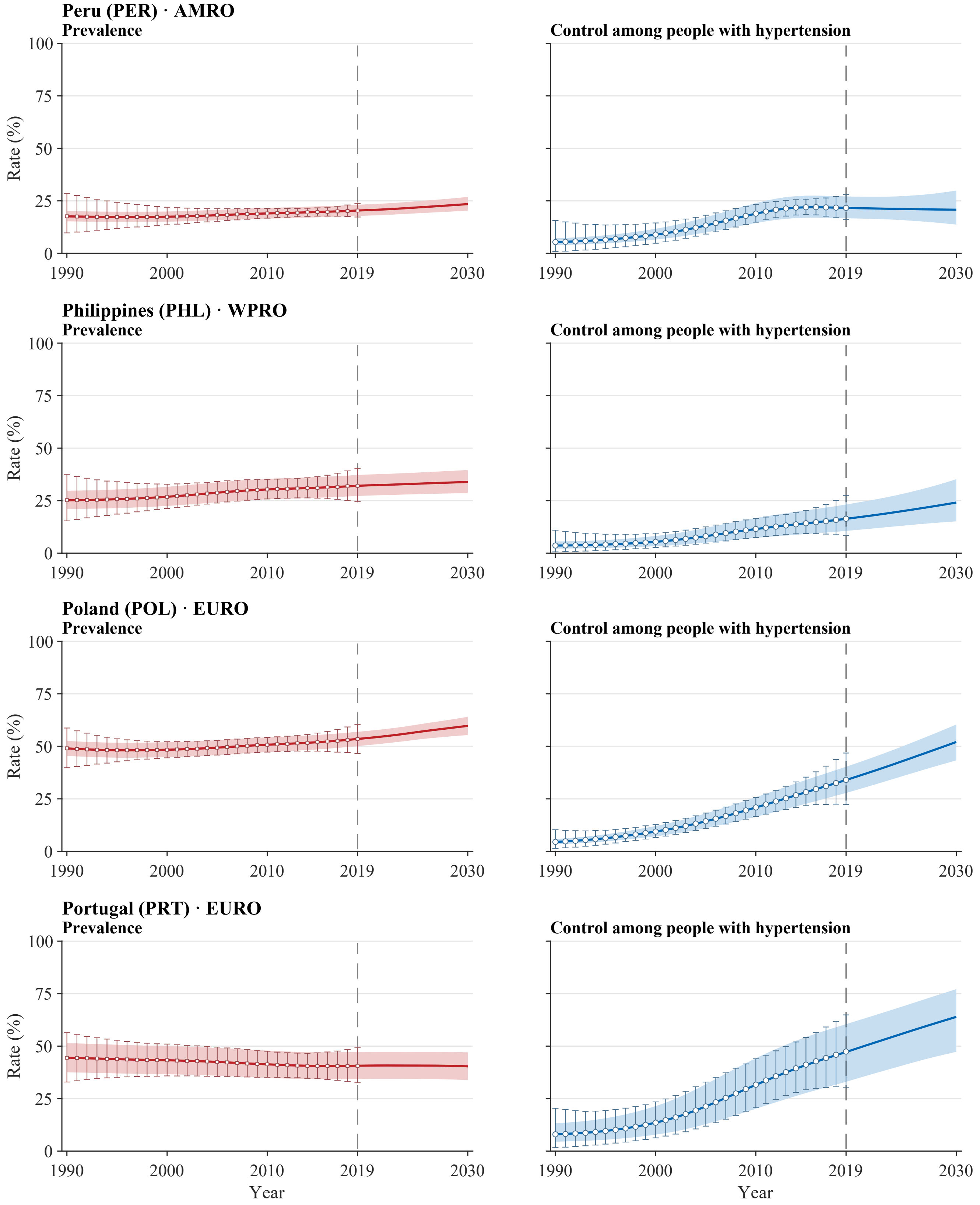


**Legend.** Open squares and error bars in prevalence panels and open circles and error bars in control panels show annual NCD-RisC central estimates with propagated 95% uncertainty intervals for 1990–2019. Solid red and blue lines and shaded bands show model medians and 95% uncertainty intervals for 1990–2030. The vertical dashed line marks 2019, the final year with reported estimates; values for 2020–2030 are forecast. Latent stratum-specific values in 2019 are anchored to the published central estimates. Propagated intervals for the published estimates are wider than the model bands during the historical period by construction of the external uncertainty layer. Model uncertainty represents a persistent level offset. All panels use a common 0–100% scale. Prevalence is defined among all adults aged 30–79 years, whereas control is defined among adults with hypertension. Estimates use CORE-P25 and the prespecified main dependence assumptions.

**Supplementary Figure S3. Country-level trajectories of aggregate hypertension prevalence and control, 1990–2030**

Atlas panel page 37 of 50; countries 145–148 of 200.

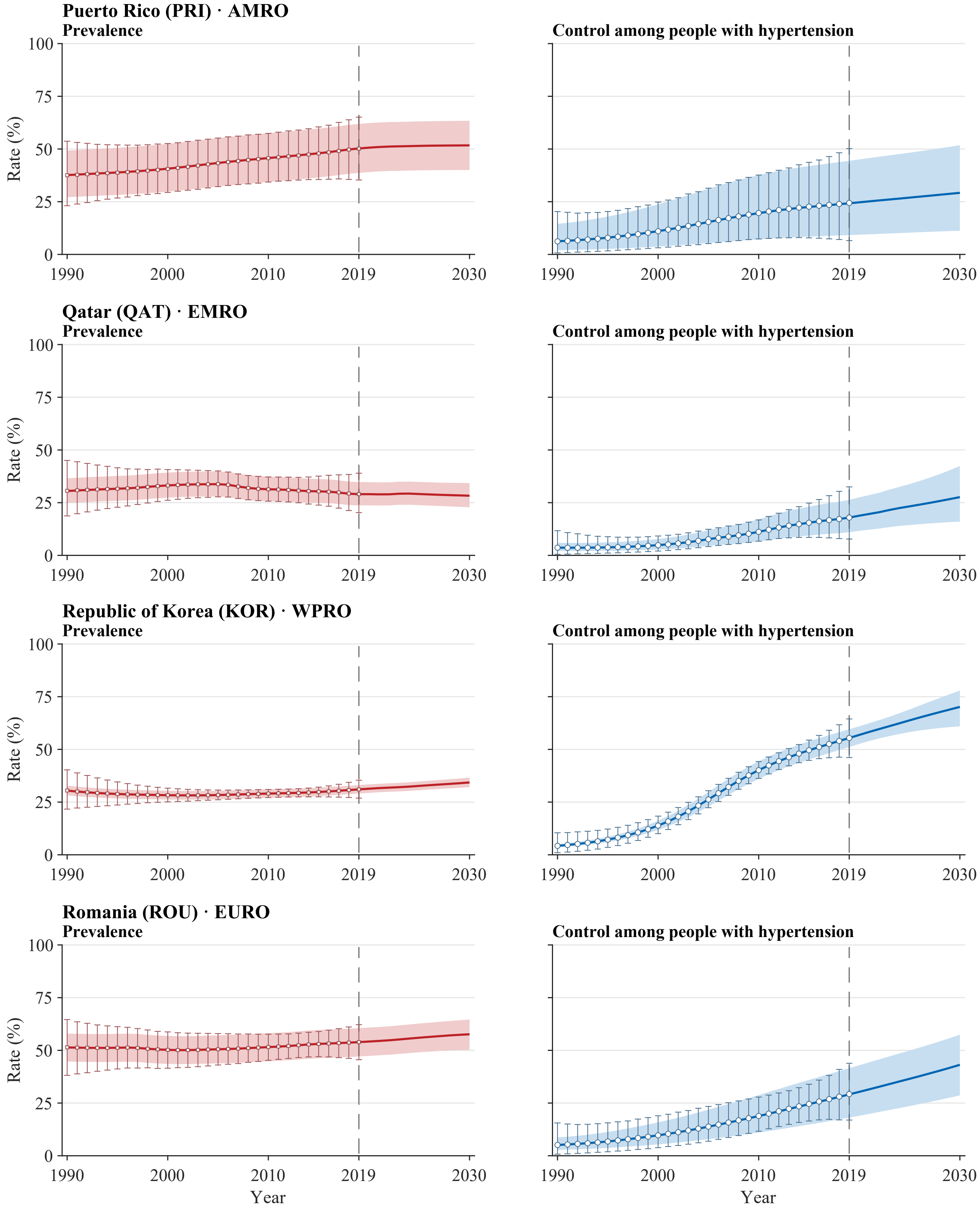


**Legend.** Open squares and error bars in prevalence panels and open circles and error bars in control panels show annual NCD-RisC central estimates with propagated 95% uncertainty intervals for 1990–2019. Solid red and blue lines and shaded bands show model medians and 95% uncertainty intervals for 1990–2030. The vertical dashed line marks 2019, the final year with reported estimates; values for 2020–2030 are forecast. Latent stratum-specific values in 2019 are anchored to the published central estimates. Propagated intervals for the published estimates are wider than the model bands during the historical period by construction of the external uncertainty layer. Model uncertainty represents a persistent level offset. All panels use a common 0–100% scale. Prevalence is defined among all adults aged 30–79 years, whereas control is defined among adults with hypertension. Estimates use CORE-P25 and the prespecified main dependence assumptions.

**Supplementary Figure S3. Country-level trajectories of aggregate hypertension prevalence and control, 1990–2030**

Atlas panel page 38 of 50; countries 149–152 of 200.

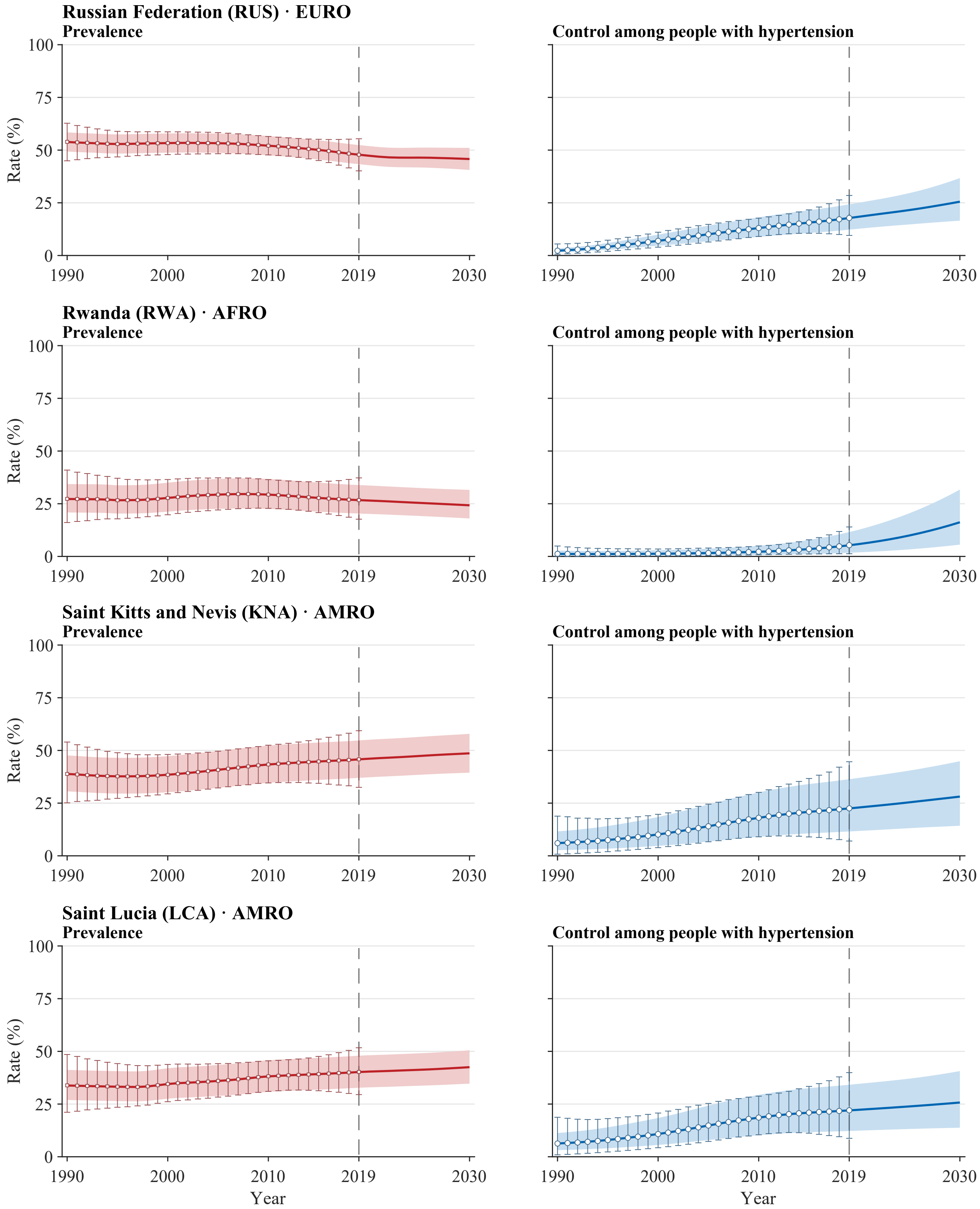


**Legend.** Open squares and error bars in prevalence panels and open circles and error bars in control panels show annual NCD-RisC central estimates with propagated 95% uncertainty intervals for 1990–2019. Solid red and blue lines and shaded bands show model medians and 95% uncertainty intervals for 1990–2030. The vertical dashed line marks 2019, the final year with reported estimates; values for 2020–2030 are forecast. Latent stratum-specific values in 2019 are anchored to the published central estimates. Propagated intervals for the published estimates are wider than the model bands during the historical period by construction of the external uncertainty layer. Model uncertainty represents a persistent level offset. All panels use a common 0–100% scale. Prevalence is defined among all adults aged 30–79 years, whereas control is defined among adults with hypertension. Estimates use CORE-P25 and the prespecified main dependence assumptions.

**Supplementary Figure S3. Country-level trajectories of aggregate hypertension prevalence and control, 1990–2030**
Atlas panel page 39 of 50; countries 153–156 of 200.

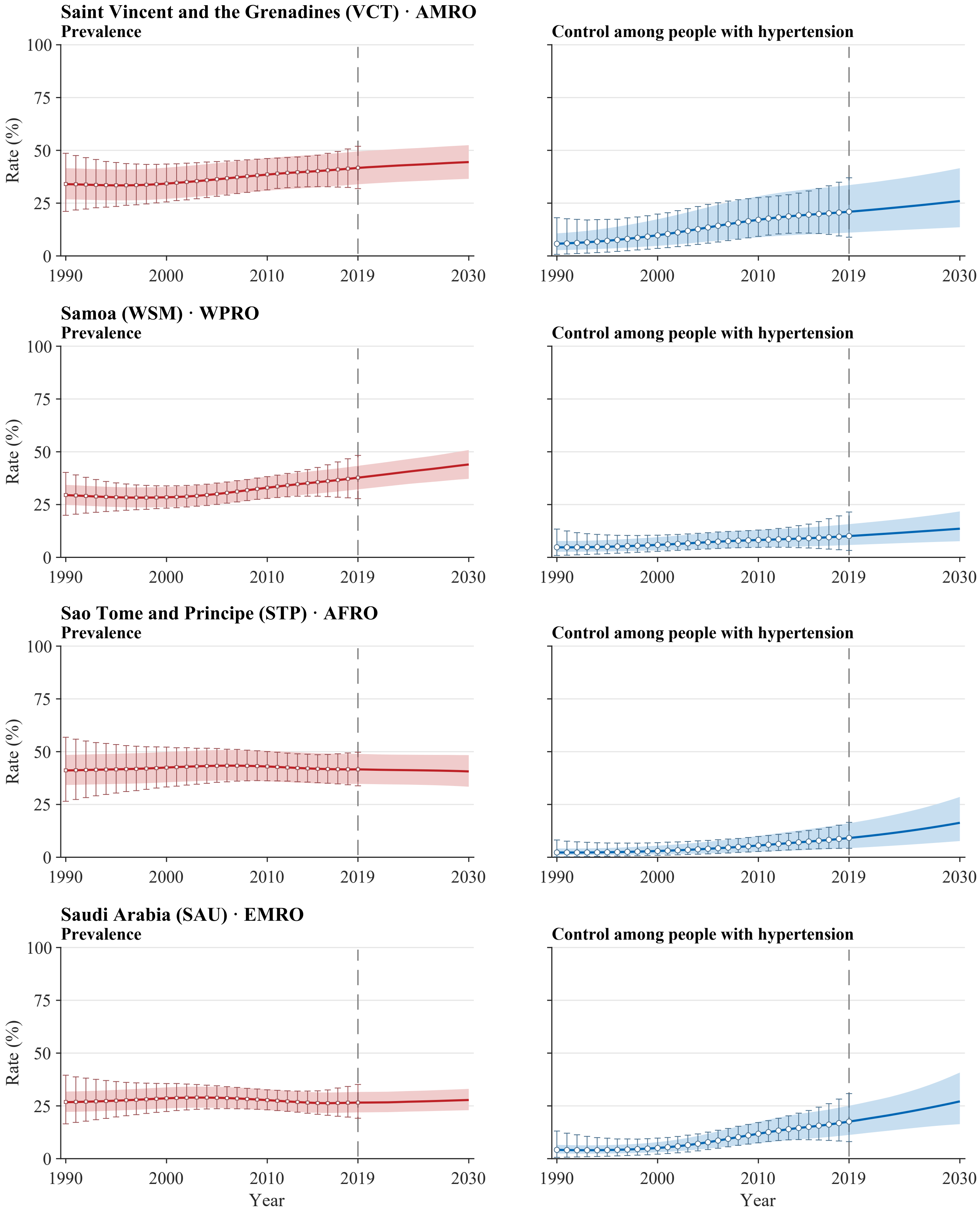


**Legend.** Open squares and error bars in prevalence panels and open circles and error bars in control panels show annual NCD-RisC central estimates with propagated 95% uncertainty intervals for 1990–2019. Solid red and blue lines and shaded bands show model medians and 95% uncertainty intervals for 1990–2030. The vertical dashed line marks 2019, the final year with reported estimates; values for 2020–2030 are forecast. Latent stratum-specific values in 2019 are anchored to the published central estimates. Propagated intervals for the published estimates are wider than the model bands during the historical period by construction of the external uncertainty layer. Model uncertainty represents a persistent level offset. All panels use a common 0–100% scale. Prevalence is defined among all adults aged 30–79 years, whereas control is defined among adults with hypertension. Estimates use CORE-P25 and the prespecified main dependence assumptions.

**Supplementary Figure S3. Country-level trajectories of aggregate hypertension prevalence and control, 1990–2030**

Atlas panel page 40 of 50; countries 157–160 of 200.

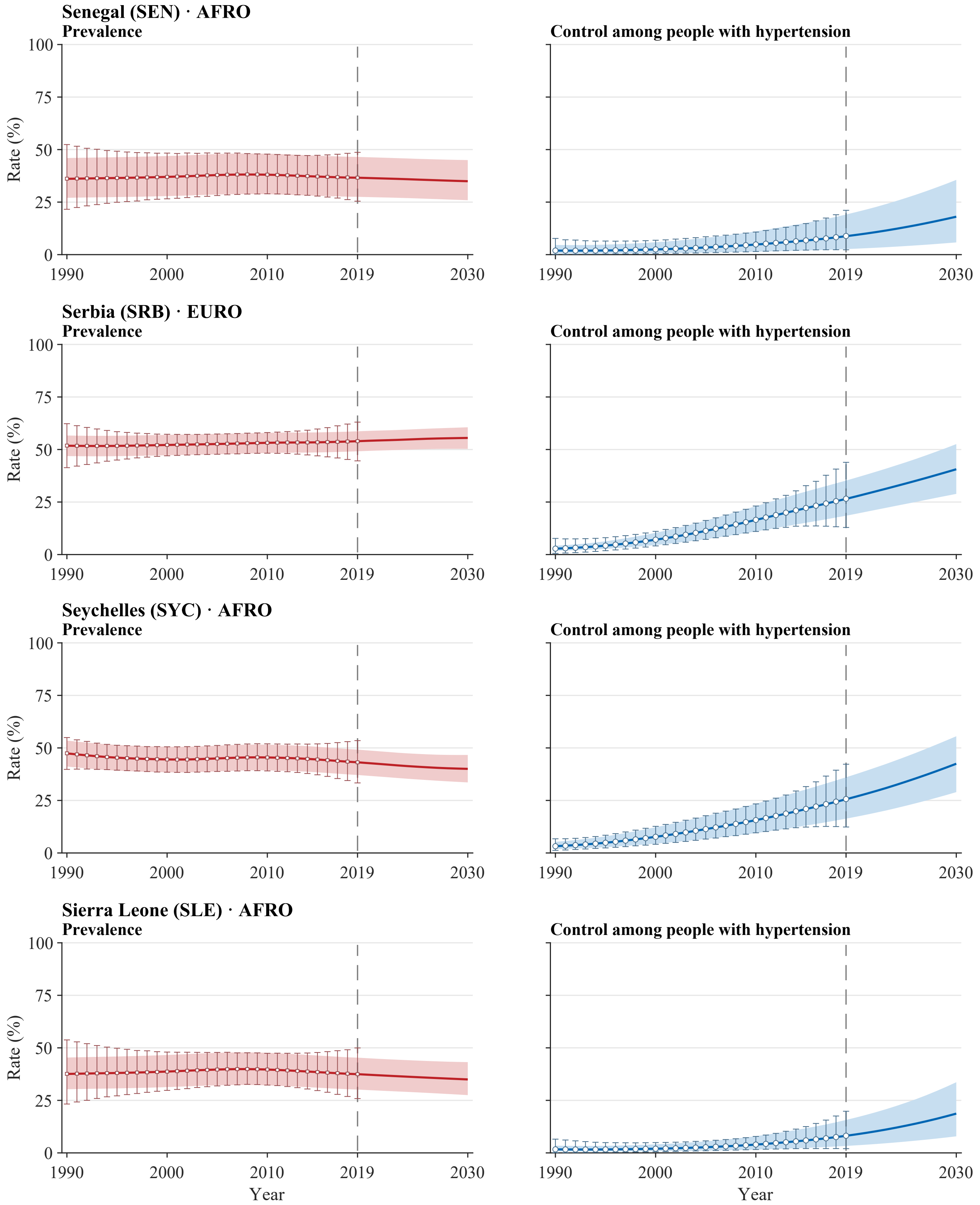


**Legend.** Open squares and error bars in prevalence panels and open circles and error bars in control panels show annual NCD-RisC central estimates with propagated 95% uncertainty intervals for 1990–2019. Solid red and blue lines and shaded bands show model medians and 95% uncertainty intervals for 1990–2030. The vertical dashed line marks 2019, the final year with reported estimates; values for 2020–2030 are forecast. Latent stratum-specific values in 2019 are anchored to the published central estimates. Propagated intervals for the published estimates are wider than the model bands during the historical period by construction of the external uncertainty layer. Model uncertainty represents a persistent level offset. All panels use a common 0–100% scale. Prevalence is defined among all adults aged 30–79 years, whereas control is defined among adults with hypertension. Estimates use CORE-P25 and the prespecified main dependence assumptions.

**Supplementary Figure S3. Country-level trajectories of aggregate hypertension prevalence and control, 1990–2030**

Atlas panel page 41 of 50; countries 161–164 of 200.

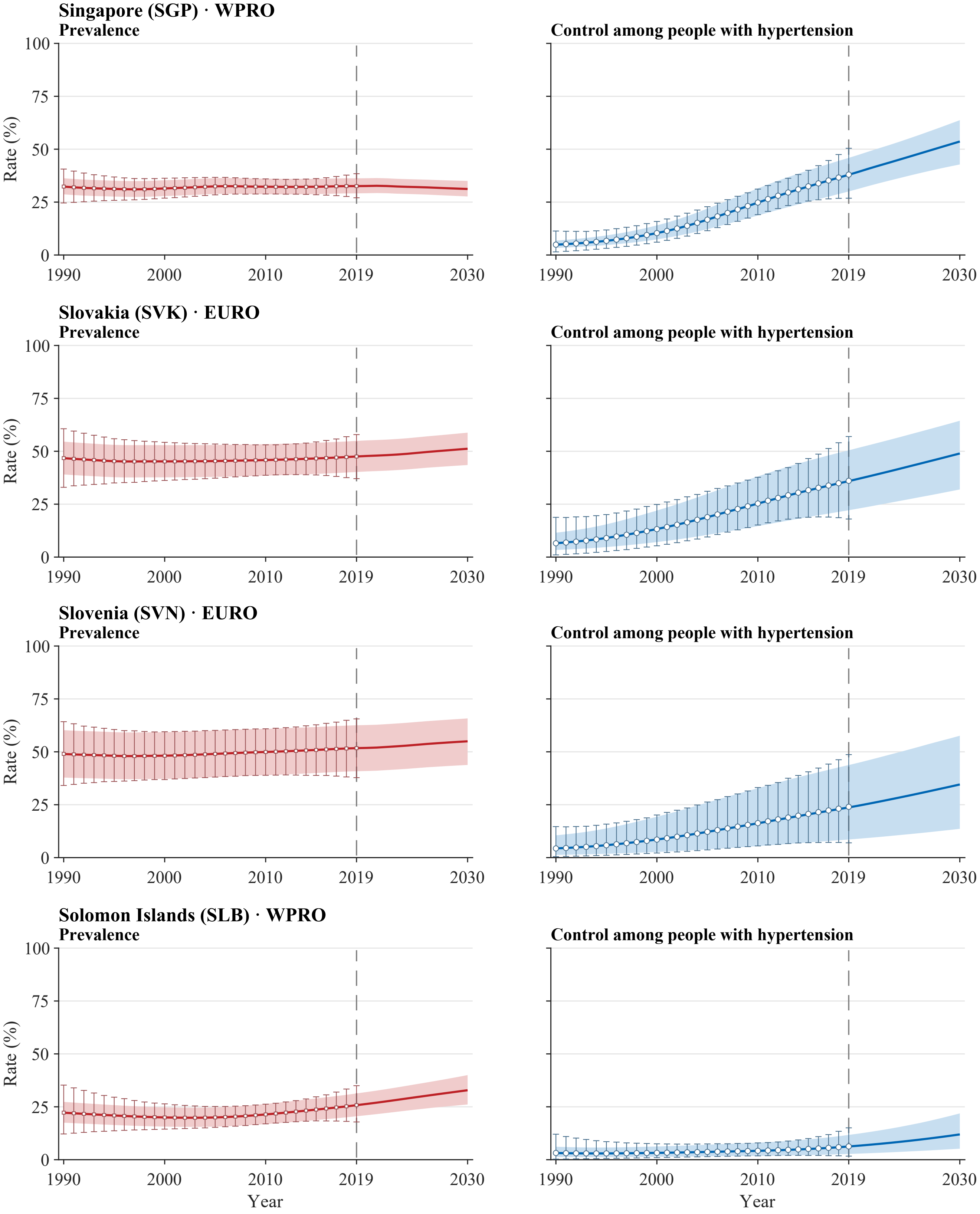


**Legend.** Open squares and error bars in prevalence panels and open circles and error bars in control panels show annual NCD-RisC central estimates with propagated 95% uncertainty intervals for 1990–2019. Solid red and blue lines and shaded bands show model medians and 95% uncertainty intervals for 1990–2030. The vertical dashed line marks 2019, the final year with reported estimates; values for 2020–2030 are forecast. Latent stratum-specific values in 2019 are anchored to the published central estimates. Propagated intervals for the published estimates are wider than the model bands during the historical period by construction of the external uncertainty layer. Model uncertainty represents a persistent level offset. All panels use a common 0–100% scale. Prevalence is defined among all adults aged 30–79 years, whereas control is defined among adults with hypertension. Estimates use CORE-P25 and the prespecified main dependence assumptions.

**Supplementary Figure S3. Country-level trajectories of aggregate hypertension prevalence and control, 1990–2030**
Atlas panel page 42 of 50; countries 165–168 of 200.

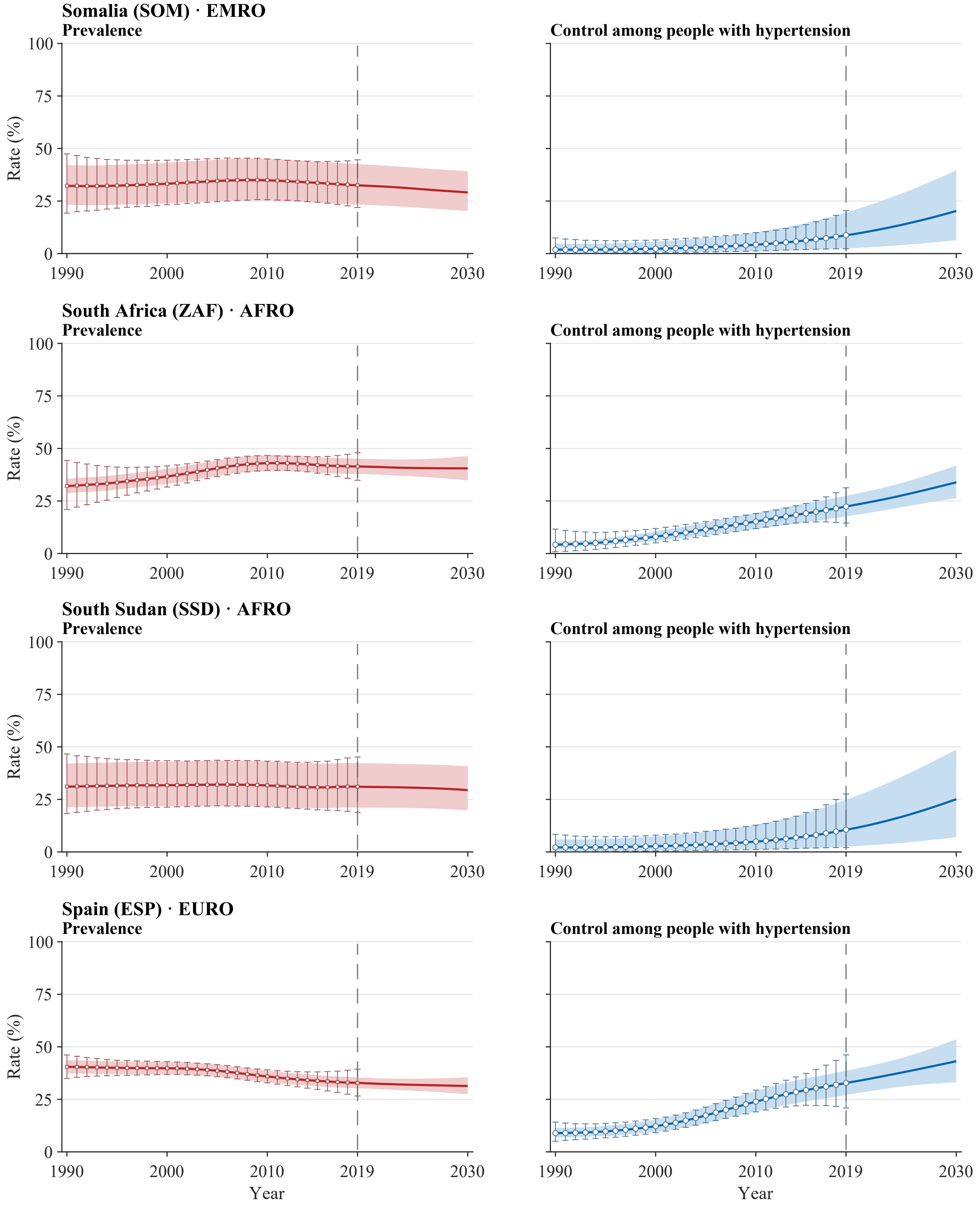


**Legend.** Open squares and error bars in prevalence panels and open circles and error bars in control panels show annual NCD-RisC central estimates with propagated 95% uncertainty intervals for 1990–2019. Solid red and blue lines and shaded bands show model medians and 95% uncertainty intervals for 1990–2030. The vertical dashed line marks 2019, the final year with reported estimates; values for 2020–2030 are forecast. Latent stratum-specific values in 2019 are anchored to the published central estimates. Propagated intervals for the published estimates are wider than the model bands during the historical period by construction of the external uncertainty layer. Model uncertainty represents a persistent level offset. All panels use a common 0–100% scale. Prevalence is defined among all adults aged 30–79 years, whereas control is defined among adults with hypertension. Estimates use CORE-P25 and the prespecified main dependence assumptions.

**Supplementary Figure S3. Country-level trajectories of aggregate hypertension prevalence and control, 1990–2030**
Atlas panel page 43 of 50; countries 169–172 of 200.

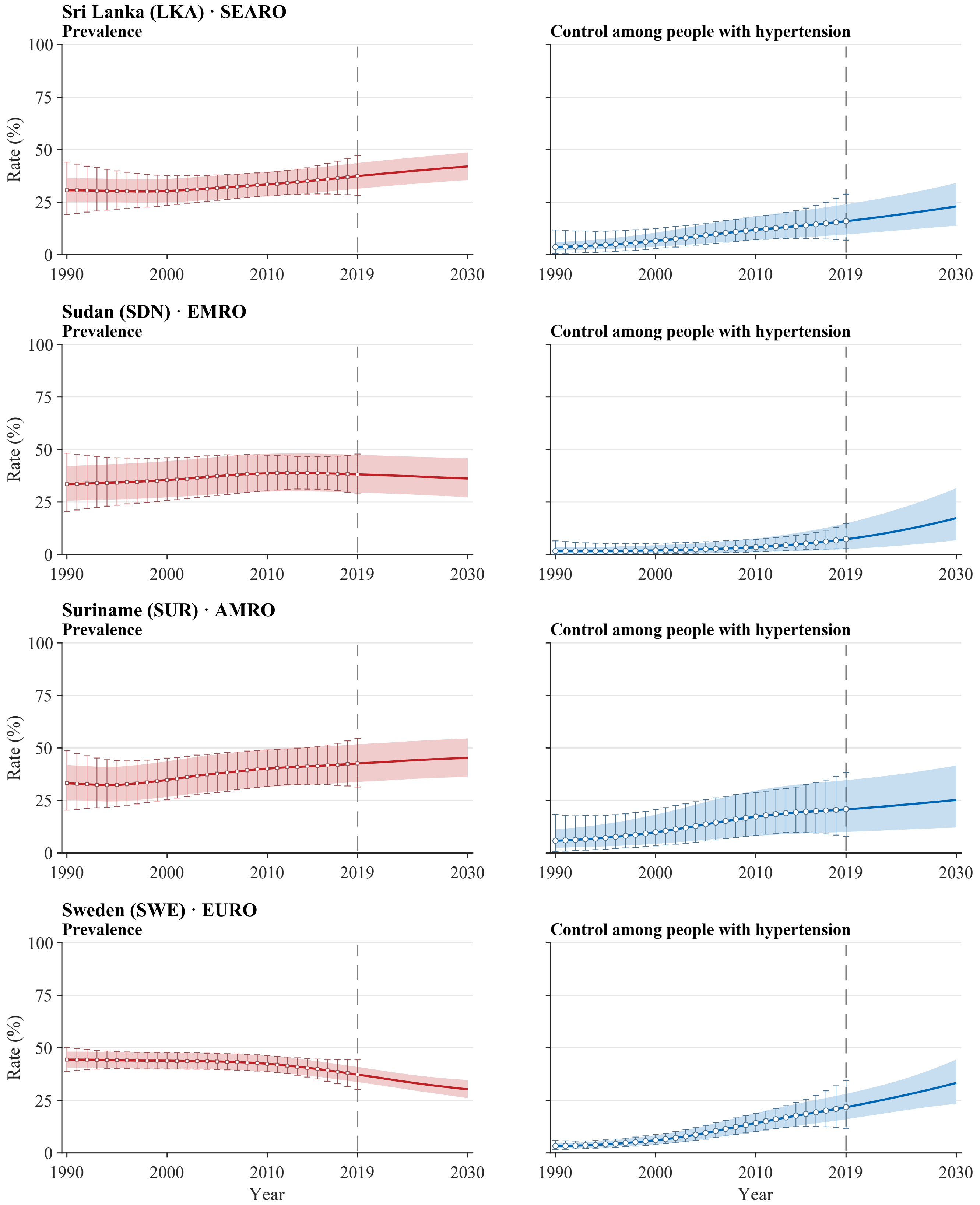


**Legend.** Open squares and error bars in prevalence panels and open circles and error bars in control panels show annual NCD-RisC central estimates with propagated 95% uncertainty intervals for 1990–2019. Solid red and blue lines and shaded bands show model medians and 95% uncertainty intervals for 1990–2030. The vertical dashed line marks 2019, the final year with reported estimates; values for 2020–2030 are forecast. Latent stratum-specific values in 2019 are anchored to the published central estimates. Propagated intervals for the published estimates are wider than the model bands during the historical period by construction of the external uncertainty layer. Model uncertainty represents a persistent level offset. All panels use a common 0–100% scale. Prevalence is defined among all adults aged 30–79 years, whereas control is defined among adults with hypertension. Estimates use CORE-P25 and the prespecified main dependence assumptions.

**Supplementary Figure S3. Country-level trajectories of aggregate hypertension prevalence and control, 1990–2030**
Atlas panel page 44 of 50; countries 173–176 of 200.

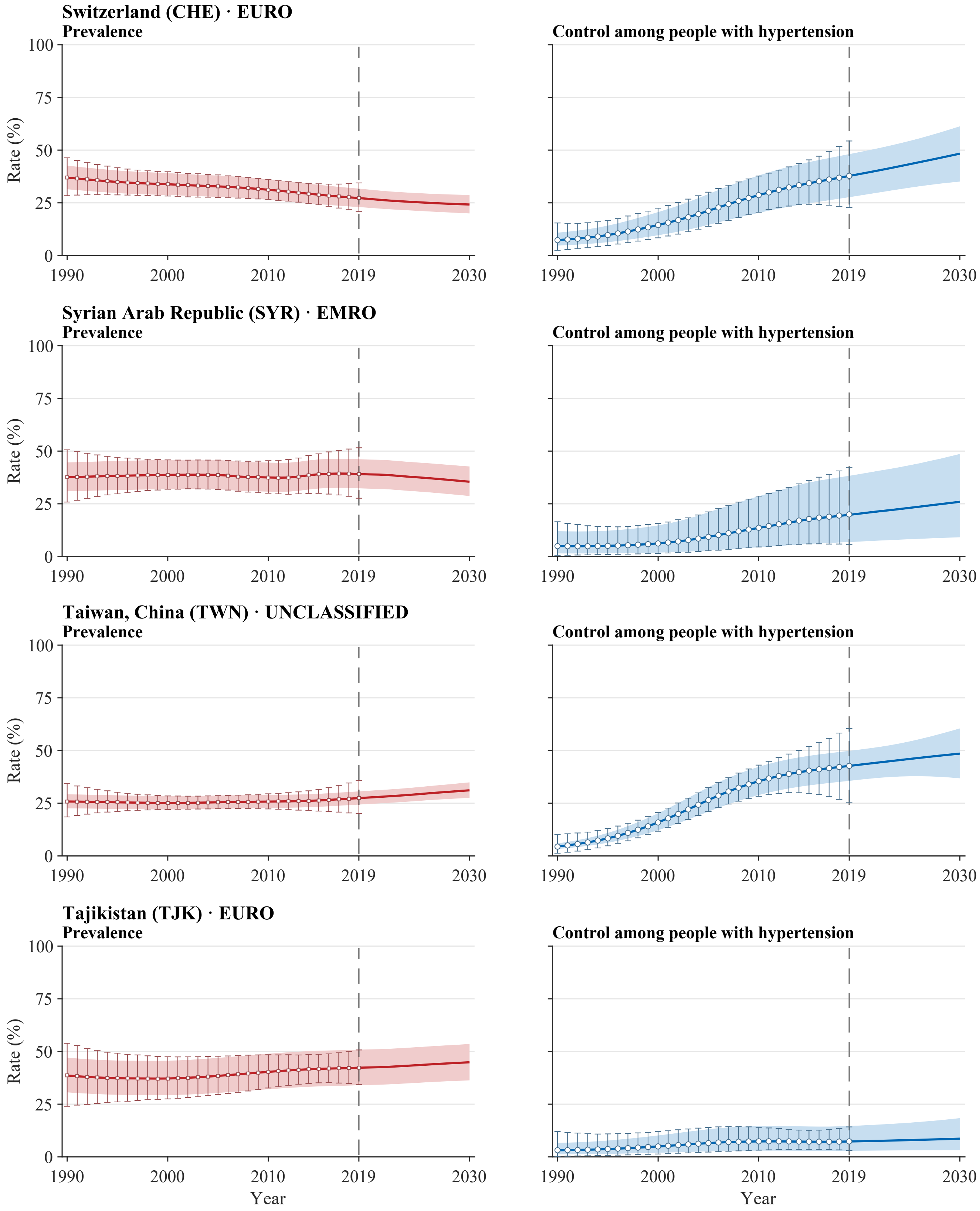


**Legend.** Open squares and error bars in prevalence panels and open circles and error bars in control panels show annual NCD-RisC central estimates with propagated 95% uncertainty intervals for 1990–2019. Solid red and blue lines and shaded bands show model medians and 95% uncertainty intervals for 1990–2030. The vertical dashed line marks 2019, the final year with reported estimates; values for 2020–2030 are forecast. Latent stratum-specific values in 2019 are anchored to the published central estimates. Propagated intervals for the published estimates are wider than the model bands during the historical period by construction of the external uncertainty layer. Model uncertainty represents a persistent level offset. All panels use a common 0–100% scale. Prevalence is defined among all adults aged 30–79 years, whereas control is defined among adults with hypertension. Estimates use CORE-P25 and the prespecified main dependence assumptions.

**Supplementary Figure S3. Country-level trajectories of aggregate hypertension prevalence and control, 1990–2030**
Atlas panel page 45 of 50; countries 177–180 of 200.

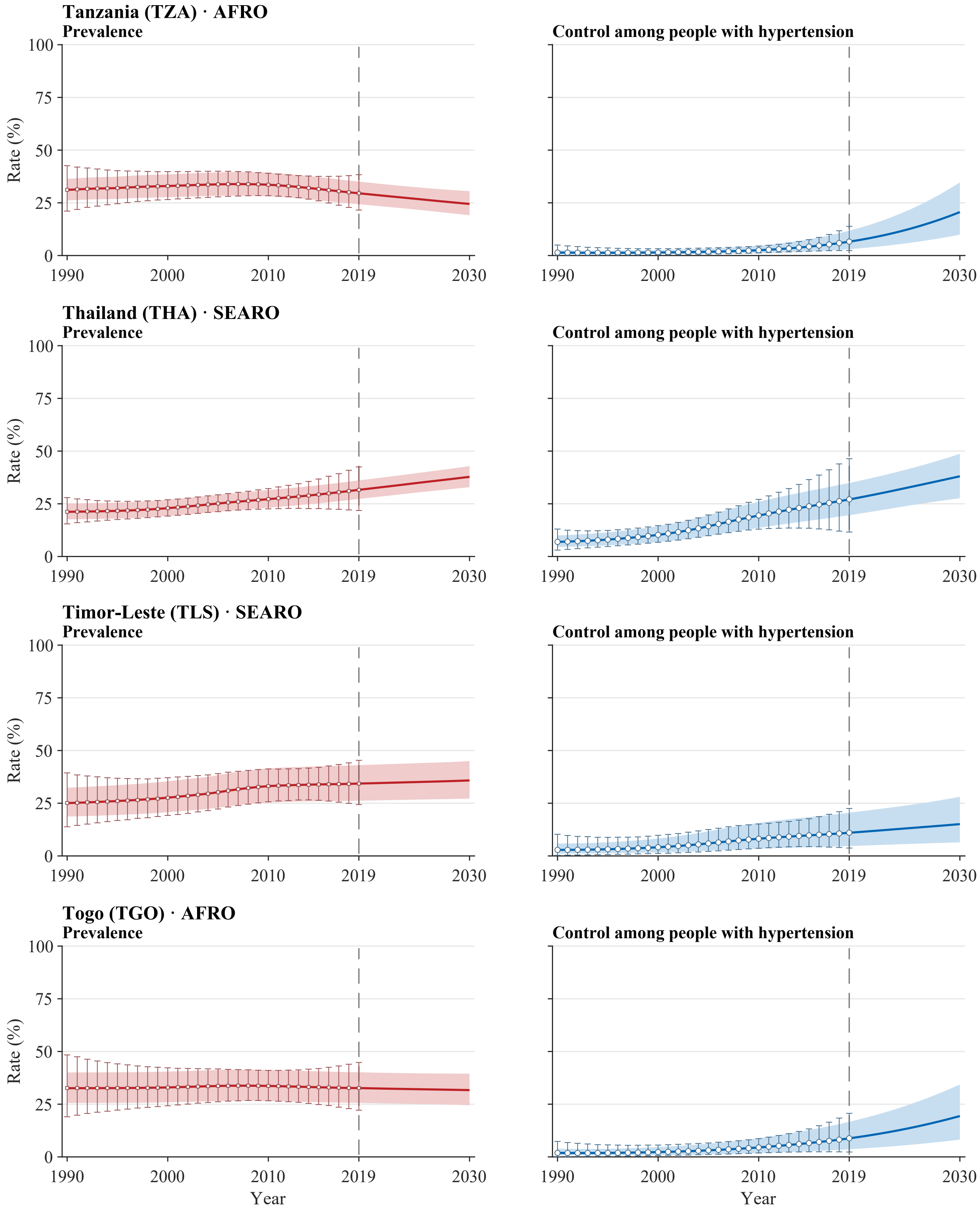


**Legend.** Open squares and error bars in prevalence panels and open circles and error bars in control panels show annual NCD-RisC central estimates with propagated 95% uncertainty intervals for 1990–2019. Solid red and blue lines and shaded bands show model medians and 95% uncertainty intervals for 1990–2030. The vertical dashed line marks 2019, the final year with reported estimates; values for 2020–2030 are forecast. Latent stratum-specific values in 2019 are anchored to the published central estimates. Propagated intervals for the published estimates are wider than the model bands during the historical period by construction of the external uncertainty layer. Model uncertainty represents a persistent level offset. All panels use a common 0–100% scale. Prevalence is defined among all adults aged 30–79 years, whereas control is defined among adults with hypertension. Estimates use CORE-P25 and the prespecified main dependence assumptions.

**Supplementary Figure S3. Country-level trajectories of aggregate hypertension prevalence and control, 1990–2030**

Atlas panel page 46 of 50; countries 181–184 of 200.

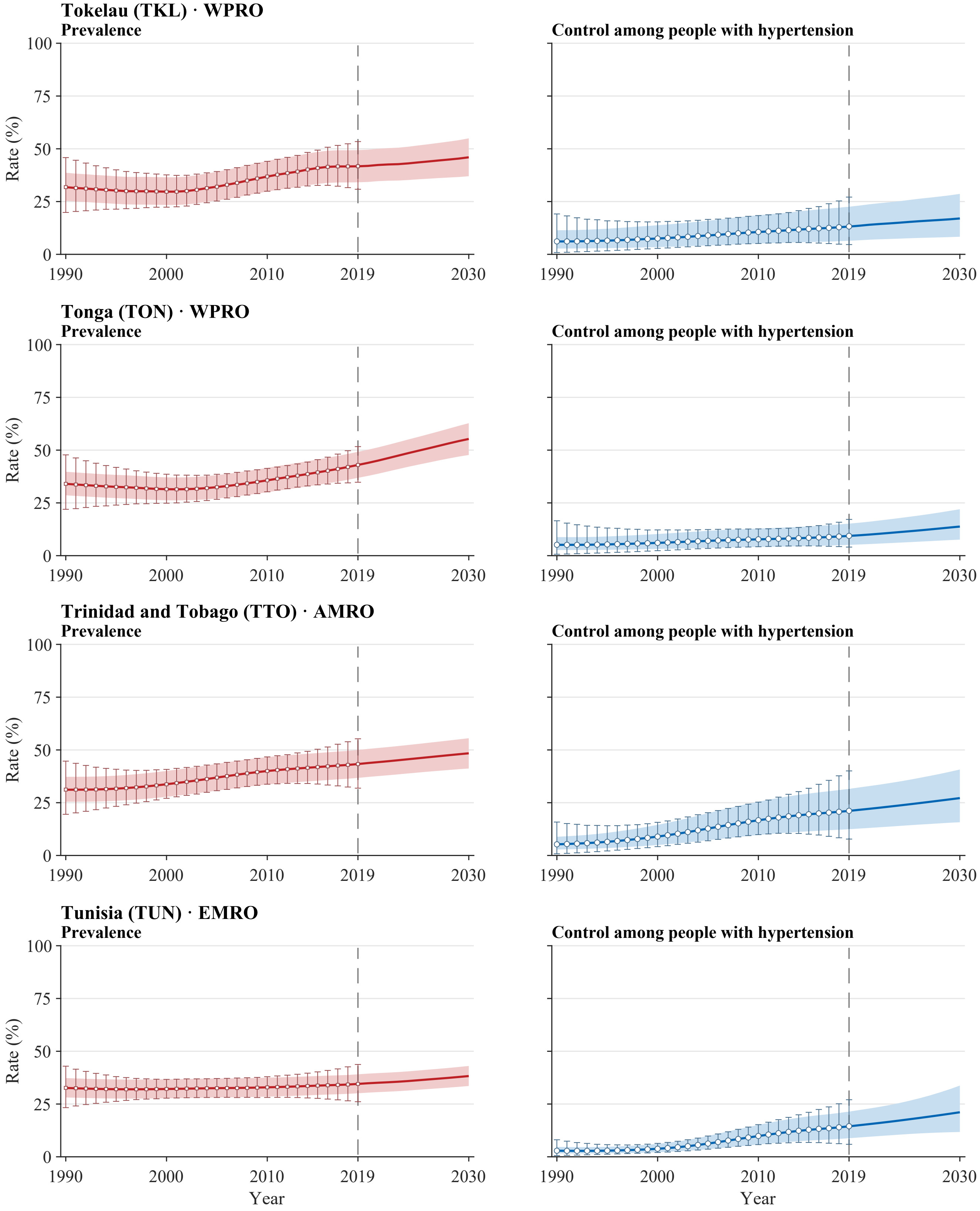


**Legend.** Open squares and error bars in prevalence panels and open circles and error bars in control panels show annual NCD-RisC central estimates with propagated 95% uncertainty intervals for 1990–2019. Solid red and blue lines and shaded bands show model medians and 95% uncertainty intervals for 1990–2030. The vertical dashed line marks 2019, the final year with reported estimates; values for 2020–2030 are forecast. Latent stratum-specific values in 2019 are anchored to the published central estimates. Propagated intervals for the published estimates are wider than the model bands during the historical period by construction of the external uncertainty layer. Model uncertainty represents a persistent level offset. All panels use a common 0–100% scale. Prevalence is defined among all adults aged 30–79 years, whereas control is defined among adults with hypertension. Estimates use CORE-P25 and the prespecified main dependence assumptions.

**Supplementary Figure S3. Country-level trajectories of aggregate hypertension prevalence and control, 1990–2030**

Atlas panel page 47 of 50; countries 185–188 of 200.

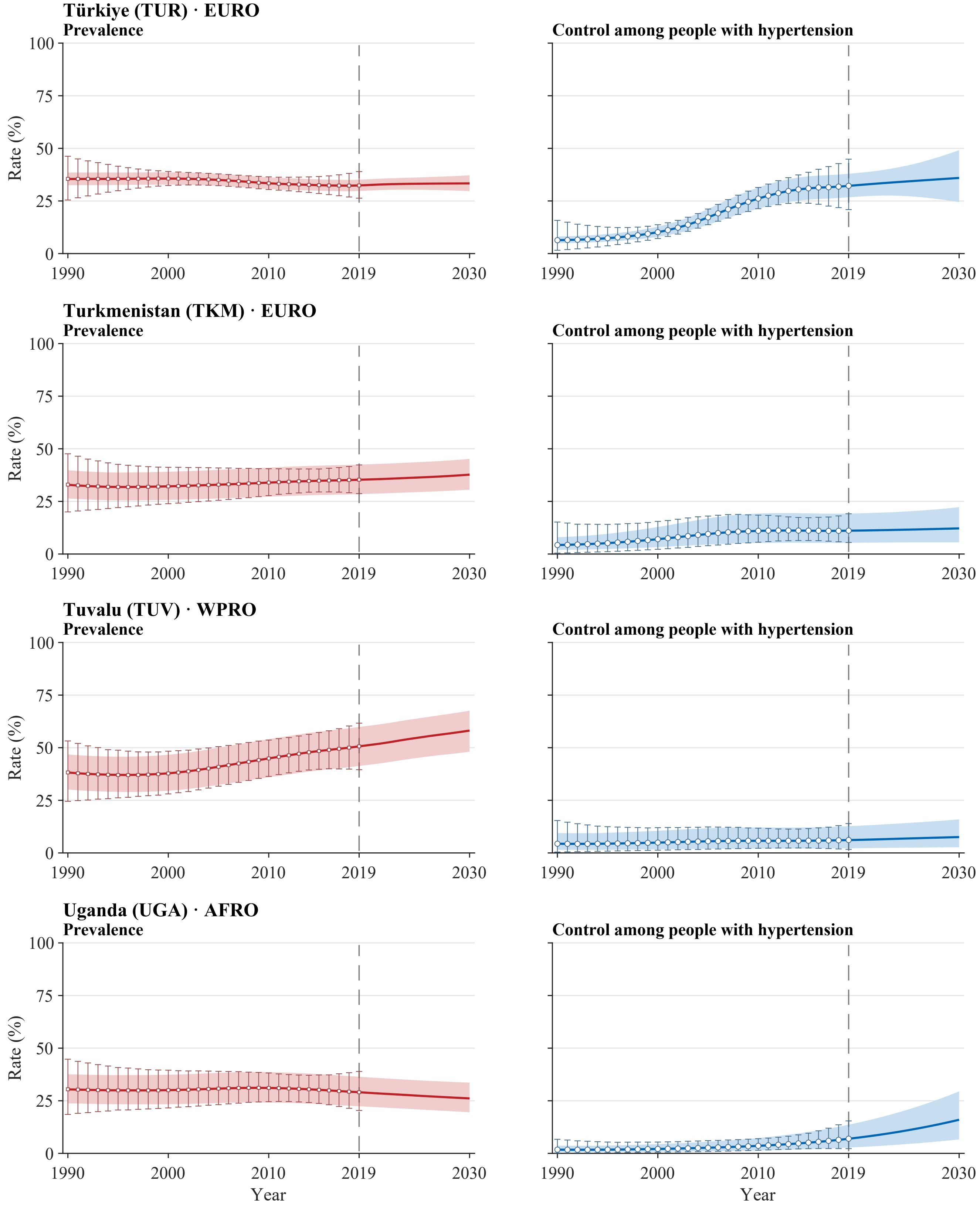


**Legend.** Open squares and error bars in prevalence panels and open circles and error bars in control panels show annual NCD-RisC central estimates with propagated 95% uncertainty intervals for 1990–2019. Solid red and blue lines and shaded bands show model medians and 95% uncertainty intervals for 1990–2030. The vertical dashed line marks 2019, the final year with reported estimates; values for 2020–2030 are forecast. Latent stratum-specific values in 2019 are anchored to the published central estimates. Propagated intervals for the published estimates are wider than the model bands during the historical period by construction of the external uncertainty layer. Model uncertainty represents a persistent level offset. All panels use a common 0–100% scale. Prevalence is defined among all adults aged 30–79 years, whereas control is defined among adults with hypertension. Estimates use CORE-P25 and the prespecified main dependence assumptions.

**Supplementary Figure S3. Country-level trajectories of aggregate hypertension prevalence and control, 1990–2030**

Atlas panel page 48 of 50; countries 189–192 of 200.

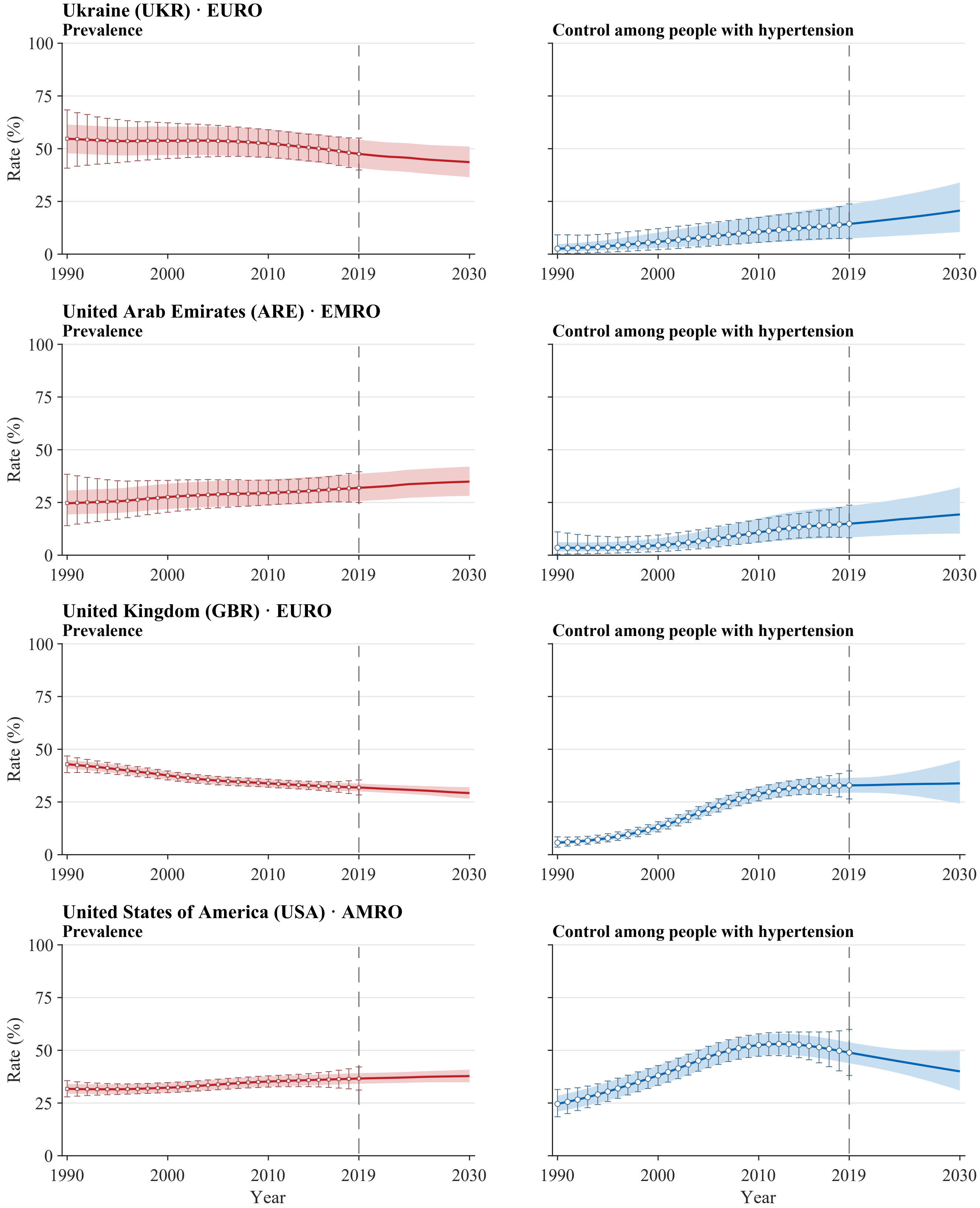


**Legend.** Open squares and error bars in prevalence panels and open circles and error bars in control panels show annual NCD-RisC central estimates with propagated 95% uncertainty intervals for 1990–2019. Solid red and blue lines and shaded bands show model medians and 95% uncertainty intervals for 1990–2030. The vertical dashed line marks 2019, the final year with reported estimates; values for 2020–2030 are forecast. Latent stratum-specific values in 2019 are anchored to the published central estimates. Propagated intervals for the published estimates are wider than the model bands during the historical period by construction of the external uncertainty layer. Model uncertainty represents a persistent level offset. All panels use a common 0–100% scale. Prevalence is defined among all adults aged 30–79 years, whereas control is defined among adults with hypertension. Estimates use CORE-P25 and the prespecified main dependence assumptions.

**Supplementary Figure S3. Country-level trajectories of aggregate hypertension prevalence and control, 1990–2030**
Atlas panel page 49 of 50; countries 193–196 of 200.

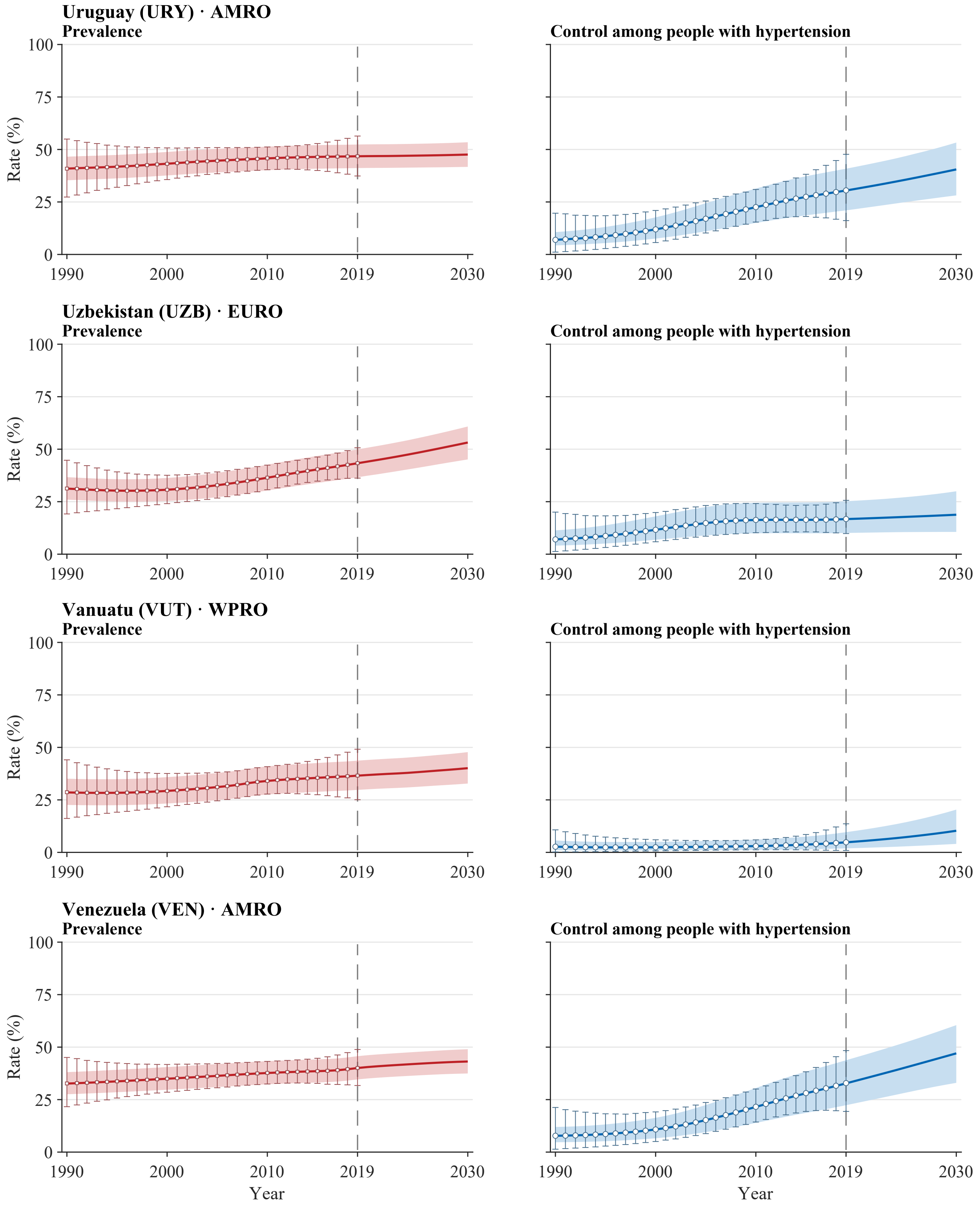


**Legend.** Open squares and error bars in prevalence panels and open circles and error bars in control panels show annual NCD-RisC central estimates with propagated 95% uncertainty intervals for 1990–2019. Solid red and blue lines and shaded bands show model medians and 95% uncertainty intervals for 1990–2030. The vertical dashed line marks 2019, the final year with reported estimates; values for 2020–2030 are forecast. Latent stratum-specific values in 2019 are anchored to the published central estimates. Propagated intervals for the published estimates are wider than the model bands during the historical period by construction of the external uncertainty layer. Model uncertainty represents a persistent level offset. All panels use a common 0–100% scale. Prevalence is defined among all adults aged 30–79 years, whereas control is defined among adults with hypertension. Estimates use CORE-P25 and the prespecified main dependence assumptions.

**Supplementary Figure S3. Country-level trajectories of aggregate hypertension prevalence and control, 1990–2030**

Atlas panel page 50 of 50; countries 197–200 of 200.

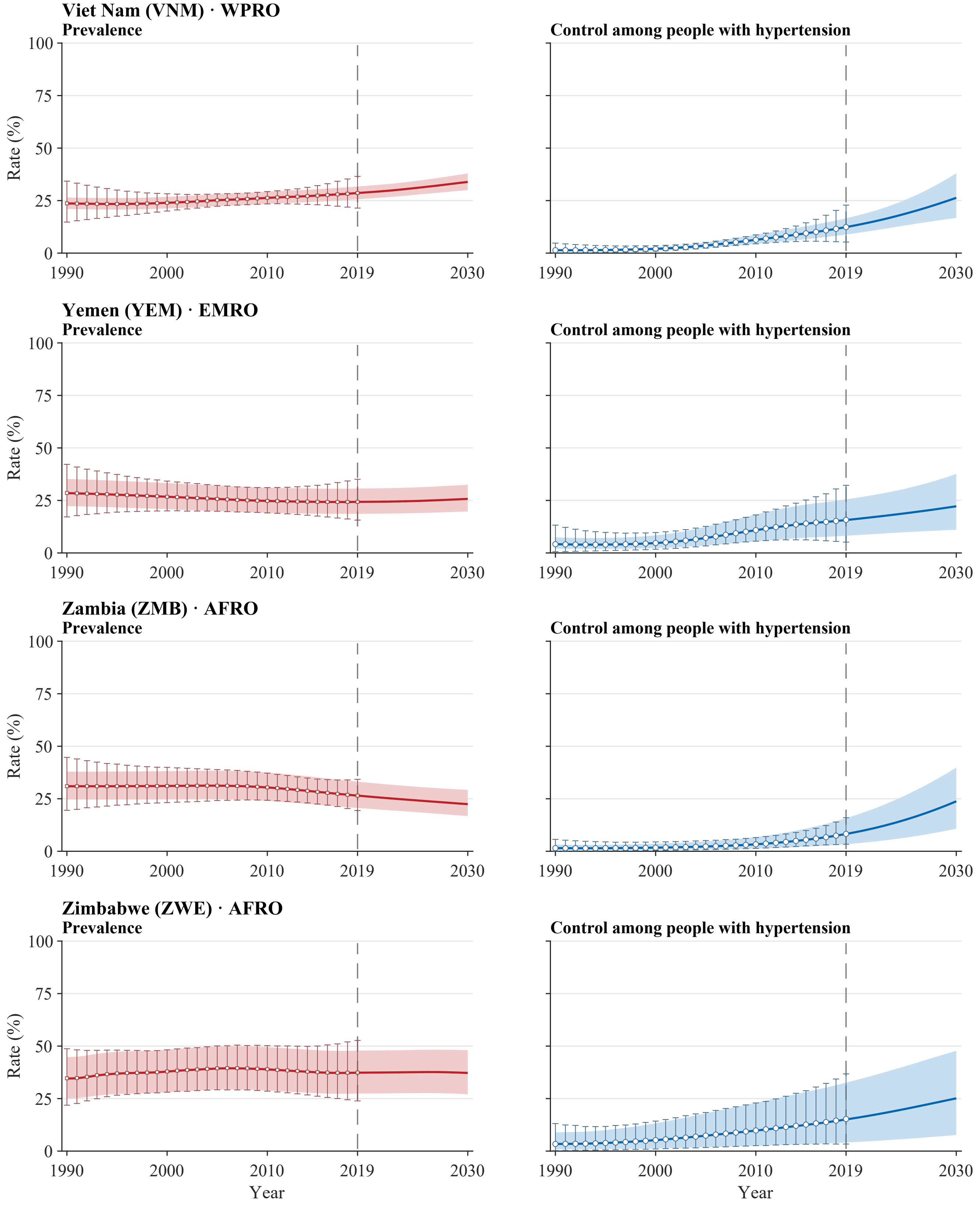


**Legend.** Open squares and error bars in prevalence panels and open circles and error bars in control panels show annual NCD-RisC central estimates with propagated 95% uncertainty intervals for 1990–2019. Solid red and blue lines and shaded bands show model medians and 95% uncertainty intervals for 1990–2030. The vertical dashed line marks 2019, the final year with reported estimates; values for 2020–2030 are forecast. Latent stratum-specific values in 2019 are anchored to the published central estimates. Propagated intervals for the published estimates are wider than the model bands during the historical period by construction of the external uncertainty layer. Model uncertainty represents a persistent level offset. All panels use a common 0–100% scale. Prevalence is defined among all adults aged 30–79 years, whereas control is defined among adults with hypertension. Estimates use CORE-P25 and the prespecified main dependence assumptions.
