## Supplementary Table S4 for "Forecasting global progress towards the United Nations hypertension control target for 2030"

#### Supplementary Table S4. Country-level forecast contributions and Horiuchi decomposition, 2024–2030

##### Section A. Forecast levels, change, and contribution to the +150 million target



| ISO3 | Country | WHO region | Unit | C2024 (95% UI) | C2030 (95% UI) | ΔC (95% UI) | Contribution to +150 M % (95% UI) | Pr(ΔC ≤ 0) % | Status |
|---|---|---|---|---|---|---|---|---|---|
| AFG | Afghanistan | EMRO | thousands | 725.4 (294.3–1395) | 1159 (474.0–2198) | 430.4 (167.5–838.9) | 0.29 (0.11–0.56) | <0.01 | Yes |
| ALB | Albania | EURO | thousands | 85.0 (37.5–155.5) | 110.3 (48.2–202.2) | 24.6 (6.93–54.6) | 0.02 (0.00–0.04) | 0.14 | Yes |
| DZA | Algeria | AFRO | thousands | 1318 (730.7–2093) | 1839 (945.7–3116) | 513.8 (86.3–1183) | 0.34 (0.06–0.79) | 0.75 | Yes |
| ASM | American Samoa | WPRO | persons | 1562 (764.4–2676) | 2002 (984.6–3392) | 431.6 (170.4–821.4) | 0.00 (0.00–0.00) | 0.03 | Yes |
| AND | Andorra | EURO | thousands | 7.01 (3.13–12.1) | 8.66 (3.94–14.6) | 1.61 (0.426–3.11) | 0.00 (0.00–0.00) | 0.27 | Yes |
| AGO | Angola | AFRO | thousands | 589.3 (187.7–1268) | 1007 (332.9–2074) | 411.9 (135.2–839.7) | 0.27 (0.09–0.56) | 0.00 | Yes |
| ATG | Antigua and Barbuda | AMRO | thousands | 6.31 (2.34–12.1) | 7.83 (2.92–14.7) | 1.46 (0.348–3.13) | 0.00 (0.00–0.00) | 0.26 | Yes |
| ARG | Argentina | AMRO | thousands | 1837 (1031–2923) | 2025 (1077–3381) | 185.6 (−152.1–672.9) | 0.12 (−0.10–0.45) | 14.48 | No |
| ARM | Armenia | EURO | thousands | 92.4 (43.4–164.9) | 114.8 (50.9–214.0) | 21.7 (0.973–59.3) | 0.01 (0.00–0.04) | 1.95 | Yes |
| AUS | Australia | WPRO | thousands | 1293 (990.0–1654) | 1171 (767.0–1717) | −120.1 (−361.5–188.3) | −0.08 (−0.24–0.13) | 80.24 | No |
| AUT | Austria | EURO | thousands | 695.5 (348.0–1123) | 772.3 (382.8–1247) | 73.9 (−42.6–215.0) | 0.05 (−0.03–0.14) | 10.54 | No |
| AZE | Azerbaijan | EURO | thousands | 352.3 (187.6–583.2) | 447.1 (224.3–774.8) | 92.7 (13.8–224.2) | 0.06 (0.01–0.15) | 0.87 | Yes |
| BHS | Bahamas | AMRO | thousands | 26.0 (13.9–41.7) | 31.7 (16.7–51.0) | 5.51 (1.17–11.5) | 0.00 (0.00–0.01) | 0.55 | Yes |
| BHR | Bahrain | EMRO | thousands | 56.3 (19.0–114.9) | 75.3 (25.4–153.6) | 18.3 (2.95–45.5) | 0.01 (0.00–0.03) | 0.60 | Yes |
| BGD | Bangladesh | SEARO | thousands | 3315 (2171–4798) | 4376 (2717–6665) | 1050 (311.9–2148) | 0.70 (0.21–1.43) | 0.21 | Yes |
| BRB | Barbados | AMRO | thousands | 31.4 (19.7–44.6) | 35.8 (22.4–50.7) | 4.35 (0.476–8.67) | 0.00 (0.00–0.01) | 1.42 | Yes |
| BLR | Belarus | EURO | thousands | 393.2 (155.5–762.9) | 459.5 (178.8–904.9) | 63.0 (−8.05–192.0) | 0.04 (−0.01–0.13) | 4.26 | No |
| BEL | Belgium | EURO | thousands | 1031 (781.6–1305) | 1027 (737.8–1355) | −3.48 (−134.2–139.5) | 0.00 (−0.09–0.09) | 52.10 | No |
| BLZ | Belize | AMRO | thousands | 14.9 (7.84–24.6) | 21.2 (11.0–35.3) | 6.22 (2.47–11.8) | 0.00 (0.00–0.01) | <0.01 | Yes |
| BEN | Benin | AFRO | thousands | 133.0 (74.4–214.9) | 205.2 (110.8–341.9) | 71.3 (29.3–138.3) | 0.05 (0.02–0.09) | <0.01 | Yes |
| BMU | Bermuda | AMRO | persons | 5542 (2044–10462) | 6424 (2397–11851) | 841.3 (60.0–1963) | 0.00 (0.00–0.00) | 1.69 | Yes |
| BTN | Bhutan | SEARO | thousands | 15.6 (7.57–27.6) | 22.7 (11.0–40.4) | 7.01 (2.75–14.2) | 0.00 (0.00–0.01) | <0.01 | Yes |
| BOL | Bolivia | AMRO | thousands | 373.3 (130.3–769.9) | 494.3 (173.9–1003) | 118.5 (33.9–257.5) | 0.08 (0.02–0.17) | 0.03 | Yes |
| BIH | Bosnia and Herzegovina | EURO | thousands | 252.2 (138.3–392.2) | 325.5 (181.1–494.2) | 71.9 (31.1–122.5) | 0.05 (0.02–0.08) | 0.03 | Yes |
| BWA | Botswana | AFRO | thousands | 79.0 (46.7–120.3) | 124.2 (74.0–187.3) | 44.8 (24.2–72.0) | 0.03 (0.02–0.05) | 0.00 | Yes |

Countries 1–25 of 200; alphabetical order. Values: posterior median (95% UI). Status: fractions reportable (Yes/No).

#### Supplementary Table S4. Country-level forecast contributions and Horiuchi decomposition, 2024–2030

##### Section A. Forecast levels, change, and contribution to the +150 million target



| ISO3 | Country | WHO region | Unit | C2024 (95% UI) | C2030 (95% UI) | ΔC (95% UI) | Contribution to +150 M % (95% UI) | Pr(ΔC ≤ 0) % | Status |
|---|---|---|---|---|---|---|---|---|---|
| BRA | Brazil | AMRO | thousands | 22132 (17795–26797) | 29481 (23173–36184) | 7339 (4226–10683) | 4.89 (2.82–7.12) | 0.00 | Yes |
| BRN | Brunei Darussalam | WPRO | thousands | 38.7 (24.9–55.0) | 55.5 (35.7–77.8) | 16.6 (8.85–25.6) | 0.01 (0.01–0.02) | 0.00 | Yes |
| BGR | Bulgaria | EURO | thousands | 668.6 (242.9–1236) | 782.6 (299.5–1368) | 105.9 (32.1–199.3) | 0.07 (0.02–0.13) | 0.24 | Yes |
| BFA | Burkina Faso | AFRO | thousands | 202.4 (76.4–401.2) | 359.0 (139.5–694.3) | 154.6 (58.3–309.6) | 0.10 (0.04–0.21) | 0.00 | Yes |
| BDI | Burundi | AFRO | thousands | 165.1 (40.1–393.8) | 296.8 (79.0–649.7) | 129.2 (37.0–270.0) | 0.09 (0.02–0.18) | 0.00 | Yes |
| CPV | Cabo Verde | AFRO | thousands | 16.9 (9.02–26.9) | 25.4 (13.8–39.8) | 8.41 (4.14–14.1) | 0.01 (0.00–0.01) | <0.01 | Yes |
| KHM | Cambodia | WPRO | thousands | 485.7 (259.7–795.2) | 707.4 (373.7–1157) | 218.7 (91.8–399.5) | 0.15 (0.06–0.27) | <0.01 | Yes |
| CMR | Cameroon | AFRO | thousands | 264.7 (125.3–470.3) | 478.4 (226.9–849.5) | 211.9 (93.0–399.9) | 0.14 (0.06–0.27) | 0.00 | Yes |
| CAN | Canada | AMRO | thousands | 4096 (3390–4859) | 4159 (3237–5117) | 68.0 (−403.0–505.5) | 0.05 (−0.27–0.34) | 38.20 | No |
| CAF | Central African Republic | AFRO | thousands | 32.5 (11.5–68.8) | 57.7 (19.9–122.3) | 24.9 (7.72–56.6) | 0.02 (0.01–0.04) | 0.00 | Yes |
| TCD | Chad | AFRO | thousands | 250.8 (83.0–527.6) | 438.3 (149.6–887.7) | 184.7 (61.8–378.0) | 0.12 (0.04–0.25) | 0.00 | Yes |
| CHL | Chile | AMRO | thousands | 1673 (1245–2153) | 2039 (1469–2674) | 364.8 (123.9–639.9) | 0.24 (0.08–0.43) | 0.16 | Yes |
| CHN | China | WPRO | thousands | 48695 (39030–59731) | 58097 (41657–79275) | 9376 (−700.3–22787) | 6.25 (−0.47–15.19) | 3.51 | No |
| COL | Colombia | AMRO | thousands | 3632 (2562–4829) | 5196 (3656–6869) | 1557 (897.7–2285) | 1.04 (0.60–1.52) | <0.01 | Yes |
| COM | Comoros | AFRO | thousands | 14.6 (6.37–26.7) | 25.1 (11.4–43.6) | 10.3 (4.61–18.2) | 0.01 (0.00–0.01) | 0.00 | Yes |
| COG | Congo | AFRO | thousands | 95.7 (26.9–217.2) | 172.0 (50.1–369.6) | 75.0 (21.7–160.7) | 0.05 (0.01–0.11) | 0.00 | Yes |
| COK | Cook Islands | WPRO | persons | 733.4 (421.4–1139) | 798.6 (455.9–1238) | 63.0 (−18.0–163.5) | 0.00 (0.00–0.00) | 6.17 | No |
| CRI | Costa Rica | AMRO | thousands | 672.7 (570.8–780.2) | 870.6 (731.1–1017) | 197.5 (130.1–267.7) | 0.13 (0.09–0.18) | 0.00 | Yes |
| CIV | Côte d'Ivoire | AFRO | thousands | 374.8 (125.8–791.3) | 701.1 (244.5–1420) | 321.0 (109.8–662.4) | 0.21 (0.07–0.44) | 0.00 | Yes |
| HRV | Croatia | EURO | thousands | 360.0 (257.4–474.4) | 429.2 (302.5–572.9) | 68.8 (22.1–124.8) | 0.05 (0.01–0.08) | 0.23 | Yes |
| CUB | Cuba | AMRO | thousands | 1137 (768.3–1568) | 1269 (834.9–1775) | 130.1 (−23.6–304.1) | 0.09 (−0.02–0.20) | 4.77 | No |
| CYP | Cyprus | EURO | thousands | 98.5 (42.9–171.1) | 117.9 (51.9–201.4) | 18.9 (3.41–39.3) | 0.01 (0.00–0.03) | 0.77 | Yes |
| CZE | Czechia | EURO | thousands | 1431 (1142–1731) | 1621 (1278–1976) | 188.8 (52.9–332.7) | 0.13 (0.04–0.22) | 0.38 | Yes |
| PRK | Democratic People's Republic of Korea | SEARO | thousands | 1164 (397.0–2296) | 1447 (505.1–2769) | 269.6 (60.8–584.1) | 0.18 (0.04–0.39) | 0.31 | Yes |
| COD | Democratic Republic of the Congo | AFRO | thousands | 1406 (496.1–2882) | 2480 (906.8–4854) | 1061 (387.4–2068) | 0.71 (0.26–1.38) | 0.00 | Yes |

Countries 26–50 of 200; alphabetical order. Values: posterior median (95% UI). Status: fractions reportable (Yes/No).

**Supplementary Table S4. Country-level forecast contributions and Horiuchi decomposition, 2024–2030**

**Section A. Forecast levels, change, and contribution to the +150 million target**



| ISO3 | Country | WHO region | Unit | C2024 (95% UI) | C2030 (95% UI) | ΔC (95% UI) | Contribution to +150 M % (95% UI) | Pr(ΔC ≤ 0) % | Status |
|---|---|---|---|---|---|---|---|---|---|
| DNK | Denmark | EURO | thousands | 181.3 (135.2–236.3) | 192.0 (126.1–281.0) | 10.5 (−27.4–63.1) | 0.01 (−0.02–0.04) | 30.88 | No |
| DJI | Djibouti | EMRO | thousands | 23.7 (5.68–55.0) | 40.5 (10.7–86.6) | 16.6 (4.78–33.6) | 0.01 (0.00–0.02) | 0.00 | Yes |
| DMA | Dominica | AMRO | persons | 4004 (1994–6663) | 4828 (2383–8032) | 805.1 (134.0–1769) | 0.00 (0.00–0.00) | 0.83 | Yes |
| DOM | Dominican Republic | AMRO | thousands | 609.8 (375.4–895.5) | 773.4 (467.3–1153) | 161.7 (53.6–307.2) | 0.11 (0.04–0.20) | 0.14 | Yes |
| ECU | Ecuador | AMRO | thousands | 728.4 (495.1–1013) | 955.3 (630.9–1356) | 225.6 (91.9–394.4) | 0.15 (0.06–0.26) | 0.03 | Yes |
| EGY | Egypt | EMRO | thousands | 3560 (2552–4763) | 4618 (2912–6891) | 1054 (43.5–2471) | 0.70 (0.03–1.65) | 2.01 | Yes |
| SLV | El Salvador | AMRO | thousands | 407.6 (269.7–569.8) | 517.3 (345.3–718.5) | 109.2 (58.1–168.8) | 0.07 (0.04–0.11) | <0.01 | Yes |
| GNQ | Equatorial Guinea | AFRO | thousands | 34.6 (7.74–86.1) | 57.9 (13.8–134.1) | 22.8 (5.72–50.2) | 0.02 (0.00–0.03) | 0.00 | Yes |
| ERI | Eritrea | AFRO | thousands | 44.1 (22.3–74.1) | 79.1 (41.2–128.1) | 34.5 (17.4–57.4) | 0.02 (0.01–0.04) | 0.00 | Yes |
| EST | Estonia | EURO | thousands | 71.3 (46.9–100.9) | 82.9 (53.0–121.0) | 11.6 (0.324–26.4) | 0.01 (0.00–0.02) | 2.14 | Yes |
| SWZ | Eswatini | AFRO | thousands | 26.4 (13.7–43.9) | 39.4 (20.4–65.2) | 12.8 (5.62–23.4) | 0.01 (0.00–0.02) | <0.01 | Yes |
| ETH | Ethiopia | AFRO | thousands | 869.8 (353.9–1691) | 1769 (730.2–3373) | 889.2 (354.6–1753) | 0.59 (0.24–1.17) | 0.00 | Yes |
| FJI | Fiji | WPRO | thousands | 24.9 (13.4–40.2) | 33.7 (18.2–54.5) | 8.73 (3.79–16.0) | 0.01 (0.00–0.01) | <0.01 | Yes |
| FIN | Finland | EURO | thousands | 477.8 (387.7–579.1) | 472.9 (343.2–625.8) | −4.75 (−81.3–83.0) | 0.00 (−0.05–0.06) | 54.57 | No |
| FRA | France | EURO | thousands | 4751 (3644–5974) | 5104 (3632–6779) | 352.2 (−396.6–1197) | 0.23 (−0.26–0.80) | 17.83 | No |
| PYF | French Polynesia | WPRO | thousands | 10.7 (5.16–18.8) | 15.1 (7.23–26.2) | 4.25 (1.82–7.95) | 0.00 (0.00–0.01) | <0.01 | Yes |
| GAB | Gabon | AFRO | thousands | 53.1 (20.8–101.5) | 88.4 (36.1–161.5) | 34.9 (14.3–63.7) | 0.02 (0.01–0.04) | 0.00 | Yes |
| GMB | Gambia | AFRO | thousands | 35.9 (14.8–67.4) | 65.7 (27.7–119.9) | 29.5 (12.0–55.2) | 0.02 (0.01–0.04) | 0.00 | Yes |
| GEO | Georgia | EURO | thousands | 221.8 (124.4–346.2) | 269.5 (147.6–426.0) | 46.7 (7.93–100.4) | 0.03 (0.01–0.07) | 0.87 | Yes |
| DEU | Germany | EURO | thousands | 9862 (8129–11667) | 10381 (8020–12803) | 524.1 (−727.9–1768) | 0.35 (−0.49–1.18) | 20.19 | No |
| GHA | Ghana | AFRO | thousands | 946.3 (627.0–1320) | 1572 (1044–2201) | 623.7 (356.7–958.7) | 0.42 (0.24–0.64) | 0.00 | Yes |
| GRC | Greece | EURO | thousands | 970.2 (697.5–1272) | 1022 (700.0–1388) | 52.5 (−86.8–205.1) | 0.03 (−0.06–0.14) | 22.86 | No |
| GRL | Greenland | EURO | persons | 3126 (1801–4815) | 3693 (2085–5724) | 554.8 (0.50–1258) | 0.00 (0.00–0.00) | 2.49 | Yes |
| GRD | Grenada | AMRO | thousands | 6.57 (3.39–10.7) | 8.15 (4.16–13.4) | 1.55 (0.406–3.20) | 0.00 (0.00–0.00) | 0.31 | Yes |
| GTM | Guatemala | AMRO | thousands | 481.9 (222.9–828.0) | 815.6 (386.3–1368) | 330.2 (145.0–576.2) | 0.22 (0.10–0.38) | 0.00 | Yes |

Countries 51–75 of 200; alphabetical order. Values: posterior median (95% UI). Status: fractions reportable (Yes/No).

#### Supplementary Table S4. Country-level forecast contributions and Horiuchi decomposition, 2024–2030

##### Section A. Forecast levels, change, and contribution to the +150 million target



| ISO3 | Country | WHO region | Unit | C2024 (95% UI) | C2030 (95% UI) | ΔC (95% UI) | Contribution to +150 M % (95% UI) | Pr(ΔC ≤ 0) % | Status |
|---|---|---|---|---|---|---|---|---|---|
| GIN | Guinea | AFRO | thousands | 158.6 (48.3–345.0) | 298.4 (94.2–624.0) | 137.4 (42.8–293.8) | 0.09 (0.03–0.20) | 0.00 | Yes |
| GNB | Guinea Bissau | AFRO | thousands | 33.3 (7.97–79.7) | 57.1 (14.5–127.9) | 23.4 (6.20–50.6) | 0.02 (0.00–0.03) | 0.00 | Yes |
| GUY | Guyana | AMRO | thousands | 30.8 (15.0–53.4) | 36.6 (17.4–64.3) | 5.65 (0.398–13.9) | 0.00 (0.00–0.01) | 1.64 | Yes |
| HTI | Haiti | AMRO | thousands | 131.0 (50.5–274.2) | 135.9 (49.8–303.4) | 4.81 (−25.6–54.2) | 0.00 (−0.02–0.04) | 36.79 | No |
| HND | Honduras | AMRO | thousands | 467.5 (235.2–773.3) | 679.3 (351.2–1091) | 209.8 (102.8–342.8) | 0.14 (0.07–0.23) | 0.00 | Yes |
| HUN | Hungary | EURO | thousands | 971.2 (420.2–1646) | 1173 (531.0–1900) | 192.7 (73.3–338.6) | 0.13 (0.05–0.23) | 0.06 | Yes |
| ISL | Iceland | EURO | thousands | 37.7 (29.5–46.7) | 41.2 (31.0–52.2) | 3.43 (−0.961–8.05) | 0.00 (0.00–0.01) | 6.11 | No |
| IND | India | SEARO | thousands | 41070 (31914–51417) | 65995 (49882–84657) | 24874 (16014–35448) | 16.58 (10.68–23.63) | 0.00 | Yes |
| IDN | Indonesia | WPRO | thousands | 3052 (1665–5154) | 4230 (2012–8328) | 1163 (−38.1–3650) | 0.78 (−0.03–2.43) | 3.03 | No |
| IRN | Iran (Islamic Republic of) | EMRO | thousands | 2870 (2308–3516) | 3371 (2238–4851) | 499.6 (−259.6–1525) | 0.33 (−0.17–1.02) | 10.83 | No |
| IRQ | Iraq | EMRO | thousands | 978.0 (445.4–1774) | 1421 (616.1–2716) | 435.9 (92.4–1064) | 0.29 (0.06–0.71) | 0.34 | Yes |
| IRL | Ireland | EURO | thousands | 301.5 (195.9–431.9) | 356.6 (220.8–528.5) | 54.6 (−3.71–127.6) | 0.04 (0.00–0.09) | 3.34 | No |
| ISR | Israel | EURO | thousands | 420.2 (303.8–557.8) | 470.3 (316.9–659.1) | 49.9 (−23.0–138.4) | 0.03 (−0.02–0.09) | 9.18 | No |
| ITA | Italy | EURO | thousands | 5392 (4325–6555) | 5882 (4270–7732) | 492.5 (−453.7–1571) | 0.33 (−0.30–1.05) | 15.52 | No |
| JAM | Jamaica | AMRO | thousands | 141.1 (91.0–203.8) | 168.9 (104.3–253.4) | 27.6 (2.66–62.1) | 0.02 (0.00–0.04) | 1.45 | Yes |
| JPN | Japan | WPRO | thousands | 11796 (10222–13469) | 12705 (10373–15231) | 909.6 (−436.2–2367) | 0.61 (−0.29–1.58) | 9.23 | No |
| JOR | Jordan | EMRO | thousands | 558.4 (412.0–724.1) | 781.9 (541.7–1059) | 223.5 (89.6–377.3) | 0.15 (0.06–0.25) | 0.05 | Yes |
| KAZ | Kazakhstan | EURO | thousands | 1607 (916.5–2398) | 2226 (1313–3201) | 611.7 (349.7–891.7) | 0.41 (0.23–0.59) | 0.00 | Yes |
| KEN | Kenya | AFRO | thousands | 400.7 (133.8–870.7) | 706.2 (242.8–1483) | 300.9 (100.9–645.1) | 0.20 (0.07–0.43) | 0.00 | Yes |
| KIR | Kiribati | WPRO | persons | 1423 (652.6–2621) | 1781 (794.1–3424) | 350.1 (54.2–938.7) | 0.00 (0.00–0.00) | 0.80 | Yes |
| KWT | Kuwait | EMRO | thousands | 326.6 (236.1–431.6) | 421.9 (284.3–587.0) | 94.9 (21.4–183.5) | 0.06 (0.01–0.12) | 0.58 | Yes |
| KGZ | Kyrgyzstan | EURO | thousands | 144.3 (69.2–256.2) | 201.3 (91.9–371.4) | 56.1 (15.6–129.5) | 0.04 (0.01–0.09) | 0.09 | Yes |
| LAO | Lao People's Democratic Republic | WPRO | thousands | 137.5 (66.9–239.0) | 203.4 (97.0–358.8) | 65.0 (23.2–132.8) | 0.04 (0.02–0.09) | 0.02 | Yes |
| LVA | Latvia | EURO | thousands | 129.7 (69.7–207.7) | 153.9 (83.0–243.2) | 23.5 (4.82–48.4) | 0.02 (0.00–0.03) | 0.67 | Yes |
| LBN | Lebanon | EMRO | thousands | 351.3 (227.8–497.9) | 439.6 (270.2–643.1) | 87.9 (11.8–180.1) | 0.06 (0.01–0.12) | 1.11 | Yes |

Countries 76–100 of 200; alphabetical order. Values: posterior median (95% UI). Status: fractions reportable (Yes/No).

### Supplementary Table S4. Country-level forecast contributions and Horiuchi decomposition, 2024–2030

#### Section A. Forecast levels, change, and contribution to the +150 million target



| ISO3 | Country | WHO region | Unit | C2024 (95% UI) | C2030 (95% UI) | ΔC (95% UI) | Contribution to +150 M % (95% UI) | Pr(ΔC ≤ 0) % | Status |
|---|---|---|---|---|---|---|---|---|---|
| LSO | Lesotho | AFRO | thousands | 64.8 (37.5–99.2) | 96.4 (55.9–146.0) | 31.2 (15.9–51.3) | 0.02 (0.01–0.03) | 0.00 | Yes |
| LBR | Liberia | AFRO | thousands | 73.1 (28.1–143.8) | 129.7 (50.9–248.3) | 55.9 (20.9–110.9) | 0.04 (0.01–0.07) | 0.00 | Yes |
| LBY | Libya | EMRO | thousands | 175.2 (77.9–316.9) | 280.6 (120.1–526.1) | 103.9 (29.1–233.3) | 0.07 (0.02–0.16) | 0.08 | Yes |
| LTU | Lithuania | EURO | thousands | 163.8 (89.6–259.1) | 208.1 (111.8–332.1) | 43.4 (10.1–90.2) | 0.03 (0.01–0.06) | 0.46 | Yes |
| LUX | Luxembourg | EURO | thousands | 48.9 (32.2–67.6) | 62.2 (40.8–85.9) | 13.2 (4.65–23.2) | 0.01 (0.00–0.02) | 0.12 | Yes |
| MDG | Madagascar | AFRO | thousands | 337.5 (98.6–755.1) | 728.2 (223.3–1524) | 384.4 (117.6–804.9) | 0.26 (0.08–0.54) | 0.00 | Yes |
| MWI | Malawi | AFRO | thousands | 195.0 (95.4–327.0) | 357.8 (185.9–581.0) | 161.1 (80.2–275.3) | 0.11 (0.05–0.18) | <0.01 | Yes |
| MYS | Malaysia | WPRO | thousands | 1544 (1120–2048) | 2195 (1539–3000) | 647.8 (325.9–1060) | 0.43 (0.22–0.71) | 0.00 | Yes |
| MDV | Maldives | SEARO | thousands | 12.9 (5.40–24.5) | 22.4 (9.56–41.4) | 9.41 (3.75–18.1) | 0.01 (0.00–0.01) | 0.00 | Yes |
| MLI | Mali | AFRO | thousands | 330.1 (136.0–626.5) | 539.7 (229.9–988.8) | 207.3 (86.1–384.2) | 0.14 (0.06–0.26) | 0.00 | Yes |
| MLT | Malta | EURO | thousands | 59.9 (38.9–83.6) | 65.8 (42.9–91.0) | 5.79 (−0.536–12.8) | 0.00 (0.00–0.01) | 3.53 | No |
| MHL | Marshall Islands | WPRO | persons | 684.9 (362.8–1152) | 685.4 (357.4–1166) | 0.012 (−93.2–103.2) | 0.00 (0.00–0.00) | 49.99 | No |
| MRT | Mauritania | AFRO | thousands | 73.5 (18.0–174.1) | 126.0 (32.7–279.1) | 51.6 (13.9–111.2) | 0.03 (0.01–0.07) | 0.00 | Yes |
| MUS | Mauritius | AFRO | thousands | 127.0 (94.1–160.7) | 166.6 (127.0–208.5) | 39.6 (22.1–58.9) | 0.03 (0.01–0.04) | 0.00 | Yes |
| MEX | Mexico | AMRO | thousands | 6429 (5168–7827) | 8531 (6550–10791) | 2093 (1022–3350) | 1.40 (0.68–2.23) | <0.01 | Yes |
| FSM | Micronesia (Federated States of) | WPRO | persons | 2298 (1385–3509) | 2999 (1760–4684) | 693.1 (274.9–1322) | 0.00 (0.00–0.00) | 0.02 | Yes |
| MDA | Moldova | EURO | thousands | 105.5 (43.6–200.9) | 128.0 (52.6–244.3) | 21.5 (2.37–55.6) | 0.01 (0.00–0.04) | 1.30 | Yes |
| MNG | Mongolia | WPRO | thousands | 188.4 (130.8–252.6) | 256.7 (177.1–345.5) | 67.9 (37.0–104.7) | 0.05 (0.02–0.07) | <0.01 | Yes |
| MNE | Montenegro | EURO | thousands | 55.3 (20.4–101.0) | 67.4 (25.9–117.1) | 11.5 (4.14–20.6) | 0.01 (0.00–0.01) | 0.03 | Yes |
| MAR | Morocco | EMRO | thousands | 677.8 (279.1–1325) | 808.2 (310.6–1688) | 126.0 (−62.0–492.6) | 0.08 (−0.04–0.33) | 10.35 | No |
| MOZ | Mozambique | AFRO | thousands | 331.1 (149.1–600.1) | 602.4 (269.8–1093) | 268.8 (110.4–519.7) | 0.18 (0.07–0.35) | 0.00 | Yes |
| MMR | Myanmar | SEARO | thousands | 1845 (1158–2722) | 2583 (1543–3969) | 730.2 (257.0–1410) | 0.49 (0.17–0.94) | 0.08 | Yes |
| NAM | Namibia | AFRO | thousands | 103.6 (63.1–153.3) | 157.9 (97.2–231.6) | 53.9 (29.7–85.1) | 0.04 (0.02–0.06) | 0.00 | Yes |
| NRU | Nauru | WPRO | persons | 268.3 (157.7–413.2) | 382.6 (225.2–587.8) | 113.4 (61.0–187.4) | 0.00 (0.00–0.00) | 0.00 | Yes |
| NPL | Nepal | SEARO | thousands | 427.9 (253.0–663.7) | 682.8 (395.0–1084) | 253.2 (123.2–449.3) | 0.17 (0.08–0.30) | 0.00 | Yes |

Countries 101–125 of 200; alphabetical order. Values: posterior median (95% UI). Status: fractions reportable (Yes/No).

#### Supplementary Table S4. Country-level forecast contributions and Horiuchi decomposition, 2024–2030

##### Section A. Forecast levels, change, and contribution to the +150 million target



| ISO3 | Country | WHO region | Unit | C2024 (95% UI) | C2030 (95% UI) | ΔC (95% UI) | Contribution to +150 M % (95% UI) | Pr(ΔC ≤ 0) % | Status |
|---|---|---|---|---|---|---|---|---|---|
| NLD | Netherlands | EURO | thousands | 1204 (911.9–1530) | 1460 (1001–2001) | 254.9 (−12.7–583.3) | 0.17 (−0.01–0.39) | 3.17 | No |
| NZL | New Zealand | WPRO | thousands | 313.0 (226.9–415.1) | 321.6 (215.4–453.3) | 8.71 (−42.6–69.3) | 0.01 (−0.03–0.05) | 37.16 | No |
| NIC | Nicaragua | AMRO | thousands | 372.2 (194.0–596.7) | 513.9 (274.6–802.6) | 140.3 (69.2–226.8) | 0.09 (0.05–0.15) | 0.00 | Yes |
| NER | Niger | AFRO | thousands | 231.6 (74.3–500.1) | 474.7 (155.2–1002) | 240.5 (75.2–524.2) | 0.16 (0.05–0.35) | 0.00 | Yes |
| NGA | Nigeria | AFRO | thousands | 3585 (1911–5871) | 6540 (3512–10555) | 2929 (1431–4996) | 1.95 (0.95–3.33) | 0.00 | Yes |
| NIU | Niue | WPRO | persons | 102.5 (54.4–166.3) | 117.2 (62.0–190.1) | 14.3 (2.03–31.6) | 0.00 (0.00–0.00) | 1.09 | Yes |
| MKD | North Macedonia | EURO | thousands | 165.1 (60.7–304.1) | 202.7 (77.9–355.1) | 36.0 (13.5–63.6) | 0.02 (0.01–0.04) | 0.02 | Yes |
| NOR | Norway | EURO | thousands | 387.7 (274.9–515.8) | 436.7 (294.5–602.0) | 48.5 (−18.4–124.4) | 0.03 (−0.01–0.08) | 7.75 | No |
| PSE | occupied Palestinian territory | EMRO | thousands | 140.4 (79.7–217.3) | 218.8 (114.1–358.8) | 78.0 (20.4–159.0) | 0.05 (0.01–0.11) | 0.21 | Yes |
| OMN | Oman | EMRO | thousands | 104.7 (58.2–170.1) | 149.6 (76.9–261.6) | 44.4 (8.55–104.7) | 0.03 (0.01–0.07) | 0.53 | Yes |
| PAK | Pakistan | EMRO | thousands | 4394 (2315–7366) | 5722 (2924–9934) | 1312 (350.7–2962) | 0.87 (0.23–1.97) | 0.22 | Yes |
| PLW | Palau | WPRO | persons | 948.8 (499.2–1534) | 1300 (695.9–2058) | 345.8 (164.2–587.2) | 0.00 (0.00–0.00) | <0.01 | Yes |
| PAN | Panama | AMRO | thousands | 274.8 (152.8–428.1) | 385.4 (220.0–584.8) | 109.6 (56.3–173.6) | 0.07 (0.04–0.12) | 0.00 | Yes |
| PNG | Papua New Guinea | WPRO | thousands | 137.0 (51.5–275.7) | 244.5 (93.1–481.8) | 106.3 (39.4–214.1) | 0.07 (0.03–0.14) | 0.00 | Yes |
| PRY | Paraguay | AMRO | thousands | 245.8 (118.4–425.9) | 352.3 (168.0–611.1) | 105.0 (40.7–204.6) | 0.07 (0.03–0.14) | <0.01 | Yes |
| PER | Peru | AMRO | thousands | 768.7 (565.3–1016) | 916.6 (593.3–1355) | 147.4 (−40.0–405.4) | 0.10 (−0.03–0.27) | 6.63 | No |
| PHL | Philippines | WPRO | thousands | 3244 (2078–4709) | 4732 (2919–7112) | 1475 (618.5–2672) | 0.98 (0.41–1.78) | 0.02 | Yes |
| POL | Poland | EURO | thousands | 5924 (4894–6983) | 7630 (6286–8981) | 1703 (1118–2302) | 1.14 (0.75–1.53) | 0.00 | Yes |
| PRT | Portugal | EURO | thousands | 1495 (1055–1947) | 1671 (1198–2143) | 172.5 (18.2–334.6) | 0.11 (0.01–0.22) | 1.50 | Yes |
| PRI | Puerto Rico | AMRO | thousands | 273.9 (103.3–514.3) | 301.2 (113.9–556.7) | 25.3 (−10.6–73.3) | 0.02 (−0.01–0.05) | 7.96 | No |
| QAT | Qatar | EMRO | thousands | 127.8 (77.6–192.7) | 159.5 (89.9–255.2) | 31.4 (−1.62–77.9) | 0.02 (0.00–0.05) | 3.18 | No |
| KOR | Republic of Korea | WPRO | thousands | 7085 (6397–7789) | 8684 (7466–9824) | 1601 (837.8–2266) | 1.07 (0.56–1.51) | 0.02 | Yes |
| ROU | Romania | EURO | thousands | 2360 (1492–3334) | 2875 (1874–3941) | 507.2 (267.9–772.4) | 0.34 (0.18–0.51) | <0.01 | Yes |
| RUS | Russian Federation | EURO | thousands | 8948 (6074–12425) | 10343 (6612–15158) | 1388 (−348.3–3645) | 0.93 (−0.23–2.43) | 6.02 | No |
| RWA | Rwanda | AFRO | thousands | 110.0 (36.2–239.8) | 222.4 (75.0–461.4) | 110.8 (36.8–232.5) | 0.07 (0.02–0.16) | 0.00 | Yes |

Countries 126–150 of 200; alphabetical order. Values: posterior median (95% UI). Status: fractions reportable (Yes/No).

#### Supplementary Table S4. Country-level forecast contributions and Horiuchi decomposition, 2024–2030

##### Section A. Forecast levels, change, and contribution to the +150 million target



| ISO3 | Country | WHO region | Unit | C2024 (95% UI) | C2030 (95% UI) | ΔC (95% UI) | Contribution to +150 M % (95% UI) | Pr(ΔC ≤ 0) % | Status |
|---|---|---|---|---|---|---|---|---|---|
| KNA | Saint Kitts and Nevis | AMRO | persons | 3216 (1635–5311) | 3965 (1978–6533) | 727.6 (182.8–1488) | 0.00 (0.00–0.00) | 0.38 | Yes |
| LCA | Saint Lucia | AMRO | thousands | 9.95 (5.39–16.0) | 12.2 (6.41–19.9) | 2.21 (0.481–4.68) | 0.00 (0.00–0.00) | 0.47 | Yes |
| VCT | Saint Vincent and the Grenadines | AMRO | persons | 5445 (2844–8928) | 6369 (3273–10470) | 897.8 (57.7–2056) | 0.00 (0.00–0.00) | 1.74 | Yes |
| WSM | Samoa | WPRO | thousands | 3.74 (2.13–5.99) | 4.82 (2.66–7.94) | 1.07 (0.334–2.26) | 0.00 (0.00–0.00) | 0.12 | Yes |
| STP | Sao Tome and Principe | AFRO | thousands | 3.81 (1.79–6.83) | 5.97 (2.76–10.8) | 2.13 (0.817–4.27) | 0.00 (0.00–0.00) | <0.01 | Yes |
| SAU | Saudi Arabia | EMRO | thousands | 995.9 (627.6–1465) | 1434 (842.4–2224) | 435.8 (125.2–869.6) | 0.29 (0.08–0.58) | 0.20 | Yes |
| SEN | Senegal | AFRO | thousands | 259.9 (79.7–565.4) | 466.2 (148.4–967.5) | 202.8 (64.3–423.7) | 0.14 (0.04–0.28) | 0.00 | Yes |
| SRB | Serbia | EURO | thousands | 782.8 (552.7–1032) | 940.7 (664.5–1238) | 157.1 (58.4–266.1) | 0.10 (0.04–0.18) | 0.12 | Yes |
| SYC | Seychelles | AFRO | thousands | 10.0 (6.48–14.1) | 14.3 (9.54–19.6) | 4.29 (2.75–6.05) | 0.00 (0.00–0.00) | 0.00 | Yes |
| SLE | Sierra Leone | AFRO | thousands | 121.7 (49.7–233.7) | 221.1 (91.8–416.3) | 98.0 (37.8–192.1) | 0.07 (0.03–0.13) | 0.00 | Yes |
| SGP | Singapore | WPRO | thousands | 509.0 (402.6–622.2) | 662.8 (517.9–814.4) | 153.4 (89.2–220.6) | 0.10 (0.06–0.15) | <0.01 | Yes |
| SVK | Slovakia | EURO | thousands | 726.4 (453.1–1023) | 878.2 (564.3–1201) | 149.1 (77.1–227.0) | 0.10 (0.05–0.15) | <0.01 | Yes |
| SVN | Slovenia | EURO | thousands | 203.7 (75.6–372.4) | 252.6 (98.5–439.2) | 47.3 (17.9–82.3) | 0.03 (0.01–0.05) | 0.02 | Yes |
| SLB | Solomon Islands | WPRO | thousands | 6.95 (2.99–13.2) | 13.6 (5.83–25.8) | 6.62 (2.69–13.1) | 0.00 (0.00–0.01) | 0.00 | Yes |
| SOM | Somalia | EMRO | thousands | 196.0 (56.6–438.5) | 366.1 (112.4–769.1) | 167.1 (52.8–347.5) | 0.11 (0.04–0.23) | 0.00 | Yes |
| ZAF | South Africa | AFRO | thousands | 3373 (2654–4171) | 4632 (3504–5920) | 1256 (649.5–1974) | 0.84 (0.43–1.32) | <0.01 | Yes |
| SSD | South Sudan | AFRO | thousands | 179.9 (45.6–420.0) | 312.7 (86.5–666.5) | 130.7 (39.0–262.0) | 0.09 (0.03–0.17) | 0.00 | Yes |
| ESP | Spain | EURO | thousands | 3708 (3006–4466) | 4164 (3116–5359) | 456.0 (−149.0–1142) | 0.30 (−0.10–0.76) | 7.01 | No |
| LKA | Sri Lanka | SEARO | thousands | 925.8 (553.4–1412) | 1257 (745.0–1918) | 328.2 (146.6–574.3) | 0.22 (0.10–0.38) | 0.02 | Yes |
| SDN | Sudan | EMRO | thousands | 634.1 (232.4–1268) | 1220 (466.9–2319) | 579.5 (224.8–1099) | 0.39 (0.15–0.73) | 0.00 | Yes |
| SUR | Suriname | AMRO | thousands | 29.9 (14.2–51.3) | 37.9 (17.9–65.1) | 7.84 (2.23–16.2) | 0.01 (0.00–0.01) | 0.16 | Yes |
| SWE | Sweden | EURO | thousands | 566.0 (414.1–739.0) | 640.5 (440.2–879.7) | 73.8 (−28.1–196.3) | 0.05 (−0.02–0.13) | 7.96 | No |
| CHE | Switzerland | EURO | thousands | 607.7 (434.6–805.1) | 667.5 (465.4–901.2) | 59.3 (−21.7–148.6) | 0.04 (−0.01–0.10) | 7.58 | No |
| SYR | Syrian Arab Republic | EMRO | thousands | 778.6 (273.6–1502) | 1121 (392.2–2166) | 335.8 (80.7–749.7) | 0.22 (0.05–0.50) | 0.13 | Yes |
| TWN | Taiwan, China | UNCLASSIFIED | thousands | 2086 (1683–2529) | 2393 (1778–3071) | 307.3 (−42.4–679.1) | 0.20 (−0.03–0.45) | 4.22 | No |

Countries 151–175 of 200; alphabetical order. Values: posterior median (95% UI). Status: fractions reportable (Yes/No).

#### Supplementary Table S4. Country-level forecast contributions and Horiuchi decomposition, 2024–2030

##### Section A. Forecast levels, change, and contribution to the +150 million target



| ISO3 | Country | WHO region | Unit | C2024 (95% UI) | C2030 (95% UI) | ΔC (95% UI) | Contribution to +150 M % (95% UI) | Pr(ΔC ≤ 0) % | Status |
|---|---|---|---|---|---|---|---|---|---|
| TJK | Tajikistan | EURO | thousands | 136.2 (52.3–283.6) | 184.2 (67.4–402.0) | 46.6 (7.86–133.3) | 0.03 (0.01–0.09) | 0.45 | Yes |
| TZA | Tanzania | AFRO | thousands | 611.0 (283.2–1089) | 1233 (573.7–2173) | 616.0 (263.4–1152) | 0.41 (0.18–0.77) | 0.00 | Yes |
| THA | Thailand | SEARO | thousands | 4885 (3524–6390) | 6555 (4687–8621) | 1658 (876.9–2565) | 1.11 (0.58–1.71) | <0.01 | Yes |
| TLS | Timor-Leste | SEARO | thousands | 21.8 (9.16–41.9) | 31.4 (13.0–61.2) | 9.31 (2.74–21.4) | 0.01 (0.00–0.01) | 0.06 | Yes |
| TGO | Togo | AFRO | thousands | 127.8 (52.2–243.9) | 225.0 (93.0–419.7) | 95.9 (37.2–186.9) | 0.06 (0.02–0.12) | 0.00 | Yes |
| TKL | Tokelau | WPRO | persons | 71.8 (35.1–124.4) | 111.5 (54.0–193.9) | 39.4 (17.2–73.3) | 0.00 (0.00–0.00) | 0.00 | Yes |
| TON | Tonga | WPRO | persons | 2091 (1151–3379) | 2803 (1538–4521) | 701.2 (305.3–1292) | 0.00 (0.00–0.00) | <0.01 | Yes |
| TTO | Trinidad and Tobago | AMRO | thousands | 99.9 (58.1–151.3) | 125.1 (71.3–191.5) | 24.8 (6.66–49.2) | 0.02 (0.00–0.03) | 0.31 | Yes |
| TUN | Tunisia | EMRO | thousands | 407.9 (246.7–613.5) | 571.3 (315.1–924.0) | 162.0 (28.3–360.2) | 0.11 (0.02–0.24) | 0.66 | Yes |
| TUR | Türkiye | EURO | thousands | 5334 (4254–6544) | 6208 (4172–8624) | 870.3 (−466.0–2470) | 0.58 (−0.31–1.65) | 10.78 | No |
| TKM | Turkmenistan | EURO | thousands | 139.8 (65.5–251.8) | 181.6 (81.0–341.4) | 40.6 (6.05–104.5) | 0.03 (0.00–0.07) | 0.82 | Yes |
| TUV | Tuvalu | WPRO | persons | 146.4 (52.8–310.0) | 167.1 (59.4–360.8) | 19.6 (−4.04–68.0) | 0.00 (0.00–0.00) | 5.35 | No |
| UGA | Uganda | AFRO | thousands | 365.4 (144.0–726.9) | 708.9 (283.4–1382) | 340.4 (131.8–682.1) | 0.23 (0.09–0.45) | 0.00 | Yes |
| UKR | Ukraine | EURO | thousands | 1930 (985.4–3224) | 2287 (1145–3874) | 345.8 (32.1–830.9) | 0.23 (0.02–0.55) | 1.46 | Yes |
| ARE | United Arab Emirates | EMRO | thousands | 361.8 (201.4–587.1) | 477.7 (248.7–822.0) | 114.9 (13.7–278.3) | 0.08 (0.01–0.19) | 1.15 | Yes |
| GBR | United Kingdom | EURO | thousands | 4209 (3576–4894) | 4153 (2947–5577) | −54.6 (−850.7–893.6) | −0.04 (−0.57–0.60) | 54.87 | No |
| USA | United States of America | AMRO | thousands | 33825 (29090–38814) | 31448 (24100–39579) | −2363 (−6847–2543) | −1.58 (−4.56–1.70) | 83.78 | No |
| URY | Uruguay | AMRO | thousands | 305.6 (210.6–411.9) | 378.0 (259.6–509.2) | 71.8 (30.7–118.1) | 0.05 (0.02–0.08) | 0.05 | Yes |
| UZB | Uzbekistan | EURO | thousands | 1383 (807.7–2163) | 1860 (1029–3040) | 470.1 (141.7–988.8) | 0.31 (0.09–0.66) | 0.11 | Yes |
| VUT | Vanuatu | WPRO | thousands | 3.02 (1.18–6.12) | 5.67 (2.20–11.5) | 2.62 (0.937–5.60) | 0.00 (0.00–0.00) | <0.01 | Yes |
| VEN | Venezuela | AMRO | thousands | 2250 (1548–3017) | 2921 (2012–3875) | 665.4 (338.5–1015) | 0.44 (0.23–0.68) | <0.01 | Yes |
| VNM | Viet Nam | WPRO | thousands | 2981 (2091–4069) | 5272 (3313–7732) | 2277 (1013–3938) | 1.52 (0.68–2.63) | <0.01 | Yes |
| YEM | Yemen | EMRO | thousands | 565.4 (285.3–970.1) | 893.5 (428.5–1596) | 323.2 (99.4–692.3) | 0.22 (0.07–0.46) | 0.04 | Yes |
| ZMB | Zambia | AFRO | thousands | 211.0 (87.6–396.6) | 424.1 (185.1–760.4) | 210.9 (91.3–383.4) | 0.14 (0.06–0.26) | 0.00 | Yes |
| ZWE | Zimbabwe | AFRO | thousands | 364.2 (102.5–780.4) | 536.2 (160.9–1082) | 167.4 (53.0–330.2) | 0.11 (0.04–0.22) | 0.00 | Yes |

Countries 176–200 of 200; alphabetical order. Values: posterior median (95% UI). Status: fractions reportable (Yes/No).

#### Section B. Absolute and fractional Horiuchi components



| ISO3 | Country | A unit | Population size D_N | Age–sex composition D_A | Age–sex-specific prevalence D_P | Control among people with hypertension, D_C | Demographic subtotal D_demo |
|---|---|---|---|---|---|---|---|
| AFG | Afghanistan | thousands | A: 235.4 (96.3–447.6)<br>F: 54.3 (42.0–76.2) | A: −8.15 (−15.8–−2.61)<br>F: −1.9 (−3.3–−0.8) | A: 43.8 (14.8–94.1)<br>F: 10.4 (5.2–17.1) | A: 155.8 (28.7–362.5)<br>F: 37.2 (12.3–51.5) | A: 227.0 (92.4–433.8)<br>F: 52.4 (40.5–73.5) |
| ALB | Albania | thousands | A: 2.38 (1.05–4.35)<br>F: 9.4 (5.9–22.9) | A: 1.65 (−0.481–4.93)<br>F: 6.5 (−3.5–20.1) | A: 1.08 (−2.54–5.60)<br>F: 4.6 (−13.3–20.7) | A: 19.3 (4.21–44.8)<br>F: 79.6 (49.8–96.5) | A: 4.05 (0.874–8.96)<br>F: 15.9 (5.8–39.9) |
| DZA | Algeria | thousands | A: 144.3 (77.8–234.6)<br>F: 27.4 (15.7–101.3) | A: 195.2 (106.9–306.1)<br>F: 37.0 (21.0–137.7) | A: −39.6 (−97.5–0.906)<br>F: −7.8 (−35.7–0.8) | A: 219.8 (−144.6–752.5)<br>F: 43.6 (−109.0–68.1) | A: 339.6 (185.0–540.2)<br>F: 64.3 (36.8–239.2) |
| ASM | American Samoa | persons | A: −154.4 (−262.5–−76.0)<br>F: −35.1 (−66.3–−23.8) | A: 210.7 (100.4–351.1)<br>F: 47.8 (31.5–90.4) | A: 30.7 (−36.6–115.9)<br>F: 7.4 (−10.7–22.8) | A: 343.4 (120.9–681.8)<br>F: 80.0 (59.4–96.1) | A: 55.1 (17.4–102.4)<br>F: 12.6 (5.1–26.6) |
| AND | Andorra | thousands | A: 0.501 (0.226–0.852)<br>F: 30.7 (17.7–83.6) | A: 0.750 (0.353–1.16)<br>F: 45.6 (26.1–125.8) | A: −0.358 (−0.782–−0.084)<br>F: −22.6 (−80.4–−4.7) | A: 0.740 (−0.235–1.92)<br>F: 46.8 (−38.7–70.5) | A: 1.25 (0.585–2.00)<br>F: 76.3 (44.5–208.4) |
| AGO | Angola | thousands | A: 146.7 (48.0–306.7)<br>F: 35.3 (26.9–50.1) | A: 10.6 (2.48–22.6)<br>F: 2.6 (1.1–4.3) | A: −6.96 (−49.2–25.8)<br>F: −1.9 (−11.2–5.3) | A: 261.3 (81.2–544.9)<br>F: 64.1 (49.3–73.9) | A: 157.5 (51.2–327.3)<br>F: 37.8 (28.8–53.7) |
| ATG | Antigua and Barbuda | thousands | A: 0.490 (0.183–0.926)<br>F: 32.7 (19.7–87.0) | A: 0.354 (0.111–0.625)<br>F: 23.3 (11.7–62.4) | A: 0.044 (−0.163–0.296)<br>F: 3.3 (−14.6–19.1) | A: 0.574 (−0.283–1.75)<br>F: 40.7 (−50.9–65.6) | A: 0.846 (0.306–1.52)<br>F: 56.0 (33.2–148.0) |
| ARG | Argentina | thousands | A: 140.4 (77.4–227.0)<br>F: NR | A: 44.0 (23.8–70.5)<br>F: NR | A: 57.0 (21.1–114.8)<br>F: NR | A: −50.4 (−386.0–346.3)<br>F: NR | A: 184.4 (101.8–296.2)<br>F: NR |
| ARM | Armenia | thousands | A: −0.718 (−1.30–−0.328)<br>F: −3.1 (−16.2–−1.5) | A: 7.05 (1.86–15.1)<br>F: 30.3 (8.9–149.5) | A: 0.520 (−1.68–3.08)<br>F: 2.4 (−14.1–21.7) | A: 14.9 (−4.09–47.3)<br>F: 70.4 (−44.2–94.2) | A: 6.33 (1.48–13.9)<br>F: 27.1 (6.4–134.5) |
| AUS | Australia | thousands | A: 63.3 (45.9–85.3)<br>F: NR | A: 39.4 (28.9–51.9)<br>F: NR | A: −11.8 (−61.0–36.4)<br>F: NR | A: −209.8 (−443.5–72.8)<br>F: NR | A: 102.7 (75.0–136.9)<br>F: NR |
| AUT | Austria | thousands | A: 2.69 (1.35–4.32)<br>F: NR | A: 43.0 (20.3–67.3)<br>F: NR | A: −58.1 (−110.4–−22.2)<br>F: NR | A: 88.5 (−26.4–227.8)<br>F: NR | A: 45.7 (21.7–71.4)<br>F: NR |
| AZE | Azerbaijan | thousands | A: 25.8 (13.4–43.4)<br>F: 27.0 (14.9–104.8) | A: 31.0 (12.2–57.6)<br>F: 32.3 (15.9–121.4) | A: 3.68 (−7.00–16.7)<br>F: 4.1 (−12.1–23.3) | A: 32.9 (−33.9–131.6)<br>F: 36.8 (−136.0–66.2) | A: 56.8 (26.2–100.1)<br>F: 59.3 (31.9–224.5) |
| BHS | Bahamas | thousands | A: 1.72 (0.916–2.75)<br>F: 30.6 (17.6–100.5) | A: 1.07 (0.539–1.66)<br>F: 18.9 (10.0–62.8) | A: 0.257 (−0.626–1.27)<br>F: 4.8 (−17.2–26.1) | A: 2.48 (−1.28–7.26)<br>F: 45.7 (−69.9–70.4) | A: 2.79 (1.49–4.35)<br>F: 49.5 (28.2–162.7) |
| BHR | Bahrain | thousands | A: 6.70 (2.28–13.6)<br>F: 35.0 (19.7–121.8) | A: 3.37 (1.20–5.88)<br>F: 17.4 (9.1–62.3) | A: −1.13 (−4.30–0.786)<br>F: −6.6 (−34.6–4.6) | A: 9.68 (−3.12–30.8)<br>F: 54.4 (−57.2–75.1) | A: 10.1 (3.53–19.3)<br>F: 52.4 (29.5–183.3) |
| BGD | Bangladesh | thousands | A: 548.0 (351.8–809.0)<br>F: 51.5 (31.9–137.4) | A: 104.1 (65.4–154.3)<br>F: 9.8 (5.9–26.2) | A: 145.6 (−19.8–364.0)<br>F: 14.0 (−2.9–40.0) | A: 255.9 (−344.4–1053)<br>F: 24.7 (−92.4–53.6) | A: 652.5 (419.4–959.5)<br>F: 61.3 (37.9–163.0) |
| BRB | Barbados | thousands | A: 0.431 (0.272–0.610)<br>F: 9.7 (4.9–43.2) | A: 1.29 (0.803–1.74)<br>F: 29.0 (13.6–130.2) | A: 0.241 (−0.614–1.16)<br>F: 5.6 (−26.3–37.4) | A: 2.39 (−1.23–6.32)<br>F: 55.7 (−84.1–81.6) | A: 1.73 (1.09–2.33)<br>F: 38.7 (18.8–173.3) |

Countries 1–16. A: absolute contribution in stated unit (95% UI); F: % of ΔC (95% UI); NR: not reportable.

### Supplementary Table S4. Country-level forecast contributions and Horiuchi decomposition, 2024–2030

#### Section B. Absolute and fractional Horiuchi components



| ISO3 | Country | A unit | Population size D_N | Age–sex composition D_A | Age–sex-specific prevalence D_P | Control among people with hypertension, D_C | Demographic subtotal D_demo |
|---|---|---|---|---|---|---|---|
| BLR | Belarus | thousands | A: −14.2 (−27.7–−5.60)<br>F: NR | A: 20.2 (5.43–44.6)<br>F: NR | A: −1.56 (−12.2–6.82)<br>F: NR | A: 58.9 (−11.0–182.7)<br>F: NR | A: 5.89 (−2.51–19.2)<br>F: NR |
| BEL | Belgium | thousands | A: 4.99 (3.71–6.41)<br>F: NR | A: 42.3 (32.5–52.1)<br>F: NR | A: −84.4 (−146.3–−30.4)<br>F: NR | A: 35.5 (−91.5–168.5)<br>F: NR | A: 47.3 (36.2–58.5)<br>F: NR |
| BLZ | Belize | thousands | A: 3.31 (1.73–5.45)<br>F: 52.5 (37.0–93.2) | A: 0.602 (0.335–0.894)<br>F: 9.5 (6.3–17.5) | A: 0.635 (−0.058–1.67)<br>F: 10.6 (−1.2–23.7) | A: 1.68 (−0.822–5.06)<br>F: 27.4 (−26.5–48.7) | A: 3.91 (2.08–6.32)<br>F: 62.0 (43.8–110.3) |
| BEN | Benin | thousands | A: 31.8 (17.6–51.9)<br>F: 44.1 (31.1–77.6) | A: 1.87 (1.01–3.06)<br>F: 2.6 (1.7–4.7) | A: −15.1 (−31.4–−3.76)<br>F: −21.5 (−55.0–−4.7) | A: 53.2 (18.9–106.9)<br>F: 74.8 (54.7–90.6) | A: 33.7 (18.7–54.9)<br>F: 46.7 (33.0–82.2) |
| BMU | Bermuda | persons | A: −82.7 (−153.9–−30.8)<br>F: −9.3 (−45.6–−4.3) | A: 326.7 (104.7–574.5)<br>F: 36.4 (14.2–174.4) | A: 37.7 (−119.7–233.5)<br>F: 4.7 (−26.5–36.0) | A: 559.4 (−159.0–1558)<br>F: 68.2 (−41.2–94.8) | A: 242.7 (67.8–434.1)<br>F: 27.0 (8.5–129.1) |
| BTN | Bhutan | thousands | A: 2.44 (1.18–4.30)<br>F: 34.2 (24.0–60.2) | A: 1.13 (0.560–1.91)<br>F: 15.8 (10.8–27.9) | A: 0.365 (−0.071–1.03)<br>F: 5.4 (−1.2–13.1) | A: 3.08 (0.111–7.91)<br>F: 44.7 (3.2–61.4) | A: 3.57 (1.75–6.18)<br>F: 50.0 (35.2–88.1) |
| BOL | Bolivia | thousands | A: 62.1 (21.8–126.7)<br>F: 51.5 (34.1–101.4) | A: 10.9 (4.27–19.1)<br>F: 9.0 (5.7–18.2) | A: 6.77 (−3.83–25.9)<br>F: 6.0 (−4.1–17.8) | A: 38.0 (−15.8–117.8)<br>F: 33.4 (−28.6–56.1) | A: 73.0 (26.2–145.2)<br>F: 60.4 (40.3–119.1) |
| BIH | Bosnia and Herzegovina | thousands | A: −10.8 (−16.5–−5.99)<br>F: −14.8 (−28.3–−9.9) | A: 11.2 (4.77–18.6)<br>F: 15.3 (8.2–30.2) | A: −1.49 (−8.95–5.34)<br>F: −2.2 (−15.2–6.9) | A: 73.1 (32.8–122.7)<br>F: 101.7 (91.1–116.1) | A: 0.381 (−3.47–4.36)<br>F: 0.5 (−6.0–6.5) |
| BWA | Botswana | thousands | A: 19.7 (11.8–29.7)<br>F: 43.6 (33.6–61.8) | A: 1.58 (0.835–2.52)<br>F: 3.6 (2.1–5.6) | A: −2.26 (−7.99–2.85)<br>F: −5.2 (−20.6–5.4) | A: 25.9 (11.8–44.1)<br>F: 58.1 (41.7–69.8) | A: 21.3 (12.8–31.9)<br>F: 47.2 (36.4–66.8) |
| BRA | Brazil | thousands | A: 1763 (1410–2140)<br>F: 24.0 (17.9–37.5) | A: 1196 (1010–1364)<br>F: 16.3 (11.9–26.2) | A: −469.2 (−1576–608.3)<br>F: −6.5 (−28.2–7.1) | A: 4868 (2204–7591)<br>F: 66.2 (47.3–79.7) | A: 2960 (2428–3493)<br>F: 40.3 (29.9–63.6) |
| BRN | Brunei Darussalam | thousands | A: 4.80 (3.11–6.75)<br>F: 28.8 (21.3–44.6) | A: 3.84 (2.64–4.92)<br>F: 22.9 (16.7–36.3) | A: 1.19 (−0.096–2.73)<br>F: 7.3 (−0.7–15.8) | A: 6.80 (0.913–13.4)<br>F: 41.0 (9.4–56.5) | A: 8.65 (5.76–11.6)<br>F: 51.7 (38.3–80.6) |
| BGR | Bulgaria | thousands | A: −51.8 (−92.7–−19.4)<br>F: −46.3 (−141.6–−25.0) | A: 25.1 (9.78–42.1)<br>F: 22.5 (12.0–67.2) | A: −0.056 (−23.8–23.4)<br>F: 0.0 (−34.4–20.2) | A: 134.0 (50.5–234.3)<br>F: 123.6 (102.5–200.7) | A: −26.5 (−53.1–−8.32)<br>F: −23.8 (−76.8–−10.8) |
| BFA | Burkina Faso | thousands | A: 55.1 (21.2–106.9)<br>F: 35.1 (26.7–50.9) | A: 3.74 (1.45–7.21)<br>F: 2.4 (1.5–3.9) | A: −10.2 (−32.7–3.88)<br>F: −7.0 (−20.4–2.3) | A: 106.7 (37.9–219.0)<br>F: 69.6 (55.3–79.5) | A: 58.8 (22.9–113.7)<br>F: 37.5 (28.6–54.4) |
| BDI | Burundi | thousands | A: 38.6 (9.94–87.3)<br>F: 29.4 (22.0–42.3) | A: 12.5 (3.33–25.7)<br>F: 9.7 (5.6–14.9) | A: −12.7 (−40.3–0.172)<br>F: −10.5 (−27.6–0.1) | A: 92.0 (26.8–188.7)<br>F: 71.6 (59.0–82.8) | A: 51.3 (13.7–111.5)<br>F: 39.0 (29.5–55.4) |
| CPV | Cabo Verde | thousands | A: 2.47 (1.34–3.87)<br>F: 29.0 (21.4–44.2) | A: 1.49 (0.784–2.27)<br>F: 17.4 (12.4–27.0) | A: −0.642 (−1.78–0.188)<br>F: −7.9 (−23.5–2.1) | A: 5.13 (2.01–9.26)<br>F: 61.6 (41.8–73.7) | A: 3.96 (2.14–6.12)<br>F: 46.4 (34.2–71.0) |
| KHM | Cambodia | thousands | A: 58.7 (31.3–95.6)<br>F: 26.6 (18.8–46.2) | A: 34.9 (18.0–55.4)<br>F: 15.7 (10.6–27.8) | A: 32.9 (7.78–71.7)<br>F: 15.3 (4.5–29.9) | A: 91.7 (2.25–212.3)<br>F: 42.4 (2.1–59.7) | A: 93.7 (49.8–150.1)<br>F: 42.3 (29.9–73.5) |

Countries 17–32. A: absolute contribution in stated unit (95% UI); F: % of ΔC (95% UI); NR: not reportable.

#### Supplementary Table S4. Country-level forecast contributions and Horiuchi decomposition, 2024–2030





| ISO3 | Country | A unit | Population size D_N | Age–sex composition D_A | Age–sex-specific prevalence D_P | Control among people with hypertension, D_C | Demographic subtotal D_demo |
|---|---|---|---|---|---|---|---|
| CMR | Cameroon | thousands | A: 71.6 (34.2–126.3)<br>F: 33.4 (25.7–48.1) | A: 11.2 (5.62–18.5)<br>F: 5.2 (3.9–7.6) | A: 6.72 (−8.98–27.4)<br>F: 3.4 (−4.7–10.4) | A: 121.9 (43.7–250.2)<br>F: 58.1 (40.1–68.4) | A: 82.7 (39.9–144.7)<br>F: 38.6 (29.8–55.5) |
| CAN | Canada | thousands | A: 210.6 (170.5–253.5)<br>F: NR | A: 88.9 (71.1–106.3)<br>F: NR | A: −191.6 (−292.4–−98.9)<br>F: NR | A: −38.7 (−501.8–385.2)<br>F: NR | A: 299.8 (246.2–354.2)<br>F: NR |
| CAF | Central African Republic | thousands | A: 11.0 (3.86–23.1)<br>F: 43.8 (32.9–66.5) | A: −2.67 (−5.37–−0.991)<br>F: −10.8 (−19.5–−5.9) | A: −0.676 (−3.24–1.08)<br>F: −3.0 (−12.0–3.9) | A: 17.3 (4.93–40.6)<br>F: 70.1 (54.0–80.0) | A: 8.31 (2.65–18.3)<br>F: 33.0 (23.8–50.4) |
| TCD | Chad | thousands | A: 77.7 (26.3–159.0)<br>F: 41.5 (31.4–60.4) | A: 0.523 (−1.12–2.43)<br>F: 0.3 (−0.7–1.2) | A: −9.05 (−34.8–6.99)<br>F: −5.4 (−17.9–3.3) | A: 116.2 (36.4–243.6)<br>F: 63.7 (47.4–74.3) | A: 78.3 (26.4–160.2)<br>F: 41.8 (31.6–60.9) |
| CHL | Chile | thousands | A: 145.9 (107.5–188.9)<br>F: 39.9 (25.2–101.1) | A: 57.9 (41.3–76.0)<br>F: 15.8 (9.6–40.5) | A: −108.8 (−215.0–−10.3)<br>F: −30.1 (−118.9–−1.9) | A: 272.4 (64.6–494.9)<br>F: 74.4 (40.6–104.4) | A: 204.0 (149.7–263.2)<br>F: 55.7 (35.0–141.3) |
| CHN | China | thousands | A: 1082 (829.3–1391)<br>F: NR | A: 4472 (3433–5704)<br>F: NR | A: −3176 (−8820–2646)<br>F: NR | A: 7084 (−1179–17596)<br>F: NR | A: 5554 (4272–7081)<br>F: NR |
| COL | Colombia | thousands | A: 532.2 (377.5–702.1)<br>F: 34.1 (25.6–50.7) | A: 176.4 (116.5–231.3)<br>F: 11.2 (7.9–17.3) | A: −90.2 (−265.2–81.2)<br>F: −5.9 (−20.5–4.6) | A: 943.8 (413.3–1509)<br>F: 60.6 (41.7–72.6) | A: 708.4 (497.3–928.9)<br>F: 45.3 (33.7–67.9) |
| COM | Comoros | thousands | A: 2.66 (1.20–4.70)<br>F: 25.6 (19.2–37.3) | A: 0.320 (0.148–0.549)<br>F: 3.1 (1.8–5.1) | A: −1.16 (−2.98–−0.014)<br>F: −11.8 (−30.1–−0.1) | A: 8.55 (3.81–14.9)<br>F: 83.2 (71.5–95.2) | A: 2.98 (1.38–5.19)<br>F: 28.7 (21.8–41.6) |
| COG | Congo | thousands | A: 21.0 (6.04–45.8)<br>F: 27.6 (20.7–40.6) | A: 5.64 (1.53–11.3)<br>F: 7.4 (4.7–11.5) | A: −2.56 (−11.4–2.69)<br>F: −3.8 (−13.4–3.2) | A: 51.0 (14.0–112.5)<br>F: 68.9 (54.1–78.1) | A: 26.7 (7.67–56.8)<br>F: 35.0 (26.1–51.5) |
| COK | Cook Islands | persons | A: −144.1 (−222.7–−82.7)<br>F: NR | A: 112.9 (63.6–172.7)<br>F: NR | A: 4.47 (−20.6–31.6)<br>F: NR | A: 89.6 (9.36–196.0)<br>F: NR | A: −31.0 (−56.5–−14.4)<br>F: NR |
| CRI | Costa Rica | thousands | A: 65.0 (55.1–75.4)<br>F: 32.9 (25.5–46.8) | A: 38.5 (35.2–41.1)<br>F: 19.4 (14.8–28.4) | A: −16.9 (−50.0–16.7)<br>F: −8.6 (−33.1–7.1) | A: 111.5 (59.5–162.4)<br>F: 56.3 (40.2–70.0) | A: 103.5 (90.6–116.0)<br>F: 52.3 (40.4–74.9) |
| CIV | Côte d'Ivoire | thousands | A: 77.4 (26.7–158.6)<br>F: 23.7 (17.9–34.8) | A: 36.2 (13.5–66.9)<br>F: 11.1 (7.9–16.6) | A: −12.6 (−52.7–12.7)<br>F: −4.3 (−15.2–3.6) | A: 220.9 (70.4–474.6)<br>F: 69.5 (55.0–78.8) | A: 113.7 (40.5–224.4)<br>F: 34.8 (26.4–50.9) |
| HRV | Croatia | thousands | A: −12.0 (−15.8–−8.55)<br>F: −17.2 (−46.7–−10.5) | A: 9.17 (5.75–12.8)<br>F: 13.2 (7.4–36.1) | A: 3.94 (−7.96–16.0)<br>F: 5.8 (−16.9–25.3) | A: 67.9 (22.4–121.7)<br>F: 98.3 (80.6–123.5) | A: −2.81 (−5.09–−0.687)<br>F: −4.1 (−12.3–−1.0) |
| CUB | Cuba | thousands | A: −6.05 (−8.38–−4.06)<br>F: NR | A: 55.9 (35.8–76.8)<br>F: NR | A: 15.5 (−17.0–52.0)<br>F: NR | A: 65.0 (−80.9–225.1)<br>F: NR | A: 49.8 (31.1–69.2)<br>F: NR |
| CYP | Cyprus | thousands | A: 5.92 (2.61–10.2)<br>F: 30.5 (16.2–106.2) | A: 8.22 (3.74–13.3)<br>F: 42.2 (22.5–147.2) | A: −5.02 (−11.2–−1.18)<br>F: −26.8 (−118.2–−4.5) | A: 10.1 (−3.46–26.5)<br>F: 54.6 (−49.1–78.9) | A: 14.1 (6.35–23.5)<br>F: 72.8 (38.8–253.8) |
| CZE | Czechia | thousands | A: −83.4 (−100.9–−66.4)<br>F: −43.9 (−135.4–−25.9) | A: 79.8 (65.1–93.9)<br>F: 42.0 (24.6–130.5) | A: −11.9 (−79.8–54.5)<br>F: −6.2 (−93.6–24.3) | A: 205.2 (85.2–328.3)<br>F: 108.1 (77.8–198.8) | A: −3.65 (−11.1–3.07)<br>F: −1.9 (−8.4–2.2) |

Countries 33–48. A: absolute contribution in stated unit (95% UI); F: % of ΔC (95% UI); NR: not reportable.

#### Supplementary Table S4. Country-level forecast contributions and Horiuchi decomposition, 2024–2030

##### Section B. Absolute and fractional Horiuchi components



| ISO3 | Country | A unit | Population size D_N | Age–sex composition D_A | Age–sex-specific prevalence D_P | Control among people with hypertension, D_C | Demographic subtotal D_demo |
|---|---|---|---|---|---|---|---|
| PRK | Democratic People's Republic of Korea | thousands | A: 58.5 (20.3–113.2)<br>F: 21.0 (12.0–60.2) | A: 62.5 (13.4–124.6)<br>F: 22.7 (7.5–63.5) | A: −4.77 (−68.2–57.4)<br>F: −1.9 (−36.4–18.1) | A: 154.0 (−10.6–390.8)<br>F: 58.3 (−6.9–82.1) | A: 121.7 (38.9–229.8)<br>F: 43.6 (23.2–120.1) |
| COD | Democratic Republic of the Congo | thousands | A: 394.9 (142.8–784.0)<br>F: 36.9 (28.4–51.4) | A: −23.1 (−43.3–−8.11)<br>F: −2.2 (−3.5–−1.2) | A: 15.9 (−65.7–116.8)<br>F: 1.7 (−6.3–8.7) | A: 666.6 (231.5–1330)<br>F: 63.6 (49.2–73.2) | A: 371.6 (133.1–744.2)<br>F: 34.7 (26.6–48.4) |
| DNK | Denmark | thousands | A: 1.80 (1.28–2.47)<br>F: NR | A: 0.161 (−0.579–0.897)<br>F: NR | A: −14.6 (−27.7–−4.31)<br>F: NR | A: 23.5 (−15.4–77.5)<br>F: NR | A: 1.95 (1.14–2.96)<br>F: NR |
| DJI | Djibouti | thousands | A: 4.18 (1.06–9.21)<br>F: 24.7 (18.2–36.8) | A: 1.45 (0.399–2.77)<br>F: 8.6 (5.5–13.1) | A: −1.69 (−5.32–0.020)<br>F: −10.9 (−29.4–0.1) | A: 12.8 (3.77–25.5)<br>F: 77.8 (65.2–89.3) | A: 5.63 (1.49–11.9)<br>F: 33.3 (24.7–49.0) |
| DMA | Dominica | persons | A: 64.8 (32.3–107.3)<br>F: 7.8 (4.2–30.2) | A: 153.7 (63.5–262.3)<br>F: 18.5 (8.3–72.4) | A: −34.8 (−210.9–121.7)<br>F: −4.4 (−48.5–15.9) | A: 627.7 (5.28–1492)<br>F: 78.4 (21.0–100.2) | A: 218.8 (99.1–363.3)<br>F: 26.2 (13.1–102.6) |
| DOM | Dominican Republic | thousands | A: 69.8 (42.8–102.8)<br>F: 42.6 (26.8–103.6) | A: 31.8 (20.0–43.4)<br>F: 19.3 (11.7–48.0) | A: 0.724 (−20.9–22.6)<br>F: 0.5 (−17.6–13.8) | A: 60.5 (−31.0–174.7)<br>F: 37.8 (−48.5–61.5) | A: 101.6 (63.1–145.5)<br>F: 61.9 (38.8–151.6) |
| ECU | Ecuador | thousands | A: 105.3 (71.0–147.3)<br>F: 46.5 (31.4–94.5) | A: 35.5 (24.7–46.9)<br>F: 15.6 (10.4–32.1) | A: 12.4 (−21.6–53.6)<br>F: 5.6 (−13.5–21.4) | A: 72.8 (−33.5–193.8)<br>F: 32.4 (−32.2–54.6) | A: 140.8 (95.9–193.8)<br>F: 62.1 (41.8–126.6) |
| EGY | Egypt | thousands | A: 530.8 (361.6–744.5)<br>F: 48.9 (23.7–261.6) | A: 156.5 (105.7–217.7)<br>F: 14.4 (6.8–77.9) | A: −27.3 (−185.4–127.6)<br>F: −2.5 (−39.4–18.3) | A: 400.1 (−488.5–1594)<br>F: 39.4 (−217.2–72.0) | A: 687.5 (467.8–960.6)<br>F: 63.3 (30.5–339.9) |
| SLV | El Salvador | thousands | A: 61.3 (40.9–85.2)<br>F: 55.9 (40.1–90.1) | A: −3.72 (−6.15–−1.75)<br>F: −3.4 (−6.2–−1.7) | A: 7.04 (−8.45–24.7)<br>F: 6.5 (−9.8–20.0) | A: 44.7 (6.00–86.8)<br>F: 41.0 (9.0–58.5) | A: 57.5 (38.4–80.0)<br>F: 52.5 (37.6–84.6) |
| GNQ | Equatorial Guinea | thousands | A: 5.66 (1.32–13.5)<br>F: 24.5 (18.1–36.1) | A: 2.71 (0.653–5.71)<br>F: 11.9 (7.1–18.2) | A: −0.607 (−3.46–1.12)<br>F: −3.1 (−13.0–4.2) | A: 15.0 (3.67–33.9)<br>F: 66.9 (51.7–77.0) | A: 8.40 (2.02–18.9)<br>F: 36.3 (26.8–52.9) |
| ERI | Eritrea | thousands | A: 12.4 (6.41–20.2)<br>F: 35.6 (27.7–48.7) | A: −2.13 (−3.31–−1.14)<br>F: −6.1 (−8.7–−4.4) | A: −1.89 (−5.85–1.32)<br>F: −5.7 (−17.9–3.3) | A: 26.3 (13.0–43.5)<br>F: 76.4 (65.6–85.6) | A: 10.2 (5.21–17.0)<br>F: 29.4 (22.7–40.6) |
| EST | Estonia | thousands | A: −3.33 (−4.77–−2.17)<br>F: −27.7 (−151.5–−12.3) | A: 2.94 (1.84–4.30)<br>F: 24.4 (10.5–133.1) | A: −2.19 (−5.18–0.282)<br>F: −18.3 (−134.6–10.6) | A: 14.2 (2.83–29.3)<br>F: 121.6 (92.5–252.9) | A: −0.382 (−0.903–0.051)<br>F: −3.2 (−20.4–2.0) |
| SWZ | Eswatini | thousands | A: 4.32 (2.25–7.12)<br>F: 33.3 (23.9–54.7) | A: 0.589 (0.247–1.04)<br>F: 4.6 (2.4–8.3) | A: −1.05 (−3.24–0.590)<br>F: −8.5 (−29.5–4.0) | A: 9.04 (3.34–17.3)<br>F: 70.8 (51.5–84.0) | A: 4.92 (2.58–8.05)<br>F: 37.9 (27.2–62.0) |
| ETH | Ethiopia | thousands | A: 273.8 (113.0–521.3)<br>F: 30.4 (23.9–42.1) | A: 5.89 (−2.02–16.6)<br>F: 0.7 (−0.3–1.7) | A: −27.5 (−135.8–60.8)<br>F: −3.4 (−15.6–5.6) | A: 640.3 (246.4–1277)<br>F: 72.3 (60.6–81.8) | A: 279.9 (115.2–532.3)<br>F: 31.1 (24.4–43.1) |
| FJI | Fiji | thousands | A: 1.73 (0.940–2.79)<br>F: 19.6 (13.7–34.1) | A: 0.846 (0.391–1.43)<br>F: 9.5 (5.8–17.3) | A: 1.22 (0.086–2.89)<br>F: 14.3 (1.2–29.3) | A: 4.91 (1.18–10.3)<br>F: 56.6 (26.4–71.7) | A: 2.58 (1.36–4.17)<br>F: 29.1 (20.2–50.7) |
| FIN | Finland | thousands | A: −12.6 (−15.8–−9.78)<br>F: NR | A: 0.459 (−0.312–1.28)<br>F: NR | A: −16.4 (−29.7–−4.44)<br>F: NR | A: 23.9 (−55.7–115.5)<br>F: NR | A: −12.1 (−15.3–−9.38)<br>F: NR |

Countries 49–64. A: absolute contribution in stated unit (95% UI); F: % of ΔC (95% UI); NR: not reportable.

#### Section B. Absolute and fractional Horiuchi components



| ISO3 | Country | A unit | Population size D_N | Age–sex composition D_A | Age–sex-specific prevalence D_P | Control among people with hypertension, D_C | Demographic subtotal D_demo |
|---|---|---|---|---|---|---|---|
| FRA | France | thousands | A: −55.6 (−71.5–−41.5)<br>F: NR | A: 137.7 (102.4–174.8)<br>F: NR | A: −149.3 (−308.5–−2.45)<br>F: NR | A: 422.4 (−326.1–1259)<br>F: NR | A: 81.9 (59.9–105.0)<br>F: NR |
| PYF | French Polynesia | thousands | A: 0.700 (0.337–1.22)<br>F: 16.3 (12.0–25.2) | A: 0.974 (0.426–1.73)<br>F: 22.5 (15.4–35.8) | A: 0.442 (−0.119–1.27)<br>F: 10.8 (−3.6–23.4) | A: 2.11 (0.603–4.45)<br>F: 50.4 (25.7–65.4) | A: 1.68 (0.769–2.93)<br>F: 38.8 (27.9–60.5) |
| GAB | Gabon | thousands | A: 9.08 (3.64–16.9)<br>F: 25.7 (19.6–36.5) | A: 4.19 (1.86–6.99)<br>F: 11.9 (8.7–17.1) | A: −0.712 (−4.05–1.96)<br>F: −2.2 (−11.4–4.9) | A: 22.3 (8.43–42.6)<br>F: 64.7 (50.1–74.0) | A: 13.3 (5.55–23.7)<br>F: 37.7 (29.0–53.0) |
| GMB | Gambia | thousands | A: 10.4 (4.40–19.2)<br>F: 35.0 (27.1–49.3) | A: 0.539 (−0.114–1.35)<br>F: 1.8 (−0.5–4.2) | A: −1.27 (−4.51–0.948)<br>F: −4.6 (−14.9–2.9) | A: 19.9 (7.59–38.2)<br>F: 67.9 (54.3–77.2) | A: 11.0 (4.54–20.1)<br>F: 36.8 (28.2–52.0) |
| GEO | Georgia | thousands | A: 0.735 (0.409–1.15)<br>F: 1.5 (0.8–5.8) | A: 10.1 (4.23–16.9)<br>F: 20.8 (9.6–79.2) | A: −1.16 (−7.81–4.75)<br>F: −2.5 (−28.2–11.9) | A: 37.3 (0.541–86.7)<br>F: 80.4 (26.5–96.1) | A: 10.8 (4.68–18.0)<br>F: 22.4 (10.6–84.9) |
| DEU | Germany | thousands | A: −150.9 (−181.2–−121.3)<br>F: NR | A: 616.7 (504.6–722.8)<br>F: NR | A: −681.1 (−1001–−382.7)<br>F: NR | A: 748.3 (−523.3–1976)<br>F: NR | A: 465.5 (379.4–546.9)<br>F: NR |
| GHA | Ghana | thousands | A: 194.1 (129.8–268.7)<br>F: 30.9 (23.7–44.9) | A: 63.9 (46.6–80.6)<br>F: 10.2 (7.6–15.1) | A: −97.8 (−200.2–−10.2)<br>F: −15.9 (−39.9–−1.4) | A: 467.5 (253.3–721.7)<br>F: 74.9 (61.2–88.7) | A: 258.1 (176.9–348.8)<br>F: 41.1 (31.5–59.9) |
| GRC | Greece | thousands | A: −40.6 (−53.9–−28.8)<br>F: NR | A: 66.1 (48.6–84.0)<br>F: NR | A: −32.8 (−72.1–1.83)<br>F: NR | A: 60.8 (−76.4–208.9)<br>F: NR | A: 25.4 (19.4–31.0)<br>F: NR |
| GRL | Greenland | persons | A: 72.7 (41.8–111.8)<br>F: 12.6 (4.2–74.5) | A: 198.6 (85.3–327.7)<br>F: 34.2 (2.2–199.2) | A: −292.3 (−551.0–−109.1)<br>F: −51.3 (−368.9–14.6) | A: 585.1 (50.7–1248)<br>F: 104.3 (54.1–218.4) | A: 271.7 (133.7–431.2)<br>F: 46.8 (11.3–273.3) |
| GRD | Grenada | thousands | A: 0.451 (0.232–0.736)<br>F: 28.5 (17.1–80.5) | A: 0.260 (0.101–0.450)<br>F: 16.3 (7.7–47.2) | A: 0.036 (−0.198–0.294)<br>F: 2.4 (−18.1–19.3) | A: 0.808 (−0.168–2.11)<br>F: 52.8 (−27.9–73.8) | A: 0.713 (0.351–1.16)<br>F: 44.8 (26.3–126.1) |
| GTM | Guatemala | thousands | A: 125.3 (59.3–211.1)<br>F: 37.6 (28.2–55.7) | A: 7.20 (2.81–12.5)<br>F: 2.2 (1.2–3.6) | A: 15.1 (−6.68–47.8)<br>F: 4.8 (−2.4–12.0) | A: 180.8 (60.1–344.4)<br>F: 55.4 (34.4–67.2) | A: 132.5 (62.7–222.7)<br>F: 39.8 (29.8–58.9) |
| GIN | Guinea | thousands | A: 47.1 (14.7–99.3)<br>F: 33.6 (25.4–49.5) | A: −5.71 (−11.0–−1.80)<br>F: −4.1 (−6.4–−2.6) | A: −7.35 (−26.5–2.80)<br>F: −5.8 (−17.2–1.9) | A: 104.3 (31.7–224.3)<br>F: 76.4 (64.0–85.1) | A: 41.4 (12.8–88.8)<br>F: 29.6 (22.2–43.6) |
| GNB | Guinea Bissau | thousands | A: 8.58 (2.13–19.7)<br>F: 36.2 (26.9–52.8) | A: 0.804 (0.128–1.76)<br>F: 3.4 (1.1–6.4) | A: −1.10 (−4.80–0.910)<br>F: −5.3 (−18.0–3.4) | A: 15.2 (3.99–33.0)<br>F: 65.8 (50.1–76.5) | A: 9.40 (2.36–21.2)<br>F: 39.6 (29.5–57.7) |
| GUY | Guyana | thousands | A: 3.52 (1.70–6.11)<br>F: 59.5 (29.3–292.6) | A: 0.119 (−0.198–0.493)<br>F: 2.0 (−7.1–15.3) | A: 0.096 (−1.08–1.36)<br>F: 1.8 (−35.5–31.1) | A: 1.98 (−2.81–8.24)<br>F: 36.8 (−197.8–71.2) | A: 3.65 (1.70–6.37)<br>F: 61.6 (29.8–300.9) |
| HTI | Haiti | thousands | A: 18.4 (6.99–39.3)<br>F: NR | A: 2.97 (1.18–6.02)<br>F: NR | A: 1.50 (−2.78–7.84)<br>F: NR | A: −16.0 (−56.3–17.7)<br>F: NR | A: 21.4 (8.20–45.2)<br>F: NR |
| HND | Honduras | thousands | A: 104.3 (53.6–169.5)<br>F: 49.5 (36.9–70.9) | A: 19.0 (10.3–27.8)<br>F: 9.0 (6.3–13.4) | A: −1.51 (−22.9–20.6)<br>F: −0.8 (−12.2–8.4) | A: 87.5 (24.9–165.8)<br>F: 42.3 (19.0–57.2) | A: 123.4 (64.2–196.5)<br>F: 58.5 (43.7–83.7) |

Countries 65–80. A: absolute contribution in stated unit (95% UI); F: % of ΔC (95% UI); NR: not reportable.

#### Section B. Absolute and fractional Horiuchi components



| ISO3 | Country | A unit | Population size D_N | Age–sex composition D_A | Age–sex-specific prevalence D_P | Control among people with hypertension, D_C | Demographic subtotal D_demo |
|---|---|---|---|---|---|---|---|
| HUN | Hungary | thousands | A: −38.6 (−63.7–−17.1)<br>F: −19.4 (−44.6–−11.5) | A: 35.5 (11.5–61.3)<br>F: 18.0 (7.4–41.5) | A: −4.89 (−39.0–24.5)<br>F: −2.6 (−27.2–11.5) | A: 202.0 (81.8–346.2)<br>F: 104.1 (87.6–134.5) | A: −2.93 (−23.4–14.8)<br>F: −1.5 (−15.1–8.8) |
| ISL | Iceland | thousands | A: 3.05 (2.35–3.80)<br>F: NR | A: 1.08 (0.816–1.34)<br>F: NR | A: −3.15 (−5.22–−1.23)<br>F: NR | A: 2.52 (−1.46–6.37)<br>F: NR | A: 4.13 (3.19–5.12)<br>F: NR |
| IND | India | thousands | A: 6457 (4975–8153)<br>F: 25.9 (20.9–34.7) | A: 2205 (1714–2717)<br>F: 8.8 (7.0–12.1) | A: −1198 (−3320–901.9)<br>F: −4.9 (−15.3–3.2) | A: 17464 (10096–26024)<br>F: 70.1 (59.1–78.5) | A: 8664 (6700–10856)<br>F: 34.7 (27.9–46.7) |
| IDN | Indonesia | thousands | A: 319.8 (165.9–577.9)<br>F: NR | A: 153.3 (75.4–281.9)<br>F: NR | A: 274.5 (113.5–570.4)<br>F: NR | A: 425.4 (−622.6–2386)<br>F: NR | A: 473.9 (243.9–855.4)<br>F: NR |
| IRN | Iran (Islamic Republic of) | thousands | A: 253.9 (188.2–334.7)<br>F: NR | A: 433.8 (324.4–564.8)<br>F: NR | A: −536.7 (−871.9–−245.6)<br>F: NR | A: 362.1 (−338.3–1240)<br>F: NR | A: 687.8 (512.6–899.2)<br>F: NR |
| IRQ | Iraq | thousands | A: 237.7 (106.5–439.2)<br>F: 53.1 (32.0–159.6) | A: 42.9 (19.3–74.0)<br>F: 9.6 (5.2–29.5) | A: 5.71 (−16.4–30.2)<br>F: 1.4 (−4.7–7.9) | A: 152.6 (−118.4–598.5)<br>F: 36.0 (−91.6–61.5) | A: 280.6 (127.5–509.9)<br>F: 62.6 (37.9–188.1) |
| IRL | Ireland | thousands | A: 15.5 (9.91–22.4)<br>F: NR | A: 19.7 (13.4–26.3)<br>F: NR | A: −14.1 (−25.3–−5.72)<br>F: NR | A: 34.2 (−21.5–101.1)<br>F: NR | A: 35.2 (23.4–48.7)<br>F: NR |
| ISR | Israel | thousands | A: 32.1 (22.7–43.5)<br>F: NR | A: 8.61 (5.85–11.8)<br>F: NR | A: −10.5 (−28.3–5.54)<br>F: NR | A: 20.2 (−48.3–99.4)<br>F: NR | A: 40.7 (28.7–55.0)<br>F: NR |
| ITA | Italy | thousands | A: −160.6 (−201.9–−123.7)<br>F: NR | A: 341.4 (270.3–415.6)<br>F: NR | A: −279.9 (−522.2–−60.8)<br>F: NR | A: 596.9 (−351.7–1664)<br>F: NR | A: 180.6 (144.0–217.1)<br>F: NR |
| JAM | Jamaica | thousands | A: 12.5 (7.94–18.3)<br>F: 44.0 (22.4–206.4) | A: 6.75 (4.38–9.45)<br>F: 23.8 (11.7–112.2) | A: 2.48 (−2.66–8.40)<br>F: 9.0 (−18.7–47.5) | A: 6.06 (−16.0–33.7)<br>F: 23.2 (−248.8–61.8) | A: 19.2 (12.4–27.5)<br>F: 67.9 (34.3–318.5) |
| JPN | Japan | thousands | A: −816.1 (−951.2–−690.4)<br>F: NR | A: 170.2 (122.1–221.8)<br>F: NR | A: −147.0 (−762.7–447.4)<br>F: NR | A: 1713 (422.7–3054)<br>F: NR | A: −645.8 (−749.9–−547.8)<br>F: NR |
| JOR | Jordan | thousands | A: 89.8 (64.8–118.2)<br>F: 40.0 (27.7–82.8) | A: 61.2 (46.7–75.2)<br>F: 27.2 (18.4–57.8) | A: −9.19 (−24.9–4.21)<br>F: −4.2 (−14.5–1.9) | A: 82.6 (−29.2–207.9)<br>F: 37.0 (−30.4–56.9) | A: 151.0 (111.7–193.1)<br>F: 67.3 (46.2–140.6) |
| KAZ | Kazakhstan | thousands | A: 107.2 (62.5–156.4)<br>F: 17.4 (12.9–24.9) | A: 88.4 (41.7–136.7)<br>F: 14.3 (8.5–22.2) | A: −8.42 (−65.6–45.5)<br>F: −1.4 (−12.1–6.8) | A: 423.7 (226.7–646.2)<br>F: 69.8 (55.6–80.7) | A: 196.1 (107.2–288.7)<br>F: 31.8 (22.2–46.3) |
| KEN | Kenya | thousands | A: 98.8 (33.7–208.5)<br>F: 32.3 (24.3–47.8) | A: 21.7 (7.53–43.2)<br>F: 7.1 (5.1–10.6) | A: −44.9 (−121.9–−6.99)<br>F: −15.6 (−37.5–−2.5) | A: 228.9 (75.8–489.3)<br>F: 76.3 (62.5–89.2) | A: 120.5 (41.3–251.4)<br>F: 39.4 (29.7–58.2) |
| KIR | Kiribati | persons | A: 229.2 (104.0–428.9)<br>F: 62.6 (34.0–244.7) | A: 25.2 (−0.74–65.6)<br>F: 7.0 (−1.0–28.9) | A: 154.5 (42.2–359.5)<br>F: 43.6 (15.1–146.4) | A: −47.2 (−327.9–264.9)<br>F: −13.2 (−308.7–37.7) | A: 254.9 (112.4–482.4)<br>F: 69.6 (37.5–269.0) |
| KWT | Kuwait | thousands | A: 28.2 (19.9–38.0)<br>F: 29.4 (17.7–94.8) | A: 13.4 (10.3–16.3)<br>F: 14.0 (7.8–47.8) | A: −7.44 (−22.1–6.46)<br>F: −7.9 (−43.8–7.0) | A: 61.3 (−6.07–140.0)<br>F: 64.6 (−11.2–82.1) | A: 41.6 (30.3–54.0)<br>F: 43.4 (25.7–142.6) |

Countries 81–96. A: absolute contribution in stated unit (95% UI); F: % of ΔC (95% UI); NR: not reportable.

#### Section B. Absolute and fractional Horiuchi components

| ISO3 | Country | A unit | Population size D_N | Age–sex composition D_A | Age–sex-specific prevalence D_P | Control among people with hypertension, D_C | Demographic subtotal D_demo |
|---|---|---|---|---|---|---|---|
| KGZ | Kyrgyzstan | thousands | A: 20.2 (9.50–36.4)<br>F: 35.3 (22.6–83.6) | A: 6.03 (1.33–14.0)<br>F: 10.4 (3.9–25.6) | A: 3.48 (−1.32–10.4)<br>F: 6.4 (−2.9–19.1) | A: 26.3 (−4.21–79.0)<br>F: 47.9 (−20.4–67.5) | A: 26.3 (11.2–49.7)<br>F: 45.7 (28.8–107.4) |
| LAO | Lao People's Democratic Republic | thousands | A: 26.5 (12.9–46.2)<br>F: 40.1 (27.5–76.7) | A: 5.68 (2.65–9.65)<br>F: 8.6 (5.6–16.6) | A: 2.76 (−4.63–12.4)<br>F: 4.5 (−8.5–16.8) | A: 30.0 (0.262–76.9)<br>F: 46.8 (1.0–64.4) | A: 32.2 (15.6–55.6)<br>F: 48.7 (33.3–93.2) |
| LVA | Latvia | thousands | A: −7.74 (−12.3–−4.18)<br>F: −32.0 (−114.3–−17.6) | A: 6.12 (2.99–9.94)<br>F: 25.2 (13.0–90.5) | A: −3.68 (−8.56–−0.396)<br>F: −15.7 (−73.7–−0.6) | A: 29.1 (9.48–55.3)<br>F: 122.6 (105.5–196.8) | A: −1.61 (−3.28–−0.359)<br>F: −6.8 (−26.8–−1.3) |
| LBN | Lebanon | thousands | A: 34.6 (22.1–49.5)<br>F: 38.7 (21.3–157.1) | A: 12.6 (8.16–17.1)<br>F: 14.0 (7.1–60.1) | A: −9.50 (−22.1–0.627)<br>F: −10.9 (−56.6–2.2) | A: 50.7 (−18.7–132.3)<br>F: 58.4 (−68.7–78.8) | A: 47.2 (30.7–65.9)<br>F: 52.8 (28.8–217.3) |
| LSO | Lesotho | thousands | A: 10.6 (6.15–16.0)<br>F: 33.6 (25.1–50.9) | A: 1.55 (0.596–2.65)<br>F: 5.0 (2.0–9.3) | A: −1.11 (−5.31–2.79)<br>F: −3.7 (−20.0–7.7) | A: 20.2 (8.60–35.3)<br>F: 65.1 (47.7–77.3) | A: 12.1 (7.28–18.1)<br>F: 38.7 (29.0–58.3) |
| LBR | Liberia | thousands | A: 17.2 (6.73–33.1)<br>F: 30.3 (23.0–44.3) | A: 0.629 (0.135–1.36)<br>F: 1.1 (0.3–2.1) | A: −3.22 (−10.7–1.47)<br>F: −6.2 (−18.5–2.4) | A: 41.6 (14.8–84.0)<br>F: 74.8 (62.1–84.0) | A: 17.8 (6.98–34.2)<br>F: 31.5 (23.9–46.0) |
| LBY | Libya | thousands | A: 26.5 (11.7–48.4)<br>F: 25.0 (16.4–57.7) | A: 29.1 (13.5–49.7)<br>F: 27.4 (17.6–64.3) | A: −7.72 (−20.2–−0.441)<br>F: −7.7 (−24.3–−0.4) | A: 56.8 (−1.26–156.0)<br>F: 55.4 (−3.1–71.4) | A: 55.7 (25.2–98.0)<br>F: 52.3 (34.2–121.5) |
| LTU | Lithuania | thousands | A: −3.55 (−5.62–−1.94)<br>F: −8.0 (−25.3–−4.7) | A: 4.50 (1.57–8.14)<br>F: 10.1 (4.1–32.9) | A: −5.59 (−12.3–−1.28)<br>F: −13.1 (−50.3–−2.4) | A: 48.3 (14.1–96.3)<br>F: 111.0 (100.0–144.9) | A: 0.932 (−1.03–3.16)<br>F: 2.1 (−3.6–10.2) |
| LUX | Luxembourg | thousands | A: 4.01 (2.66–5.50)<br>F: 30.2 (18.8–73.9) | A: 3.57 (2.37–4.76)<br>F: 26.8 (16.2–67.0) | A: −7.24 (−12.0–−3.32)<br>F: −55.3 (−185.7–−18.6) | A: 13.0 (5.54–21.3)<br>F: 98.4 (72.9–153.9) | A: 7.58 (5.05–10.2)<br>F: 56.9 (35.1–140.9) |
| MDG | Madagascar | thousands | A: 109.6 (33.3–233.2)<br>F: 28.1 (21.6–40.0) | A: 3.57 (−0.700–9.26)<br>F: 0.9 (−0.3–2.2) | A: −34.0 (−99.4–−2.46)<br>F: −9.5 (−22.8–−0.7) | A: 308.5 (94.3–639.7)<br>F: 80.5 (69.8–89.7) | A: 113.3 (33.9–240.3)<br>F: 29.0 (22.1–41.5) |
| MWI | Malawi | thousands | A: 64.7 (33.2–105.3)<br>F: 39.5 (29.5–58.7) | A: 1.46 (−2.00–5.98)<br>F: 0.9 (−1.7–3.0) | A: −38.1 (−79.2–−11.4)<br>F: −24.0 (−55.5–−6.7) | A: 135.4 (69.6–221.0)<br>F: 83.7 (69.7–101.9) | A: 66.1 (33.2–109.9)<br>F: 40.4 (30.0–59.7) |
| MYS | Malaysia | thousands | A: 249.4 (179.0–333.8)<br>F: 38.3 (27.6–64.8) | A: 116.2 (86.7–148.0)<br>F: 17.8 (12.6–30.6) | A: −25.6 (−121.9–65.3)<br>F: −4.0 (−24.3–8.8) | A: 311.3 (52.6–625.5)<br>F: 48.1 (14.6–64.1) | A: 365.6 (266.1–481.1)<br>F: 56.1 (40.4–95.5) |
| MDV | Maldives | thousands | A: 2.33 (0.990–4.32)<br>F: 24.5 (18.4–36.5) | A: 2.82 (1.26–4.84)<br>F: 29.6 (21.8–44.2) | A: 0.279 (−0.332–1.15)<br>F: 3.2 (−3.7–10.1) | A: 3.93 (0.773–9.12)<br>F: 42.7 (15.9–57.0) | A: 5.16 (2.27–9.12)<br>F: 54.1 (40.8–80.1) |
| MLI | Mali | thousands | A: 85.0 (35.8–157.3)<br>F: 40.5 (30.6–58.3) | A: 4.75 (1.28–9.52)<br>F: 2.3 (0.8–4.4) | A: −7.76 (−33.0–11.4)<br>F: −4.0 (−16.1–4.7) | A: 125.7 (46.7–242.4)<br>F: 61.2 (44.7–72.1) | A: 89.8 (38.5–165.1)<br>F: 42.9 (32.5–61.6) |
| MLT | Malta | thousands | A: 1.64 (1.07–2.27)<br>F: NR | A: 0.971 (0.469–1.60)<br>F: NR | A: −3.08 (−5.70–−0.935)<br>F: NR | A: 6.33 (0.300–12.7)<br>F: NR | A: 2.61 (1.62–3.80)<br>F: NR |
| MHL | Marshall Islands | persons | A: −176.4 (−297.1–−93.2)<br>F: NR | A: 154.3 (84.0–249.4)<br>F: NR | A: 7.93 (−33.6–55.3)<br>F: NR | A: 13.6 (−66.6–113.9)<br>F: NR | A: −21.8 (−54.0–−5.48)<br>F: NR |

Countries 97–112. A: absolute contribution in stated unit (95% UI); F: % of ΔC (95% UI); NR: not reportable.

#### Section B. Absolute and fractional Horiuchi components



| ISO3 | Country | A unit | Population size D_N | Age–sex composition D_A | Age–sex-specific prevalence D_P | Control among people with hypertension, D_C | Demographic subtotal D_demo |
|---|---|---|---|---|---|---|---|
| MRT | Mauritania | thousands | A: 19.4 (4.95–44.1)<br>F: 37.2 (27.6–54.7) | A: 0.799 (0.161–1.63)<br>F: 1.5 (0.7–2.6) | A: −2.43 (−10.3–1.87)<br>F: −5.3 (−17.6–3.2) | A: 33.8 (8.99–73.6)<br>F: 66.7 (50.9–77.2) | A: 20.2 (5.15–45.6)<br>F: 38.7 (28.7–56.9) |
| MUS | Mauritius | thousands | A: 5.10 (3.86–6.37)<br>F: 12.8 (8.9–22.0) | A: 7.63 (5.94–9.02)<br>F: 19.1 (12.8–33.7) | A: −11.4 (−22.5–−0.798)<br>F: −29.0 (−91.4–−1.5) | A: 38.5 (26.5–51.5)<br>F: 97.2 (75.8–138.5) | A: 12.7 (9.88–15.3)<br>F: 31.9 (21.8–55.6) |
| MEX | Mexico | thousands | A: 783.3 (619.2–966.9)<br>F: 37.3 (26.3–67.3) | A: 358.9 (293.3–426.5)<br>F: 17.1 (11.8–31.4) | A: −453.5 (−946.2–30.7)<br>F: −21.8 (−70.4–1.2) | A: 1417 (545.5–2366)<br>F: 67.4 (43.4–89.2) | A: 1142 (913.8–1391)<br>F: 54.4 (38.2–98.6) |
| FSM | Micronesia (Federated States of) | persons | A: 249.6 (149.0–384.3)<br>F: 35.6 (24.0–69.9) | A: −78.5 (−109.2–−52.6)<br>F: −11.4 (−24.9–−6.1) | A: 97.5 (−62.7–298.5)<br>F: 14.3 (−13.0–35.5) | A: 425.3 (108.9–878.2)<br>F: 61.5 (30.6–83.2) | A: 169.4 (84.9–295.8)<br>F: 24.2 (15.3–47.6) |
| MDA | Moldova | thousands | A: −8.49 (−16.1–−3.52)<br>F: −37.3 (−166.6–−18.9) | A: 4.65 (1.03–10.7)<br>F: 20.5 (5.3–90.8) | A: −2.99 (−7.97–−0.351)<br>F: −13.8 (−76.1–0.2) | A: 28.6 (7.29–66.0)<br>F: 130.7 (108.5–254.2) | A: −3.71 (−7.78–−0.959)<br>F: −16.7 (−83.3–−2.8) |
| MNG | Mongolia | thousands | A: 16.9 (11.8–22.6)<br>F: 24.8 (18.2–38.9) | A: 20.6 (13.9–27.1)<br>F: 30.0 (21.3–48.2) | A: −13.5 (−26.4–−2.47)<br>F: −20.1 (−51.3–−3.0) | A: 44.4 (20.0–72.6)<br>F: 65.4 (45.9–81.2) | A: 37.5 (25.8–49.6)<br>F: 54.8 (39.7–86.9) |
| MNE | Montenegro | thousands | A: −1.59 (−2.81–−0.600)<br>F: −13.3 (−28.5–−8.1) | A: 2.08 (0.560–3.83)<br>F: 17.4 (7.5–38.7) | A: −0.020 (−2.02–1.96)<br>F: −0.2 (−22.3–14.7) | A: 11.1 (4.08–19.7)<br>F: 96.4 (78.9–117.7) | A: 0.467 (−0.381–1.43)<br>F: 4.0 (−4.3–13.9) |
| MAR | Morocco | thousands | A: 68.0 (27.3–136.0)<br>F: NR | A: 50.8 (20.3–99.0)<br>F: NR | A: −28.8 (−74.8–−5.24)<br>F: NR | A: 42.1 (−146.1–338.9)<br>F: NR | A: 118.8 (47.9–234.5)<br>F: NR |
| MOZ | Mozambique | thousands | A: 100.0 (45.2–180.2)<br>F: 36.8 (28.3–53.1) | A: −5.66 (−13.0–−0.428)<br>F: −2.1 (−4.4–−0.2) | A: −17.6 (−55.9–8.89)<br>F: −6.9 (−21.4–2.9) | A: 193.6 (74.0–382.5)<br>F: 72.3 (58.9–82.6) | A: 94.2 (42.9–169.6)<br>F: 34.7 (26.7–50.1) |
| MMR | Myanmar | thousands | A: 178.6 (110.1–267.8)<br>F: 24.2 (15.9–53.7) | A: 74.5 (44.0–110.4)<br>F: 10.0 (6.3–22.7) | A: 55.7 (−21.3–155.9)<br>F: 7.8 (−3.7–22.3) | A: 423.0 (20.2–974.3)<br>F: 58.1 (8.0–73.7) | A: 253.2 (154.9–376.8)<br>F: 34.2 (22.4–76.4) |
| NAM | Namibia | thousands | A: 25.3 (15.6–37.0)<br>F: 46.5 (35.5–67.1) | A: 3.00 (1.90–4.14)<br>F: 5.5 (4.0–8.2) | A: −4.35 (−12.2–2.35)<br>F: −8.2 (−26.9–3.7) | A: 30.2 (13.6–51.0)<br>F: 56.4 (38.7–69.0) | A: 28.3 (17.5–41.0)<br>F: 52.0 (39.8–75.0) |
| NRU | Nauru | persons | A: 19.0 (11.2–29.1)<br>F: 16.6 (12.9–23.4) | A: 29.1 (16.8–44.4)<br>F: 25.4 (18.9–36.4) | A: 17.5 (−3.02–44.8)<br>F: 15.8 (−3.5–29.8) | A: 47.4 (19.6–86.8)<br>F: 42.3 (24.7–56.9) | A: 48.2 (28.2–73.2)<br>F: 42.0 (32.2–59.3) |
| NPL | Nepal | thousands | A: 61.0 (35.9–95.3)<br>F: 23.9 (18.0–36.0) | A: 2.54 (0.515–5.13)<br>F: 1.0 (0.2–2.0) | A: −1.77 (−15.7–12.0)<br>F: −0.7 (−6.6–4.4) | A: 191.6 (82.6–356.7)<br>F: 75.8 (63.1–82.9) | A: 63.6 (37.4–99.0)<br>F: 24.9 (18.8–37.6) |
| NLD | Netherlands | thousands | A: 32.5 (23.7–42.6)<br>F: NR | A: 11.1 (5.77–16.6)<br>F: NR | A: −127.9 (−258.9–−10.3)<br>F: NR | A: 343.6 (84.1–640.3)<br>F: NR | A: 43.7 (31.6–56.7)<br>F: NR |
| NZL | New Zealand | thousands | A: 12.1 (8.50–16.4)<br>F: NR | A: 12.9 (9.38–16.8)<br>F: NR | A: −12.7 (−21.1–−5.75)<br>F: NR | A: −3.25 (−53.5–54.6)<br>F: NR | A: 25.0 (18.0–33.0)<br>F: NR |
| NIC | Nicaragua | thousands | A: 67.3 (35.6–106.2)<br>F: 47.7 (34.8–71.7) | A: 17.6 (9.79–25.0)<br>F: 12.4 (8.7–19.1) | A: −5.59 (−23.9–11.3)<br>F: −4.2 (−20.1–7.0) | A: 61.2 (16.0–115.1)<br>F: 44.0 (18.7–59.8) | A: 85.0 (45.7–130.6)<br>F: 60.1 (44.0–90.2) |

Countries 113–128. A: absolute contribution in stated unit (95% UI); F: % of ΔC (95% UI); NR: not reportable.

### Supplementary Table S4. Country-level forecast contributions and Horiuchi decomposition, 2024–2030

#### Section B. Absolute and fractional Horiuchi components



| ISO3 | Country | A unit | Population size D_N | Age–sex composition D_A | Age–sex-specific prevalence D_P | Control among people with hypertension, D_C | Demographic subtotal D_demo |
|---|---|---|---|---|---|---|---|
| NER | Niger | thousands | A: 80.0 (26.2–169.0)<br>F: 32.8 (25.1–47.9) | A: −4.15 (−8.06–−1.47)<br>F: −1.7 (−2.9–−1.0) | A: −11.3 (−42.2–5.63)<br>F: −5.2 (−15.6–2.2) | A: 177.1 (52.8–391.9)<br>F: 74.1 (61.4–82.7) | A: 75.8 (24.5–161.8)<br>F: 31.1 (23.7–45.4) |
| NGA | Nigeria | thousands | A: 744.9 (401.8–1200)<br>F: 25.2 (19.5–36.2) | A: 85.9 (51.2–126.1)<br>F: 2.9 (2.1–4.4) | A: −22.6 (−221.0–168.6)<br>F: −0.8 (−7.9–5.1) | A: 2118 (939.1–3763)<br>F: 72.7 (60.5–80.1) | A: 831.6 (456.0–1322)<br>F: 28.2 (21.9–40.3) |
| NIU | Niue | persons | A: −2.83 (−4.58–−1.50)<br>F: −19.2 (−81.4–−10.0) | A: −0.32 (−3.23–2.78)<br>F: −2.1 (−50.0–22.3) | A: 4.11 (−0.60–10.7)<br>F: 29.0 (−11.7–99.4) | A: 13.2 (2.33–28.0)<br>F: 92.4 (49.5–174.6) | A: −3.11 (−6.45–−0.27)<br>F: −21.3 (−119.1–0.3) |
| MKD | North Macedonia | thousands | A: −2.97 (−5.31–−1.12)<br>F: −8.0 (−16.1–−5.0) | A: 5.62 (1.64–10.2)<br>F: 15.0 (7.1–31.5) | A: −0.217 (−6.33–5.68)<br>F: −0.7 (−21.7–13.4) | A: 33.7 (12.8–59.3)<br>F: 93.9 (77.3–112.1) | A: 2.60 (0.171–5.47)<br>F: 7.0 (0.7–17.0) |
| NOR | Norway | thousands | A: 11.6 (8.09–15.6)<br>F: NR | A: 10.9 (7.79–14.1)<br>F: NR | A: −29.3 (−49.0–−13.5)<br>F: NR | A: 56.0 (−10.8–130.7)<br>F: NR | A: 22.5 (16.0–29.5)<br>F: NR |
| PSE | occupied Palestinian territory | thousands | A: 32.5 (17.9–51.4)<br>F: 41.2 (26.6–108.9) | A: 6.46 (3.58–9.93)<br>F: 8.2 (5.2–21.8) | A: −4.88 (−12.8–0.886)<br>F: −6.5 (−24.1–1.2) | A: 44.3 (−3.71–109.7)<br>F: 57.1 (−13.1–73.1) | A: 39.0 (21.6–61.2)<br>F: 49.4 (31.8–130.6) |
| OMN | Oman | thousands | A: 20.3 (11.0–34.0)<br>F: 44.7 (26.3–153.5) | A: 8.34 (4.74–13.1)<br>F: 18.3 (10.5–63.2) | A: −4.34 (−11.0–0.065)<br>F: −10.0 (−43.6–0.6) | A: 20.6 (−9.69–68.0)<br>F: 47.0 (−79.4–69.5) | A: 28.7 (15.8–47.0)<br>F: 63.1 (37.0–216.1) |
| PAK | Pakistan | thousands | A: 770.7 (401.5–1304)<br>F: 57.2 (34.9–158.5) | A: 41.2 (15.9–76.7)<br>F: 3.1 (1.4–8.8) | A: 129.9 (33.1–286.7)<br>F: 9.9 (2.7–28.6) | A: 382.1 (−414.6–1554)<br>F: 29.9 (−93.2–57.2) | A: 812.0 (422.9–1374)<br>F: 60.3 (36.7–167.0) |
| PLW | Palau | persons | A: −20.7 (−32.9–−11.0)<br>F: −5.9 (−9.5–−4.2) | A: 79.9 (39.1–128.5)<br>F: 22.7 (15.2–37.5) | A: −6.86 (−64.8–46.9)<br>F: −2.1 (−22.8–11.3) | A: 294.8 (139.1–501.8)<br>F: 85.5 (70.7–101.3) | A: 59.2 (27.6–96.5)<br>F: 16.8 (10.7–28.2) |
| PAN | Panama | thousands | A: 37.5 (21.2–57.3)<br>F: 34.0 (24.9–51.6) | A: 13.6 (7.76–19.3)<br>F: 12.3 (8.5–19.1) | A: −2.07 (−13.7–9.16)<br>F: −2.0 (−14.3–7.4) | A: 60.6 (22.3–106.6)<br>F: 55.7 (34.0–68.6) | A: 51.1 (29.3–76.2)<br>F: 46.3 (33.8–70.2) |
| PNG | Papua New Guinea | thousands | A: 29.9 (11.3–59.2)<br>F: 27.8 (22.0–37.8) | A: 7.76 (2.65–15.1)<br>F: 7.2 (4.7–10.5) | A: 19.7 (4.38–49.6)<br>F: 19.2 (6.9–30.5) | A: 47.7 (15.6–102.6)<br>F: 45.8 (29.0–58.1) | A: 37.6 (14.1–73.8)<br>F: 35.0 (27.5–47.6) |
| PRY | Paraguay | thousands | A: 38.7 (18.6–66.7)<br>F: 36.3 (25.6–63.6) | A: 8.83 (4.20–14.6)<br>F: 8.3 (5.3–15.0) | A: −3.63 (−16.9–7.18)<br>F: −3.7 (−18.4–6.0) | A: 61.5 (14.1–135.0)<br>F: 59.2 (29.4–72.7) | A: 47.6 (23.1–80.8)<br>F: 44.6 (31.4–78.2) |
| PER | Peru | thousands | A: 92.9 (65.2–128.5)<br>F: NR | A: 38.2 (27.1–51.9)<br>F: NR | A: 21.7 (−14.1–66.6)<br>F: NR | A: −4.81 (−167.8–203.4)<br>F: NR | A: 131.1 (92.3–180.2)<br>F: NR |
| PHL | Philippines | thousands | A: 546.6 (346.2–802.8)<br>F: 36.7 (25.5–69.7) | A: 112.5 (74.3–153.5)<br>F: 7.5 (5.0–14.7) | A: 56.5 (−92.0–232.1)<br>F: 3.9 (−7.5–14.9) | A: 762.9 (64.7–1690)<br>F: 51.8 (9.9–67.4) | A: 659.4 (422.1–954.4)<br>F: 44.2 (30.7–84.3) |
| POL | Poland | thousands | A: −176.6 (−207.3–−146.4)<br>F: −10.4 (−14.8–−8.0) | A: 284.8 (231.1–335.3)<br>F: 16.7 (12.6–24.1) | A: 177.1 (−33.6–390.8)<br>F: 10.4 (−2.3–21.3) | A: 1420 (899.4–1941)<br>F: 83.3 (71.9–95.1) | A: 107.8 (75.3–138.6)<br>F: 6.3 (4.2–9.7) |
| PRT | Portugal | thousands | A: −47.9 (−61.7–−34.3)<br>F: −27.3 (−128.2–−12.6) | A: 75.0 (56.8–89.7)<br>F: 42.5 (19.8–201.2) | A: −67.3 (−120.9–−20.0)<br>F: −38.3 (−230.2–−4.0) | A: 214.6 (63.6–369.0)<br>F: 123.0 (92.2–259.6) | A: 26.8 (20.8–31.8)<br>F: 15.2 (6.7–73.6) |

Countries 129–144. A: absolute contribution in stated unit (95% UI); F: % of ΔC (95% UI); NR: not reportable.

#### Supplementary Table S4. Country-level forecast contributions and Horiuchi decomposition, 2024–2030

##### Section B. Absolute and fractional Horiuchi components



| ISO3 | Country | A unit | Population size D_N | Age–sex composition D_A | Age–sex-specific prevalence D_P | Control among people with hypertension, D_C | Demographic subtotal D_demo |
|---|---|---|---|---|---|---|---|
| PRI | Puerto Rico | thousands | A: −2.91 (−5.41–−1.11)<br>F: NR | A: 0.531 (−1.50–2.84)<br>F: NR | A: 1.97 (−5.64–11.5)<br>F: NR | A: 25.4 (−9.84–72.6)<br>F: NR | A: −2.30 (−5.39–−0.166)<br>F: NR |
| QAT | Qatar | thousands | A: 6.86 (4.05–10.6)<br>F: NR | A: 3.65 (2.27–5.21)<br>F: NR | A: −6.40 (−15.5–0.578)<br>F: NR | A: 27.6 (−4.13–71.2)<br>F: NR | A: 10.5 (6.40–15.7)<br>F: NR |
| KOR | Republic of Korea | thousands | A: 218.4 (193.2–242.9)<br>F: 13.7 (10.2–24.0) | A: 761.9 (701.4–812.6)<br>F: 47.6 (34.8–86.2) | A: −68.9 (−237.7–106.3)<br>F: −4.4 (−19.0–6.3) | A: 690.7 (−12.4–1285)<br>F: 43.1 (−1.3–58.7) | A: 980.3 (896.0–1054)<br>F: 61.3 (45.1–110.2) |
| ROU | Romania | thousands | A: −103.9 (−143.9–−67.0)<br>F: −20.2 (−36.5–−13.6) | A: 58.9 (22.2–96.8)<br>F: 11.5 (4.7–22.9) | A: 16.0 (−68.5–102.8)<br>F: 3.2 (−17.8–18.1) | A: 536.9 (308.7–785.8)<br>F: 105.6 (90.1–131.5) | A: −44.6 (−82.1–−11.3)<br>F: −8.8 (−19.7–−2.3) |
| RUS | Russian Federation | thousands | A: −385.0 (−545.5–−255.4)<br>F: NR | A: 421.6 (240.7–652.0)<br>F: NR | A: −631.1 (−1111–−274.4)<br>F: NR | A: 1991 (191.1–4341)<br>F: NR | A: 37.6 (−58.0–146.5)<br>F: NR |
| RWA | Rwanda | thousands | A: 27.7 (9.33–58.1)<br>F: 24.7 (19.2–35.0) | A: 3.83 (0.702–8.75)<br>F: 3.5 (1.1–6.3) | A: −9.87 (−28.3–−0.574)<br>F: −9.5 (−22.7–−0.5) | A: 89.9 (29.8–187.5)<br>F: 81.3 (71.0–90.8) | A: 31.6 (10.6–65.8)<br>F: 28.2 (21.7–40.0) |
| KNA | Saint Kitts and Nevis | persons | A: 193.6 (97.5–317.8)<br>F: 26.1 (15.5–74.0) | A: 207.0 (91.0–334.8)<br>F: 27.7 (14.7–79.6) | A: 1.03 (−117.8–122.2)<br>F: 0.2 (−23.8–16.6) | A: 331.4 (−127.0–922.9)<br>F: 46.2 (−47.6–69.9) | A: 401.1 (193.7–642.9)<br>F: 53.8 (31.3–152.8) |
| LCA | Saint Lucia | thousands | A: 0.900 (0.482–1.45)<br>F: 40.0 (23.6–125.5) | A: 0.405 (0.187–0.662)<br>F: 17.9 (9.6–56.0) | A: 0.093 (−0.241–0.482)<br>F: 4.4 (−16.1–24.1) | A: 0.821 (−0.614–2.69)<br>F: 37.8 (−87.5–64.3) | A: 1.31 (0.683–2.09)<br>F: 57.8 (33.8–181.3) |
| VCT | Saint Vincent and the Grenadines | persons | A: 28.2 (14.7–46.1)<br>F: 3.0 (1.4–15.3) | A: 312.1 (155.0–491.5)<br>F: 33.2 (14.9–168.6) | A: 23.9 (−145.6–208.2)<br>F: 2.8 (−30.1–32.3) | A: 539.0 (−243.2–1559)<br>F: 61.2 (−86.1–86.4) | A: 340.4 (170.5–536.1)<br>F: 36.2 (16.4–183.8) |
| WSM | Samoa | thousands | A: 0.076 (0.043–0.123)<br>F: 7.0 (4.4–17.3) | A: 0.047 (−0.0017–0.123)<br>F: 4.3 (−0.3–12.8) | A: 0.240 (−0.00079–0.583)<br>F: 22.7 (−0.3–54.8) | A: 0.700 (0.093–1.66)<br>F: 66.0 (23.4–87.3) | A: 0.124 (0.048–0.241)<br>F: 11.3 (5.7–28.5) |
| STP | Sao Tome and Principe | thousands | A: 0.703 (0.329–1.26)<br>F: 32.6 (23.5–54.5) | A: 0.152 (0.062–0.275)<br>F: 7.1 (4.2–12.3) | A: −0.099 (−0.404–0.131)<br>F: −4.9 (−20.6–5.5) | A: 1.38 (0.404–2.99)<br>F: 65.4 (42.4–77.6) | A: 0.856 (0.397–1.52)<br>F: 39.6 (28.4–66.2) |
| SAU | Saudi Arabia | thousands | A: 118.8 (73.1–178.0)<br>F: 27.0 (17.2–70.8) | A: 99.2 (66.1–135.6)<br>F: 22.5 (13.8–61.2) | A: −18.5 (−62.8–21.2)<br>F: −4.4 (−21.4–4.8) | A: 238.1 (−29.7–599.1)<br>F: 54.9 (−18.7–72.2) | A: 218.2 (139.8–312.4)<br>F: 49.5 (31.2–132.2) |
| SEN | Senegal | thousands | A: 80.5 (25.3–169.6)<br>F: 39.1 (29.9–55.6) | A: 0.456 (−1.16–2.34)<br>F: 0.2 (−0.7–1.2) | A: −5.77 (−29.0–9.74)<br>F: −3.2 (−13.0–4.1) | A: 128.0 (38.4–274.4)<br>F: 63.9 (48.8–73.8) | A: 81.1 (25.4–170.3)<br>F: 39.3 (30.1–55.9) |
| SRB | Serbia | thousands | A: −42.4 (−55.5–−30.1)<br>F: −26.8 (−64.5–−16.8) | A: 11.4 (4.41–18.8)<br>F: 7.2 (2.9–18.4) | A: −2.82 (−25.6–19.1)<br>F: −1.8 (−24.1–11.9) | A: 191.1 (90.9–301.2)<br>F: 121.4 (106.2–166.0) | A: −30.9 (−41.1–−21.4)<br>F: −19.6 (−48.2–−11.8) |
| SYC | Seychelles | thousands | A: 1.47 (0.970–2.03)<br>F: 34.1 (26.7–46.0) | A: 0.635 (0.400–0.853)<br>F: 14.6 (10.7–20.5) | A: −0.735 (−1.44–−0.168)<br>F: −17.3 (−39.7–−3.4) | A: 2.94 (1.87–4.13)<br>F: 68.8 (56.7–81.4) | A: 2.11 (1.38–2.87)<br>F: 48.7 (37.9–66.2) |
| SLE | Sierra Leone | thousands | A: 32.3 (13.4–60.9)<br>F: 32.5 (24.7–47.9) | A: 0.739 (0.269–1.38)<br>F: 0.7 (0.4–1.3) | A: −5.28 (−17.7–2.94)<br>F: −5.8 (−18.1–2.7) | A: 70.6 (25.5–142.0)<br>F: 72.5 (58.8–81.9) | A: 33.0 (13.7–62.2)<br>F: 33.3 (25.3–49.0) |

Countries 145–160. A: absolute contribution in stated unit (95% UI); F: % of ΔC (95% UI); NR: not reportable.

Supplementary Table S4. Country-level forecast contributions and Horiuchi decomposition, 2024–2030

#### Section B. Absolute and fractional Horiuchi components



| ISO3 | Country | A unit | Population size D_N | Age–sex composition D_A | Age–sex-specific prevalence D_P | Control among people with hypertension, D_C | Demographic subtotal D_demo |
|---|---|---|---|---|---|---|---|
| SGP | Singapore | thousands | A: 71.2 (56.3–86.9)<br>F: 46.4 (34.4–72.4) | A: 11.7 (8.13–15.1)<br>F: 7.6 (5.1–12.3) | A: −28.7 (−48.1–−10.5)<br>F: −18.9 (−40.6–−6.0) | A: 99.6 (43.6–155.9)<br>F: 64.9 (45.4–76.9) | A: 83.0 (65.1–101.4)<br>F: 54.1 (39.9–84.5) |
| SVK | Slovakia | thousands | A: −15.1 (−20.9–−9.62)<br>F: −10.0 (−18.1–−6.7) | A: 43.7 (26.3–60.0)<br>F: 28.9 (18.4–52.5) | A: −0.201 (−24.8–24.2)<br>F: −0.1 (−21.8–14.5) | A: 121.0 (57.4–189.6)<br>F: 81.4 (62.7–97.6) | A: 28.5 (16.1–39.9)<br>F: 18.8 (11.3–34.7) |
| SVN | Slovenia | thousands | A: −4.53 (−8.04–−1.73)<br>F: −9.3 (−18.3–−5.9) | A: 10.7 (3.59–18.7)<br>F: 21.9 (12.0–43.5) | A: −0.306 (−7.80–6.81)<br>F: −0.7 (−20.0–12.5) | A: 41.5 (15.4–73.3)<br>F: 88.4 (70.7–104.1) | A: 6.07 (1.56–11.3)<br>F: 12.5 (4.9–26.4) |
| SLB | Solomon Islands | thousands | A: 1.94 (0.834–3.65)<br>F: 29.0 (23.1–39.3) | A: 0.368 (0.151–0.689)<br>F: 5.5 (3.8–8.0) | A: 0.989 (0.263–2.34)<br>F: 15.3 (5.9–24.7) | A: 3.28 (1.11–7.05)<br>F: 50.1 (33.7–61.2) | A: 2.31 (0.996–4.32)<br>F: 34.6 (27.4–46.8) |
| SOM | Somalia | thousands | A: 69.9 (21.1–149.8)<br>F: 41.2 (31.7–57.7) | A: −5.35 (−10.7–−1.64)<br>F: −3.2 (−5.0–−1.8) | A: −13.1 (−42.3–1.49)<br>F: −8.4 (−22.6–0.9) | A: 116.9 (37.4–239.8)<br>F: 70.5 (57.7–80.8) | A: 64.5 (19.1–140.0)<br>F: 38.0 (29.0–53.5) |
| ZAF | South Africa | thousands | A: 390.5 (303.3–488.0)<br>F: 31.0 (22.3–52.6) | A: 227.9 (183.7–272.2)<br>F: 18.1 (12.7–31.4) | A: −129.5 (−412.5–143.1)<br>F: −10.4 (−48.7–9.0) | A: 774.3 (301.6–1300)<br>F: 61.5 (38.4–81.3) | A: 618.4 (487.7–758.9)<br>F: 49.1 (35.1–83.9) |
| SSD | South Sudan | thousands | A: 35.6 (9.57–78.6)<br>F: 26.9 (19.9–39.3) | A: 11.7 (2.71–24.0)<br>F: 9.1 (4.0–14.6) | A: −12.9 (−40.9–0.379)<br>F: −10.7 (−28.4–0.3) | A: 97.4 (29.7–191.7)<br>F: 75.0 (62.2–86.8) | A: 47.6 (13.0–100.7)<br>F: 35.9 (26.4–51.8) |
| ESP | Spain | thousands | A: −43.2 (−53.4–−34.0)<br>F: NR | A: 381.6 (308.2–458.0)<br>F: NR | A: −346.5 (−625.8–−91.0)<br>F: NR | A: 471.9 (−89.8–1074)<br>F: NR | A: 338.4 (274.1–404.6)<br>F: NR |
| LKA | Sri Lanka | thousands | A: 56.9 (34.0–86.6)<br>F: 17.2 (12.1–30.6) | A: 40.0 (21.9–60.8)<br>F: 12.0 (7.9–21.7) | A: 36.4 (7.97–76.5)<br>F: 11.3 (2.8–23.1) | A: 194.6 (47.0–389.1)<br>F: 59.6 (28.8–72.8) | A: 97.0 (56.4–146.7)<br>F: 29.1 (20.2–52.1) |
| SDN | Sudan | thousands | A: 213.3 (80.7–412.0)<br>F: 36.4 (28.8–49.0) | A: 3.54 (−4.66–12.5)<br>F: 0.6 (−1.0–2.3) | A: −23.8 (−88.1–18.9)<br>F: −4.5 (−14.7–2.8) | A: 387.5 (149.9–737.7)<br>F: 67.5 (56.0–76.4) | A: 217.4 (81.5–415.9)<br>F: 37.1 (29.2–49.9) |
| SUR | Suriname | thousands | A: 3.42 (1.63–5.86)<br>F: 42.7 (26.5–108.1) | A: 1.31 (0.569–2.13)<br>F: 16.2 (8.9–41.9) | A: 0.230 (−0.786–1.39)<br>F: 3.1 (−13.0–17.6) | A: 2.93 (−1.58–8.94)<br>F: 38.1 (−53.6–62.1) | A: 4.74 (2.26–7.88)<br>F: 58.9 (36.5–149.4) |
| SWE | Sweden | thousands | A: 4.02 (2.88–5.33)<br>F: NR | A: 7.31 (4.60–10.4)<br>F: NR | A: −61.6 (−103.6–−27.8)<br>F: NR | A: 125.4 (21.6–248.4)<br>F: NR | A: 11.3 (7.80–15.4)<br>F: NR |
| CHE | Switzerland | thousands | A: 9.20 (6.54–12.2)<br>F: NR | A: 44.4 (32.4–57.0)<br>F: NR | A: −67.2 (−107.5–−33.4)<br>F: NR | A: 74.1 (−2.75–155.4)<br>F: NR | A: 53.6 (39.0–69.2)<br>F: NR |
| SYR | Syrian Arab Republic | thousands | A: 271.4 (95.8–520.0)<br>F: 78.7 (49.7–187.8) | A: −26.5 (−51.4–−9.18)<br>F: −7.8 (−19.4–−4.3) | A: −38.2 (−106.0–−2.61)<br>F: −11.9 (−40.0–−0.7) | A: 133.9 (−46.3–402.5)<br>F: 41.2 (−37.7–63.5) | A: 244.5 (85.6–472.1)<br>F: 70.9 (44.6–169.0) |
| TWN | Taiwan, China | thousands | A: 0.456 (0.357–0.566)<br>F: NR | A: 227.5 (181.5–274.0)<br>F: NR | A: −19.0 (−103.0–70.2)<br>F: NR | A: 99.9 (−222.1–427.5)<br>F: NR | A: 227.9 (181.9–274.5)<br>F: NR |
| TJK | Tajikistan | thousands | A: 26.9 (10.1–56.9)<br>F: 55.3 (32.1–182.3) | A: 5.13 (1.03–12.9)<br>F: 10.6 (3.7–35.0) | A: −0.629 (−6.73–5.32)<br>F: −1.4 (−21.0–10.6) | A: 15.8 (−14.6–73.1)<br>F: 35.6 (−105.9–63.2) | A: 32.0 (11.6–69.1)<br>F: 65.9 (38.2–215.5) |

Countries 161–176. A: absolute contribution in stated unit (95% UI); F: % of ΔC (95% UI); NR: not reportable.

#### Supplementary Table S4. Country-level forecast contributions and Horiuchi decomposition, 2024–2030

##### Section B. Absolute and fractional Horiuchi components



| ISO3 | Country | A unit | Population size D_N | Age–sex composition D_A | Age–sex-specific prevalence D_P | Control among people with hypertension, D_C | Demographic subtotal D_demo |
|---|---|---|---|---|---|---|---|
| TZA | Tanzania | thousands | A: 194.9 (91.5–341.5)<br>F: 31.3 (24.1–45.3) | A: 1.29 (−5.38–8.05)<br>F: 0.2 (−0.9–1.4) | A: −97.7 (−216.4–−24.2)<br>F: −16.4 (−36.7–−3.8) | A: 523.4 (223.5–964.1)<br>F: 84.9 (73.1–97.8) | A: 196.1 (92.9–343.0)<br>F: 31.5 (24.3–45.5) |
| THA | Thailand | thousands | A: 127.0 (91.7–165.9)<br>F: 7.6 (5.5–12.6) | A: 477.9 (365.0–583.4)<br>F: 28.6 (20.3–48.3) | A: 289.2 (83.4–538.1)<br>F: 17.6 (5.8–32.3) | A: 765.3 (118.0–1493)<br>F: 46.2 (12.5–62.1) | A: 605.0 (457.5–747.8)<br>F: 36.2 (25.9–60.9) |
| TLS | Timor-Leste | thousands | A: 4.59 (1.92–8.84)<br>F: 48.0 (31.1–107.6) | A: −0.928 (−1.95–−0.241)<br>F: −9.7 (−25.0–−3.6) | A: 0.581 (−0.336–2.11)<br>F: 6.5 (−4.5–20.2) | A: 5.04 (0.068–13.8)<br>F: 55.2 (2.9–72.1) | A: 3.64 (1.54–7.16)<br>F: 38.3 (24.6–84.9) |
| TGO | Togo | thousands | A: 28.0 (11.6–52.4)<br>F: 28.8 (21.8–42.3) | A: 4.42 (1.81–7.76)<br>F: 4.5 (3.2–6.9) | A: −3.64 (−14.8–4.42)<br>F: −4.1 (−15.3–4.1) | A: 67.4 (24.1–136.2)<br>F: 70.8 (56.5–80.1) | A: 32.4 (13.4–60.0)<br>F: 33.3 (25.2–49.0) |
| TKL | Tokelau | persons | A: 22.7 (11.1–39.2)<br>F: 57.1 (44.0–81.6) | A: −4.42 (−8.73–−0.92)<br>F: −11.3 (−26.2–−2.3) | A: 5.86 (0.69–14.4)<br>F: 15.4 (2.4–27.1) | A: 15.1 (4.27–31.6)<br>F: 39.1 (17.8–54.4) | A: 18.2 (7.76–33.5)<br>F: 45.6 (33.3–64.6) |
| TON | Tonga | persons | A: −115.3 (−185.3–−63.6)<br>F: −16.2 (−29.0–−11.3) | A: 258.1 (144.7–400.1)<br>F: 36.2 (25.0–64.9) | A: 162.3 (50.1–334.6)<br>F: 23.4 (9.3–43.5) | A: 393.7 (89.5–840.3)<br>F: 56.6 (25.1–72.2) | A: 142.4 (79.4–218.6)<br>F: 20.0 (13.2–36.2) |
| TTO | Trinidad and Tobago | thousands | A: 3.03 (1.75–4.59)<br>F: 12.0 (7.2–34.5) | A: 7.28 (4.05–10.8)<br>F: 28.7 (16.6–83.5) | A: 1.97 (−1.79–6.57)<br>F: 8.2 (−10.2–30.3) | A: 12.6 (−3.20–32.7)<br>F: 51.2 (−34.8–72.7) | A: 10.3 (5.86–15.3)<br>F: 40.7 (24.1–118.0) |
| TUN | Tunisia | thousands | A: 32.5 (19.0–50.3)<br>F: 19.7 (11.5–71.9) | A: 48.6 (28.5–73.1)<br>F: 29.4 (16.9–108.2) | A: −0.0078 (−11.2–10.6)<br>F: 0.0 (−10.2–8.1) | A: 81.7 (−33.8–247.1)<br>F: 51.0 (−78.7–71.8) | A: 81.1 (47.6–123.2)<br>F: 49.1 (28.5–179.9) |
| TUR | Türkiye | thousands | A: 520.1 (387.0–673.8)<br>F: NR | A: 300.4 (225.7–381.7)<br>F: NR | A: −218.5 (−543.4–79.8)<br>F: NR | A: 278.2 (−936.8–1686)<br>F: NR | A: 820.7 (613.8–1054)<br>F: NR |
| TKM | Turkmenistan | thousands | A: 25.8 (11.9–47.1)<br>F: 61.2 (33.7–236.7) | A: 10.1 (3.99–19.0)<br>F: 23.7 (12.0–90.6) | A: −0.380 (−5.37–4.28)<br>F: −1.0 (−20.7–11.8) | A: 6.08 (−23.8–48.2)<br>F: 16.1 (−217.2–54.1) | A: 35.9 (16.1–65.7)<br>F: 85.0 (46.6–325.4) |
| TUV | Tuvalu | persons | A: −7.54 (−16.1–−2.70)<br>F: NR | A: 3.02 (−1.83–10.9)<br>F: NR | A: 7.00 (0.69–20.2)<br>F: NR | A: 16.6 (−5.68–60.6)<br>F: NR | A: −4.19 (−11.9–0.65)<br>F: NR |
| UGA | Uganda | thousands | A: 145.9 (58.4–285.1)<br>F: 42.5 (33.5–58.0) | A: −3.36 (−8.10–−0.028)<br>F: −1.0 (−2.2–0.0) | A: −25.5 (−77.0–5.87)<br>F: −8.0 (−21.4–1.5) | A: 225.2 (84.2–457.1)<br>F: 66.6 (53.9–76.4) | A: 142.5 (56.7–279.1)<br>F: 41.4 (32.7–56.6) |
| UKR | Ukraine | thousands | A: 37.1 (18.8–62.1)<br>F: 10.3 (5.2–47.7) | A: 50.9 (9.84–113.9)<br>F: 14.0 (2.4–65.2) | A: −145.8 (−290.6–−56.0)<br>F: −41.4 (−229.6–−10.0) | A: 408.2 (92.3–884.6)<br>F: 117.0 (89.2–226.6) | A: 88.2 (31.2–173.5)<br>F: 24.3 (9.6–111.8) |
| ARE | United Arab Emirates | thousands | A: 48.6 (26.4–80.7)<br>F: 41.1 (22.1–177.3) | A: −13.2 (−23.1–−5.98)<br>F: −11.2 (−50.0–−4.7) | A: 14.7 (2.50–35.1)<br>F: 12.8 (1.2–53.0) | A: 65.1 (−25.2–199.7)<br>F: 57.3 (−78.5–78.2) | A: 35.3 (18.1–60.8)<br>F: 29.9 (15.6–128.4) |
| GBR | United Kingdom | thousands | A: 99.2 (78.3–123.1)<br>F: NR | A: 61.5 (47.9–76.7)<br>F: NR | A: −229.3 (−432.5–−39.8)<br>F: NR | A: 17.0 (−782.2–953.6)<br>F: NR | A: 160.8 (127.1–198.9)<br>F: NR |
| USA | United States of America | thousands | A: 774.8 (638.1–922.9)<br>F: NR | A: 752.1 (613.4–894.1)<br>F: NR | A: −230.1 (−1173–744.1)<br>F: NR | A: −3651 (−7966–974.7)<br>F: NR | A: 1527 (1261–1805)<br>F: NR |

Countries 177–192. A: absolute contribution in stated unit (95% UI); F: % of ΔC (95% UI); NR: not reportable.

#### Section B. Absolute and fractional Horiuchi components



| ISO3 | Country | A unit | Population size D_N | Age–sex composition D_A | Age–sex-specific prevalence D_P | Control among people with hypertension, D_C | Demographic subtotal D_demo |
|---|---|---|---|---|---|---|---|
| URY | Uruguay | thousands | A: 16.1 (11.1–21.7)<br>F: 22.3 (14.8–45.8) | A: 5.50 (3.68–7.21)<br>F: 7.6 (4.7–16.2) | A: 0.634 (−8.04–9.47)<br>F: 0.9 (−14.6–12.9) | A: 49.6 (12.2–91.0)<br>F: 69.2 (37.6–82.4) | A: 21.6 (15.0–28.6)<br>F: 29.9 (19.7–61.8) |
| UZB | Uzbekistan | thousands | A: 184.9 (105.6–294.3)<br>F: 38.8 (24.7–95.0) | A: 24.3 (1.42–57.6)<br>F: 5.1 (0.4–13.9) | A: 138.2 (46.1–274.0)<br>F: 29.6 (12.0–69.9) | A: 123.5 (−123.4–472.2)<br>F: 26.7 (−70.2–53.6) | A: 209.3 (113.5–345.1)<br>F: 43.8 (27.7–106.7) |
| VUT | Vanuatu | thousands | A: 0.674 (0.264–1.36)<br>F: 25.4 (19.6–36.5) | A: 0.138 (0.028–0.329)<br>F: 5.2 (1.8–9.3) | A: 0.138 (0.0035–0.394)<br>F: 5.5 (0.2–11.3) | A: 1.66 (0.526–3.75)<br>F: 63.9 (48.3–73.0) | A: 0.814 (0.308–1.66)<br>F: 30.6 (23.2–44.0) |
| VEN | Venezuela | thousands | A: 113.4 (78.5–150.6)<br>F: 17.0 (12.0–29.3) | A: 123.1 (85.2–157.6)<br>F: 18.4 (12.6–32.1) | A: −9.25 (−78.7–58.8)<br>F: −1.4 (−14.4–8.4) | A: 439.7 (152.1–743.3)<br>F: 66.1 (42.1–78.0) | A: 236.6 (164.9–306.1)<br>F: 35.3 (24.7–61.2) |
| VNM | Viet Nam | thousands | A: 259.7 (173.4–366.4)<br>F: 11.3 (8.4–19.3) | A: 457.3 (304.6–628.1)<br>F: 19.9 (14.3–34.7) | A: 118.3 (−52.8–323.6)<br>F: 5.3 (−2.7–13.7) | A: 1446 (400.5–2802)<br>F: 63.5 (37.7–74.3) | A: 716.9 (479.0–993.3)<br>F: 31.3 (22.7–53.9) |
| YEM | Yemen | thousands | A: 158.0 (78.6–274.5)<br>F: 48.1 (32.1–106.0) | A: 32.2 (17.5–51.1)<br>F: 9.9 (6.3–22.1) | A: 3.70 (−21.4–33.0)<br>F: 1.2 (−8.1–9.5) | A: 130.5 (−34.4–386.0)<br>F: 40.9 (−29.2–60.7) | A: 190.4 (96.7–324.4)<br>F: 58.0 (38.7–127.9) |
| ZMB | Zambia | thousands | A: 74.0 (31.9–134.0)<br>F: 34.7 (26.8–48.1) | A: 16.9 (7.74–28.1)<br>F: 7.9 (5.9–11.1) | A: −33.3 (−76.3–−7.98)<br>F: −16.3 (−36.0–−3.8) | A: 155.6 (67.0–278.9)<br>F: 73.9 (61.8–85.6) | A: 90.8 (39.7–161.8)<br>F: 42.5 (33.1–58.9) |
| ZWE | Zimbabwe | thousands | A: 57.1 (16.7–118.2)<br>F: 33.4 (23.3–53.4) | A: 12.5 (−2.92–32.9)<br>F: 7.7 (−2.5–17.6) | A: −9.28 (−42.6–11.8)<br>F: −6.2 (−25.8–5.8) | A: 108.1 (33.0–215.9)<br>F: 65.3 (46.1–80.8) | A: 70.1 (20.2–142.7)<br>F: 41.1 (27.5–64.2) |

Countries 193–200. A: absolute contribution in stated unit (95% UI); F: % of ΔC (95% UI); NR: not reportable.

#### Definitions and reporting rules



Scope. All estimates use CORE-P25, the main dependence scenario (ρ_series = 0; ρ_country = 0), and the 2024–2030 forecast window.

Section A. C2024 and C2030 denote the number of adults aged 30–79 years with controlled hypertension. ΔC = C2030 − C2024.

Contribution to +150 M is 100 × ΔC/150 million. It is a linear rescaling of ΔC and is shown for all 200 countries, including those whose ΔC UI includes zero.

Section B. D_N is population size change; D_A is age–sex composition; D_P is age–sex-specific prevalence; D_C is control among people with hypertension; D_demo is the demographic subtotal D_N + D_A.

A denotes the absolute component in the unit stated for each country. F denotes 100 × D_g/ΔC, calculated draw by draw. Medians of components need not sum to the median of ΔC.

A component fraction can exceed 100% when its contribution alone exceeds ΔC and is offset by one or more components with the opposite sign.

NR means not reportable because the 2.5th percentile of ΔC does not exceed zero. Absolute contributions remain reportable. Exactly 160 countries have reportable fractions and 40 are marked NR.

Population estimates from the United Nations World Population Prospects were treated as deterministic. Uncertainty therefore reflects the modeled prevalence and control series, not population uncertainty.

Country order is alphabetical by displayed English name. Current WHO/UN display names are applied through a prespecified ISO3 map; analytical keys and frozen canonical names are unchanged. TWN remains UNCLASSIFIED.

Numeric presentation. Absolute values are shown in persons when |median ΔC| < 1,000 and in thousands otherwise; the unit is stated in each row. Percentages are rounded once, with half-up ties and no double rounding.

UI: uncertainty interval; WHO: World Health Organization.
