## Supplementary Methods S5 for "Forecasting global progress towards the United Nations hypertension control target for 2030"

Global forecasting and decomposition of hypertension control to 2030

### S5.1 Study design and data sources

We conducted a global model-based forecasting and decomposition study of hypertension prevalence and control among adults aged 30–79 years. Epidemiological estimates for 1990–2019 were obtained from the NCD Risk Factor Collaboration (NCD-RisC; NCD denotes noncommunicable disease), and age- and sex-specific population estimates and projections were obtained from the United Nations (UN) *World Population Prospects 2024* (WPP 2024), Medium variant.[1,2]

NCD-RisC provided estimates separately for men and women in ten 5-year age groups (30–34 to 75–79 years) for 200 countries and territories. Hypertension was defined as systolic blood pressure at least 140 mmHg, diastolic blood pressure at least 90 mmHg, or use of antihypertensive medication. Controlled hypertension was defined among people with hypertension as antihypertensive treatment with systolic blood pressure below 140 mmHg and diastolic blood pressure below 90 mmHg.[1] Prevalence and control were modelled separately and combined only when calculating absolute counts.

The 200 NCD-RisC entities were matched to WPP 2024 and represented approximately 99.8% of the global population aged 30–79 years in 2024 and 2030. WPP 2024 entities without corresponding NCD-RisC series—the largest being Hong Kong SAR (China), Kosovo, Réunion, and Macao SAR (China)—together accounted for the remaining approximately 0.2%; among WHO Member States, only Monaco and San Marino were affected. Results were aggregated to the six World Health Organization (WHO) regions: African Region (AFRO), Region of the Americas (AMRO), Eastern Mediterranean Region (EMRO), European Region (EURO), South-East Asia Region (SEARO), and Western Pacific Region (WPRO). Following World Health Assembly resolution 78.25 (WHA78.25), adopted on 27 May 2025, Indonesia was assigned to WPRO throughout the analytical series.[4] Taiwan, China was included in the global total in an analytical category labelled UNCLASSIFIED and was not assigned to one of the six WHO regions.

The primary policy estimand was progress toward the UN General Assembly target of 150 million additional people with hypertension under control by 2030, specified in paragraph 41 of the political declaration annexed to resolution A/RES/80/117.[3] Because the resolution does not specify an analytical baseline, the projected 2024 level was prespecified as the primary analytical baseline; 2020, 2023, and 2025 were examined in sensitivity analyses. Progress toward the target was calculated as the projected increase in the number controlled from 2024 to 2030 divided by 150 million. Regional values were expressed as contributions to this same global target and were not interpreted as region-specific targets.

### S5.2 Forecasting model

#### S5.2.1 State-space model

Hypertension prevalence and control were modelled on the logit scale using a Bayesian local-linear-trend state-space model. For age–sex stratum $j$ and year $t$,

$$
\begin{aligned}
y_{j,t} &= \ell_{j,t} + \epsilon_{j,t}, & \epsilon_{j,t} &\sim N(0, 0.02^2), \\
\ell_{j,t} &= \ell_{j,t-1} + b_{j,t-1}, \\
b_{j,t} &= b_{j,t-1} + \sigma_\beta \eta_{j,t}. \qquad (1)
\end{aligned}
$$

Here, $\ell_{j,t}$ is the latent level and $b_{j,t}$ its local slope. The model therefore allows the direction and rate of change to evolve over time rather than imposing a fixed linear trend. Related Bayesian approaches have been used for population-level blood-pressure trajectories.[5]

Within each country and epidemiological series, slope innovations were correlated across the 20 age–sex strata using nested common, sex-specific, and stratum-specific Gaussian components, with $0 \leq \rho_{\text{common}} \leq \rho_{\text{sex}} \leq 1$. Prevalence and control were fitted separately for each country.

The slope-innovation scale $\sigma_\beta$ had a half-normal prior with scale 0.01; transformed correlation parameters had $N(0, 2^2)$ priors. Posterior inference for the primary model used four Markov chain Monte Carlo (MCMC) chains. Convergence was assessed using effective sample size and split-$\widehat{R}$, with all primary global fits satisfying the prespecified convergence criteria.

##### S5.2.2 Treatment of modelled uncertainty and construction of 95% uncertainty intervals

A key feature of the analysis was that uncertainty was modelled for the *entire* epidemiological trajectory rather than copied year by year from the published NCD-RisC intervals. NCD-RisC annual estimates and their 95% credible intervals are themselves outputs of a Bayesian hierarchical model, not independent sampling estimates for each calendar year.[1] Consequently, year-specific interval widths also reflect temporal smoothing and model behaviour near the boundaries of the fitted time series. In particular, intervals near the beginning and end of a modelled series may be affected by boundary-related widening or narrowing and were therefore not treated as direct observations of annual measurement variance.

We separated two sources of uncertainty. First, the state-space likelihood represented uncertainty around the latent temporal trajectory. Second, a persistent external level-uncertainty component represented uncertainty in the NCD-RisC epidemiological level. This second component was calibrated from the published NCD-RisC uncertainty intervals using the CORE-P25 rule: for each age–sex stratum, lower- and upper-side uncertainty scales were derived separately on the logit scale as the 25th percentile of their annual values over the interior 1997–2012 reference window. This rule was selected during preliminary model development and fixed before global production, thereby reducing sensitivity to interval behaviour at the temporal boundaries. The published 2019 central estimate was retained as the anchor for the forecast.

Three correlation parameters governed uncertainty propagation. $\rho_{\text{strata}}$ represented correlation of persistent level uncertainty across the 20 age–sex strata within the same country and epidemiological series and was fixed at 0.70 in all reported analyses. $\rho_{\text{series}}$ represented prevalence–control dependence within the same country. In the dependence sensitivity analyses, it was applied to both the persistent level component and the rank pairing of the prevalence and control forecast-dynamic draws, thereby changing their joint dependence while preserving each marginal distribution. $\rho_{\text{country}}$ represented dependence of persistent level uncertainty across countries and did not induce shared dynamic innovations between countries. These parameters were distinct from $\rho_{\text{common}}$ and $\rho_{\text{sex}}$, which governed slope innovations across age–sex strata within each fitted epidemiological series. Because cross-series and cross-country dependence cannot be identified from the published marginal NCD-

RisC uncertainty intervals, the primary analysis used $\rho_{\text{series}} = 0$ and $\rho_{\text{country}} = 0$, with alternative values examined as sensitivity analyses.

For the annual NCD-RisC points displayed in the historical portions of the main trajectory figures and Supplementary Figure S3, the published age–sex-specific lower and upper bounds were converted to asymmetric logit-scale deviations and propagated to country, regional, and global aggregates under the primary dependence assumptions. These error bars are therefore propagated aggregate intervals rather than uncertainty intervals published directly by NCD-RisC, and they are distinct from the historical model band.

For 1990–2019, the displayed model-based historical 95% uncertainty intervals (95% UIs) combined the smoothed fitted trajectory with the calibrated persistent level uncertainty. For 2020–2030, the 95% UIs additionally incorporated uncertainty from the evolving latent slope and future state innovations. Thus, uncertainty was represented continuously across the full 1990–2030 curve, while acknowledging that historical and forecast intervals have different components. The 2019–2024 segment is entirely model-based because NCD-RisC provides no observations after 2019; this interval also spans the coronavirus disease 2019 (COVID-19) pandemic.

#### S5.2.3 Forecast generation

For each reported scenario, 100,000 posterior draws were propagated from 2019 through 2030. Forecasts were anchored to the published 2019 NCD-RisC central estimate and incorporated the calibrated level uncertainty, uncertainty in the terminal slope, and future slope innovations. Draws were transformed from the logit scale back to probabilities before aggregation.

The primary model preserved the local slope dynamically throughout the forecast horizon.

### S5.3 Absolute counts and aggregation

For country $p$, age–sex stratum $j$, year $t$, and posterior draw $r$, the number of adults with controlled hypertension was calculated as

$$C_{p,t}^{(r)} = \sum_{j} N_{pjt}\, Prev_{pjt}^{(r)}\, Ctrl_{pjt}^{(r)}, \tag{2}$$

where $N_{pjt}$ is the WPP population, $Prev_{pjt}^{(r)}$ is hypertension prevalence, and $Ctrl_{pjt}^{(r)}$ is control among adults with hypertension.

Country counts were summed draw by draw to obtain WHO regional and global estimates. Aggregate hypertension control was calculated as the number controlled divided by the number with hypertension within each draw; stratum- or country-level control percentages were not averaged directly. Because posterior summaries were calculated after draw-level aggregation, separately reported regional medians need not sum exactly to the global median.

For the country-level presentation in Figure 4, the 2024–2030 change in the number controlled was also expressed per 1,000 adults aged 30–79 years, using the 2024 WPP population as denominator. Countries highlighted for absolute contribution were ranked by posterior mean change, because posterior means are additive across countries. The display rule selected the smallest set reaching 80% of the global posterior mean change, with a minimum of five and a maximum of 12 countries; if the 80% threshold was not reached by 12 countries, the display remained capped at 12. Countries with a negative median change were additionally displayed. Cartographic boundaries and small-country points were obtained from Natural Earth 1:50m cultural vectors, version 5.1.1; Natural Earth uses predominantly de facto international boundaries, a cartographic convention that did not alter the analytical country assignments.[7]

### S5.4 Decomposition of change in controlled hypertension

The change in the absolute number of adults with controlled hypertension was decomposed using the continuous-change method of Horiuchi, Wilmoth, and Pletcher.[6] Four components were considered: total population size, age–sex composition, age–sex-specific hypertension prevalence, and control among adults with hypertension. For each posterior draw,

$$\Delta C = D_N + D_A + D_P + D_C, \tag{3}$$

where $D_N$ is the contribution of population size, $D_A$ of age–sex composition, $D_P$ of hypertension prevalence, and $D_C$ of control. The demographic subtotal was defined as $D_{\text{demo}} = D_N + D_A$.

The continuous-change integral was evaluated numerically and closure was verified within each posterior draw. Country-level components were calculated first and then summed to regional and global levels, preserving additivity.

Component shares were calculated draw by draw as $100D_g/\Delta C$. Shares were reported only when the 2.5th percentile of $\Delta C$ was greater than zero; absolute component contributions remained reportable when this criterion was not met. Because posterior medians were taken after draw-level calculations, median components need not sum exactly to the median total change, and median component shares need not sum exactly to 100%. A component share could be negative or exceed 100% when components acted in opposing directions.

### S5.5 Historical versus future decomposition

To compare recent historical dynamics with the forecast period, the same decomposition was applied to matched six-year windows: 2013–2019 and 2024–2030. Absolute changes were annualized, and future-minus-historical differences in component shares were calculated within aligned posterior draws.

For selected contrasts, we report the posterior proportion of paired draws in which the difference was above or below zero. These are posterior probabilities and are not frequentist $P$ values. Because forecast intervals additionally contain future process uncertainty, their width is not directly comparable with historical interval width as a measure of calibration.

### S5.6 Sensitivity analyses

Reported sensitivity analyses examined alternative analytical baselines (2020, 2023, and 2025) and the dependence assumptions underlying the 95% UIs. All baseline-year analyses used the same posterior draws. Supplementary Figure S1B presents differences between marginal posterior median component shares relative to the primary 2024 baseline; paired uncertainty intervals for these baseline-to-baseline differences were not generated in the frozen analytical output.

Dependence assumptions were assessed factorially using $\rho_{\text{series}} = -0.5, 0,$ or $+0.5$ and $\rho_{\text{country}} = 0$ or $+0.5$. As defined above, $\rho_{\text{series}}$ varied prevalence–control dependence within countries in both the persistent level and forecast-dynamic components, whereas $\rho_{\text{country}}$ varied only the persistent level dependence across countries. $\rho_{\text{strata}}$ remained fixed at 0.70. These analyses changed uncertainty propagation and joint dependence while preserving the marginal epidemiological forecasts. WPP 2024 population projections were treated as deterministic throughout.

### S5.7 Reporting and reproducibility

The study used secondary modelled estimates and population projections and did not involve identifiable individual-level data. Reporting follows the Guidelines for Accurate and Transparent Health Estimates Reporting (GATHER) principles for health estimates.[8] NCD-RisC estimates and WPP 2024 population data are available from their respective public sources. Analytical code and supplementary outputs are available at Zenodo.[9]
